# A new lineage nomenclature to aid genomic surveillance of dengue virus

**DOI:** 10.1101/2024.05.16.24307504

**Authors:** Verity Hill, Sara Cleemput, Vagner Fonseca, Houriiyah Tegally, Anderson F. Brito, Robert Gifford, Vi Thuy Tran, Duong Thi Hue Kien, Tuyen Huynh, Sophie Yacoub, Idrissa Dieng, Mignane Ndiaye, Diamilatou Balde, Moussa M. Diagne, Oumar Faye, Richard Salvato, Gabriel Luz Wallau, Tatiana S. Gregianini, Fernanda M.S. Godinho, Chantal B.F. Vogels, Mallery I. Breban, Mariana Leguia, Suraj Jagtap, Rahul Roy, Chanditha Hapuarachchi, Gaspary Mwanyika, Marta Giovanetti, Luiz C.J. Alcantara, Nuno R. Faria, Christine V.F. Carrington, Kathryn A. Hanley, Edward C. Holmes, Wim Dumon, Tulio de Oliveira, Nathan D. Grubaugh

## Abstract

Dengue virus (DENV) is currently causing epidemics of unprecedented scope in endemic settings and expanding to new geographical areas. It is therefore critical to track this virus using genomic surveillance. However, the complex patterns of viral genomic diversity make it challenging to use the existing genotype classification system. Here we propose adding two sub-genotypic levels of virus classification, named major and minor lineages. These lineages have high thresholds for phylogenetic distance and clade size, rendering them stable between phylogenetic studies. We present an assignment tool to show that the proposed lineages are useful for regional, national and sub-national discussions of relevant DENV diversity. Moreover, the proposed lineages are robust to classification using partial genome sequences. We provide a standardized neutral descriptor of DENV diversity with which we can identify and track lineages of potential epidemiological and/or clinical importance. Information about our lineage system, including methods to assign lineages to sequence data and propose new lineages, can be found at: dengue-lineages.org.

## Introduction

Dengue virus (DENV: *Flaviviridae*; *Orthoflavivirus*) inflicts the heaviest global burden on public health of any mosquito-borne virus, causing more than 100 million infections per year (Bhatt et al. 2013). Dengue incidence is increasing worldwide, with major outbreaks across endemic regions in the tropics in 2023 (World Health Organization 2023), and sustained local transmission in non-endemic regions such as the state of Florida in the US (Jones et al. 2024) and Italy (Vita et al. 2024; Branda et al. 2023). As DENV continues to spread, tracking the evolution at a high resolution is key to understanding its transmission patterns on local, regional, and global scales.

Dengue virus in its human-endemic cycle consists of four serotypes (DENV-1-4) that likely correspond to at least four successful spillover events from its ancestral sylvatic cycle that took place several centuries ago (Vasilakis et al. 2011). Each serotype includes several genotypes that were designated in the late 1990s and early 2000s based on partial genetic sequences (Rico-Hesse 1990; Twiddy, Holmes, and Rambaut 2003). In addition, DENV-2 and DENV-4 include genotypes encompassing viruses from the sylvatic cycle. These serotypes and genotypes have provided the basis for decades of work characterizing the natural history, phenotypic diversity, and transmission dynamics of DENV. However, with recent large increases in global sequencing capacity and its integration into public health systems, additional granularity of DENV diversity is required. Several previous studies have already classified sub-genotypic diversity on a country or regional level (e.g. (OhAinle et al. 2011; de Araújo et al. 2012; Adelino et al. 2021), but there is a need to standardize this discussion between research groups and countries to aid communication and facilitate identification of common patterns. The continued evolution and spread of DENV has led to the emergence of distinct evolutionary lineages within recognized genotypes. Further, with the implementation of interventions (e.g. vaccines, *Wolbachia*-infected mosquitoes) that may eventually select for specific viral lineages, it is imperative to have a precise and common language to monitor continued DENV transmission in different spatio-temporal scales; and that this is communicable to clinicians and public health officials who may not have a background in genomics.

Here, we propose a system for the classification and nomenclature of DENV lineages that builds on existing serotype and genotype classifications to (**1**) provide additional temporal and spatial granularity and (**2**) standardize the discussion of important diversity globally. We take inspiration from the design of the pango nomenclature system, a hierarchical lineage system set up to track SARS-CoV-2 evolution (Rambaut et al. 2020), as well as lessons learned from its design and implementation. We discuss the design, validation, and application of our proposed DENV lineage system, show how it enhances resolution when monitoring circulating lineages, and introduce tools enabling end-users to assign lineages to their own sequences. By making the system compatible with existing classifications and showcasing its utility, we aim to achieve widespread uptake and introduce a truly standardized global language with which to discuss DENV genetic diversity.

## Results and Discussion

### Previously defined genotypes provide useful but not sufficient resolution

Dengue virus is currently classified into four serotypes, which in turn include varying numbers of genotypes: five for DENV-1, six for DENV-2, five for DENV-3, and four for DENV-4 (**Figure 1**). Genotype classification systems were originally based on greater than 6% pairwise genetic distance within the genotype using a 240 nucleotide sequence (i.e. a single amplicon) of the envelope (E)/nonstructural protein 1 (NS1) protein coding region, as this arbitrary threshold split then-known DENV-1 diversity into manageable groups (Rico-Hesse 1990). As more sequence data was generated, these were replaced by systems based on entire protein-coding regions, especially E, with which many of the current genotypes were designated 20-30 years ago (e.g. (Twiddy et al. 2002; Lanciotti et al. 1994; Lanciotti, Gubler, and Trent 1997)).

**Figure 1.**
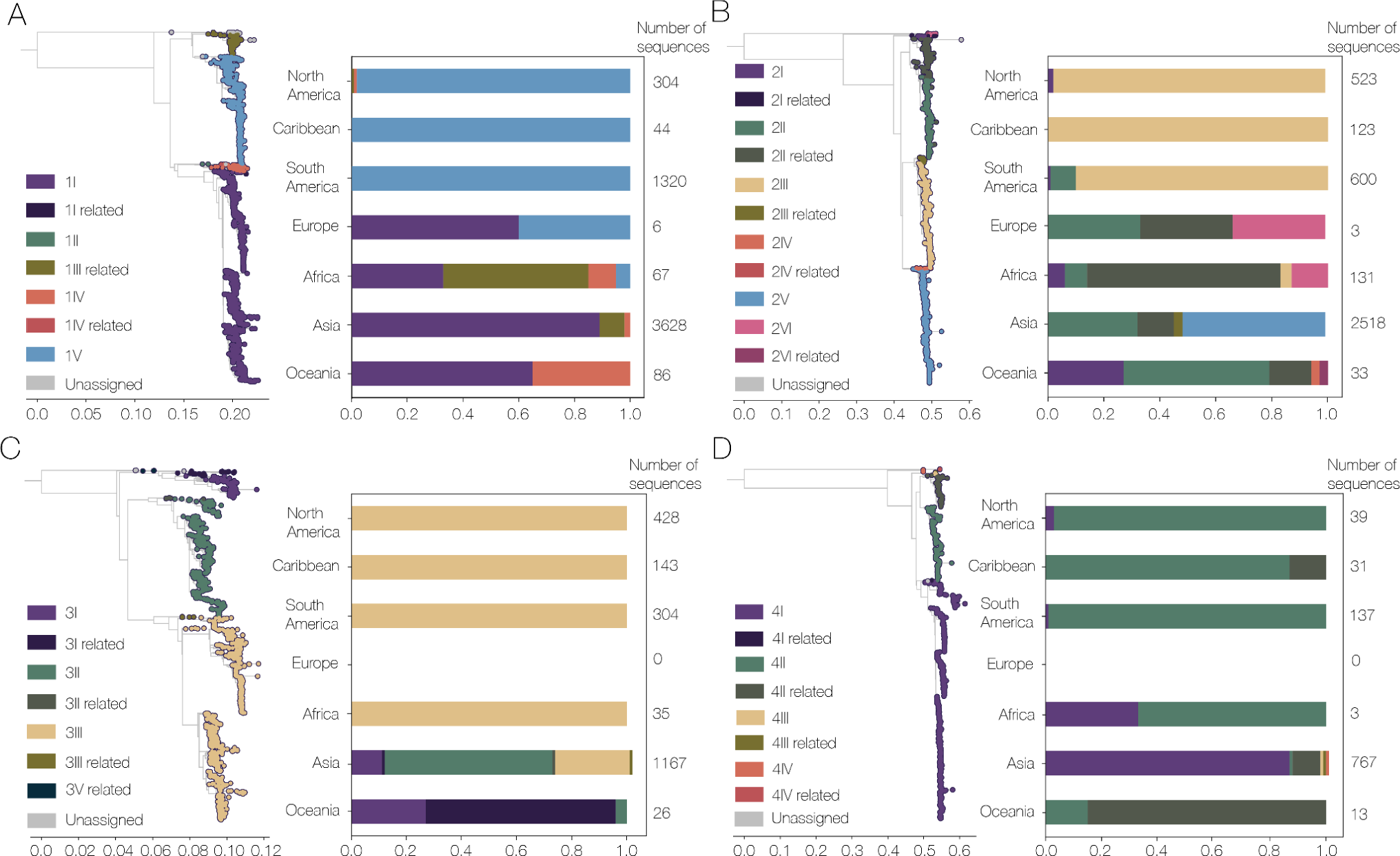
| Current system of classifying dengue virus serotypes into distinct genotypes provides insufficient geographical resolution. Maximum likelihood phylogenetic trees scaled by genetic distance for each DENV serotype: A) DENV-1; B) DENV-2; C) DENV-3 and D) DENV-4. They are colored by the current genotype classification obtained using the Genome Detective Dengue Virus Typing Tool (Fonseca et al. 2019; Vilsker et al. 2019). Bar charts indicate the frequency of whole genomes sampled in each continent assigned to each existing genotype, numbers at the end of each bar indicate the number of sequences in each dataset. Note that every serotype has a dominant genotype across the Americas (i.e. North America, Caribbean, and South America). “Related” refers to sequences that are not reliably placed into the clade as there is considerable bootstrap support for the clade without the query as well as with the query.

This serotype/genotype nomenclature system still holds well with newer whole genome sequences, with some geographic distinction between continents. For example, within DENV-1 we found that the Americas are dominated by genotype V, whereas Asia and Oceania have more sequences of genotype I (**Figure 1A**). Further, much of the existing research uses genotypes to characterize circulating variants (e.g. (S. C. Hill et al. 2019), ensure adequate genomic diversity for sequencing panels (Santiago et al. 2013; Vogels et al. 2024), identify new introductions leading to outbreaks (Ma et al. 2021), explore differences in viral fitness, and disease association (Cologna, Armstrong, and Rico-Hesse 2005). However, there has been a huge increase in publicly available sequence data, both in terms of number and regions from which samples are being sequenced, since these genotype classification systems were established. This has led to a similarly large increase in the known genetic diversity within each serotype. In particular, there are now groups of genomes that do not reliably fall into genotypes: 6.41% of DENV-1 sequences, 12.8% of DENV-2, 2.14% of DENV-3, and 9.75% of DENV-4 sequences are related to defined genotypes, but cluster basally to them (designated as “related” by Genome Detective Dengue Virus Typing tool (Fonseca et al. 2019; Vilsker et al. 2019); **Figure 1**). A smaller number of genomes in each serotype do not fall into any currently-defined genotype and are designated as “unassigned” (0.60% DENV-1, 0.25% DENV-2, 0.14% DENV-3, 0.10% DENV-4). This increase in the known genetic diversity of circulating lineages also leads to complexities on the sub-genotypic level, as some genotypes are now very large and highly diverse - 5 out of 17 genotypes contain more than 1000 whole genome sequences. Of particular note, DENV-1 genotype I contains 3293 published sequences, which is more than three times the size of the entire DENV-4 whole genome data set that we used here (n = 995). The large number of genomes in many of these genotypes, combined with increased air travel and the expanded range of the mosquito vectors (Ryan et al. 2019; Kraemer et al. 2019), lead us to conclude that genotypes alone do not provide sufficient resolution for many epidemiological questions (**Figure 1**). For example, we found that the current genotype assignment does not provide additional epidemiological information than the serotype for DENV-1, DENV-2, and DENV-3 in the Americas, as they are dominated overwhelmingly by a single genotype for each serotype. Indeed, many previous studies in this region have already named sub-lineages within genotypes (e.g., lineage classification systems in Brazil and Nicaragua) (Williams et al. 2014; OhAinle et al. 2011; Drumond et al. 2013; Thongsripong et al. 2023; de Carvalho Marques et al. 2023). Therefore, while genotypes provide an important base for research, there is clearly a need for an updated classification system to include newly recognized global diversity and provide additional sub-genotype resolution in a systematic and standardized way.

### New lineage classification system design

To better describe the circulating diversity of DENV, we propose a new system that builds on the existing serotype/genotype system discussed above (**Figure 2**). First, we updated the genotype definitions so that fewer genomes are unassigned or ambiguously classified as “related”. Then, we added two additional layers of resolution within the genotypes, major and minor lineages, with associated nomenclature.

**Figure 2.**
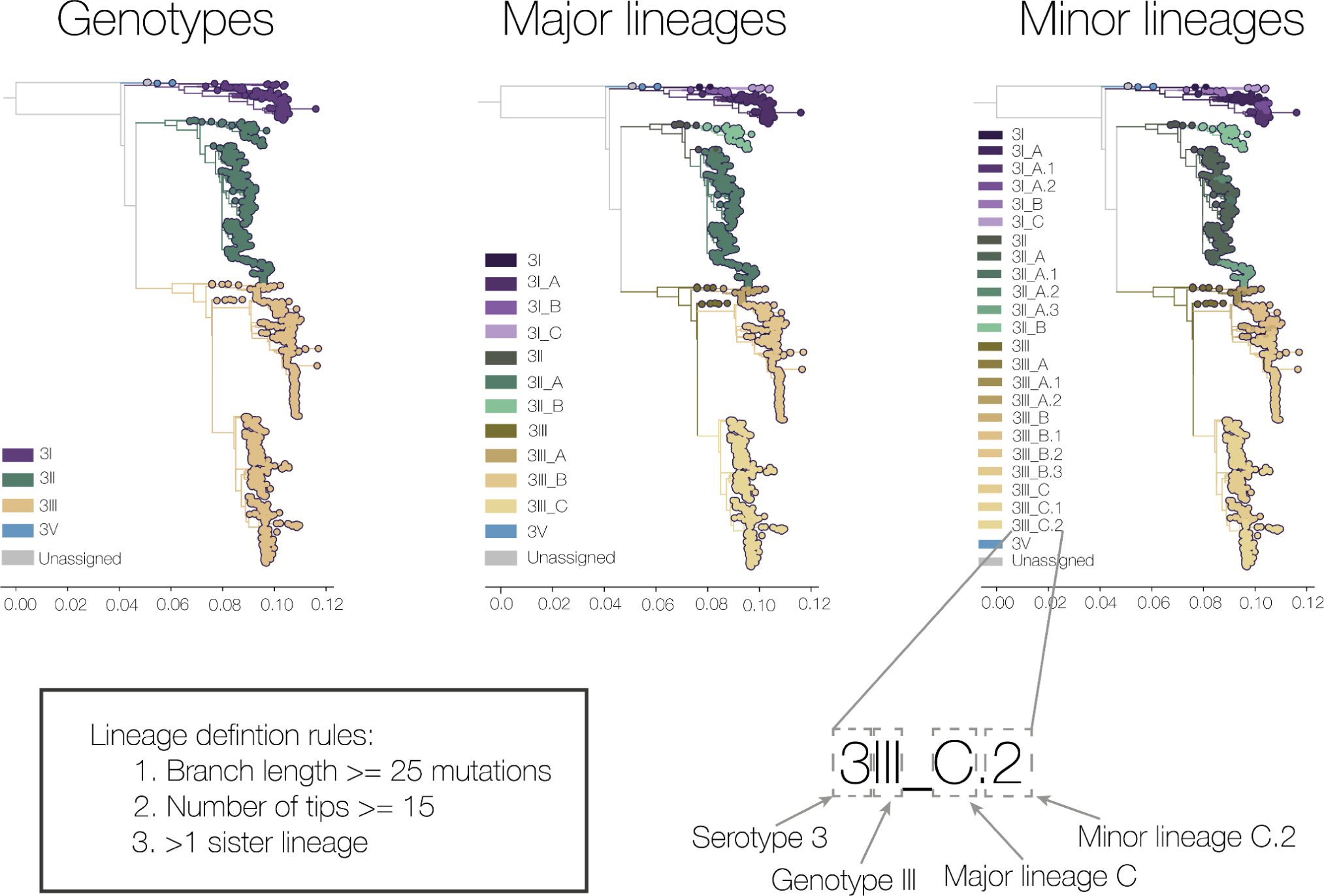
| Description of proposed dengue virus lineage classification system using serotype 3 as an example. Genotype level classification has been expanded to include most previously unassigned genomes. Two additional layers of classification have been proposed, major and minor lineages, the rules of which are shown here. The nomenclature is described here in its shorthand form (e.g. DENV-3III_C.2), with each element highlighted by a dotted box. The new lineage classifications for serotypes 1, 2, and 4 are shown in **Figure S2**.

We found that many DENV sequences fall basal to the genotype-defining nodes, meaning that they do not fit cleanly within the current classification system and are thus classified as “related” to a genotype (**Figure 1**). We identified 16 categories of “related” genotypes within our global DENV genomic data set. We therefore moved the genotype-defining node closer to the root of the tree so that they were included in the main definition of the genotype. While we did not remove any of the already defined genotypes, we did redefine DENV-1 genotype VI from a proposed definition (Pyke et al. 2016). The original DENV-1 genotype VI proposition still contains only one sequence (GenBank accession number: KR919820), so we instead propose that the genotype VI name be used on a previously undefined but related clade containing 27 sequences primarily from the Democratic Republic of the Congo. The KR919820 sequence (Pyke *et al* 2016) is currently “Unassigned” in our system, but should more sequences fall into that clade, it will likely receive a genotype definition as it would have established transmission and become more public health relevant. Even after our adjustments, some unassigned sequences remain (two sequences for each serotype, DENV-1= 0.037%, DENV-2 = 0.051%, DENV-3 = 0.095%, DENV-4 = 0.20%), but these are mostly basal to other defined genotypes or singleton outliers. For DENV-1, DENV-2, and DENV-4 the sequences are sylvatic sequences from Malaysia, Borneo, and Australia. For DENV-3, these sequences are older (1953 and 1963) from Puerto Rico.These updated genotype definitions also still fit the original arbitrary definition of less than 6% pairwise genetic distance within a genotype, even across the whole genome. It is of note, however, that the pairwise distance within genotypes is highly variable (**Figure S1A**), and this variation is not related to the number of sequences within a genotype (**Figure S1B**).

We propose to consistently use Roman numerals to refer to genotypes, thereby removing the current geography-based names for DENV-2 genotypes. We are motivated by two key reasons: (**1**) some of these genotypes are now very widespread and are not limited to the region that the name implies and (**2**) geographical names can lead to discrimination, especially when they cause large outbreaks, and as such are against best practices for naming pathogens (World Health Organization 2015). We use the standard Roman numerals for these genotypes instead, and the comparison between the systems can be found in **Table S1**.

The new genotype definitions that we propose succeed in reducing the number of unassigned DENV sequences. Large geographical spaces, however, are still dominated by single genotypes within a serotype. Therefore, we propose two additional levels of classification: major and minor lineages (**Figure 2 and S2**). Major lineages, designated using letters of the Roman alphabet, are designed to help answer regional scale questions. Minor lineages, designated using numbers and full-stops, provide more fine-scale resolution, and therefore have a nomenclature more similar to SARS-CoV-2 pango lineages (Rambaut et al. 2020). Importantly, like the existing genotypes, this lineage nomenclature system is *evolutionarily neutral* - i.e. they are designated based entirely on phylogenetic metrics and not on any possible phenotypic differences. This provides an *a priori* system for naming clades, thereby providing a framework to identify possible phenotypic changes when they arise. For example, the sudden growth of a single clade, which may indicate a change in transmissibility or immune evasion, may be more easily identified when many sequences are rapidly assigned to a lineage that is named consistently throughout the world. We also note that lineage definitions are based on the *nodes* of the phylogeny, rather than the tips.

Major and minor lineages are strictly hierarchical. We use the same defining rules for both levels of at least (**1**) 15 sequences (arbitrary cutoff), (**2**) 25 inferred nucleotide substitutions (across the whole genome) along the ancestral branch (arbitrary), and (**3**) one sister lineage at the same level - in other words, there cannot be an A lineage without a B lineage (**Figure S3**). The first two rules aim to capture epidemiologically important lineages, which are stable between iterations of phylogenetic inference. The high phylogenetic distance is possible due to the high genomic diversity of DENV, and builds on experience in the pre-variant era of SARS-CoV-2. Due to its low global genetic diversity, SARS-CoV-2 lineages were defined on a single evolutionary event (i.e. a substitution, an indel or a recombination event (Rambaut et al. 2020) and so would sometimes break monophyly when new trees were inferred, causing issues with communication. Our thresholds are also high to avoid high levels of nesting in the names of the lineages at this stage, also a lesson learned from SARS-CoV-2 lineage system designs. The final rule on compulsory sister lineages is to ensure that each designation level provides new information that is usable for public health - i.e. not simply the same lineage circulating in the same area year after year, but a distinguishable clade that differentiates it from other geographical areas. Some manual curation was also performed on this initial designation step for major lineages that were still very large (see **Methods**), but we do not anticipate needing to do this in future lineage releases as we will be designating major lineages prospectively, obviating the same very large unbroken diversity observed with retrospective designation.

Combining our updates to the genotype placements and the addition of lineages, sequences can be discussed using a formal longhand and a simpler shorthand nomenclature. For example, “Dengue virus serotype 3, genotype III, lineage C.2” can be abbreviated as “DENV-3III_C.2” (read as: “dengue three-three-C-dot-two”; **Figure 2**).

### Applications of the new lineage system

After designing a new lineage classification system and applying it to the global DENV genomic dataset, we evaluated its utility for addressing real-world public health questions. We specifically tested the lineage system based on its two key design principles: (**1**) to increase resolution with which to discuss genetic diversity and (**2**) to standardize the discussion.

We first examined whether splitting the phylogenetic trees of each DENV serotype beyond genotypes led to increased temporal and spatial resolution. While some continents are now dominated by a single major lineage (e.g. South America for DENV-3 and DENV-4), most continents have at least two major lineages designated for each serotype (**Figure 3**). For example, almost all DENV-1 whole genome sequences in the Americas in this dataset are genotype V, but we can now split the genotype V viruses circulating in this region into seven major lineages (**Figure 3**).

**Figure 3.**
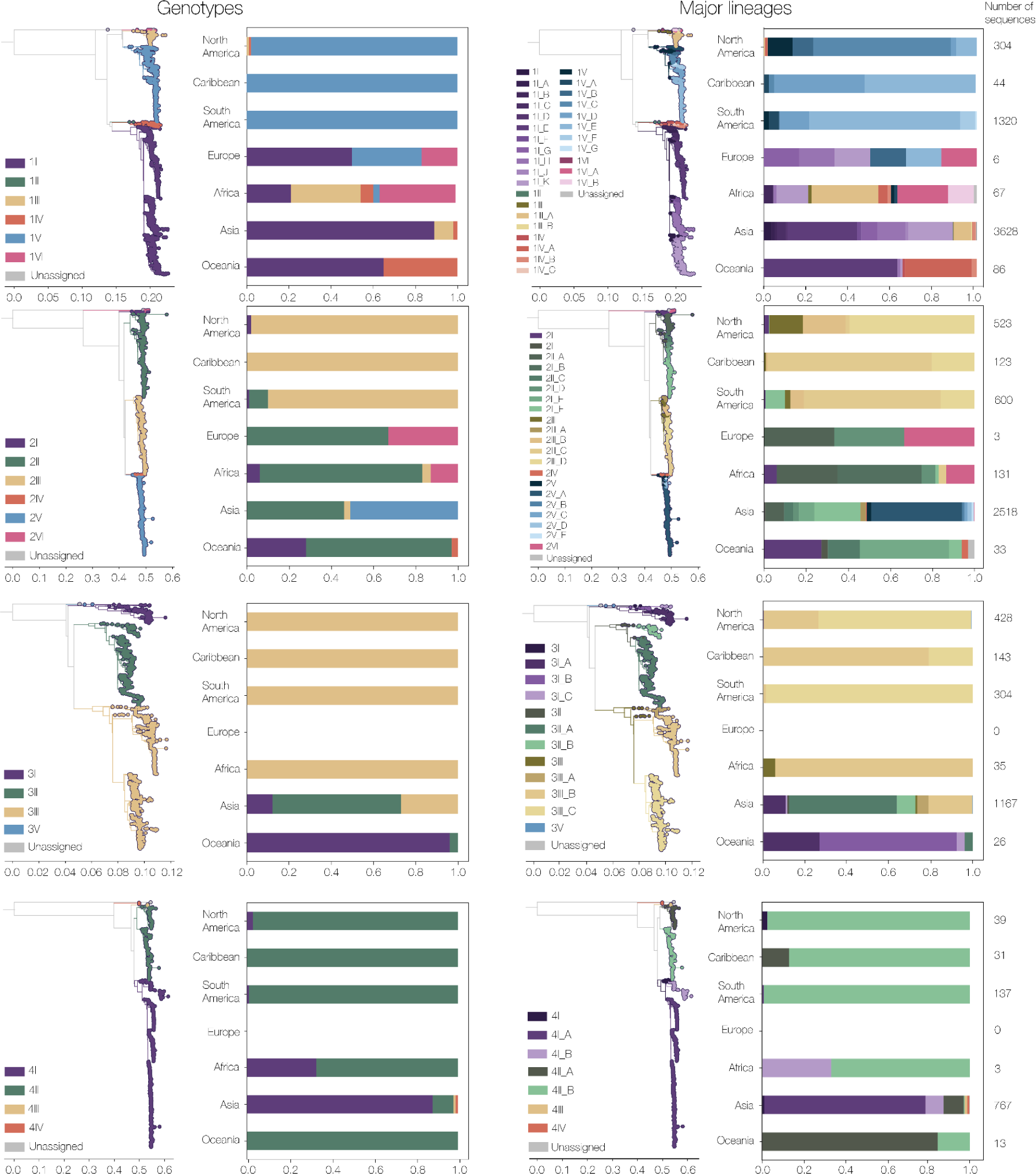
| Global distributions of the new classifications of dengue virus genotypes and major lineages. Each serotype’s new genotype (first column) and major lineage (second column). Genetic distance trees are colored by genotype or major lineage, and bar charts show the percentage of whole genome sequences in each continent by classification level. Note that major lineages break down genotypic diversity further and provide additional resolution and a continent level. Numbers by each bar represent the number of sequences in each continent by serotype.

To further explore this apparent increased resolution, we mapped the sampling location of every sequence in each level of lineage designation to identify whether geographic scope also narrows as classification level decreases. While some minor lineages are relatively widespread, we find several examples of increased geographic resolution with increased phylogenetic nesting (**Figure 4A**). For example, DENV-2 genotype II has been identified in all regions where DENV circulates. When we assign sequences from this genotype to major lineages, the major lineage DENV-2II_A sampling locations occur only in the eastern hemisphere. When further exploring the minor lineages, 2II_A.2.1 has so far only been detected in India and Kenya while 2II_A.2.2 has a larger known distribution in south and east Asia (**Figure 4A**). In this scenario, if a related DENV was sequenced in East Africa, the minor lineage assignment would immediately provide clues about whether it was a potential new introduction from Asia to Africa (i.e. 2II_A.2.2) or whether it may have arisen autochthonously within the region (i.e. 2II_A.2.1). In comparison, under the existing system, the hypothetical new lineage would simply be assigned to DENV-2 genotype II, also known as the Cosmopolitan genotype, which is one of the most diverse and widespread genotypes of DENV and little additional information would be gleaned without conducting phylogenetic analysis. We also show how the major lineages of DENV-1 genotype V, which dominates DENV-1 transmission in the Americas (**Figure 1**), provide additional geographic resolution in this region (**Figure 4B**). For example, DENV-1V_B is sampled in Nicaragua, the US, Mexico, Venezuela, and Guatemala; whereas DENV-1V_E is dominated by sequences sampled in Brazil. Notably, DENV-1V_G is only found in Colombia and Trinidad and Tobago. These major lineages therefore provide information on different patterns of circulation of DENV-1 in the Americas.

**Figure 4.**
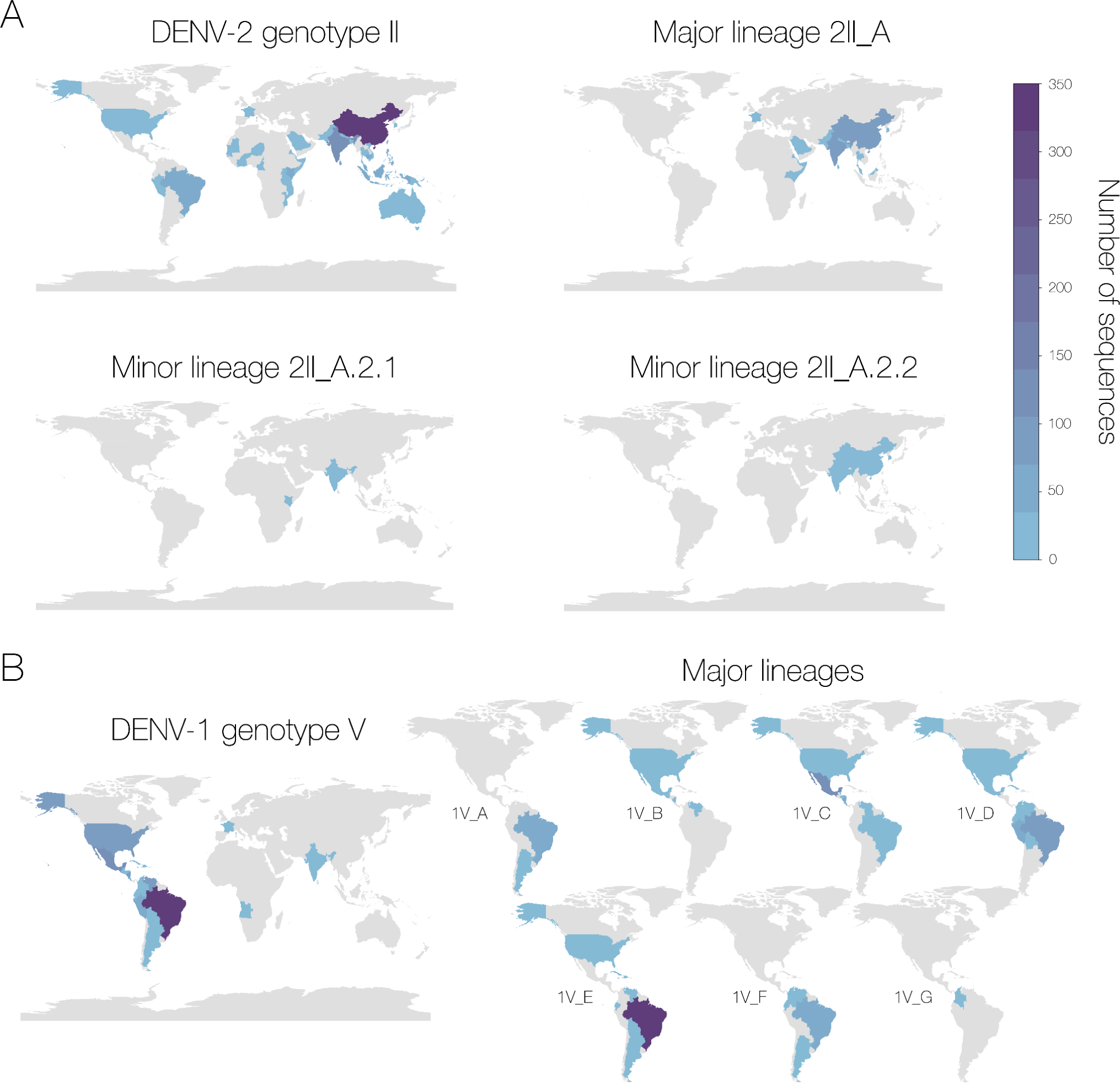
| Examples of increased geographical resolution using the new classifications of dengue virus lineages. A) Each map shows the number of sequences in the dataset in each country classified as, respectively, DENV serotype 2 genotype II, serotype 2 genotype II major lineage A, and then two minor lineages of A.2.1 and A.2.2. The color represents the number of genome sequences from blue to purple running from low to high. B) Map shows the distribution of the whole of DENV-1 genotype V and all of its constituent major lineages in the Americas. Major lineage 1V_A also has sequences from India and 1V_B and 1V_E have some from France although these are not shown here in the interests of space. Color represents the number of genome sequences on the same scale as panel A.

In the absence of a global lineage classification system, individual research groups have labeled lineages using their own nomenclature systems to aid research and surveillance efforts. While changing names can be challenging, it is important to have a standardized nomenclature system to aid discussion between different regions. This makes it easier to rapidly identify which lineages are the same in different countries and therefore which are spreading internationally, possibly indicating a relevant phenotypic property. For example, there has been a new introduction of DENV-3 genotype III from Asia into the Caribbean, which has since spread across the Americas and has been reintroduced into Asia and Africa (Jones et al. 2024; Naveca et al. 2023; Miguel et al. 2024; Branda et al. 2023; Taylor-Salmon et al. 2023). This lineage spread may be connected to the large DENV-3 outbreaks in 2023 in the region, and so there is a risk of different research groups naming this lineage separately (e.g., “DENV-3 GIII-American-II lineage”) making it harder to detect the wider pattern of spread and phylogenetic relatedness. In the new system, this introduction has been designated a minor lineage 3III_B.3 (**Figure S4A**). We note that a few basal sequences in Asia are included in this lineage and so not every sequence assigned to this minor lineage is necessarily part of the same introduction, but it provides a common language to discuss this epidemiologically important lineage.

Further, our system has equivalents to many of the existing sublineages individually defined by other research groups. For example, four lineages of DENV-3 genotype III have been described in Brazil (de Araújo et al. 2012). BR-I and BR-II fall into DENV-3III_C, and while the single sequence in BR-III is not in our dataset as it is only an E sequence, its closest whole genome relative (genbank accession: FJ898462) is also in 3III_C. The single sequence representing BR-IV is assigned to 3III_C.1. While three of the named Brazil lineages fall into the same major lineage in our system, it still provides a separation from the newly introduced 3III_B.3 lineage of the same genotype that we describe above (**Figure S4A**). Similarly, in DENV-2 genotype III, there are lineages in Brazil defined as BR1-4 (Drumond et al. 2013; de Carvalho Marques et al. 2023; Adelino et al. 2021). In our system, BR1 is DENV2-2III_B, BR2 does not get assigned to a major lineage because it does not meet the size threshold required and so it is just 2III, BR3 is DENV-2III_C, and BR4 is 2III_C.1.1 (**Figure S4B**). We note that only BR4 in DENV-2 is a 1:1 match with our lineage system and thus there may still be reason for research groups to use finer resolution of lineage classification for their specific needs. Our goal is not to restrict these activities, rather to persuade the use of common classifications for external communications. To aid this effort, we included lineage assignments for every whole genome sequence in our dataset (**Table S2**).

While we deliberately do not take phenotypic differences into account in the designation of lineages, it is important that they are captured by the neutral process. For example, in a study of DENV-2 in Nicaragua, the authors describe three sublineages which underwent lineage turnover from NI-1 to NI-2A and then to NI-2B (OhAinle et al. 2011). They also describe an apparent increase in relative fitness of NI-2B compared to NI-1. We compared these lineages to our standardized nomenclature and found that NI-1 corresponds mostly to DENV-2III_D.1.3 (and some to 2III_D.1), NI-2A to 2III_D.1, and NI-2B to 2III_D.1.1 (**Figure S4B**). We therefore capture the lineage replacement and the apparent phenotypic difference between NI-1 and NI-2B (going from 2III_D.1.3/2III_D.1 to 2III_D.1.1). Further, 2III_D.1.1 (NI-2B), while mostly sampled in Nicaragua, has a subclade which was sampled in Cuba and Costa Rica from 2019-2022 (circled in **Figure S4B**), highlighting the importance of a standardized naming system between countries. A more recent paper (Thongsripong et al. 2023) also described NI-3 sublineages, which correspond to 2III_D.1.2 in our system.

We attempted to explore potential phenotypic differences between lineages in a more systematic way by using antigenic distance data generated from different serotypes and genotypes in Thailand over a 20 year period (Katzelnick et al. 2021). We began by comparing within-classification pairwise antigenic distances by serotype (**Figure S5A**) and found that while there was a slight decrease between serotype, genotype, and major lineage, it was not significant or consistent. Indeed, DENV-1 had a gradual decrease in antigenic distance across all three levels, DENV-2 only decreased at the major lineage level, DENV-3 mostly decreased at the genotype level, and DENV-4 had no noticeable difference between classification levels. We then mapped the antigenic distances in 3D space (**Figure S5B and S5C**) and found that major lineages did not form distinct clusters. These results, however, may be complicated by not having a broad representation of the global major lineages. Therefore, we may expect to find more noticeable differences in pairwise antigenic distances among the serotype, genotype, and major lineage levels if this antigenic dataset also included more diverse viruses (e.g. lineages from the Americas). Our results could also indicate that antigenic distance is more complex than a simple phylogenetically clustered trait. Indeed, the original authors found that antigenic distance varied more over time than between other circulating clades (Katzelnick et al. 2021). Regardless, our lineage system still provides a way to identify distinct lineages that may have an epidemiological advantage, which will be essential for evaluating the impact of antigenic distance on the immune landscape and therefore real-world effectiveness of new dengue vaccines.

By capturing known phenotypic diversity, providing additional geographic resolution, and standardizing discussion of important lineages, the lineage system proposed here builds on the success of the currently used genotypes and provides a necessary tool for monitoring DENV as it continues to spread worldwide.

### Lineage assignment using Genome Detective

The previous sections of this paper discuss the process of lineage *designation* - the development of a standardized new lineages classification system based on expert opinion. In addition, there is *assignation*, the process of providing a lineage call to a new sequence, which should be possible to do easily by research groups and public health professionals globally. Here, we provide an option for lineage assignment - Genome Detective. In the supplementary information, we also provide alignments and phylogenies of representative sequences from each of the lineages so that the reader may also try their own methods.

The Genome Detective Platform (Vilkser et al. 2019) is a microbial bioinformatics software suite that includes a generic framework for phylogenetic subtyping tools, allowing the creation of subtyping tools for any virus species. Currently, Genome Detective includes subtyping tools for nineteen virus species, developed with subject matter experts globally (de Oliveira et al. 2005; Alcantara et al. 2009; Kroneman et al. 2011; Pineda-Peña et al. 2013; Faria et al. 2018; Cleemput et al. 2020; Fonseca et al. 2019). A Dengue virus subtyping tool was first developed in 2019 (Fonseca et al. 2019), and since version 4.0 it was updated to use the lineage designation scheme introduced in this work (https://www.genomedetective.com/app/typingtool/dengue/, see Methods and figures S8-10).

### System stability and assignment validation

After designing this system and showing that it is useful to describe existing genomic diversity and to standardize discussions of DENV evolution at a higher resolution than before, we performed a series of validation checks. A key element of any lineage system is that it is reliable, in regards to the sampling structure of the dataset and genome coverage. When testing both, we found that the proposed rules generate lineages which are stable with low coverage genomes, and with different subsamples.

It can be challenging to obtain high coverage DENV genomes as viral load tends to decrease rapidly after the short viremic phase that often occurs prior to sample collection (Guzman et al. 2010). Further, while capacity for whole genome sequencing is increasing globally, mostly due to the genomic capacity accrued by public health and research institutes during the SARS-CoV-2 pandemic, many groups will preferentially sequence only the E coding region as this is all that is required for genotyping, and it is faster than whole genome sequencing. We therefore simulated low coverage genomes from a subset of the dataset (n = 309) by replacing nucleotides with N’s in runs of 20 at different percentages from 90% to 10% in 10% intervals, and re-assigned them using the Genome Detective typing tool. We found that overall the assignment accuracy for serotypes, genotypes, and major and minor lineages was high, even with very low genome coverage (**Figure S6A**). The lowest assignment accuracy was 86% for minor lineages at 30% coverage, while all other levels of classification were above 90% accurate. We found the accuracy to be above 95% for with genome coverages above 70%.

We further tested the tool and the system by trimming whole genome sequences keeping only the E coding region and again assigned them using Genome Detective. Serotypes, major lineages, and minor lineages were all correctly assigned using E sequences only. Seven out of 309 had incorrect genotype assignments, which were all given “unassigned” but should have been DENV2 genotype I. This is therefore an accuracy of 98% for genotypes and 100% for other classification levels using only complete E sequences.

We also tested the impact of sampling artifacts on lineage designation by randomly subsampling the dataset ten times and rebuilding the trees to see if the same monophyletic clades emerged. We found that they generally did (**Figure S6B**), with DENV-1 having the highest mean of different clades (0.01, 95% CI: 0.0-0.034), and DENV-2 having no different clades in any of the subsamples. DENV-3 and −4 were in between with 0.004 (95% CI: 0.0-0.017) and 0.006 (95% CI: 0.0 - 0.023), respectively.

Each level of classification can therefore be reliably assigned using a freely available, user-friendly tool even with only E sequences and with very low coverage genomes. Our lineage system is also robust to the sampling bias inherent in almost any genomic dataset. This system is therefore robust and reliable for real DENV genomic datasets.

### Case studies with additional sequencing data

To test our proposed system on new real-world data, we created a series of case studies based on genomic data not included in the system design from different geographical regions.

#### Vietnam

Vietnam experiences a severe dengue burden, with an average of 90,000 cases reported each year (Hung et al. 2018), although this is an underestimate. The virus is hyperendemic in the southern part of the country, around the densely populated Ho Chi Minh City, and causes seasonal outbreaks in the north (Gibb et al. 2023). To test our proposed lineage system, we generated genome sequences (>70% coverage) from 596 stored DENV samples mostly from hospitals and clinics in Ho Chi Minh City and southern Vietnam (some samples from Hanoi) from 2010-2023 and detected patterns of new lineage introductions and patterns of lineage turnover (**Figure 5 and S7**).

**Figure 5.**
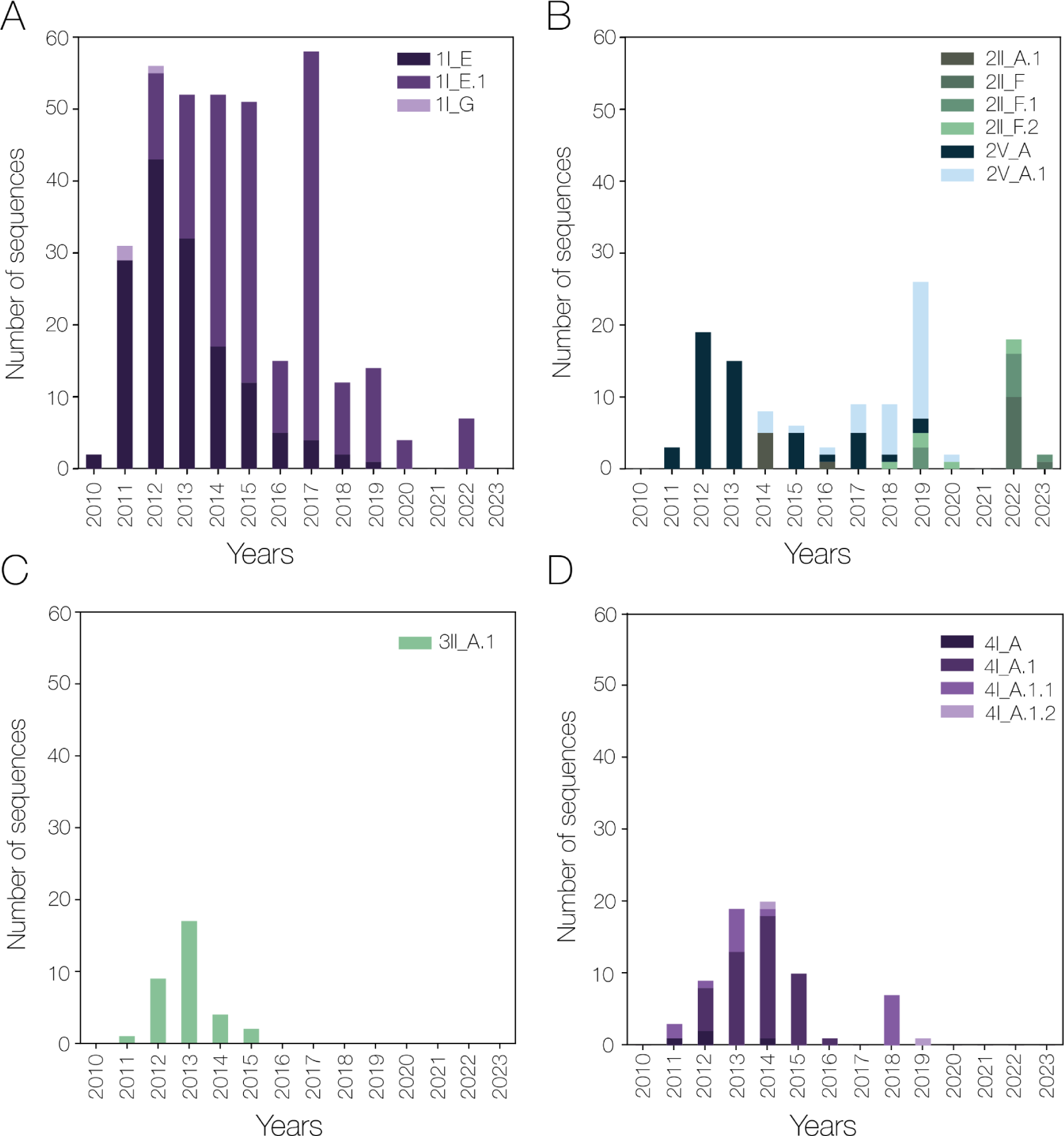
| Case study 1: Temporal dengue virus lineage distributions from Vietnam. Number of dengue virus whole genome sequences mostly from Ho Chi Minh City, Vietnam assigned to each lineage over time for A) DENV-1 B) DENV-2 C) DENV-3 and D) DENV-4.

We mostly found a single genotype in each of the four serotypes in this sample set. Further, most of the sequences for DENV-1 and all of the sequences for DENV-4 also fell into a single major lineage designation, and DENV-3 into a single minor lineage (**Figure 5**). However, increasingly nested minor lineages of DENV-1 and DENV-4 accumulated over our sampling time period (i.e. 1I_E.1 and 4I_A.1, 4I_A.1.1, and 4I_A.1.2). This implies that, rather than new successful introductions, the time period analyzed is dominated by continuous circulation and evolution of local DENV lineages, confirmed by estimating the phylogenies (**Figure S7**) and supported by other work (Ashall et al. 2023). It is also of note that the minor lineages 3II_A.1 and 4I_A.1 that we detected are currently only reported in Vietnam.

In comparison, DENV-2 was more diverse, with sequences falling into two major lineages in the same genotype (2II_A and 2II_F), as well as major lineage A in genotype V. Within genotype II, we stopped detecting 2II_A in 2016, with a year without any genotype II detection, and then a possible novel introduction of 2II_F which then becomes the only major lineage that we sampled in 2022 and 2023, as described in another study (Tran et al. 2023). We therefore can document patterns of lineage turnover within DENV-2 in Vietnam.

#### Rio Grande do Sul, Brazil

Dengue is a serious public health threat in Brazil, with 3 million cases reported in 2023, representing two thirds of all dengue cases reported in the Americas (World Health Organization 2023). DENV is also becoming more common in Brazil’s previously non-endemic areas, including in the far south in the state of Rio Grande do Sul, which experiences periodic outbreaks due to introductions from other regions in Brazil into its mostly immunologically naive population (Marques-Toledo et al. 2019). Understanding the difference between imports and sustained transmission in Rio Grande do Sul is therefore an important national public health question, as it provides information on the tipping points of local endemicity. We used 665 near full-length genomes of DENV-1 and DENV-2 from the region sampled from 2015-2023 to examine the utility of the lineage system for discerning importations vs. continued circulation.

We found that most of the genomic data set is dominated by four different major lineages of DENV-1 genotype V: A, D, E, and F (**Figure 6A and S7B**). We found that DENV-1V_A was prevalent early from 2015-2016, but that DENV-1V_D and DENV-1V_E were the most prevalent in 2021 and 2022 (**Figure 6A**). We detected the major lineage DENV-1V_F in April 2015, but did not detect it again until 2022. This is similar to wider DENV dynamics in Brazil where 1V_A circulated at a low frequency until 2022, with 1V_E in particular dominating 2021-2023 (**Figure 6B**).

**Figure 6.**
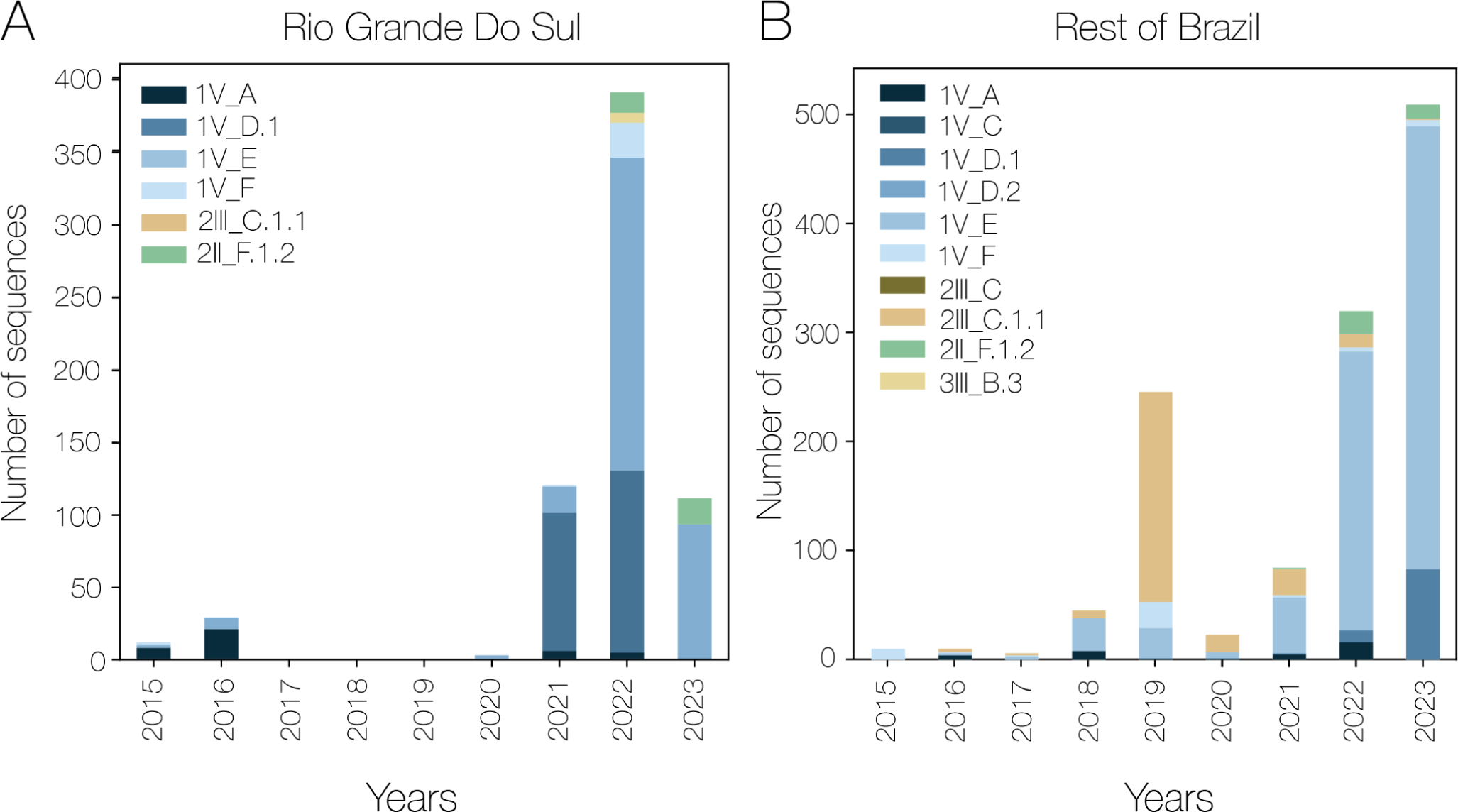
| Case study 2: Temporal dengue virus lineage distributions from Brazil. A) Time series of whole genome sequences from Rio Grande do Sul, Brazil by year. Note that not every year is present in the X-axis as there are no sequences from these years. B) Lineage assignments of whole genome sequences from the rest of Brazil in this dataset (non case study sequences from Rio Grande Do Sul have been removed).

In DENV-2, we identified two minor lineages from two different genotypes (2II_F.1.2 and 2III_C.1.1) which co-circulated for two months in 2022 (**Figure 6A**), before DENV-2II_F.1.2 took over and was the only DENV-2 lineage sequenced until the end of the dataset in March 2023. In the rest of Brazil (**Figure 6B**), DENV-2III_C.1.1 started circulating earlier, and was first detected in 2016, rising to prominence in 2019 and remained detected until 2023. There was therefore a small delay in the introduction of this lineage to Rio Grande Do Sul, and it was not reintroduced into the region after its extinction. 2II_F.1.2 was mostly sampled in 2022 and 2023, like in the region of interest. 2III_C.1.1 is only currently sampled in Brazil, separating it from its sister 2III_C.1.2 which is found in the Dominican Republic and Haiti. Further, we found the two parent lineages of 2II_F.1.2 (2II_F and 2II_F.1) in southeast and east Asia, and its sister (2II_F.1.1) only in Cambodia. Lineage 2II_F.1.2, however, is mostly found in South America, with sequences sampled in Peru and in Brazil.

The lineage dynamics in Rio Grande do Sul were similar to the rest of Brazil, with a lag in lineage frequency for DENV-2III_C.1.1. This suggests that the same lineages are being imported to this state from elsewhere in the country, rather than continued endemic circulation of the same lineages. This would of course need to be further evaluated with more robust phylodynamic analysis, but our lineage designation system provides a hypothesis that continued introductions of DENV from elsewhere in Brazil are currently the primary drivers of DENV transmission in Rio Grande do Sul.

#### Senegal

While dengue has a lower prevalence in Africa than in South America and Asia, there is under-reporting due to a general attribution of febrile disease to malaria (the “malaria umbrella”) (Amarasinghe et al. 2011). Hence, there is limited genomic or epidemiological data for DENV in Africa, and the transmission dynamics of DENV in Africa remain poorly understood. Due to this under-sampling, a new lineage system must be tested specifically with sequences from different countries in Africa to ensure that the diversity of the continent can be captured.

DENV has been known to circulate in Senegal since the 1970s (Robin et al. 1980), although it was mostly confined to the sylvatic cycle until the 2010s, when there were multiple urban outbreaks in 2017 and 2018 (Gaye et al. 2021; I. Dieng et al. 2021). Senegal is one of the 15 African countries reporting an outbreak in 2023 (World Health Organization 2023) and has strong sequencing capability. Therefore, we used 77 sequenced isolates of DENV-1, DENV-2, and DENV-3 from 2015 to 2023 from Senegal as an important test of our new lineage system.

All sequences from each serotype fell into a single lineage: DENV-1III_A, DENV-2II_B, and DENV-3III_B.2 (**Figure 7 and S7D**). Major lineage 1III_A is globally distributed, but is most commonly sequenced in China (**Figure 7A**). The 1III_A sequence from Senegal in 2023, however, is more recent than any in this major lineage in our data set. This highlights the importance of global sequencing and timely data sharing for finding continuing spread and circulation of existing lineages. Lineages 2II_B and 3III_B.2 are both detected in Africa and east/southeast Asia (**Figure 7B and C**). While strong conclusions about other African countries are difficult to draw, it is of note that 2II_B is mostly found in Burkina Faso, the African country with the most dengue cases reported in 2023 (World Health Organization 2023). Lineage 3III_B.2 is small (∼30 whole genomes in the original dataset) with most sequences sampled in Senegal (**Figure 7D**), and China being the only non-African country where it has been detected. Its sister lineages are currently found in Asia (3III_B.1) and America (3III_B.3), with the latter making up the large Cuban and subsequent Caribbean outbreaks (Taylor-Salmon et al. 2023).

**Figure 7.**
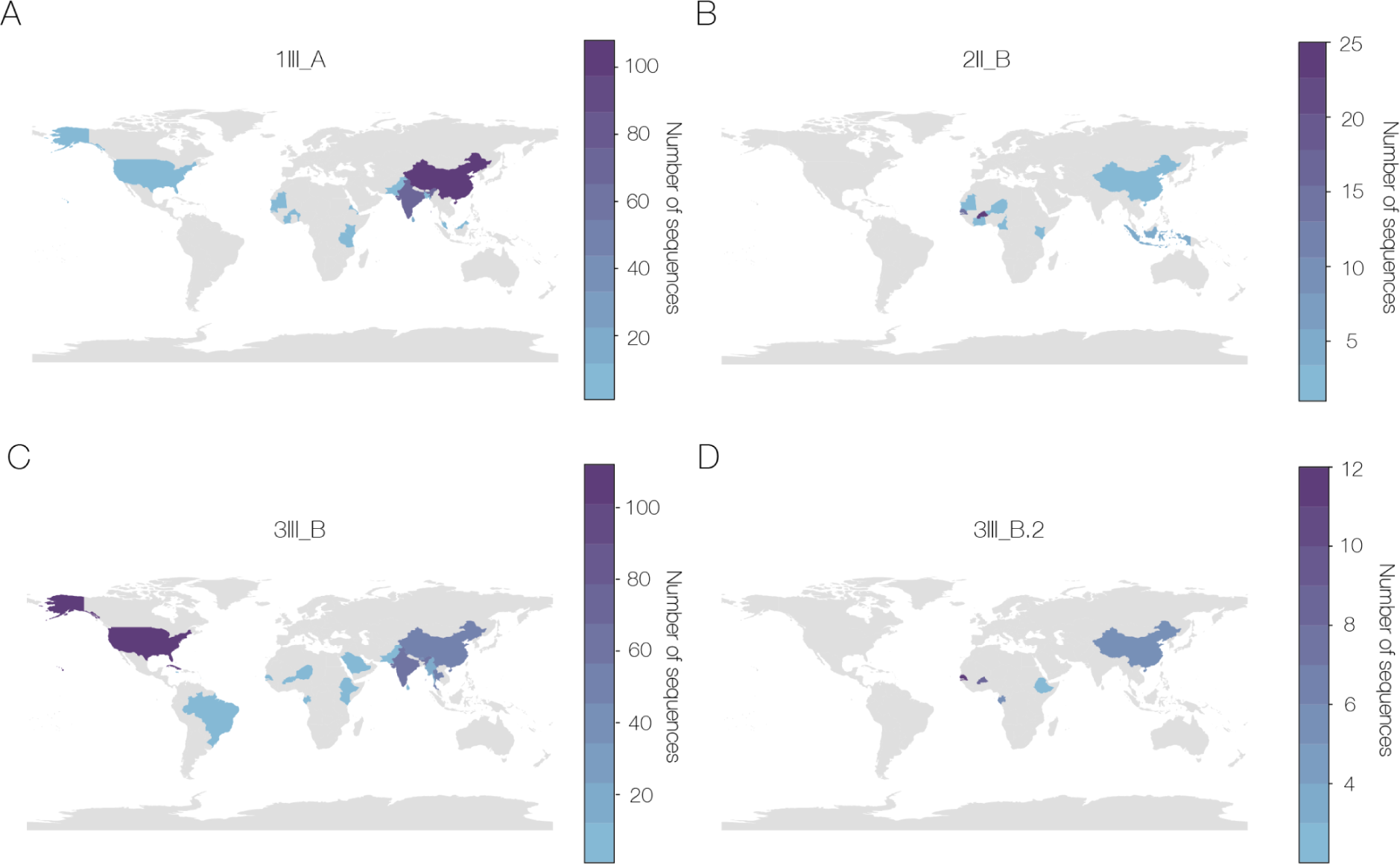
| Case study 3: Geographical dengue virus lineage distributions assigned to sequences from Senegal and Tanzania. A) Major lineage 1III_A which all DENV-1 sequences in these datasets are assigned to. B) Major lineage 2II_B which all Senegal DENV-2 sequences are assigned to. C) Major lineage 3III_B which all Tanzania DENV-3 sequences are assigned to. D) Minor lineage 3III_B.2 which all Senegal DENV-3 sequences are assigned to. All maps include all sublineages of each lineage, and colors show the number of whole genome sequences in the training dataset which are in each country or territory.

These sequences from Senegal fit well into our lineage system despite the initial training set containing far fewer sequences from Africa than from South America and Asia. By assigning major lineages, we reveal a stronger connection of DENV in Africa to Asia (particularly China) than to South America; as well as possible circulating lineages within Africa, although this needs to be confirmed by sequencing in more locations. Further, by assigning a minor lineage to the DENV-3 genomes, we provide evidence that the 2023 outbreak in Senegal is likely not connected epidemiologically to those in the Caribbean/Americas.

#### Dar es Salaam, Tanzania

In many situations, sequencing the DENV E coding region, as compared to whole genome sequencing, is common due to its relative affordability. As the lineage system we propose here was designed based solely on whole genome sequences, it is necessary to test it with E sequences as these are still being produced. Further, the relative lack of sequences from Africa (especially whole genome sequences) could lead to a section of DENV global genetic diversity which the current system misses. It is therefore important to test this system on E sequences from Africa to check its robustness.

Dengue outbreaks occur roughly every two years in Tanzania (Mustafa et al. 2023). While historically dengue cases have been low, this is likely at least partly due to under-diagnosis as mentioned above, and it is a growing concern especially in rural areas (Kajeguka et al. 2023). A previous paper characterized three outbreaks in Tanzania between 2017 and 2019, partially by generating 423 E sequences in these years (Kelly et al. 2023). We used this dataset to test our lineage system on a potentially unincluded source of genetic diversity.

The authors of the original paper found 32 DENV-3 sequences and 341 DENV-1 sequences, with none of the other serotypes circulating. We assigned all DENV-3 E sequences to DENV-3III_B with no minor lineage, 340 DENV-1 sequences to DENV-1III_A, and one DENV-1 sequence to DENV-1I_K.2 (**Figure S7C**). This last minor lineage is currently composed of only whole genomes from east Asia, mostly China, as does its sister lineage (1I_K.1). Their parent lineage 1I_K is also almost entirely detected in Asia, although there are sequences from Réunion and the Maldives, and more recently from Italy.

Major lineage 3III_B is global, while 1III_A is mainly found in Asia (**Figure 7A and C**), especially China. Therefore, Tanzania, at least in this time period, appears to be strongly connected to DENV circulation in Asia, especially from China and other parts of east and southeast Asia. Notably, these are the same major lineages which were sequenced in Senegal (discussed above), but without any minor lineage assignments. It is unknown if these assignments are sufficient, or are under-assigned due to missing defining substitutions from the rest of the genome. While we found that E sequences are accurately assigned using our lineage system, additional detail is possible with whole genome sequences which can, in this case for example, provide specific information on within-continental spread.

### Limitations and future sustainability

There are limitations to the system we have proposed here. First, there is likely unrecognized diversity due to global inequity in sequencing capacity, especially in Africa (**Figure S11A**). This unrecognized diversity also limits the conclusions we can draw with lineage assignments, as similar circulating lineages in undersampled regions may not be epidemiologically connected to each other, and may be independent introductions. This is compounded by our decision to use whole genomes to build the system and not explicitly include E coding region sequences, which tend to be older. Our lineages are therefore influenced by the number of whole genome sequences available (**Figure S11B**), and indeed the number of lineages is strongly correlated with the number of sequences on the country level (p<0.001, **Figure S11C**). We hope that as sequencing capacity continues to build and expand globally, this disparity will decrease, while still being able to define new lineages when new diversity is captured.

On a local level, our system does not always provide enough resolution for specific epidemiological questions - some minor lineages are still relatively widespread. This is due to trying to have fewer lineages at the start of the system implementation to ease discussion, and thereby help with uptake. We also note that the new system is not a 1:1 relationship with existing regionally defined lineages. This is partially because, in some cases, previous lineages were defined based on a single sequence or a small group of sequences, which does not meet our criteria for lineage designation; but may be helpful on a regional level for defining introductions. Therefore, this system does not replace local-level phylogenetic analysis, and simply provides a first, quick pass at describing the diversity in a dataset.

Any lineage system must be easy to maintain and have an intrinsic stability, especially as the DENV genomic data set continues to grow. We deliberately chose rules to define lineages which are computer-readable (and indeed initially designated them using custom scripts) to reduce the person-time required to generate new lineages.

We envision two main ways that new lineages could be suggested going forward. First, individual research teams can submit a github issue to a public github repository (information at https://dengue-lineages.org/) using a standard form. If accepted, these suggestions will be given a “putative” designation and the label claimed until they have been reviewed so that the study can proceed with relevant lineage information. These putative lineages will then be formally incorporated into the nomenclature system during an annual review process. The second method of new lineage designation will be during this annual review, where proposals for previously undesignated diversity would be generated using automated scripts. Alternatively, these proposals could be generated with an automated lineage designation tool such as recently described (McBroome et al. 2024). We did not use ‘autolin’ because our initial designation required some manual curation and integration into the current serotype/genotype system, but this will not be required in the future and so this tool may be useful. In other words, we would generate new minor lineages nested into existing minor and major lineages, as well as new major lineages or genotypes which may arise. We hope that these two methods of lineage proposal balance the need for up-to-date information for those conducting surveillance, without requiring a large amount of effort to maintain.

These new lineage proposals would then be submitted to a designated international committee for decision-making. At this stage, the lineage designation rules and thresholds would also be reviewed. We note in particular the phylogenetic distance threshold, which we selected in the interests of stability and keeping the number of lineages manageable. However, part of the reason these long branches exist is the relative undersampling of DENV in some geographic regions. In an ideal world with more routine DENV sequencing there would be fewer very long branches in the phylogeny, and we hope this may be the case going forwards. We would not, however, change thresholds for existing lineages so as not to change results of already completed work, or change the defining nodes of existing lineages or genotypes to ensure stability.

Finally, in the interests of usability and fitting better into existing work, we provided an assignment tool already used for subtyping DENV and other viruses - Genome Detective. We note that our system is open and we are supportive of the development of any other tools to assign sequences to these lineages. We believe this will also help with sustainability, as opposed to designing a new tool specifically for this lineage system which would also require maintenance.

## Conclusions

DENV is a globally important virus that causes high levels of morbidity each year. With increasing globalization and climate change, viruses can traverse continents with ease, leading to increased global genetic mixing. As worldwide genomic sequencing capacity increases, genomic epidemiology becomes a more powerful tool for understanding how DENV spreads and causes outbreaks. The recent influx of genomes must be categorized beyond existing genotypes to provide rapid and understandable epidemiological conclusions.

Here, we present a lineage system, drawing inspiration from the design and uses similar systems implemented for SARS-CoV-2 (Rambaut et al. 2020), rabies (Campbell et al. 2022) and mpox viruses (Happi et al. 2022). We propose major and minor lineages in addition to slightly adjusting the existing genotypes to capture current diversity. We ensure that these lineages are mostly stable despite phylogenetic uncertainty and low coverage genomes by having a high inferred substitution threshold for defining a lineage.

Our system assigns E-only sequences at least to major lineages and in some cases to minor lineages. This is important as many previous studies only include E sequences, and there are still approximately double the number of E sequences as whole genomes on GenBank. We still encourage the sequencing of whole genomes going forward as they contain more genetic information for phylogenetic placement and analyses, as well as requiring a shorter time period of sampling to accurately estimate rates of evolution (Dudas and Bedford 2019). Further, natural selection acts on the whole genome and so losing information on how the nonstructural proteins evolve may hamper efforts to design and monitor the effectiveness of potential antivirals and vaccines. For these reasons, while a lot of important conclusions can and have been obtained with E sequencing, we encourage, where possible, for the whole genome sequencing of DENV.

Our proposed lineage system provides additional resolution for the discussion of potentially important global DENV diversity, and provides a conceptual framework that could be extended to incorporate hierarchical lineage classifications for other arboviruses with broadly defined genotypes (e.g., chikungunya and West Nile viruses). As DENV continues to circulate, the volume of genomic data increases, and new interventions are rolled out that may lead to important viral adaptation, this will become an imperative. By creating stable lineages which work well in different dengue-endemic regions, our system has the potential to enhance DENV genomic surveillance and epidemiology across the globe (https://dengue-lineages.org/).

## Methods

### Data set generation

We downloaded all DENV sequences with more than 70% of the genome covered and a year of collection listed released on GenBank until the 28th July 2023 and released on GISAID between 1st January 2022 and 28th July 2023. We matched sequences between these two databases to remove duplicates to create a near-complete publicly available full whole genome dataset (DENV-1 = 5657, DENV-2 = 4106, DENV-3 = 2166, DENV-4 =1045). We then aligned all of the sequences by serotype using MAFFT v.7.490 (Katoh and Standley 2013) and manually curated it in Geneious v.2022.1.1, including trimming the untranslated regions (UTRs). We removed sequences which were extremely divergent and those with frame-breaking insertions (DENV-1 = 2, DENV-2 = 1, DENV-3 = 1).

We then inferred first pass maximum likelihood trees by serotype using IQTree v2.1.4 (Minh et al. 2020). We used the program’s model selector on the smallest dataset (DENV-4) to gauge the best nucleotide substitution model to use, and it returned a transition model with empirical base frequencies and a free rate model with six categories (TIM +F+R6). We then rooted this tree using a molecular clock assumption in TempEST (Rambaut et al. 2016) and a heuristic residual mean squared model. We used the root-to-tip plot produced by this rooted tree to prune molecular clock outliers, a reliable indicator of quality control issues (V. Hill and Baele 2019). Most of the outliers that we found here were resequencing of commonly used virus stocks.

The final dataset sizes used for developing the lineage system were DENV-1 = 5456, DENV-2 = 3932, DENV-3 = 2103, and DENV-4 = 990. We assigned each of these sequences to existing genotypes using Genome Detective Dengue Virus Typing tool (Fonseca et al. 2019).

### Lineage system design

Our aims in the design of this system were to break up large clades in the genotypes to provide sufficient resolution to capture epidemiologically relevant patterns; but, drawing on our experience from SARS-CoV-2 nomenclature, not to have many fine-scaled lineages which are hard to discuss, and require regular updating.

With the above datasets, we inferred a new maximum likelihood tree for each serotype separately using the TIM+F+R6 nucleotide substitution model as suggested by IQTree’s model finder.

We separated clades using custom pre-order tree traversal scripts using the criteria of (**1**) 15 sequences, (**2**) a branch length of 25 substitutions, and (**3**) presence of a sister lineage. Branch length was calculated by multiplying the IQTree divergence length by the relevant alignment length. The length and size thresholds were obtained by testing different combinations to obtain an optimal level of resolution where major clades were split up, but no more than two sublevels of minor lineages were present. Major and minor lineages were assigned using the same rules and at the same time but were given different nomenclature.

At this point, we performed a second designation step to ensure that as many potentially epidemiologically significant lineages as possible have been captured by at least a major lineage. The aim here was to avoid a situation where a lineage, possibly from a country or region which is undersampled, could cause an outbreak in the near future without having any assignment beyond a genotype. To do so, we found all clusters which contained only sequences sampled after 2000 without a major lineage designation, and designated further major lineages using a more generous threshold of a minimum size of 5 sequences and a minimum branch length of 10 substitutions. Any clusters that didn’t meet these criteria, even those with sequences after 2000, were left unassigned, as they would not be very stable between tree building iterations if they did not meet the substitution threshold.

Finally, we performed some manual curation. As the current dataset sizes between serotypes vary by a factor of 5, rules which work well for DENV-3 and DENV-4 can lead to a high level of nesting in DENV-1 and DENV-2. For DENV-1, we moved the defining node for 1V_B closer to the present to enable us to maintain the strict hierarchy of the lineages and add up to major lineage G to break up the diversity. We also did this for 1I_A and added up lineages up to K, 1VI_A to add a sister lineage B (this was a lineage generated by the second designation step where sister lineages were not mandatory), 2II_B to add lineages up to G, and 2III_B to add lineages C and D. It is worth noting that for all of the manual changes we made, a lineage never has fewer than ten substitutions along the branch before it.

### Generating representative trees

For the assignment tools, it was necessary to make representative trees and alignments of each of the levels of designation. To do this, we used the phylogenetic distance matrix, calculated using the python package DendroPy (Sukumaran and Holder 2010), which gives pairwise phylogenetic distance between each pair of sequences. We took five sequences with coverage of over 90% which were furthest apart from each other in each of the genotypes, major lineages, and minor lineages.

It was also important to ensure that the most basal sequence of each clade was present, otherwise other basal sequences may be under-assigned. We did this first by checking if the lineage-defining node had an immediate descendent node which was a tip, and this node was added to the representative dataset. If there was not a node which was a tip, the sequence in the lineage with the lowest distance to the root (i.e. fewest SNPs) was included.

Alignments were generated, and trees were obtained by pruning them from the larger trees using jclusterfunk (https://github.com/snake-flu/jclusterfunk).

### Genome Detective

The subtyping tool for genome detective (https://www.genomedetective.com/app/typingtool/dengue/) takes a fasta file or sequences as a text input (**Figure S8)** and has two main steps to assign a lineage to a sequence.

The first step is species identification, which for the dengue virus subtyping tool is to identify the serotype. Basic Local Alignment Search Tool (BLAST) N and BLASTX are used to identify a maximum of three potential hits. For each of these potential hits we then use the Advanced Genome Aligner (AGA) (Deforche 2017) to align the query against the reference, and compute the overlap and concordance score. Finally, we select the reference with the highest product of overlap and concordance score. If this species corresponds to the species for which the tool was developed, we proceed to the clade identification. In practice, the species identification step will sometimes identify at a taxonomic level deeper than species, if there is a reference sequence in NCBI RefSeq for this deeper level. In particular, for the dengue virus subtyping tool this first step will identify the serotype.

Once the serotype has been identified, maximum likelihood phylogenies containing the query sequence and representative sequences for each lineage are constructed to identify the clade which is most likely to contain the query. The most recent dengue subtyping tool uses IQ-TREE 2 for its analysis, a change from previous versions which used PAUP* (Swofford 1993). This is because the former is not a pure hill-climbing algorithm and so is less likely to get stuck in a local optimum. Every clade we wish to identify is defined by approximately five representative sequences (see above). We compare the bootstrap value of the node which contains some or all of the representative sequences and the query, and identify the most likely cluster. Then, we compare the support values for nodes containing only the representative sequences (outer support) against the representative group plus the query (inner support). This allows us to estimate how likely it is that the query sequence falls inside the cluster. If the bootstrap support for the most likely cluster is lower than 50%, or the most likely cluster is the outgroup, the sequence cannot be assigned. If the bootstrap support for the most likely cluster is higher than 50%, but the inner support is not significantly higher than the outer support, the sequence is assigned as “related to, but not part of” the cluster. If the bootstrap support is higher than 50%, and the inner support is significantly higher than the outer support, the sequence is assigned to the most likely cluster.

Hierarchical clade identification is a stepwise process. Once a clade is identified, the process will reiterate to identify the subclade, if any subclades exist. In other words, once the genotype (e.g., 3II) has been identified, the tool will proceed to try assigning the major lineage (e.g., 3II_A), and once this has been assigned, it will continue to the minor lineage (e.g., 3II_A.1), and then to further minor lineages (e.g., 3 II_A.1.2). Unlike previously developed subtyping tools, which could only identify clade and subclade, the current dengue tool can support an arbitrary amount of assignment levels. This is necessary to enable an evolving lineage system particularly useful when deeper levels of minor lineage are possible. Outputs are a live-updating page with lineage assignments (**Figure S9**), and on clicking the sequence ID, details of the phylogenetic analysis which lead to the lineage assignment (**Figure S10**).

### Case studies

The Institutional Review Boards (IRB) from the Yale University Human Research Protection Program determined that pathogen genomic sequencing of de-identified remnant diagnostic samples as conducted in this study is not research involving human subjects (Yale IRB Protocol ID: 2000033281).

Clinical specimens from Vietnam were sampled mostly from Southern Vietnam between 2010 and 2023. They were sequenced at the Yale School of Public Health using the recently developed amplicon sequencing DengueSeq protocol (Vogels et al. 2024). Libraries were prepared using the Illumina COVIDSeq test (RUO version) with the pan-serotype primer pools, and sequenced on the Illumina NovaSeq 6000 or X Plus (paired-end 150) at the Yale Center for Genome Analysis. Consensus sequences were generated by mapping reads to dengue reference genomes through the DengueSeq bioinformatics pipeline using default settings (minimum frequency threshold of 0.75 and minimum depth of 10 to call consensus).

The sequences from Senegal were generated using both Nanopore et Illumina sequencing technologies. Details can be found here (Idrissa Dieng et al. 2022, 2024). Methods for generating sequences from Brazil can be found here (Bermann et al. 2024) and from Tanzania can be found here (Kelly et al. 2023).

To assign lineages to each of the case study datasets, we used the representative sequences described above. We always split up serotypes for each analysis. We aligned new data with representative sequences using MAFFT v7.490, and then built a maximum likelihood tree using IQTree using the representative tree as a constraints tree and the TIM+F+R6 substitution model, as above. We rooted trees using the same roots as we inferred using the molecular clock assumption as above.

Finally, we annotated the resultant rooted trees using custom scripts to find the lineage-defining nodes. Lineages were assigned to new sequences based on them being descendents of these lineage-defining nodes.

### Validation tests

For the completeness analysis, we took a subset of the full dataset (n = 309) evenly sampled from different lineages and serotypes. We replaced bases with Ns in runs of 20 to replicate how low coverage genomes look using next generation sequencing techniques. We therefore tested genomes which we artificially lowered from 10% to 90% coverage in 10% intervals. We also artificially cropped the whole genome sequences to only the E coding region. We assigned these sequences using the genome detective dengue subtyping tool.

For the stability analysis, we built trees with 10 different subsamples of the data, and assessed whether the same lineages would be designated using the algorithm developed here using the same custom python scripts.

### Connecting new lineages to currently used lineages

For DENV-2 lineages from Brazil denoted BR1-4 (Drumond et al. 2013; de Carvalho Marques et al. 2023) and NI-3 from Nicaragua (Thongsripong et al. 2023), we were able to use tip labels on tree figures in the papers to compare to assigned sequences in the new system. For NI-1 and NI-2 lineages, sampled in Nicaragua (OhAinle et al. 2011), we used lineage defining amino acid substitutions (table 2 in the referenced paper). We took the sequences from the paper, identified substitutions at the relevant position, and put them in sets based on having the same amino acid substitution in common.

## Data Availability

All sequences used to design the lineage system are from Genbank and GISAID, accession numbers in Table S2. Custom scripts and alignments of representative sequences from Genbank can be found on our github (https://github.com/DENV-lineages/lineages-paper).
Sequences for the Vietnam case study can be found on Genbank under accession numbers PP269455-PP270050, in bioproject PRJNA1072696. For the case study from Tanzania, sequences were drawn from (Kelly et al. 2023), and can be found on Genbank under accession numbers OM920035-OM920066 for DENV-3 and OM920075-OM920415 for DENV-1.
Sequences for the Brazil and Senegal case studies are currently private awaiting separate publications, but are available on request from, respectively.

https://github.com/DENV-lineages/lineages-paper

## Data availability

All sequences used to design the lineage system are from Genbank and GISAID, accession numbers in **Table S2**. Custom scripts and alignments of representative sequences from Genbank can be found on our github (https://github.com/DENV-lineages/lineages-paper).

Sequences for the Vietnam case study can be found on Genbank under accession numbers PP269455-PP270050, in bioproject PRJNA1072696. For the case study from Tanzania, sequences were drawn from (Kelly et al. 2023), and can be found on Genbank under accession numbers OM920035-OM920066 for DENV-3 and OM920075-OM920415 for DENV-1.

Sequences for the Brazil and Senegal case studies are currently private awaiting separate publications, but are available on request from, respectively.

## Acknowledgements

We thank the participants of Pan-American Dengue Conference 2023, members of the American Committee on Arthropod-Borne Viruses and Zoonotic Viruses (ACAV) at the American Society of Tropical Medicine and Hygiene (ASTMH) meeting 2023, and participants of the Pan American Health Organization arboviral genomic surveillance meeting in Puerto Rico 2024 for their feedback on this system. We also thank Dr Gilberto Santiago, Dr Maria Kelly, and Dr Jorge Munoz for their feedback. We thank Dr Angkana Huang, Dr Lin Wan, Dr Henrik Salje and Dr Derek Cummings for their help with the antigenic data analysis. We thank all of the groups over the years that have made their data available on Genbank and GISAID, without which projects like this would not be possible. An acknowledgements table for GISAID sequences can be found in **Table S4**. This publication was made possible by the National Institute of Allergy and Infectious Diseases of the National Institutes of Health (NIH) under Award Number DP2AI176740 (NDG), CTSA Grant Number UL1 TR001863 from the National Center for Advancing Translational Science (NCATS), a component of the NIH (CBFV), NIH Award Number 1 U01 AI151807-01 (KAH), Fundação de Amparo à Pesquisa do Estado do Rio Grande do Sul (FAPERGS/FIOCRUZ 13/2022 – REDE SAÚDE-RS, grant process 23/2551-0000510-7 and FAPERGS 14/2022 - ARD/ARC, grant process 23/2551-0000852-1), Conselho Nacional de Desenvolvimento Científico e Tecnológico (CNPq) through their productivity research fellowships (307209/2023-7; GLW), European Union’s Horizon 2020 research and innovation programme under grant agreement No 101000570, and the Medical Research Council-São Paulo Research Foundation (FAPESP) CADDE partnership award (MR/S0195/1 and FAPESP 18/14389-0). The findings and conclusions in this report are those of the author(s) and do not necessarily represent the official position of the NIH.

## Supplementary information

**Table S1** | *Roman numeral equivalents for geographical names of DENV-2 genotypes*
| Geographical name of genotype | Roman numeral of genotype |
| --- | --- |
| American | I |
| Cosmopolitan | II |
| Southern Asian-American | III |
| Asian II | IV |
| Asian I | V |
| Sylvatic | VI |

**Table S1** | Roman numeral equivalents for geographical names of DENV-2 genotypes

**Table S2** | Genbank and GISAID accession numbers of sequences used with assignments

**Table S3** | Information on each of the major and minor lineages

**Table S4** | GISAID acknowledgements table

**Figure S1.**
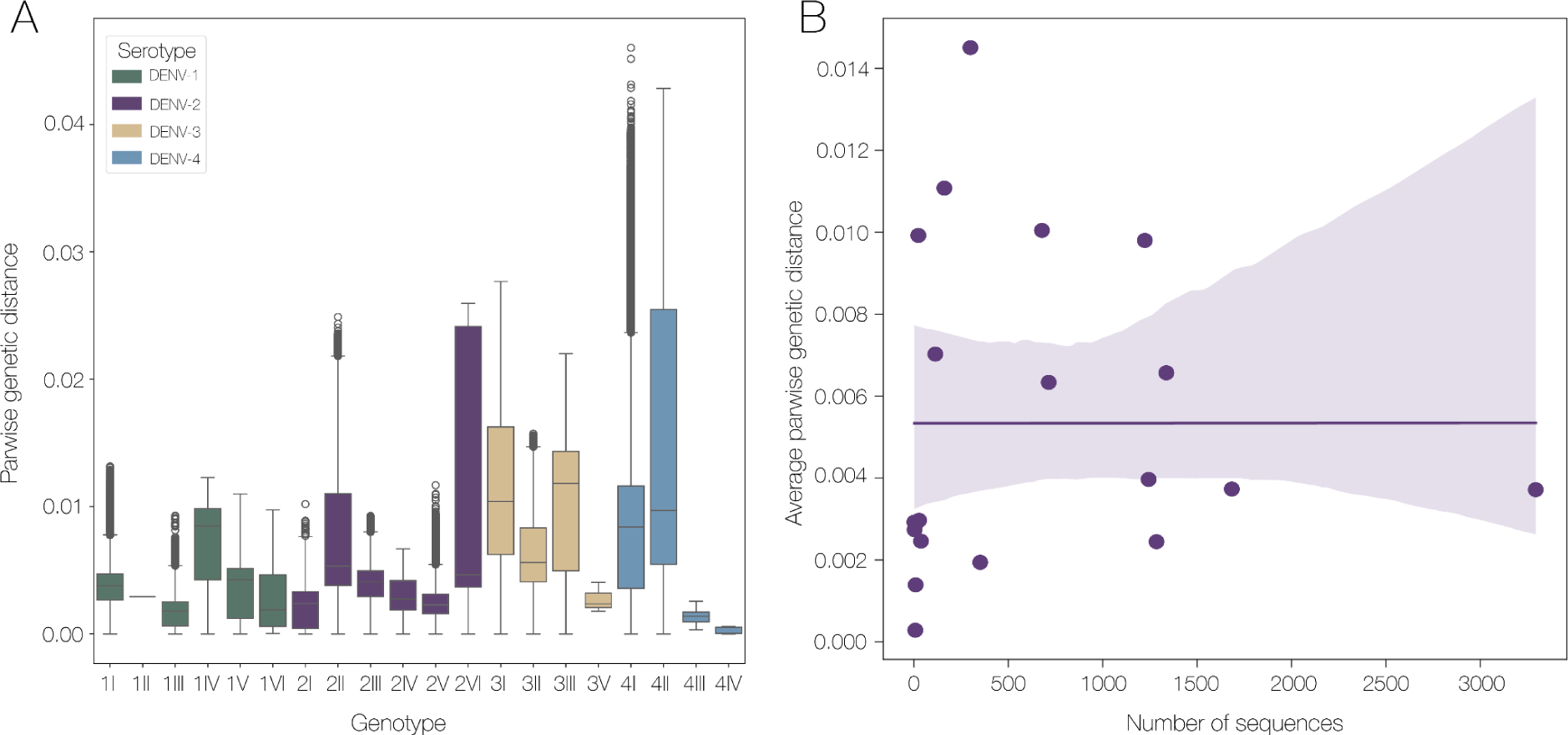
| Pairwise genetic distance of new genotypes A) Distribution of pairwise genetic distance within each genotype, colored by serotype. B) Regression of the number of sequences in each genotype compared to the average pairwise genetic distance.

**Figure S2.**
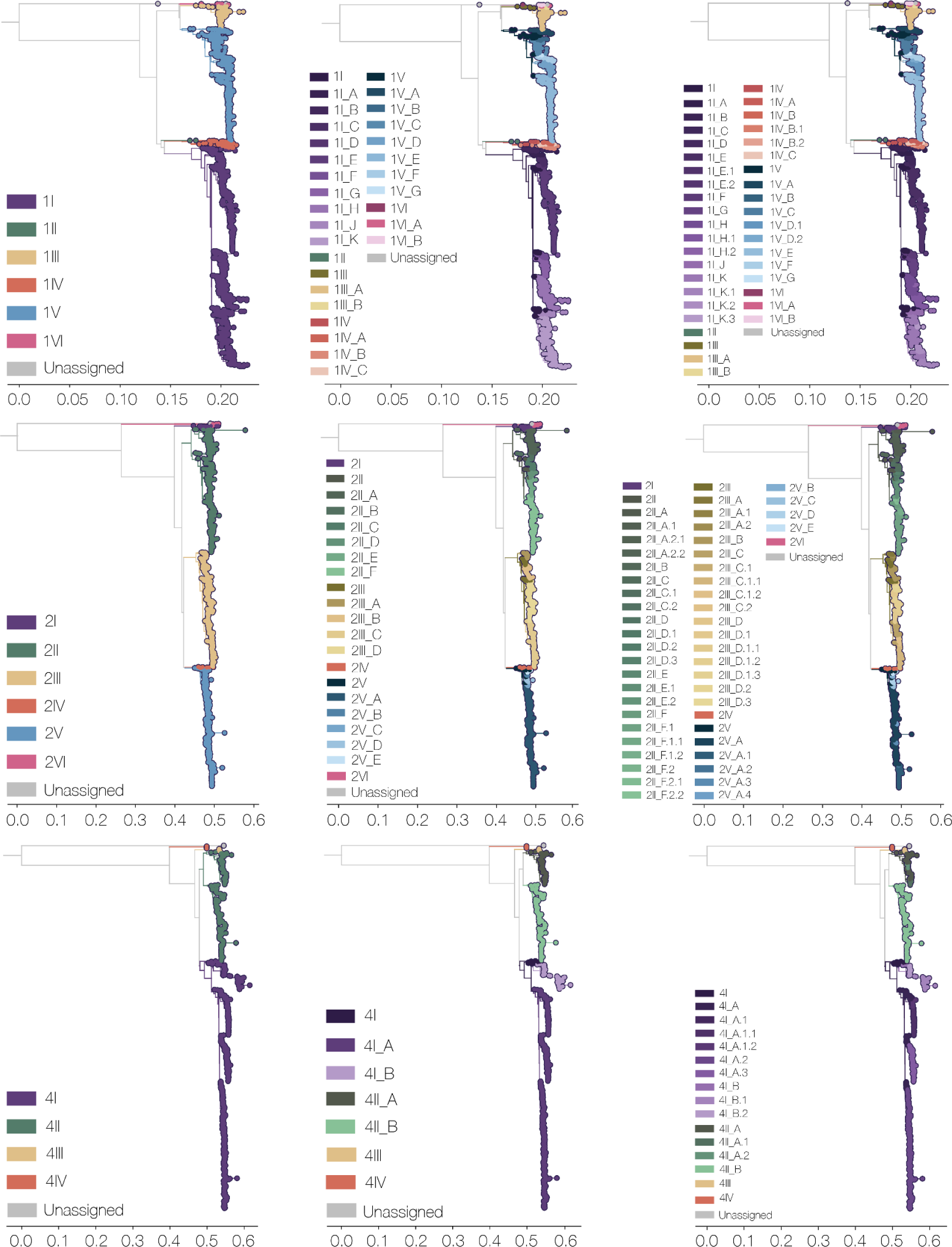
| New lineage system for serotypes 1, 2, and 4. Each row is a serotype and each column, respectively, is new genotype, major lineage and minor lineage. Note that there are many more minor lineages for serotypes 1 and 2, as they have much larger datasets currently compared to serotype 4. Serotype 3 shown in **Figure 2**.

**Figure S3.**
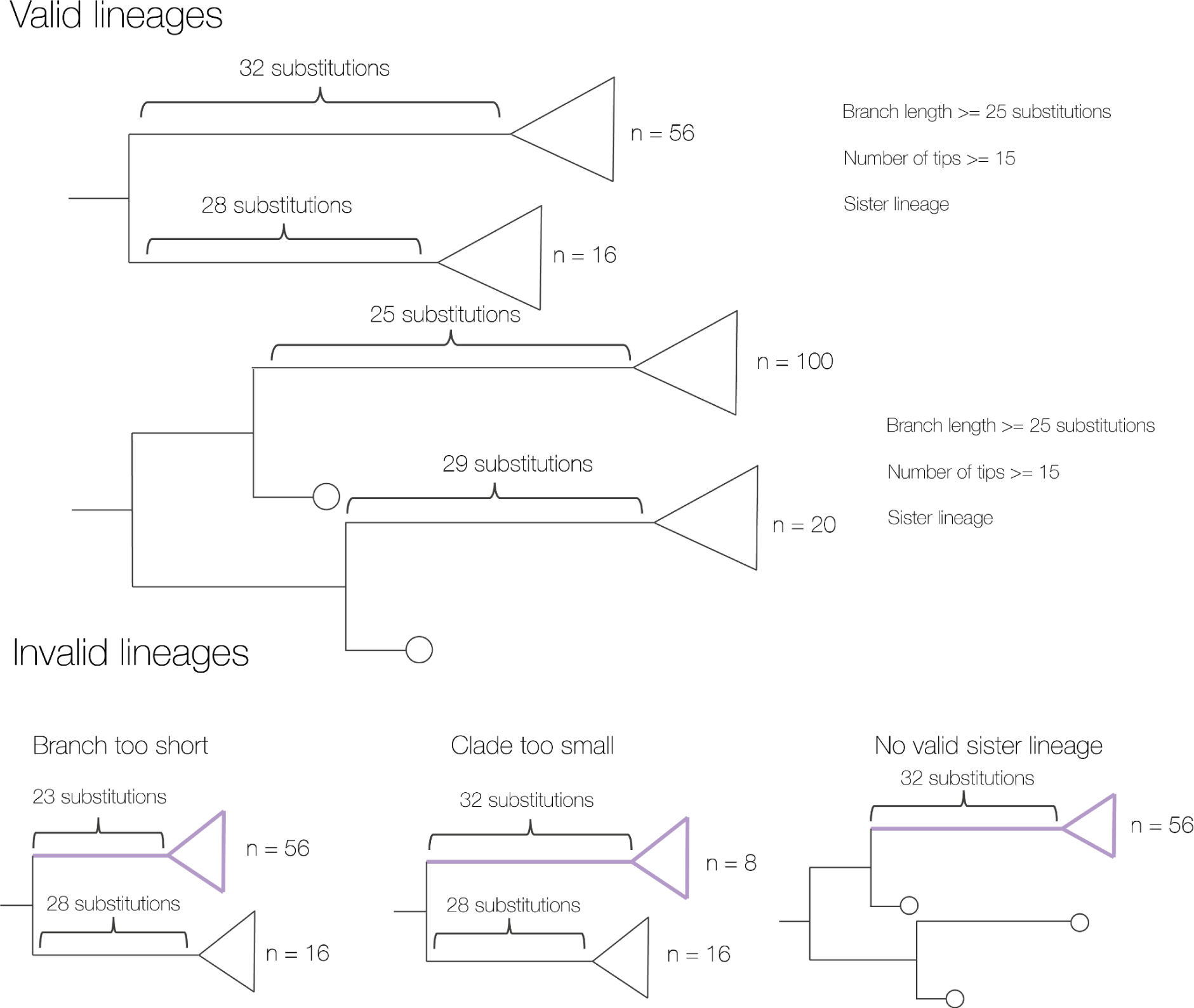
| Schematic displaying assigning new lineages. All four putative lineages displayed in the top two trees are valid lineages as they meet all three criteria of branch length, clade size, and having a sister lineage at the same distance from the root in terms of node number. Invalid lineages are shown along the bottom, with the focal putative lineage shown in purple.

**Figure S4.**
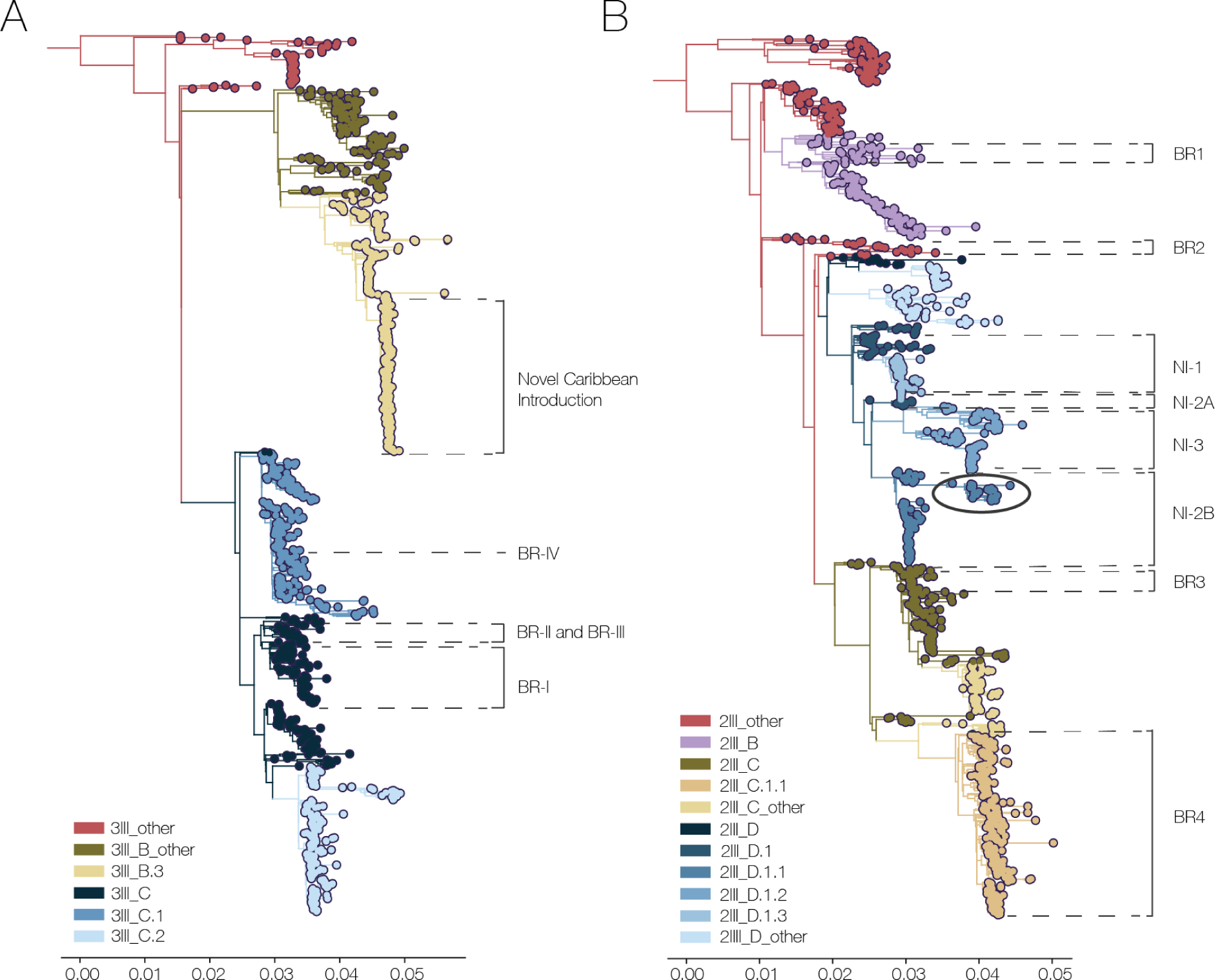
| Comparison of previously used sublineages to proposed system. A) Maximum likelihood tree of DENV-3 genotype III, colored by new lineage designation, with lineages BRI-IV and novel Caribbean introduction indicated. B) Maximum likelihood tree of DENV-2 genotype III, colored by new lineage designation, with lineages BR1-4 and NI-1 to NI-3 indicated. The circled clade indicates recent circulation of NI-2B/2III_D.1.1, which is suggested to have a transmission advantage (OhAinle et al. 2011), in Cuba and Puerto Rico.

**Figure S5.**
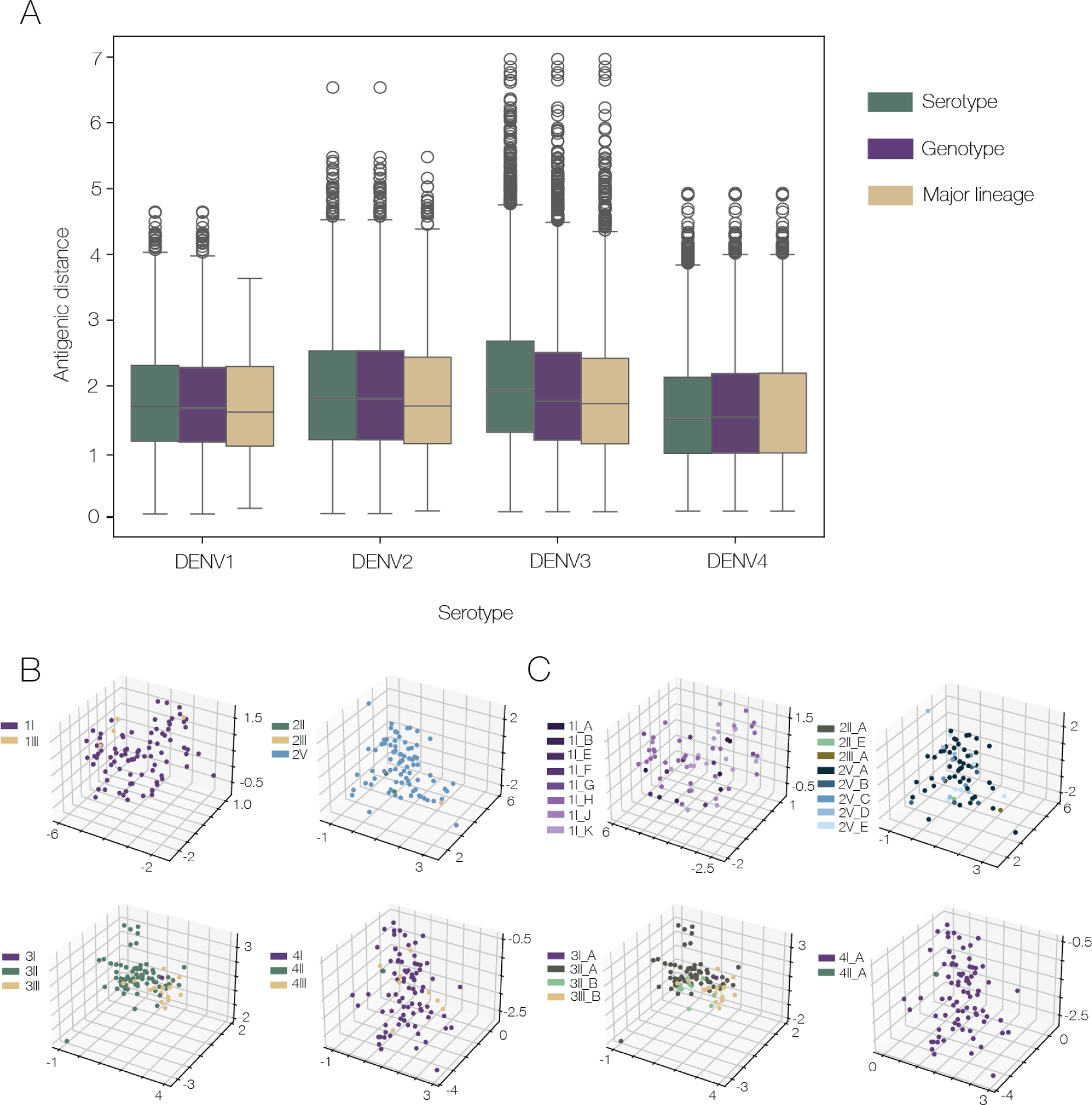
| Antigenic distance at different levels of classification. A) Distribution of antigenic distance in each level of classification, with serotype in green, genotype in purple and major lineage in yellow. Minor lineage is excluded due to a lack of antigenic data across minor lineages. B) 3-dimensional map of sequences in antigenic space by serotype, coloured by genotype. C) 3-dimensional map of sequences in antigenic space by serotype, coloured by major lineage

**Figure S6.**
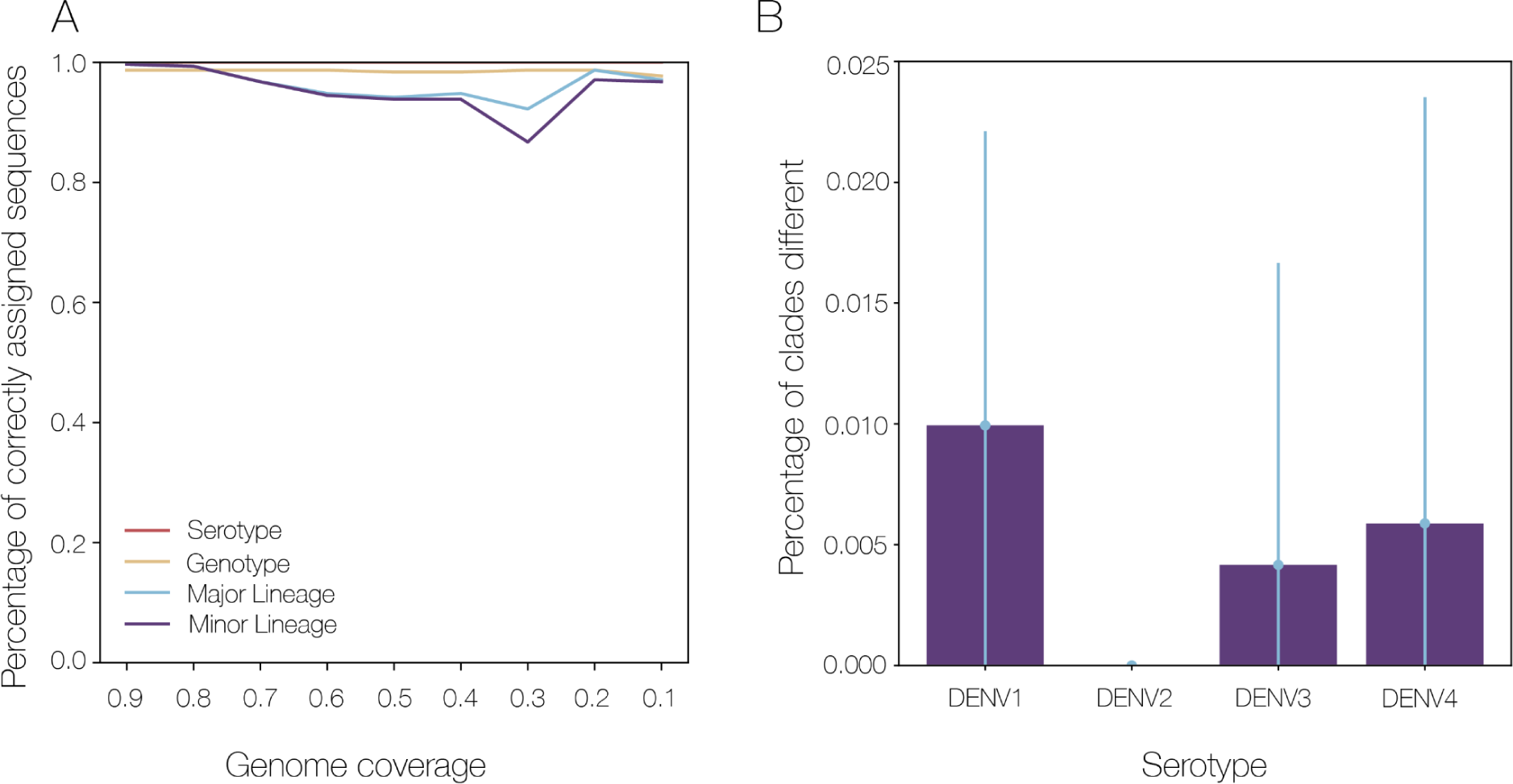
| Validation. A) Correctness of genome detective assignments at different classification levels using artificially downsampled sequences. Each line corresponds to a different classification level. B) Assessment of clade stability compared to different subsamples of the sequence dataset.

**Figure S7.**
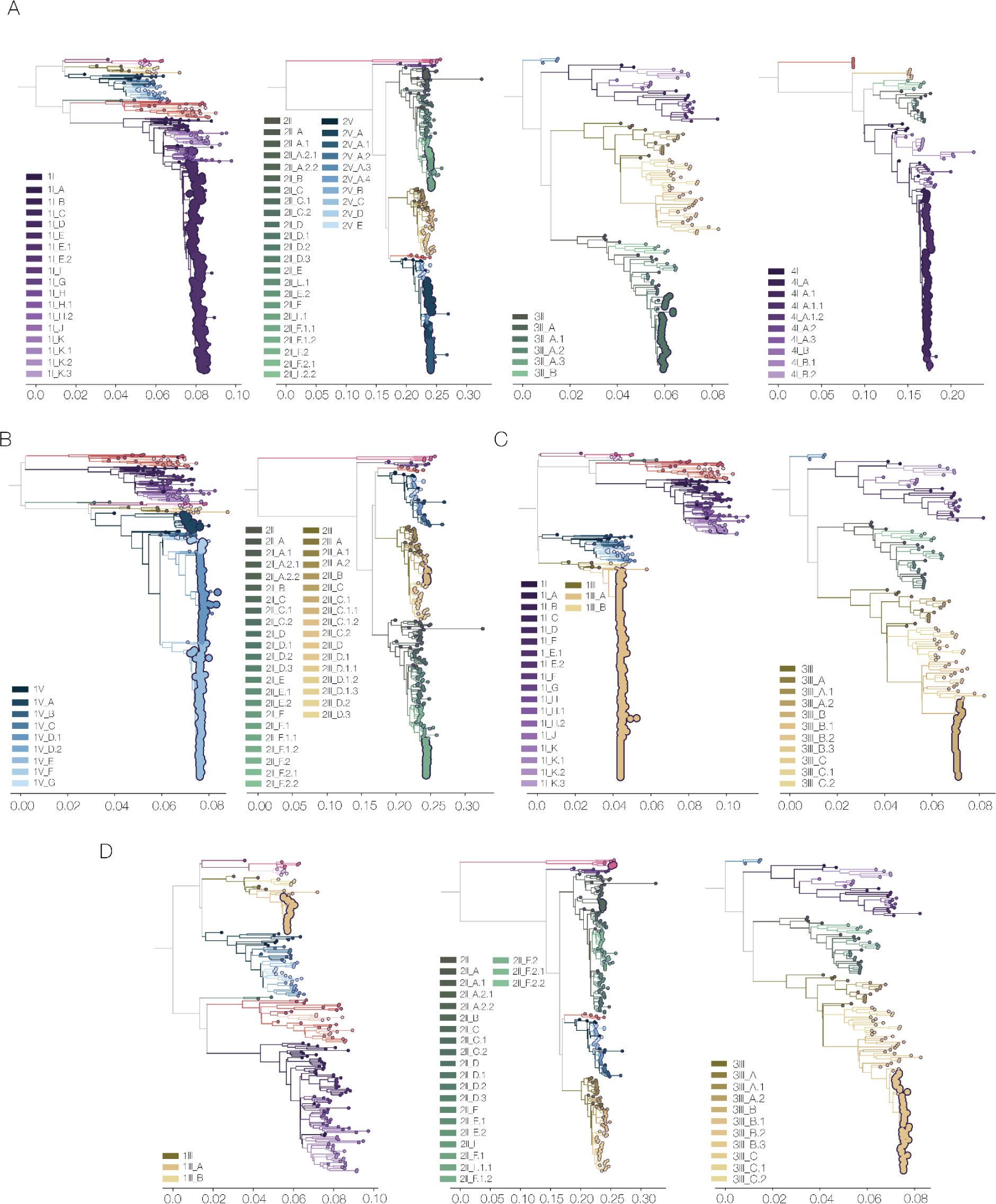
| Phylogenies of lineage assignments for case studies. A) DENV-1-4 whole genome sequences from Vietnam, time series 2010-2023. B) DENV-1 and DENV-2 whole genome sequences from Brazil, time series from 2015-2023. C) DENV-1 and DENV-3 E sequences from Tanzania. D) DENV-1, DENV-2, and DENV-3 whole genome sequences from Senegal

**Figure S8.**
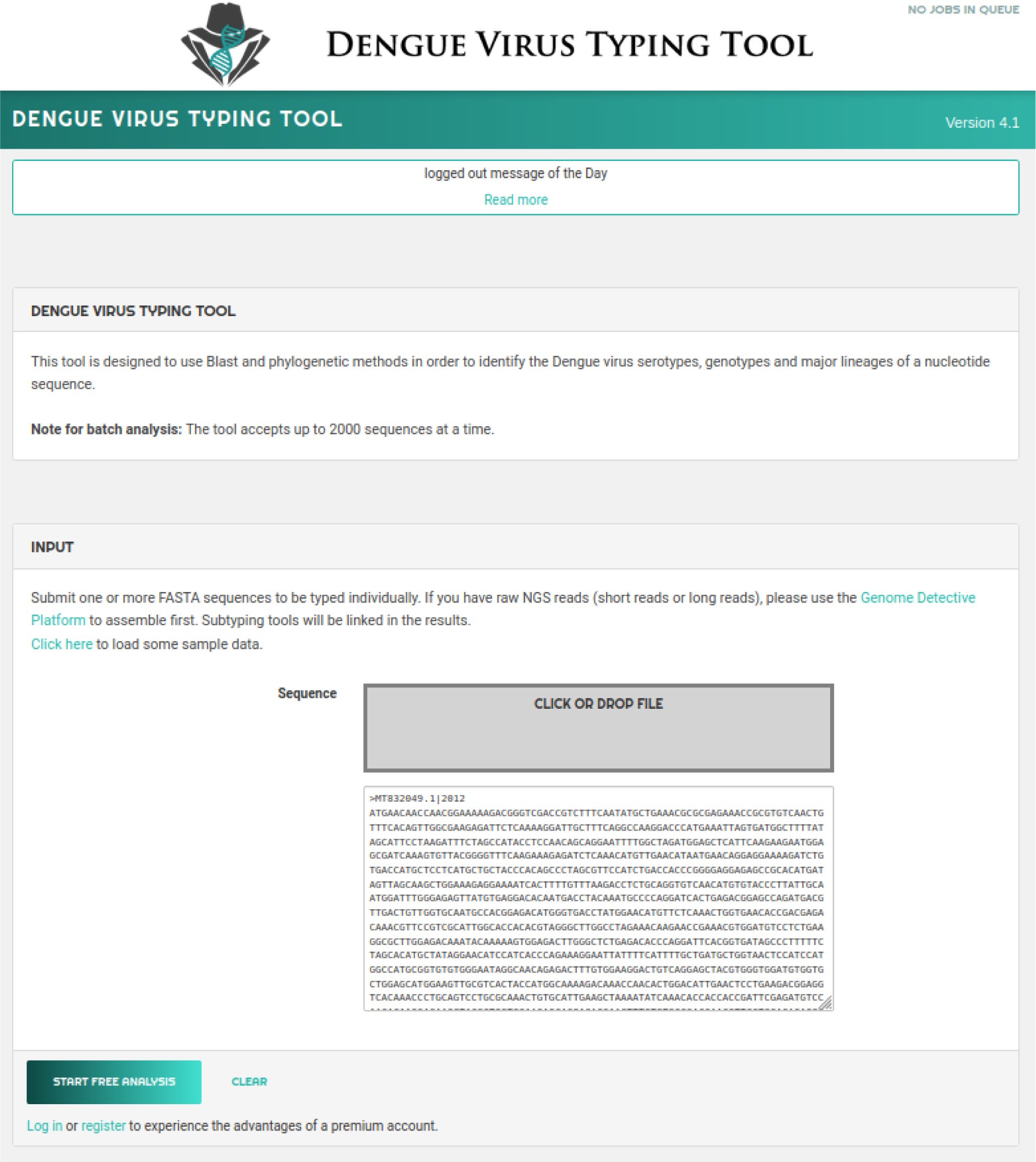
| Genome Detective dengue subtyping tool starting page. The user can either upload a .fasta file, or manually add sequences in the text field.

**Figure S9.**
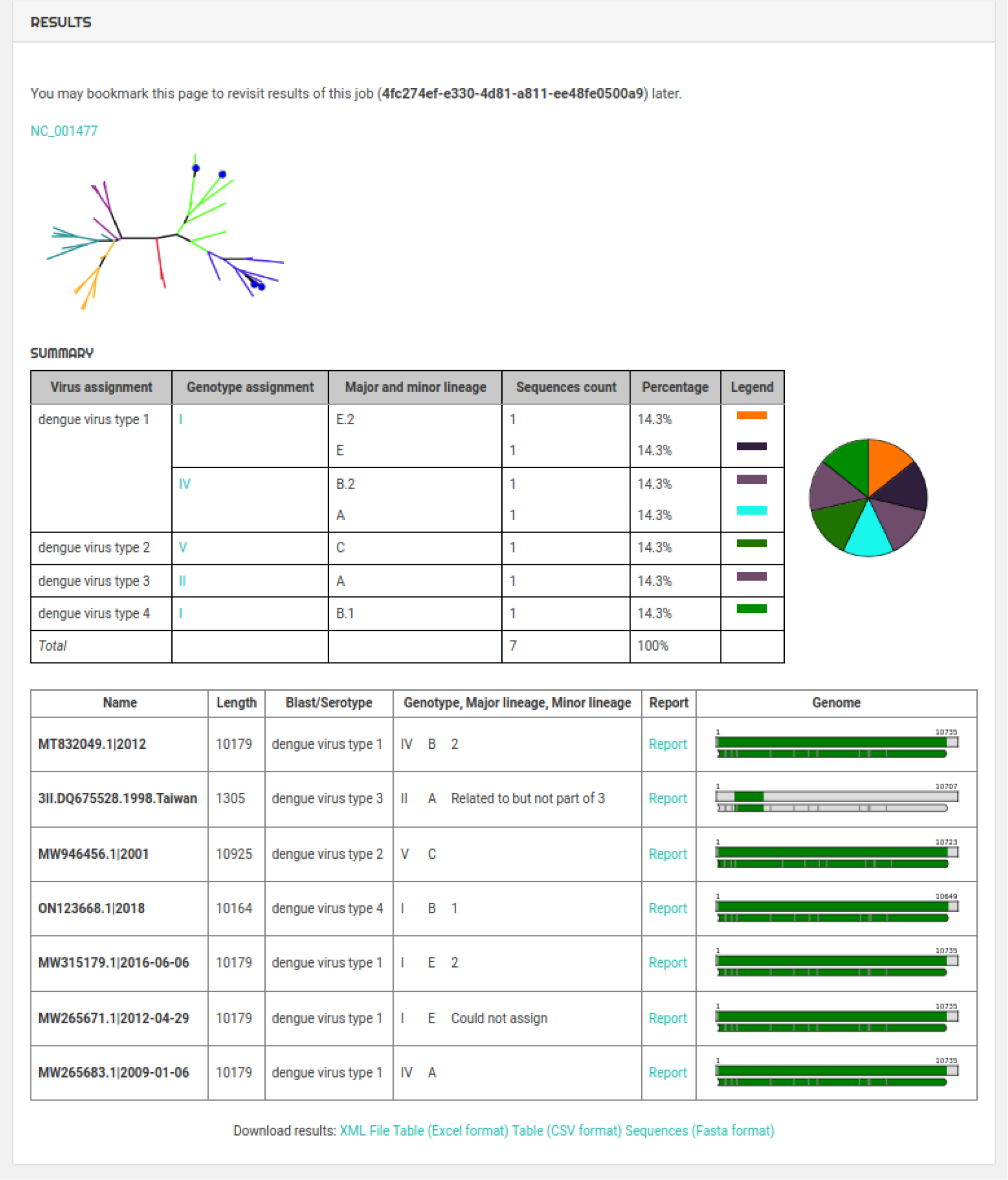
| Genome Detective dengue subtyping tool results overview. The results overview shows a short summary of the different assignments

**Figure S10.**
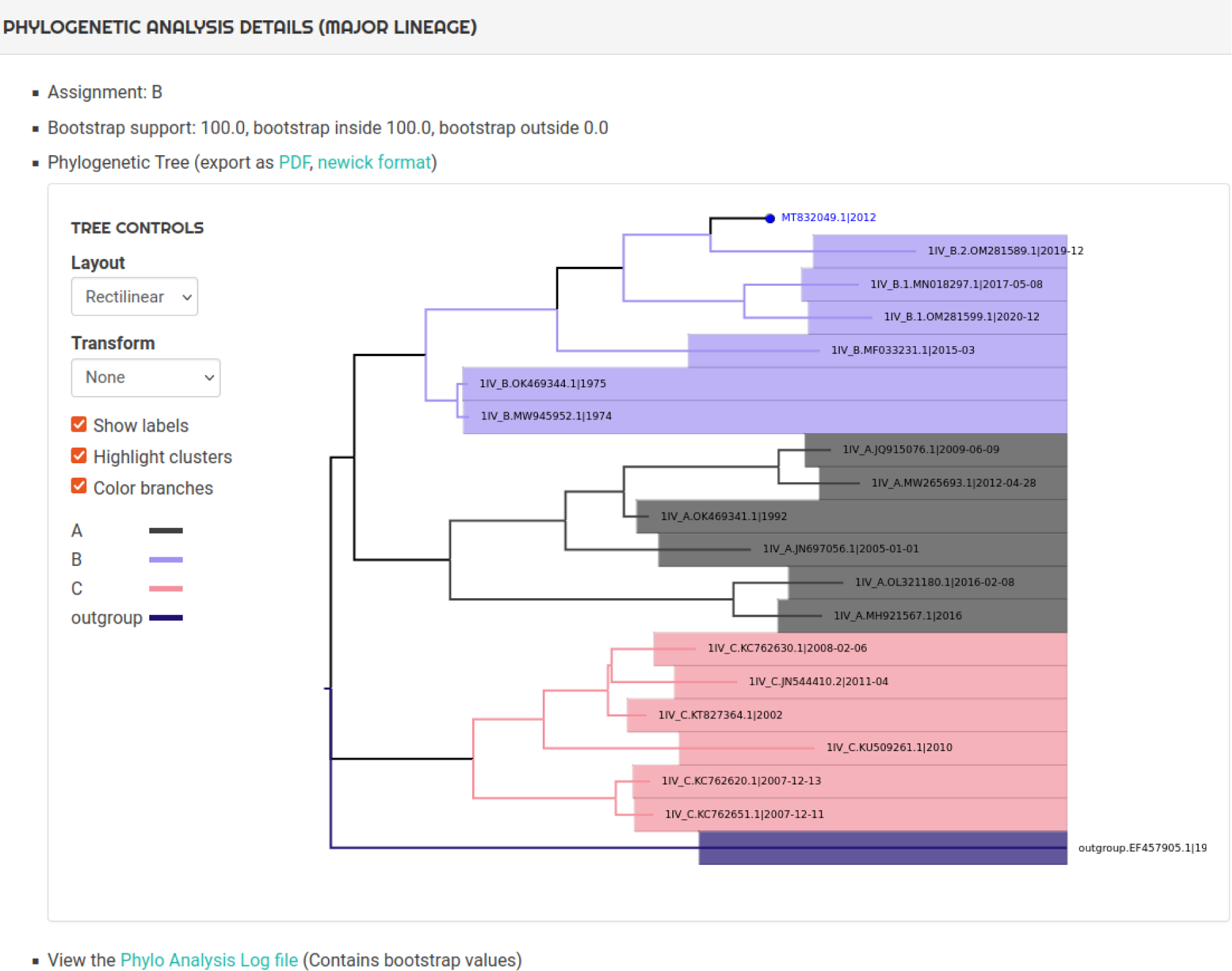
| Genome Detective dengue subtyping tool phylogenetic analysis details An example of the analysis details for a major lineage.

**Figure S11.**
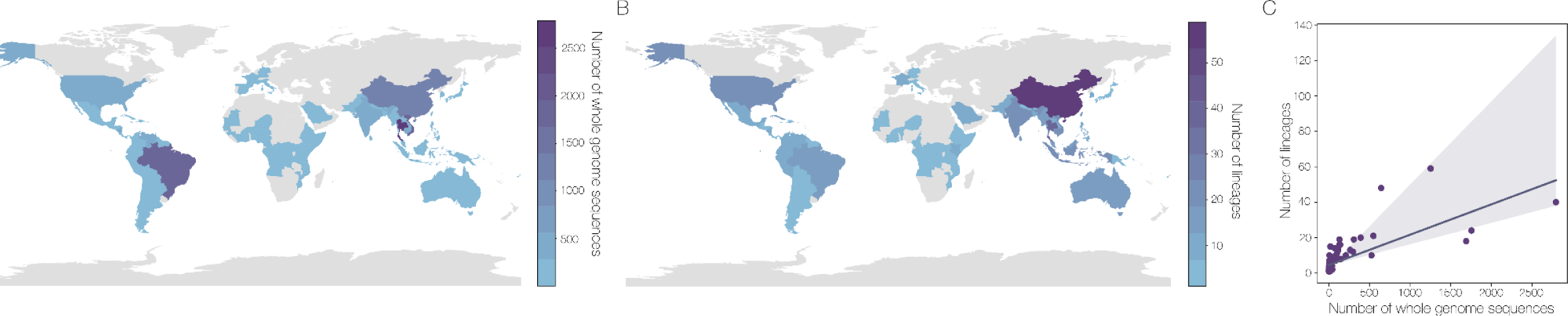
| Sampling distribution of DENV whole genome sequences. A) Sampling location of whole genome sequences by country B) Number of lineages sampled in each country C) Linear regression of the number of whole genome sequences against the number of lineages in each country (p<0.001).

