## Supplementary material for "A new lineage nomenclature to aid genomic surveillance of dengue virus": Table S4

We gratefully acknowledge the following Authors from the Originating laboratories responsible for obtaining the specimens, as well as the Submitting laboratories where the genome data were generated and shared via GISAID, on which this research is based.

All Submitters of data may be contacted directly via www.gisaid.org

Authors are sorted alphabetically.

| Accession ID | Originating Laboratory | Submitting Laboratory | Authors |
|---|---|---|---|
| EPI_ISL_10238963, EPI_ISL_10239118, EPI_ISL_10239375 | Centers for Disease Control and Prevention | Centers for Disease Control and Prevention | Santiago,G.A., Flores,B., Gonzalez-Morales,G., Charriez,K.N., Marcelo-Nique,A., Garcia-Mendoza,M.P. and Munoz-Jordan,J.L. |
| EPI_ISL_10331061, EPI_ISL_10331624, EPI_ISL_10331626, EPI_ISL_10331627, EPI_ISL_10331628, EPI_ISL_10331629, EPI_ISL_10331630, EPI_ISL_10333565, EPI_ISL_10333566, EPI_ISL_10334285, EPI_ISL_10334633, EPI_ISL_10334634, EPI_ISL_10334635, EPI_ISL_10334636 | | | |
| see above | Centers for Disease Control and Prevention | Centers for Disease Control and Prevention | Santiago,G.A., Gonzalez-Morales,G., Charriez,K., Flores,B., Stanek,D., Morrison,A. and Munoz-Jordan,J.L. |
| EPI_ISL_10656152, EPI_ISL_10656413 | French National Center of Arbovirus - Institut de recherche biomédicale des armées (IRBA) | French National Center of Arbovirus - Institut de recherche biomédicale des armées (IRBA) | Leparc-Goffart,I., Grard,G., Durand,G.A., Piorkowski,G., Noel,H. and de Lamballerie,X. |
| EPI_ISL_10707292 | Indian Institute of Technology (IIT) Roorkee | Indian Institute of Technology (IIT) Roorkee | Bhutkar,M.R. |
| EPI_ISL_10724043 | University of Hyderabad | University of Hyderabad | Maisnam,D. and Musturi,V. |
| EPI_ISL_10952927, EPI_ISL_10968441 | General Sir John Kotelawala Defence University, Faculty of Medicine | General Sir John Kotelawala Defence University, Faculty of Medicine | Abeygoonawardena,H., De Silva,A.D., Wijesinghe,N., Navaratne,V., Dilshani,P., Sundralingam,T. and Punchihewa,C. |
| EPI_ISL_10981739 | University of Hyderabad | University of Hyderabad | Maisnam,D. and Musturi,V. |
| EPI_ISL_11104976, EPI_ISL_11104977, EPI_ISL_11104978, EPI_ISL_11104979 | US Army Medical Directorate of the Armed Forces Research Institute of Medical Sciences (USAMD-AFRIMS) | US Army Medical Directorate of the Armed Forces Research Institute of Medical Sciences (USAMD-AFRIMS) | Jones,A.R., Rutvisuttinunt,W., Chinnawirotpisan,P., Manasatienkit,W., Ongajchaowlerd,P., Ellison,D.W., Kalayanarooj,S., Suntarattiwong,P., Fernandez,S. and Klungthong,C. |
| EPI_ISL_11177149 | Ahmadu Bello University | Ahmadu Bello University | Tizhe,D.T., Kwaga,J.K.P. and Kia,G.S.N. |
| EPI_ISL_11254706, EPI_ISL_11254719, EPI_ISL_11255073, EPI_ISL_11255150, EPI_ISL_11255151, EPI_ISL_11255347, EPI_ISL_11255348, EPI_ISL_11255349, EPI_ISL_11255350, EPI_ISL_11256147, EPI_ISL_11256148, EPI_ISL_11256156, EPI_ISL_11256326, EPI_ISL_11256327, EPI_ISL_11256478, EPI_ISL_11256479, EPI_ISL_11256480, EPI_ISL_11256481, EPI_ISL_11256482, EPI_ISL_11256483, EPI_ISL_11256484, EPI_ISL_11256485, EPI_ISL_11256486, EPI_ISL_11256643, EPI_ISL_11256644, EPI_ISL_11256645, EPI_ISL_11256698, EPI_ISL_11256709, EPI_ISL_11256742, EPI_ISL_11256763, EPI_ISL_11256765, EPI_ISL_11256766, EPI_ISL_11256767, EPI_ISL_11256773, EPI_ISL_11256791, EPI_ISL_11256795, EPI_ISL_11256805, EPI_ISL_11256812, EPI_ISL_11256835, EPI_ISL_11256839, EPI_ISL_11256842, EPI_ISL_11256854, EPI_ISL_11256858, EPI_ISL_11256867, EPI_ISL_11256882, EPI_ISL_11256896, EPI_ISL_11256902, EPI_ISL_11256909, EPI_ISL_11256915, EPI_ISL_11256919, EPI_ISL_11256922, EPI_ISL_11256931, EPI_ISL_11256932, EPI_ISL_11256933, EPI_ISL_11256938, EPI_ISL_11256947, EPI_ISL_11256957, EPI_ISL_11256972, EPI_ISL_11256988, EPI_ISL_11256991, EPI_ISL_11256995, EPI_ISL_11256996, EPI_ISL_11257096, EPI_ISL_11257105, EPI_ISL_11257111, EPI_ISL_11257112, EPI_ISL_11257114, EPI_ISL_11257118, EPI_ISL_11257123, EPI_ISL_11257133, EPI_ISL_11257156, EPI_ISL_11257174, EPI_ISL_11257177, EPI_ISL_11257183, EPI_ISL_11257185, EPI_ISL_11257186, EPI_ISL_11257196, EPI_ISL_11257200, EPI_ISL_11257207, EPI_ISL_11257208, EPI_ISL_11257210, EPI_ISL_11257218, EPI_ISL_11257244, EPI_ISL_11257249, EPI_ISL_11257254, EPI_ISL_11257308, EPI_ISL_11257328, EPI_ISL_11257338, EPI_ISL_11257347, EPI_ISL_11257352, EPI_ISL_11257354, EPI_ISL_11257358, EPI_ISL_11257366, EPI_ISL_11257377, EPI_ISL_11257398, EPI_ISL_11257425, EPI_ISL_11257488, EPI_ISL_11257497, EPI_ISL_11257503, EPI_ISL_11257518, EPI_ISL_11257519, EPI_ISL_11257520, EPI_ISL_11257544, EPI_ISL_11257546, EPI_ISL_11257551, EPI_ISL_11257571, EPI_ISL_11257587, EPI_ISL_11257653, EPI_ISL_11257659, EPI_ISL_11257679, EPI_ISL_11257680, EPI_ISL_11257682, EPI_ISL_11257708, EPI_ISL_11257709, EPI_ISL_11300437, EPI_ISL_11300438, EPI_ISL_11301540, EPI_ISL_11301541, EPI_ISL_11309849, EPI_ISL_11309850, EPI_ISL_11309945, EPI_ISL_11309946, EPI_ISL_11309974, EPI_ISL_11310228, EPI_ISL_11310229, EPI_ISL_11310230, EPI_ISL_11310557, EPI_ISL_11310558, EPI_ISL_11310559, EPI_ISL_11311037, EPI_ISL_11311353, EPI_ISL_11311450, EPI_ISL_11311451, EPI_ISL_11311452, EPI_ISL_11311921, EPI_ISL_11311922, EPI_ISL_11311923, EPI_ISL_11311924, EPI_ISL_11311925, EPI_ISL_11311926, EPI_ISL_11311927, EPI_ISL_11311928, EPI_ISL_11312524, EPI_ISL_11312525, EPI_ISL_11312526, EPI_ISL_11312527, EPI_ISL_11312528, EPI_ISL_11312529, EPI_ISL_11312530, EPI_ISL_11312623, EPI_ISL_11313367, EPI_ISL_11313368, EPI_ISL_11313369, EPI_ISL_11313370, EPI_ISL_11313371, EPI_ISL_11313372, EPI_ISL_11313373, EPI_ISL_11313374, EPI_ISL_11313375, EPI_ISL_11313376, EPI_ISL_11313377, EPI_ISL_11313378, EPI_ISL_11313379, EPI_ISL_11313675, EPI_ISL_11313676, EPI_ISL_11313677, EPI_ISL_11313678, EPI_ISL_11313679, EPI_ISL_11313680, EPI_ISL_11313681, EPI_ISL_11313682, EPI_ISL_11313683, EPI_ISL_11314027, EPI_ISL_11314028, EPI_ISL_11314029, EPI_ISL_11314030, EPI_ISL_11314031, EPI_ISL_11314364, EPI_ISL_11314365, EPI_ISL_11314366, EPI_ISL_11314367, EPI_ISL_11314368, EPI_ISL_11314369, EPI_ISL_11314370, EPI_ISL_11314371, EPI_ISL_11314372, EPI_ISL_11315090, EPI_ISL_11315091, EPI_ISL_11316855, EPI_ISL_11317555, EPI_ISL_11317556, EPI_ISL_11317557, EPI_ISL_11317558, EPI_ISL_11317559, EPI_ISL_11317560, EPI_ISL_11317561, EPI_ISL_11317956, EPI_ISL_11317957, EPI_ISL_11317958, EPI_ISL_11317959, EPI_ISL_11317960, EPI_ISL_11317961, EPI_ISL_11318993, EPI_ISL_11318994, EPI_ISL_11318995, EPI_ISL_11318996, EPI_ISL_11318997, EPI_ISL_11318998, EPI_ISL_11318999, EPI_ISL_11319000, EPI_ISL_11319001, EPI_ISL_11319002, EPI_ISL_11319324, EPI_ISL_11319325, EPI_ISL_11319326, EPI_ISL_11319327, EPI_ISL_11319328, EPI_ISL_11319329, EPI_ISL_11319330, EPI_ISL_11319331, EPI_ISL_11319332, EPI_ISL_11319333, EPI_ISL_11319334, EPI_ISL_11319335, EPI_ISL_11319336, EPI_ISL_11319337, EPI_ISL_11319338, EPI_ISL_11319339, EPI_ISL_11319340, EPI_ISL_11319341, EPI_ISL_11319342, EPI_ISL_11319343, EPI_ISL_11319344, EPI_ISL_11319345, EPI_ISL_11319346, EPI_ISL_11319347, EPI_ISL_11319348, EPI_ISL_11319349, EPI_ISL_11319350, EPI_ISL_11319351, EPI_ISL_11319352, EPI_ISL_11319353, EPI_ISL_11319354, EPI_ISL_11319355, EPI_ISL_11319522, EPI_ISL_11319523, EPI_ISL_11319524, EPI_ISL_11319768, EPI_ISL_11319769, EPI_ISL_11319770, EPI_ISL_11319771, EPI_ISL_11319772, EPI_ISL_11319773, EPI_ISL_11319774, EPI_ISL_11319775, EPI_ISL_11319776, EPI_ISL_11319777, EPI_ISL_11319778, EPI_ISL_11319779, EPI_ISL_11319780, EPI_ISL_11319781, EPI_ISL_11319782, EPI_ISL_11319783, EPI_ISL_11319784, EPI_ISL_11319785, EPI_ISL_11319786, EPI_ISL_11319787, EPI_ISL_11319788, EPI_ISL_11319789, EPI_ISL_11320119, EPI_ISL_11320120, EPI_ISL_11320121, EPI_ISL_11320122, EPI_ISL_11320123, EPI_ISL_11320124, EPI_ISL_11320257, EPI_ISL_11320258, EPI_ISL_11320259, EPI_ISL_11320260, EPI_ISL_11320261, EPI_ISL_11320262, EPI_ISL_11320263, EPI_ISL_11320264, EPI_ISL_11320265, EPI_ISL_11320266, EPI_ISL_11320267, EPI_ISL_11320268, EPI_ISL_11320269, EPI_ISL_11320270, EPI_ISL_11320271, EPI_ISL_11320272, EPI_ISL_11320273, EPI_ISL_11320274, EPI_ISL_11320275, EPI_ISL_11320276, EPI_ISL_11320277, EPI_ISL_11320278, EPI_ISL_11320279, EPI_ISL_11320280, EPI_ISL_11320281, EPI_ISL_11320282, EPI_ISL_11320283, EPI_ISL_11320284, EPI_ISL_11320285, EPI_ISL_11320286, EPI_ISL_11320287, EPI_ISL_11320288, EPI_ISL_11320289, EPI_ISL_11320290, EPI_ISL_11320291, EPI_ISL_11320292, EPI_ISL_11320293, EPI_ISL_11320294, EPI_ISL_11320295, EPI_ISL_11320296, EPI_ISL_11320297 | | | |
| see above | US Army Medical Directorate of the Armed Forces Research Institute of Medical Sciences (USAMD-AFRIMS) | US Army Medical Directorate of the Armed Forces Research Institute of Medical Sciences (USAMD-AFRIMS) | Klungthong,C., Phonpakobsin,T., Thaisomboonsuk,B., Chinnawirotpisan,P., Manasatienkit,W., Ong-ajchaowlerd,P., Rodpradit,P., Nisalak,A., Kalayanarooj,S., Punpanich Vandepitt,W., Macareo,L.R., Gibbons,R.V., Jarman,R.G. and Fernandez,S. |
| EPI_ISL_11330490, EPI_ISL_11330583, EPI_ISL_11330672 | Shree Guru Gobind Singh Tricentenary University (SGT) | Shree Guru Gobind Singh Tricentenary University (SGT) | Sharma,M. |
| EPI_ISL_11536036, EPI_ISL_11536037, EPI_ISL_11536765, EPI_ISL_11537627, EPI_ISL_11537628, EPI_ISL_11537629, EPI_ISL_11537630, EPI_ISL_11537631, EPI_ISL_11537632, EPI_ISL_11537633, EPI_ISL_11537634, EPI_ISL_11537635, EPI_ISL_11538419, EPI_ISL_11538420 | | | |
| see above | US Naval Medical Research Unit 6 | US Naval Medical Research Unit 6 | Espejo,V.I., Troncos,G.M., Cruz,C.D., Olivos,J.M., Ramos,R.E., Chuquirachi,F.J., Ocana,V.R., Holguin,C.E., Cedano,L.E., Ampuero,J.S., Tinoco,Y.O. and Graf,P.C. |
| EPI_ISL_11538421 | US Naval Medical Research Unit 6 | US Naval Medical Research Unit 6 | Espejo,V.I., Troncos,G.M., Cruz,C.D., Olivos,J.M., Ramos,R.E., Chuquirachi,F.J., Ocana,V.R., Holguin,C.E., Ampuero,J.S., Tinoco,Y.O. and Graf,P.C. |
| EPI_ISL_11538422, EPI_ISL_11538423, EPI_ISL_11538424 | US Naval Medical Research Unit 6 | US Naval Medical Research Unit 6 | Espejo,V.I., Troncos,G.M., Cruz,C.D., Olivos,J.M., Ramos,R.E., Chuquirachi,F.J., Ocana,V.R., Holguin,C.E., Cedano,L.E., Ampuero,J.S., Tinoco,Y.O. and Graf,P.C. |
| EPI_ISL_11538425 | US Naval Medical Research Unit 6 | US Naval Medical Research Unit 6 | Espejo,V.I., Troncos,G.M., Cruz,C.D., Olivos,J.M., Ramos,R.E., Chuquirachi,F.J., Ocana,V.R., Holguin,C.E., Ampuero,J.S., Tinoco,Y.O. and Graf,P.C. |
| EPI_ISL_11539117, EPI_ISL_11539118, EPI_ISL_11539119, EPI_ISL_11539120, EPI_ISL_11539121, EPI_ISL_11539122 | US Naval Medical Research Unit 6 | US Naval Medical Research Unit 6 | Espejo,V.I., Troncos,G.M., Cruz,C.D., Olivos,J.M., Ramos,R.E., Chuquirachi,F.J., Ocana,V.R., Holguin,C.E., Cedano,L.E., Ampuero,J.S., Tinoco,Y.O. and Graf,P.C. |
| EPI_ISL_11539123, EPI_ISL_11539124, EPI_ISL_11539125, EPI_ISL_11539126, EPI_ISL_11539127, EPI_ISL_11540732 | US Naval Medical Research Unit 6 | US Naval Medical Research Unit 6 | Espejo,V.I., Troncos,G.M., Cruz,C.D., Olivos,J.M., Ramos,R.E., Chuquirachi,F.J., Ocana,V.R., Holguin,C.E., Ampuero,J.S., Tinoco,Y.O. and Graf,P.C. |
| EPI_ISL_11542198 | Bangladesh Council of Scientific and Industrial Research (BCSIR) | Bangladesh Council of Scientific and Industrial Research (BCSIR) | Sarkar,M.M., Rabbi,M.F., Akter,S., Islam,M.S., Banu,T.A., Jahan,I., Miah,M.I., Goswami,B., Habib,M.A., Akhteruzzaman,S., Shaikh,M.A., Rubaiya,A., Ilias,M. and Khan,M.S. |
| EPI_ISL_11542199, EPI_ISL_11542200, EPI_ISL_11542201, EPI_ISL_11542202, EPI_ISL_11542203, EPI_ISL_11542204, EPI_ISL_11542205, EPI_ISL_11542206, EPI_ISL_11542207, EPI_ISL_11542208, EPI_ISL_11542209, EPI_ISL_11542210, EPI_ISL_11542211 | | | |
| see above | Bangladesh Council of Scientific and Industrial Research (BCSIR) | Bangladesh Council of Scientific and Industrial Research (BCSIR) | Sarkar,M.M., Islam,M.S., Miah,M.I., Akter,S., Banu,T.A., Goswami,B., Jahan,I., Habib,M.A., Shaikh,M.A., Rubaiya,A., Ilias,M. and Khan,M.S. |
| EPI_ISL_11579708, EPI_ISL_11579724, EPI_ISL_11579725, EPI_ISL_11579726, EPI_ISL_11579754, EPI_ISL_11579755, EPI_ISL_11579756, EPI_ISL_11579757, EPI_ISL_11579758, EPI_ISL_11579759, EPI_ISL_11579760, EPI_ISL_11579761, EPI_ISL_11579762, EPI_ISL_11579763, EPI_ISL_11579764, EPI_ISL_11579765, EPI_ISL_11579766, EPI_ISL_11579767, EPI_ISL_11579768, EPI_ISL_11579769, EPI_ISL_11579770, EPI_ISL_11579771 | | | |
| see above | Faculdade de Medicina de Sao Jose do Rio Preto, Parasitarias e Dermatologicas | Faculdade de Medicina de Sao Jose do Rio Preto, Parasitarias e Dermatologicas | Marques,B.C., Candido,D.S., Estofolete,C.F., Sacchetto,L., Banho,C.A., Dutra,K.R., Salles,F.C., Dourado,F.S., Jesus,J.G., Sabino,E.C., Faria,N. and Nogueira,M.L. |
| EPI_ISL_11579772, EPI_ISL_11579773, EPI_ISL_11579774, EPI_ISL_11579775, EPI_ISL_11579776, EPI_ISL_11579777, EPI_ISL_11579778, EPI_ISL_11579779, EPI_ISL_11579780, EPI_ISL_11579781, EPI_ISL_11579782, EPI_ISL_11580637 | | | |
| see above | Bangladesh Council of Scientific and Industrial Research (BCSIR) | Bangladesh Council of Scientific and Industrial Research (BCSIR) | Sarkar,M.M., Islam,M.S., Miah,M.I., Akter,S., Banu,T.A., Goswami,B., Jahan,I., Habib,M.A., Shaikh,M.A., Rubaiya,A., Ilias,M. and Khan,M.S. |
| EPI_ISL_11629403 | Bangladesh Council of Scientific and Industrial Research (BCSIR) | Bangladesh Council of Scientific and Industrial Research (BCSIR) | Sarkar,M.M., Rabbi,M.F., Akter,S., Islam,M.S., Banu,T.A., Jahan,I., Miah,M.I., Goswami,B., Habib,M.A., Akhteruzzaman,S., Shaikh,M.A., Rubaiya,A., Ilias,M. and Khan,M.S. |
| EPI_ISL_11631874, EPI_ISL_11631875, EPI_ISL_11631876, EPI_ISL_11631877, EPI_ISL_11631878, EPI_ISL_11631879, EPI_ISL_11631880, EPI_ISL_11631881, EPI_ISL_11631882 | Bangladesh Council of Scientific and Industrial Research (BCSIR) | Bangladesh Council of Scientific and Industrial Research (BCSIR) | Sarkar,M.M., Islam,M.S., Miah,M.I., Akter,S., Banu,T.A., Goswami,B., Jahan,I., Habib,M.A., Shaikh,M.A., Rubaiya,A., Ilias,M. and Khan,M.S. |
| EPI_ISL_11971623 | Institut de recherche biomédicale des armées (IRBA) | Institut de recherche biomédicale des armées (IRBA) | Durand,G.A., Grard,G., Leparc-Goffart,I. and Piorkowski,G. |
| EPI_ISL_12001738, EPI_ISL_12001740, EPI_ISL_12001744, EPI_ISL_12001835, EPI_ISL_12001836, EPI_ISL_12001837, EPI_ISL_12001838, EPI_ISL_12001917, EPI_ISL_12002622 | Institut Pasteur de Dakar | Institut Pasteur de Dakar | Dieng,I., Diallo,A., Ndiaye,M., Mahmadi,M., Diagne,M.M., Sanke,S., Ndione,M.H.D., Gaye,A., Sagne,S.N., Heraud,J.M., Sall,A.A., Fall,G., Loucoubar,C., Faye,O. and Faye,O. |
| EPI_ISL_12235795, EPI_ISL_12235796 | Nagasaki University | Nagasaki University | Ushijima,Y., Abe,H., Mbadinga,M.J.V.M., Nguema-Ondo,G., Bikangui,R., Agnandji,S.T., Lell,B. and Yasuda,J. |
| EPI_ISL_12387067 | DDBJ | DDBJ | Arunraj,D. |
| EPI_ISL_12391669 | Bernhard Nocht Institute for Tropical Medicine | Bernhard Nocht Institute for Tropical Medicine | Tchibozo,C., Hounknownanrin,G., Bialonski,A., Schmidt-Chanasit,J., Joest,H. and Yadouleton,A. |
| EPI_ISL_12665303, EPI_ISL_12665304, EPI_ISL_12665305 | US Naval Medical Research Unit 6 | US Naval Medical Research Unit 6 | Espejo,V.I., Troncos,G.M., Cruz,C.D., Gomez-Camargo,D., Cano-Perez,E., Ampuero,J.S., Tinoco,Y.O., Silva,M.E. and Graf,P.C. |
| EPI_ISL_12869518, EPI_ISL_12869609, EPI_ISL_12869610, EPI_ISL_12869611, EPI_ISL_12869612, EPI_ISL_12869613, EPI_ISL_12869614, EPI_ISL_12869615, EPI_ISL_12869696, EPI_ISL_12869697, EPI_ISL_12869698, EPI_ISL_12869699, EPI_ISL_12869700, EPI_ISL_12869701, EPI_ISL_12869702, EPI_ISL_12869703, EPI_ISL_12869704, EPI_ISL_12869705, EPI_ISL_12869706, EPI_ISL_12869707, EPI_ISL_12869708, EPI_ISL_12869772, EPI_ISL_12869773, EPI_ISL_12869774, EPI_ISL_12869775, EPI_ISL_12869776, EPI_ISL_12869777 | | | |
| see above | Eijkman Institute for Molecular Biology | Eijkman Institute for Molecular Biology | Masyeni,S., Hayati,R.F., Santoso,M.S. and Sasmono,R.T. |
| EPI_ISL_12996324 | National Institute of Cholera and Enteric Diseases | National Institute of Cholera and Enteric Diseases | Verma,P. and Sadhukhan,P.C. |
| EPI_ISL_13021966, EPI_ISL_13022216, EPI_ISL_13022217, EPI_ISL_13022218, EPI_ISL_13022219, EPI_ISL_13022220, EPI_ISL_13022221, EPI_ISL_13022222 | UMR PIMIT The Mixed Research Unit (UMR), Infectious Processes in Tropical Island Environments (PIMIT) | UMR PIMIT The Mixed Research Unit (UMR), Infectious Processes in Tropical Island Environments (PIMIT) | Hafsia,S., Barbar,T., Wilkinson,D.A., Atyame,C., Biscornet,L., Bibi,J., Louange,M., Gedeon,J., De Santis,O., Flahault,A., Cabie,A., Bertolotti,A. and Mavingui,P. |
| EPI_ISL_13101925 | General Sir John Kotelawala Defence University, Faculty of Medicine | General Sir John Kotelawala Defence University, Faculty of Medicine | Abeygoonawardena,H., De Silva,A.D., Wijesinghe,N., Navaratne,V., Dilshani,P., Sundralingam,T. and Punchihewa,C. |

| | | | |
|---|---|---|---|
| EPI_ISL_13154917 | US Naval Medical Research Unit 6 | US Naval Medical Research Unit 6 | Cruz,C.D., Alba,M., Mores,C., Torre,A.J., Pollet,S. and Leguia,M. |
| EPI_ISL_13167664 | Pondicherry Institute of Medical Sciences (PIMS) | Pondicherry Institute of Medical Sciences (PIMS) | Bhat,S., Nair,S., Natarajan,V. and Pandian,B. |
| EPI_ISL_13168052, EPI_ISL_13168053, EPI_ISL_13168765, EPI_ISL_13168766, EPI_ISL_13168767, EPI_ISL_13168768, EPI_ISL_13168769, EPI_ISL_13168770, EPI_ISL_13168899, EPI_ISL_13168992, EPI_ISL_13169617, EPI_ISL_13169687, EPI_ISL_13169688, EPI_ISL_13169689, EPI_ISL_13169690, EPI_ISL_13169691, EPI_ISL_13170382, EPI_ISL_13170383, EPI_ISL_13172332, EPI_ISL_13172333, EPI_ISL_13172334, EPI_ISL_13172335, EPI_ISL_13172336, EPI_ISL_13172337, EPI_ISL_13172338 | | | |
| see above | King George's Medical University | King George's Medical University | Prakash,O., Khan,D.N., Verma,A.K., Ramakrishna,V. and Jain,A. |
| EPI_ISL_13198227, EPI_ISL_13198228, EPI_ISL_13198229, EPI_ISL_13198230, EPI_ISL_13198384, EPI_ISL_13198492, EPI_ISL_13199356, EPI_ISL_13199357, EPI_ISL_13199358, EPI_ISL_13199390, EPI_ISL_13199579, EPI_ISL_13199681 | | | |
| see above | Pondicherry Institute of Medical Sciences (PIMS) | Pondicherry Institute of Medical Sciences (PIMS) | Bhat,S., Nair,S., Natarajan,V. and Pandian,B. |
| EPI_ISL_13321094, EPI_ISL_13321190, EPI_ISL_13321316, EPI_ISL_13321317, EPI_ISL_13321666, EPI_ISL_13321667, EPI_ISL_13321668, EPI_ISL_13321669, EPI_ISL_13321670, EPI_ISL_13321671, EPI_ISL_13321672, EPI_ISL_13321673, EPI_ISL_13321674, EPI_ISL_13321675, EPI_ISL_13321676, EPI_ISL_13321677, EPI_ISL_13321960, EPI_ISL_13321961, EPI_ISL_13321962, EPI_ISL_13321963, EPI_ISL_13321964, EPI_ISL_13321965, EPI_ISL_13321966, EPI_ISL_13321967, EPI_ISL_13321968, EPI_ISL_13321969, EPI_ISL_13321970, EPI_ISL_13321971, EPI_ISL_13321972, EPI_ISL_13321973, EPI_ISL_13321974, EPI_ISL_13321975, EPI_ISL_13321976, EPI_ISL_13337360, EPI_ISL_13337805, EPI_ISL_13337815, EPI_ISL_13337910, EPI_ISL_13340696, EPI_ISL_13340697, EPI_ISL_13340698, EPI_ISL_13340699, EPI_ISL_13340700, EPI_ISL_13340701, EPI_ISL_13340702, EPI_ISL_13340703, EPI_ISL_13340704 | | | |
| see above | Chulalongkorn University, Faculty of Medicine | Chulalongkorn University, Faculty of Medicine | Khongwichit,S., Chuchaona,W., Vongpunsawad,S. and Poovorawan,Y. |
| EPI_ISL_13465463, EPI_ISL_13469902, EPI_ISL_13496557, EPI_ISL_13496558, EPI_ISL_13512178, EPI_ISL_13512179 | Guangzhou Center for Disease Control and Prevention | Guangzhou Center for Disease Control and Prevention | Su,W., Cao,Y. and Lu,W. |
| EPI_ISL_13512580 | Guangzhou Center for Disease Control and Prevention | Guangzhou Center for Disease Control and Prevention | Su,W., Jiang,L. and Lu,W. |
| EPI_ISL_13531025, EPI_ISL_13531026, EPI_ISL_13531027, EPI_ISL_13531028, EPI_ISL_13531029, EPI_ISL_13531030, EPI_ISL_13531031, EPI_ISL_13531032, EPI_ISL_13531033, EPI_ISL_13531034, EPI_ISL_13531035, EPI_ISL_13531036, EPI_ISL_13531037, EPI_ISL_13531038, EPI_ISL_13531039, EPI_ISL_13531040, EPI_ISL_13531041, EPI_ISL_13531042, EPI_ISL_13531046, EPI_ISL_13531088, EPI_ISL_13531089 | | | |
| see above | Guangzhou Center for Disease Control and Prevention | Guangzhou Center for Disease Control and Prevention | Su,W., Cao,Y. and Lu,W. |
| EPI_ISL_13564328, EPI_ISL_13564401 | Guangzhou Center for Disease Control and Prevention | Guangzhou Center for Disease Control and Prevention | Su,W., Jiang,L. and Lu,W. |
| EPI_ISL_13564402 | Guangzhou Center for Disease Control and Prevention | Guangzhou Center for Disease Control and Prevention | Su,W., Cao,Y. and Lu,W. |
| EPI_ISL_13564403, EPI_ISL_13564404 | Guangzhou Center for Disease Control and Prevention | Guangzhou Center for Disease Control and Prevention | Su,W., Jiang,L. and Lu,W. |
| EPI_ISL_13564405 | Guangzhou Center for Disease Control and Prevention | Guangzhou Center for Disease Control and Prevention | Su,W., Cao,Y. and Lu,W. |
| EPI_ISL_13564406, EPI_ISL_13564407, EPI_ISL_13564408, EPI_ISL_13564409, EPI_ISL_13564410 | Guangzhou Center for Disease Control and Prevention | Guangzhou Center for Disease Control and Prevention | Su,W., Jiang,L. and Lu,W. |
| EPI_ISL_13564411 | Guangzhou Center for Disease Control and Prevention | Guangzhou Center for Disease Control and Prevention | Su,W., Cao,Y. and Lu,W. |
| EPI_ISL_13564412, EPI_ISL_13564413, EPI_ISL_13564414 | Guangzhou Center for Disease Control and Prevention | Guangzhou Center for Disease Control and Prevention | Su,W., Jiang,L. and Lu,W. |
| EPI_ISL_13564415, EPI_ISL_13564416 | Guangzhou Center for Disease Control and Prevention | Guangzhou Center for Disease Control and Prevention | Su,W., Cao,Y. and Lu,W. |
| EPI_ISL_13564417 | Guangzhou Center for Disease Control and Prevention | Guangzhou Center for Disease Control and Prevention | Su,W., Jiang,L. and Lu,W. |
| EPI_ISL_13564418 | Guangzhou Center for Disease Control and Prevention | Guangzhou Center for Disease Control and Prevention | Su,W., Jiang,L. and Cao,Y. |
| EPI_ISL_13564419, EPI_ISL_13564420, EPI_ISL_13564421, EPI_ISL_13564422 | Guangzhou Center for Disease Control and Prevention | Guangzhou Center for Disease Control and Prevention | Su,W., Jiang,L. and Lu,W. |
| EPI_ISL_13564423 | Guangzhou Center for Disease Control and Prevention | Guangzhou Center for Disease Control and Prevention | Su,W., Cao,Y. and Lu,W. |
| EPI_ISL_13564424 | Guangzhou Center for Disease Control and Prevention | Guangzhou Center for Disease Control and Prevention | Su,W., Jiang,L. and Lu,W. |
| EPI_ISL_13564625 | Guangzhou Center for Disease Control and Prevention | Guangzhou Center for Disease Control and Prevention | Su,W., Jiang,L. and Cao,Y. |
| EPI_ISL_13564626, EPI_ISL_13564627, EPI_ISL_13564628, EPI_ISL_13564629, EPI_ISL_13564630 | Guangzhou Center for Disease Control and Prevention | Guangzhou Center for Disease Control and Prevention | Su,W., Jiang,L. and Lu,W. |
| EPI_ISL_13564631 | Guangzhou Center for Disease Control and Prevention | Guangzhou Center for Disease Control and Prevention | Su,W., Cao,Y. and Lu,W. |
| EPI_ISL_13564632 | Guangzhou Center for Disease Control and Prevention | Guangzhou Center for Disease Control and Prevention | Su,W., Jiang,L. and Lu,W. |
| EPI_ISL_13564633 | Haizhu District Center for Disease Control and Prevention | Guangdong Center for Disease Control and Prevention | Wu,D., Zhang,H., Zhou,H. and Li,H. |
| EPI_ISL_13564913 | Guangzhou Center for Disease Control and Prevention | Guangzhou Center for Disease Control and Prevention | Su,W., Jiang,L. and Lu,W. |
| EPI_ISL_13564914 | Guangzhou Center for Disease Control and Prevention | Guangzhou Center for Disease Control and Prevention | Su,W., Cao,Y. and Lu,W. |
| EPI_ISL_13565025, EPI_ISL_13566018, EPI_ISL_13566020, EPI_ISL_13570033, EPI_ISL_13570625, EPI_ISL_13573945 | Guangzhou Center for Disease Control and Prevention | Guangzhou Center for Disease Control and Prevention | Su,W., Jiang,L. and Lu,W. |
| EPI_ISL_13573946 | Guangzhou Center for Disease Control and Prevention | Guangzhou Center for Disease Control and Prevention | Su,W., Cao,Y. and Lu,W. |
| EPI_ISL_13573947, EPI_ISL_13612002 | Guangzhou Center for Disease Control and Prevention | Guangzhou Center for Disease Control and Prevention | Su,W., Jiang,L. and Lu,W. |
| EPI_ISL_13631826, EPI_ISL_13631827, EPI_ISL_13632153, EPI_ISL_13632154, EPI_ISL_13632155, EPI_ISL_13632157, EPI_ISL_13632158, EPI_ISL_13632159, EPI_ISL_13632160, EPI_ISL_13632161, EPI_ISL_13632162, EPI_ISL_13632163, EPI_ISL_13632164, EPI_ISL_13632165 | | | |
| see above | National Institute of Health | National Institute of Health | Umair,M., Hakim,R., Bibi,S. and Rehman,Z. |
| EPI_ISL_13632166, EPI_ISL_13632167, EPI_ISL_13632168, EPI_ISL_13632169, EPI_ISL_13632170, EPI_ISL_13632171, EPI_ISL_13632172, EPI_ISL_13632173, EPI_ISL_13632174, EPI_ISL_13632175, EPI_ISL_13632176, EPI_ISL_13632177, EPI_ISL_13632178, EPI_ISL_13632179, EPI_ISL_13632180, EPI_ISL_13632181, EPI_ISL_13632182, EPI_ISL_13632183, EPI_ISL_13632184, EPI_ISL_13632185, EPI_ISL_13632186, EPI_ISL_13632187, EPI_ISL_13632188, EPI_ISL_13632189, EPI_ISL_13632190, EPI_ISL_13632191, EPI_ISL_13632192, EPI_ISL_13632193 | | | |
| see above | Sichuan Center of Disease Control and Prevention | Sichuan Center of Disease Control and Prevention | Li,W., Jiang,M.F., Zhong,H.R., Cao,Y.O. and Feng,Y.L. |
| EPI_ISL_13738963, EPI_ISL_13738964, EPI_ISL_13765791, EPI_ISL_13765792, EPI_ISL_13765793, EPI_ISL_13765794, EPI_ISL_13766664, EPI_ISL_13766665, EPI_ISL_13767732, EPI_ISL_13767733, EPI_ISL_13767734, EPI_ISL_13767735, EPI_ISL_13767736, EPI_ISL_13767737, EPI_ISL_13767738, EPI_ISL_13770576, EPI_ISL_13770577, EPI_ISL_13770578, EPI_ISL_13770579, EPI_ISL_13770580, EPI_ISL_13770581, EPI_ISL_13770582, EPI_ISL_13770583, EPI_ISL_13770584, EPI_ISL_13770585, EPI_ISL_13770586, EPI_ISL_13770587, EPI_ISL_13770588, EPI_ISL_13770589, EPI_ISL_13770590, EPI_ISL_13770591 | | | |
| see above | King George's Medical University | King George's Medical University | Prakash,O., Khan,D.N., Verma,A.K., Ramakrishna,V. and Jain,A. |
| EPI_ISL_13859884 | Vector Control Research Centre | Vector Control Research Centre | Sankari,T., Jayasree,A., Rameela Sanya,K., Varghese,B., Kumar,M., Kumar,A. and Niranjan,R. |
| EPI_ISL_14081633, EPI_ISL_14081634, EPI_ISL_14081635, EPI_ISL_14081636, EPI_ISL_14081637, EPI_ISL_14081698, EPI_ISL_14081699, EPI_ISL_14081700, EPI_ISL_14081701, EPI_ISL_14081702, EPI_ISL_14083175, EPI_ISL_14083176, EPI_ISL_14083177, EPI_ISL_14083178, EPI_ISL_14083179, EPI_ISL_14083180, EPI_ISL_14083181, EPI_ISL_14083182, EPI_ISL_14083183, EPI_ISL_14083184 | | | |
| see above | Oxford University Clinical Research Unit | Oxford University Clinical Research Unit | Cao,T.T., Dong,T.H., Nguyen,N.M., Huynh,D.T., Ho,C.Q., Nguyen,P.T., Nguyen,C.V., Phan,Q.T., Huynh,T.T., Luong,T.T., Nguyen,H.V., Duong,T.B., Duong,K.T., Tran,V.T. and Yacoub,S. |
| EPI_ISL_14253126, EPI_ISL_14253359, EPI_ISL_14253360, EPI_ISL_14253571 | ICMR-National Institute of Cholera and Enteric Diseases | ICMR-National Institute of Cholera and Enteric Diseases | Baskey,U., Verma,P. and Sadhukhan,P.C. |
| EPI_ISL_14287342, EPI_ISL_14287343, EPI_ISL_14287344, EPI_ISL_14287345, EPI_ISL_14287346, EPI_ISL_14287620, EPI_ISL_14288925, EPI_ISL_14288926, EPI_ISL_14288927, EPI_ISL_14289595, EPI_ISL_14289596, EPI_ISL_14289597, EPI_ISL_14289598, EPI_ISL_14289599, EPI_ISL_14289600, EPI_ISL_14289601, EPI_ISL_14289602, EPI_ISL_14289603, EPI_ISL_14289604, EPI_ISL_14289605, EPI_ISL_14289606 | | | |
| see above | ICMR-National Institute of Cholera and Enteric Diseases | ICMR-National Institute of Cholera and Enteric Diseases | Baskey,U., Verma,P., Ghosh,A. and Sadhukhan,P.C. |
| EPI_ISL_14675766, EPI_ISL_14675767 | Laboratory of Tropical Disease Control of Ministry of Education, Sun Yat-sen University, | Laboratory of Tropical Disease Control of Ministry of Education, Sun Yat-sen University, | Hu,M., Wu,T., Yang,Y., Chen,T., Hao,J., Wei,Y., Luo,T., Wu and Li,Y.P. |
| EPI_ISL_14810174, EPI_ISL_14810175, EPI_ISL_14810176, EPI_ISL_14810177, EPI_ISL_14810178, EPI_ISL_14810179, EPI_ISL_14810180, EPI_ISL_14810181, EPI_ISL_14810182, EPI_ISL_14810273, EPI_ISL_14810368 | | | |
| see above | National Institute of Cholera and Enteric | National Institute of Cholera and Enteric Diseases | Sadhukhan,P.C. and Verma,P. |

| | | | |
|---|---|---|---|
| | Diseases | | |
| EPI_ISL_14818767 | Bernhard Nocht Institute for Tropical Medicine (BNITM) | Bernhard Nocht Institute for Tropical Medicine (BNITM) | Tchibozo,C., Hounknownanrin,G., Yadouleton,A., Bialonski,A., Agboli,E., Luhken,R., Schmidt-Chanasit,J. and Jost,H. |
| EPI_ISL_14853337 | Guangzhou Center for Disease Control and Prevention | Guangzhou Center for Disease Control and Prevention | Su,W., Cao,Y. and Lu,W. |
| EPI_ISL_14907012, EPI_ISL_14907478, EPI_ISL_14907479, EPI_ISL_14907480, EPI_ISL_14907481, EPI_ISL_14907482, EPI_ISL_14907483, EPI_ISL_14907615, EPI_ISL_14907616, EPI_ISL_14907617, EPI_ISL_14907618, EPI_ISL_14907619, EPI_ISL_14907620, EPI_ISL_14907621, EPI_ISL_14908177, EPI_ISL_14908230, EPI_ISL_14908231, EPI_ISL_14908232 | | | |
| see above | The National Environment Agency | The National Environment Agency | Hapuarachchi,H.C., Setoh,Y.X., Wong,J.C.C. and Ng,L.C. |
| EPI_ISL_14908638, EPI_ISL_14908639, EPI_ISL_14908640, EPI_ISL_14908641, EPI_ISL_14908642, EPI_ISL_14908643, EPI_ISL_14908644, EPI_ISL_14908645, EPI_ISL_14908646, EPI_ISL_14908647, EPI_ISL_14908648, EPI_ISL_14908649, EPI_ISL_14908650, EPI_ISL_14908651, EPI_ISL_14908652, EPI_ISL_14908653, EPI_ISL_14908654, EPI_ISL_14908655, EPI_ISL_14908656, EPI_ISL_14908657, EPI_ISL_14908658, EPI_ISL_14908659, EPI_ISL_14908660, EPI_ISL_14908661, EPI_ISL_14908662, EPI_ISL_14908663, EPI_ISL_14908664, EPI_ISL_14908665, EPI_ISL_14908666, EPI_ISL_14908667, EPI_ISL_14908668, EPI_ISL_14908669, EPI_ISL_14908670, EPI_ISL_14908671, EPI_ISL_14908672, EPI_ISL_14908673, EPI_ISL_14908674, EPI_ISL_14908675, EPI_ISL_14908676, EPI_ISL_14908677, EPI_ISL_14908678, EPI_ISL_14908679, EPI_ISL_14908680, EPI_ISL_14908681, EPI_ISL_14908682, EPI_ISL_14908683, EPI_ISL_14908684, EPI_ISL_14908685, EPI_ISL_14908686, EPI_ISL_14908687, EPI_ISL_14908688, EPI_ISL_14908689, EPI_ISL_14908690, EPI_ISL_14908691, EPI_ISL_14908692, EPI_ISL_14908693, EPI_ISL_14908694, EPI_ISL_14908695, EPI_ISL_14908696, EPI_ISL_14908697, EPI_ISL_14908698, EPI_ISL_14908699, EPI_ISL_14908700, EPI_ISL_14908701, EPI_ISL_14908702, EPI_ISL_14908703, EPI_ISL_14908704, EPI_ISL_14908705, EPI_ISL_14908706, EPI_ISL_14908707, EPI_ISL_14908708, EPI_ISL_14908709, EPI_ISL_14908710, EPI_ISL_14908711, EPI_ISL_14908712, EPI_ISL_14908713, EPI_ISL_14908714, EPI_ISL_14908715, EPI_ISL_14908716, EPI_ISL_14908717, EPI_ISL_14908718, EPI_ISL_14908719, EPI_ISL_14908720, EPI_ISL_14908721, EPI_ISL_14908722, EPI_ISL_14908723, EPI_ISL_14908724, EPI_ISL_14908725, EPI_ISL_14908726, EPI_ISL_14908727, EPI_ISL_14908728, EPI_ISL_14908729, EPI_ISL_14908730, EPI_ISL_14908731, EPI_ISL_14908732, EPI_ISL_14908733, EPI_ISL_14908734, EPI_ISL_14908735, EPI_ISL_14908736, EPI_ISL_14908737, EPI_ISL_14908738, EPI_ISL_14908739, EPI_ISL_14908740, EPI_ISL_14908741, EPI_ISL_14908742, EPI_ISL_14908743, EPI_ISL_14908744, EPI_ISL_14908745, EPI_ISL_14908746, EPI_ISL_14908747, EPI_ISL_14908748, EPI_ISL_14908749, EPI_ISL_14908750, EPI_ISL_14908751, EPI_ISL_14908752, EPI_ISL_14908753, EPI_ISL_14908754, EPI_ISL_14908755, EPI_ISL_14908756, EPI_ISL_14908757, EPI_ISL_14908758, EPI_ISL_14908759, EPI_ISL_14908760, EPI_ISL_14908761, EPI_ISL_14908762, EPI_ISL_14908763 | | | |
| see above | Instituto Adolfo Lutz - Vector Transmission Diseases | Instituto Adolfo Lutz - Strategic Laboratory | Claudio Tavares Sacchi, Karoline Rodrigues Campos, Marlon Benedito Nascimento Santos, Alex Domingos Reis, Juliana da Silva Nogueira, Adriana Yurika Maeda |
| EPI_ISL_14908764 | LACEN do Estado do Amazonas | Instituto Adolfo Lutz - Strategic Laboratory | Claudio Tavares Sacchi, Karoline Rodrigues Campos, Marlon Benedito Nascimento Santos, Alex Domingos Reis, Juliana da Silva Nogueira, Adriana Yurika Maeda |
| EPI_ISL_14908765, EPI_ISL_14908766, EPI_ISL_14908767, EPI_ISL_14908768, EPI_ISL_14908769, EPI_ISL_14908770, EPI_ISL_14908771, EPI_ISL_14908772, EPI_ISL_14908773, EPI_ISL_14908774, EPI_ISL_14908775, EPI_ISL_14908776, EPI_ISL_14908777, EPI_ISL_14908778, EPI_ISL_14908779, EPI_ISL_14908780, EPI_ISL_14908781, EPI_ISL_14908782, EPI_ISL_14908783, EPI_ISL_14908784, EPI_ISL_14908785, EPI_ISL_14908786, EPI_ISL_14908787, EPI_ISL_14908788, EPI_ISL_14908789, EPI_ISL_14908790, EPI_ISL_14908791, EPI_ISL_14908792, EPI_ISL_14908793, EPI_ISL_14908794, EPI_ISL_14908795, EPI_ISL_14908796, EPI_ISL_14908797, EPI_ISL_14908798, EPI_ISL_14908799, EPI_ISL_14908800, EPI_ISL_14908801, EPI_ISL_14908802, EPI_ISL_14908803, EPI_ISL_14908804, EPI_ISL_14908805, EPI_ISL_14908806, EPI_ISL_14908807, EPI_ISL_14908808, EPI_ISL_14908809, EPI_ISL_14908810, EPI_ISL_14908811, EPI_ISL_14908812, EPI_ISL_14908813, EPI_ISL_14908814, EPI_ISL_14908815, EPI_ISL_14908816, EPI_ISL_14908817, EPI_ISL_14908818, EPI_ISL_14908819, EPI_ISL_14908820, EPI_ISL_14908821, EPI_ISL_14908822, EPI_ISL_14908823, EPI_ISL_14908824, EPI_ISL_14908825, EPI_ISL_14908826, EPI_ISL_14908827, EPI_ISL_14908828, EPI_ISL_14908829, EPI_ISL_14908830, EPI_ISL_14908831, EPI_ISL_14908832, EPI_ISL_14908833, EPI_ISL_14908834, EPI_ISL_14908835, EPI_ISL_14908836, EPI_ISL_14908837, EPI_ISL_14908838, EPI_ISL_14908839, EPI_ISL_14908840, EPI_ISL_14908841, EPI_ISL_14908842, EPI_ISL_14908843, EPI_ISL_14908844, EPI_ISL_14908845, EPI_ISL_14908846, EPI_ISL_14908847, EPI_ISL_14908848, EPI_ISL_14908849, EPI_ISL_14908850, EPI_ISL_14908851, EPI_ISL_14908852, EPI_ISL_14908853, EPI_ISL_14908854, EPI_ISL_14908855, EPI_ISL_14908856, EPI_ISL_14908857, EPI_ISL_14908858, EPI_ISL_14908859, EPI_ISL_14908860, EPI_ISL_14908861, EPI_ISL_14908862, EPI_ISL_14908863, EPI_ISL_14908864, EPI_ISL_14908865, EPI_ISL_14908866, EPI_ISL_14908867, EPI_ISL_14908868, EPI_ISL_14908869, EPI_ISL_14908870, EPI_ISL_14908871, EPI_ISL_14908872, EPI_ISL_14908873, EPI_ISL_14908874, EPI_ISL_14908875, EPI_ISL_14908876, EPI_ISL_14908877, EPI_ISL_14908878, EPI_ISL_14908879, EPI_ISL_14908880, EPI_ISL_14908881, EPI_ISL_14908882, EPI_ISL_14908883, EPI_ISL_14908884, EPI_ISL_14908885, EPI_ISL_14908886, EPI_ISL_14908887, EPI_ISL_14908888, EPI_ISL_14908889, EPI_ISL_14908890, EPI_ISL_14908891, EPI_ISL_14908892, EPI_ISL_14908893, EPI_ISL_14908894, EPI_ISL_14908895, EPI_ISL_14908896, EPI_ISL_14908897, EPI_ISL_14908898, EPI_ISL_14908899, EPI_ISL_14908900, EPI_ISL_14908901, EPI_ISL_14908902, EPI_ISL_14908903 | | | |
| see above | Instituto Adolfo Lutz - Vector Transmission Diseases | Instituto Adolfo Lutz - Strategic Laboratory | Claudio Tavares Sacchi, Karoline Rodrigues Campos, Marlon Benedito Nascimento Santos, Alex Domingos Reis, Juliana da Silva Nogueira, Adriana Yurika Maeda |
| EPI_ISL_14908904 | Dengue Branch, Centers for Disease Control and Prevention | Dengue Branch, Centers for Disease Control and Prevention | Santiago,G.A., Gonzalez-Morales,G., Charriez,K., Flores,B., Stanek,D., Morrison,A. and Munoz-Jordan,J.L. |
| EPI_ISL_14916174, EPI_ISL_14916175, EPI_ISL_14916176, EPI_ISL_14916177, EPI_ISL_14916178, EPI_ISL_14916179, EPI_ISL_14916180, EPI_ISL_14916181, EPI_ISL_14916182, EPI_ISL_14916183, EPI_ISL_14916184, EPI_ISL_14916185 | | | |
| see above | National Institute of Cholera and Enteric Diseases | National Institute of Cholera and Enteric Diseases | Sadhukhan,P.C. and Verma,P. |
| EPI_ISL_14931901, EPI_ISL_14932897 | Punjab University | Punjab University | Munir,R., Rafique,S., Ali,A., Idrees,M. and Amin,I. |
| EPI_ISL_14946121 | Instituto Nacional de Salud | Instituto Nacional de Salud | Rosales-Munar,A., Rivera,J., Rengifo,A., Diaz-Herrera,T.H., Parra,E., Laiton-Donato,K., Alvarez-Diaz,D.A., Usme-Ciro,J.A. and Caldas,M.L. |
| EPI_ISL_15017943, EPI_ISL_15017944, EPI_ISL_15018597, EPI_ISL_15018598, EPI_ISL_15019070, EPI_ISL_15019071, EPI_ISL_15019072 | Oxford University Clinical Research Unit | Oxford University Clinical Research Unit | Cao,T.T., Dong,T.H., Nguyen,N.M., Huynh,D.T., Ho,C.Q., Nguyen,P.T., Nguyen,C.V., Phan,Q.T., Huynh,T.T., Luong,T.T., Nguyen,H.V., Duong,T.B., Duong,K.T., Tran,V.T. and Yacoub,S. |
| EPI_ISL_15052681, EPI_ISL_15052682, EPI_ISL_15052683, EPI_ISL_15052684, EPI_ISL_15052685, EPI_ISL_15052686 | Instituto Nacional de Salud | Instituto Nacional de Salud | Rosales-Munar,A., Rivera,J., Rengifo,A., Diaz-Herrera,T.H., Parra,E., Laiton-Donato,K., Alvarez-Diaz,D.A., Usme-Ciro,J.A. and Caldas,M.L. |
| EPI_ISL_15052687 | Instituto Nacional de Salud | Instituto Nacional de Salud | Rosales-Munar,A., Rivera,J., Rengifo,A., Diaz-Herrera,T., Parra,E., Laiton-Donato,K., Alvarez-Diaz,D., Usme-Ciro,J. and Caldas,M. |
| EPI_ISL_15067517, EPI_ISL_15067890, EPI_ISL_15068082, EPI_ISL_15068185, EPI_ISL_15068186, EPI_ISL_15068187, EPI_ISL_15068188, EPI_ISL_15068201, EPI_ISL_15072557, EPI_ISL_15072558, EPI_ISL_15072559, EPI_ISL_15072669, EPI_ISL_15072670, EPI_ISL_15072713 | | | |
| see above | Center for Disease Control and Prevention | Center for Disease Control and Prevention | Guo,L., Xu,M., Chen,Q., Yang,B. and Wang,J. |
| EPI_ISL_15078295, EPI_ISL_15078717, EPI_ISL_15078718, EPI_ISL_15078719, EPI_ISL_15078720, EPI_ISL_15078958, EPI_ISL_15079570, EPI_ISL_15079848, EPI_ISL_15079849, EPI_ISL_15079850, EPI_ISL_15079851, EPI_ISL_15082327, EPI_ISL_15083530, EPI_ISL_15083531, EPI_ISL_15083586, EPI_ISL_15083636, EPI_ISL_15083681, EPI_ISL_15083926, EPI_ISL_15083929, EPI_ISL_15083935, EPI_ISL_15084659, EPI_ISL_15085326, EPI_ISL_15085528, EPI_ISL_15085529, EPI_ISL_15088380, EPI_ISL_15088390, EPI_ISL_15088514, EPI_ISL_15088515, EPI_ISL_15088516, EPI_ISL_15088517, EPI_ISL_15088518, EPI_ISL_15088519, EPI_ISL_15088520, EPI_ISL_15088521, EPI_ISL_15088522, EPI_ISL_15088523, EPI_ISL_15088524, EPI_ISL_15088525, EPI_ISL_15088526 | | | |
| see above | Oxford University Clinical Research Unit | Oxford University Clinical Research Unit | Duong,K.T., Tran,V.T., Dong,T.H., Farrar,J., Tran,N.V., Tran,T.T., Guzman,M.G., Wills,B.A. and Simmons,C.P. |
| EPI_ISL_15292346, EPI_ISL_15292347, EPI_ISL_15292424, EPI_ISL_15292425, EPI_ISL_15292426 | Institute for Medical Research | Infectious Disease Research Centre | Suppiah,J., Ramly,M.N. and Robert,F. |
| EPI_ISL_15292427 | Institute for Medical Research, National Institutes of Health | Institute for Medical Research, National Institutes of Health | Suppiah,J., Ramly,M.N. and Robert,F. |
| EPI_ISL_15292428, EPI_ISL_15292429 | Institute for Medical Research | Infectious Disease Research Centre | Suppiah,J., Ramly,M.N. and Robert,F. |
| EPI_ISL_15292430, EPI_ISL_15292431, EPI_ISL_15292432, EPI_ISL_15292433 | Institute for Medical Research, National Institutes of Health | Institute for Medical Research, National Institutes of Health | Suppiah,J., Ramly,M.N. and Robert,F. |
| EPI_ISL_15292434 | Institute for Medical Research | Infectious Disease Research Centre | Suppiah,J., Ramly,M.N. and Robert,F. |
| EPI_ISL_15314344 | Institute for Medical Research, National Institutes of Health | Institute for Medical Research, National Institutes of Health | Suppiah,J., Ramly,M.N. and Robert,F. |
| EPI_ISL_15381632, EPI_ISL_15381633, EPI_ISL_15381634 | Bangalore University | Bangalore University | Doddamane,M. and Ramachandra,S. |
| EPI_ISL_15416610, EPI_ISL_15416611, EPI_ISL_15416612, EPI_ISL_15416613, EPI_ISL_15416928, EPI_ISL_15416929, EPI_ISL_15416930, EPI_ISL_15416931, EPI_ISL_15416983, EPI_ISL_15416984, EPI_ISL_15416985, EPI_ISL_15417893, EPI_ISL_15417894, EPI_ISL_15417895, EPI_ISL_15417896, EPI_ISL_15417897, EPI_ISL_15417898 | | | |
| see above | Hangzhou Center for Disease Control and Prevention | Hangzhou Center for Disease Control and Prevention | Yu,H. |
| EPI_ISL_15417899, EPI_ISL_15417900 | Shenzhen Center for Disease Control and Prevention | Shenzhen Center for Disease Control and Prevention | Yang,F. |
| EPI_ISL_15417901, EPI_ISL_15417902, EPI_ISL_15417903, EPI_ISL_15417904, EPI_ISL_15417905, EPI_ISL_15417906, EPI_ISL_15417907, EPI_ISL_15417908, EPI_ISL_15417909, EPI_ISL_15417910, EPI_ISL_15417911, EPI_ISL_15417912, EPI_ISL_15417913, EPI_ISL_15417914, EPI_ISL_15417915, EPI_ISL_15417916, EPI_ISL_15417917, EPI_ISL_15417918, EPI_ISL_15417919, EPI_ISL_15417920, EPI_ISL_15417921, EPI_ISL_15417922, EPI_ISL_15417923, EPI_ISL_15417924, EPI_ISL_15417925, EPI_ISL_15417926, EPI_ISL_15417927, EPI_ISL_15417928, EPI_ISL_15417929, EPI_ISL_15417930, EPI_ISL_15417931, EPI_ISL_15417932, EPI_ISL_15417933, EPI_ISL_15417934, EPI_ISL_15417935, EPI_ISL_15417936, EPI_ISL_15417937, EPI_ISL_15417938, EPI_ISL_15417939, EPI_ISL_15417940, EPI_ISL_15417941, EPI_ISL_15417942, EPI_ISL_15417943, EPI_ISL_15417944, EPI_ISL_15417945, EPI_ISL_15417946, EPI_ISL_15417947, EPI_ISL_15419452, EPI_ISL_15420000, EPI_ISL_15420001, EPI_ISL_15420002, EPI_ISL_15420003, EPI_ISL_15420004, EPI_ISL_15420005, EPI_ISL_15420006, EPI_ISL_15420007, EPI_ISL_15420008 | | | |
| see above | Hangzhou Center for Disease Control and Prevention | Hangzhou Center for Disease Control and Prevention | Yu,H. |
| EPI_ISL_15572066 | Jashore University of Science and Technology | Jashore University of Science and Technology | Rahman,M., Sarker,M.M.H. and Khan,S. |
| EPI_ISL_15576656, EPI_ISL_15576657 | Jashore University of Science and Technology | Jashore University of Science and Technology | Rahman,M.S., Sarker,M.M.H. and Khan,S. |
| EPI_ISL_15609136, EPI_ISL_15609137, EPI_ISL_15609138, EPI_ISL_15609139 | National Institute of Cholera and Enteric Diseases | National Institute of Cholera and Enteric Diseases | Verma,P. and Sadhukhan,D.P. |
| EPI_ISL_15609140, EPI_ISL_15609141, EPI_ISL_15609145, EPI_ISL_15609146, EPI_ISL_15609147 | ICMR- National Institute of Cholera and Enteric Diseases | ICMR- National Institute of Cholera and Enteric Diseases | Verma,P. and Sadhukhan,D.P. |
| EPI_ISL_15609148, EPI_ISL_15609149, EPI_ISL_15609150 | National Institute of Cholera and Enteric Diseases | National Institute of Cholera and Enteric Diseases | Verma,P. and Sadhukhan,D.P. |
| EPI_ISL_15693125, EPI_ISL_15693223, EPI_ISL_15693224, EPI_ISL_15693225, EPI_ISL_15693304, EPI_ISL_15693396, EPI_ISL_15693935, EPI_ISL_15693936, EPI_ISL_15693937, EPI_ISL_15693938, EPI_ISL_15694080, EPI_ISL_15694081, EPI_ISL_15694082, EPI_ISL_15694083, EPI_ISL_15694084, EPI_ISL_15694085, EPI_ISL_15694086, EPI_ISL_15694087, EPI_ISL_15694088, EPI_ISL_15694089, EPI_ISL_15694091, EPI_ISL_15694092, EPI_ISL_15694093, EPI_ISL_15694094, EPI_ISL_15694095, EPI_ISL_15694096, EPI_ISL_15694097, EPI_ISL_15694098, EPI_ISL_15694099, EPI_ISL_15694100, EPI_ISL_15694101, EPI_ISL_15694102, EPI_ISL_15694103, EPI_ISL_15694104, EPI_ISL_15694105, EPI_ISL_15694106, EPI_ISL_15694107, EPI_ISL_15694527, EPI_ISL_15694528 | | | |
| see above | Oxford University Clinical Research Unit | Oxford University Clinical Research Unit | Duong,K.T.H., Tran,V.T., Dong,T.H.T., Tran,N.V., Tran,T., Wills,B.A. and Simmons,C.P. |
| EPI_ISL_15732740, EPI_ISL_15733266, EPI_ISL_15733891, EPI_ISL_15737306, EPI_ISL_15738309, EPI_ISL_15739460, EPI_ISL_15739461, EPI_ISL_15739462 | National Institute of Health | National Institute of Health | Umair,M., Rehman,Z., Haider,S.A., Salman,M. and Ikram,A. |
| EPI_ISL_15739463, EPI_ISL_15739464, EPI_ISL_15739782, EPI_ISL_15739783, EPI_ISL_15739784, EPI_ISL_15739785, EPI_ISL_15739786, EPI_ISL_15739787, EPI_ISL_15739788, EPI_ISL_15739789, EPI_ISL_15739790, EPI_ISL_15739791, EPI_ISL_15739792 | | | |
| see above | Department of Virology | Department of Virology | Sharma,V., Satapathy,P., Kang,M., Sarkar,S., Dhingra,S., Bora,I., Kaur,K., Goel,K., Aggarwal,A., Arora,N. and Ratho,R.K. |
| EPI_ISL_15823980 | Institute of Medical Sciences | Institute of Medical Sciences | Dinkar,A., Singh,J. and Prakash,P. |
| EPI_ISL_15917022, EPI_ISL_15917023, EPI_ISL_15917114, EPI_ISL_15917153, EPI_ISL_15917154, EPI_ISL_15917736, EPI_ISL_15917737, EPI_ISL_15917798, EPI_ISL_15917799, EPI_ISL_15917800, EPI_ISL_15917801 | | | |
| see above | National Public Health Center | National Public Health Center | Nagy,O. and Takacs,M. |
| EPI_ISL_15923145, EPI_ISL_15926913, EPI_ISL_15926989 | National Institute of Health | National Institute of Health | Umair,M., Rehman,Z., Haider,S.A., Salman,M. and Ikram,A. |
| EPI_ISL_15952840, EPI_ISL_15952841, EPI_ISL_15952842 | Bangalore University | Bangalore University | Doddamane,M. and Ramachandra,S. |
| EPI_ISL_15997588, EPI_ISL_15997589, EPI_ISL_15997590, EPI_ISL_15997591, EPI_ISL_15997592, EPI_ISL_15997593, EPI_ISL_15997594, EPI_ISL_15997595, EPI_ISL_15997596, EPI_ISL_15997597, EPI_ISL_15997598, EPI_ISL_15997599, EPI_ISL_15997600 | | | |
| see above | Fundação Oswaldo Cruz - Instituto Leônidas e | Fundação Oswaldo Cruz - Instituto Leônidas e Maria | Nascimento,V.A., Souza,V.C., Goncalves,L., Nascimento,F., Mejia,M., Silva,D. and Naveca,F.G. |

| | | | |
|---|---|---|---|
| | Maria Deane (FIOCRUZ) | Deane (FIOCRUZ) | |
| EPI_ISL_16014609, EPI_ISL_16014610, EPI_ISL_16014676, EPI_ISL_16014677, EPI_ISL_16015137, EPI_ISL_16015138 | King George's Medical University | King George's Medical University | Prakash,O., Khan,D.N., Verma,A.K., Mishra,V.K., Srivastava,A.K. and Jain,A. |
| EPI_ISL_16072648 | University of Sao Paulo | University of Sao Paulo | La Serra,L., Cazarotti,R.L.S., Scrich,V.M., Beuno,L.M., Figueiredo,L.T.M., Santos,G.S., de Carvalho,A.N., Amaral,R. and Cassiano,M.H.A. |
| EPI_ISL_16073484, EPI_ISL_16073485 | Guangzhou Center for Disease Control and Prevention | Guangzhou Center for Disease Control and Prevention | Su,W., Cao,Y. and Lu,W. |
| EPI_ISL_16092157, EPI_ISL_16092158, EPI_ISL_16092166, EPI_ISL_16092323, EPI_ISL_16092324, EPI_ISL_16092325, EPI_ISL_16092326, EPI_ISL_16092327, EPI_ISL_16092328, EPI_ISL_16092329, EPI_ISL_16092330, EPI_ISL_16092331, EPI_ISL_16092332, EPI_ISL_16092333, EPI_ISL_16092334, EPI_ISL_16092335, EPI_ISL_16092336, EPI_ISL_16092337, EPI_ISL_16092338, EPI_ISL_16092339, EPI_ISL_16092340, EPI_ISL_16092341, EPI_ISL_16092342, EPI_ISL_16092343, EPI_ISL_16092344, EPI_ISL_16092345, EPI_ISL_16092346, EPI_ISL_16092347 | | | |
| see above | Oxford University Clinical Research Unit | Oxford University Clinical Research Unit | Tran,V.T., Duong,K.T.H., Nguyen,N.M., Huynh,D.T.L., Ho,C.Q., Nguyen,P.T., Nguyen,C.V.V., Phan,Q.T., Huynh,T., Luong,T.H., Nguyen,H.V., Duong,T.B., Cao,T., Dong,T.H., Nguyen,H.V., McBride,A., Wills,B., Simmons,C.P. and Yacoub,S. |
| EPI_ISL_16092348, EPI_ISL_16092349, EPI_ISL_16092350 | National Institutes of Health (NIH) | National Institutes of Health (NIH) - Turning Discovery into Health | Bohl,J.A., Lay,S., Chea,S., Ahyong,V., Parker,D.M., Gallagher,S., Fintzi,J., Man,S., Ponce,A., Sreng,S., Kong,D., Oliveira,F., Kalantar,K., Tan,M., Fahsbender,L., Sheu,J., Neff,N., Detweiler,A.M., Yek,C., Ly,S., Sath,R., Huch,C., Kry,H., Leang,R., Huy,R., Lon,C., Tato,C.M., DeRisi,J.L. and Manning,J.E. |
| EPI_ISL_16178334, EPI_ISL_16178483, EPI_ISL_16179152, EPI_ISL_16179858, EPI_ISL_16179897 | General Sir John Kotelawala Defence University, Faculty of Medicine | General Sir John Kotelawala Defence University, Faculty of Medicine | Dassanayake,A.K.K., De Silva,A.D., Abeygoonawardena,H., Kariyawasam,J.C., Chathuranga,T., Sundralingam,T., Dilshani,P., Gunasekara,H., Premawansa,G., Premawansa,S., Weiskopf,D., Sette,A. and Punchihewa,C. |
| EPI_ISL_16191497 | University of Hyderabad | University of Hyderabad | Maisnam,D. and Musturi,V. |
| EPI_ISL_16212171 | General Sir John Kotelawala Defence University, Faculty of Medicine | General Sir John Kotelawala Defence University, Faculty of Medicine | Abeygoonawardena,H., de Silva,A.D., Wijesinghe,N., Navaratne,V., Dilshani,P., Sundralingam,T. and Punchihewa,C. |
| EPI_ISL_16212210, EPI_ISL_16212211, EPI_ISL_16212212, EPI_ISL_16215353, EPI_ISL_16215354, EPI_ISL_16215355, EPI_ISL_16215356 | Universidade Federal de Mato Grosso (UFMT) | Universidade Federal de Mato Grosso (UFMT) | Santos,M.A.M., Pavon,J.A.R., Viniski,A.E., Souza,C.L.C., Oliveira,E.C., Medeiros,D.B.A., Silva,S.P., Nunes,M.R.T. and Slhessarenko,R.D. |
| EPI_ISL_16222604 | General Sir John Kotelawala Defence University, Faculty of Medicine | General Sir John Kotelawala Defence University, Faculty of Medicine | Abeygoonawardena,H., De Silva,A.D., Dassanayake,A.K.K., Kariyawasam,J.C., Chathuranga,T., Sundralingam,T., Dilshani,P., Gunasekara,H., Premawansa,G., Premawansa,S., Weiskopf,D., Sette,A. and Punchihewa,C. |
| EPI_ISL_16222605 | General Sir John Kotelawala Defence University, Faculty of Medicine | General Sir John Kotelawala Defence University, Faculty of Medicine | De Silva,A.D., Dassanayake,A.K.K., Abeygoonawardena,H., Kariyawasam,J.C., Chathuranga,T., Sundralingam,T., Dilshani,P., Gunasekara,H., Premawansa,G., Premawansa,S., Weiskopf,D., Sette,A. and Punchihewa,C. |
| EPI_ISL_16222606 | General Sir John Kotelawala Defence University, Faculty of Medicine | General Sir John Kotelawala Defence University, Faculty of Medicine | Abeygoonawardena,H., De Silva,A.D., Dassanayake,A.K.K., Kariyawasam,J.C., Chathuranga,T., Sundralingam,T., Dilshani,P., Gunasekara,H., Premawansa,G., Premawansa,S., Weiskopf,D., Sette,A. and Punchihewa,C. |
| EPI_ISL_16222607, EPI_ISL_16222608, EPI_ISL_16222609, EPI_ISL_16222610 | General Sir John Kotelawala Defence University, Faculty of Medicine | General Sir John Kotelawala Defence University, Faculty of Medicine | De Silva,A.D., Dassanayake,A.K.K., Abeygoonawardena,H., Kariyawasam,J.C., Chathuranga,T., Sundralingam,T., Dilshani,P., Gunasekara,H., Premawansa,G., Premawansa,S., Weiskopf,D., Sette,A. and Punchihewa,C. |
| EPI_ISL_16222611, EPI_ISL_16222612, EPI_ISL_16222613 | General Sir John Kotelawala Defence University, Faculty of Medicine | General Sir John Kotelawala Defence University, Faculty of Medicine | Abeygoonawardena,H., De Silva,A.D., Dassanayake,A.K.K., Kariyawasam,J.C., Chathuranga,T., Sundralingam,T., Dilshani,P., Gunasekara,H., Premawansa,G., Premawansa,S., Weiskopf,D., Sette,A. and Punchihewa,C. |
| EPI_ISL_16282284 | Universidade Federal de Mato Grosso (UFMT) | Universidade Federal de Mato Grosso (UFMT) | Santos,M.A.M., Pavon,J.A.R., Viniski,A.E., Souza,C.L.C., Oliveira,E.C., Medeiros,D.B.A., Silva,S.P., Nunes,M.R.T. and Slhessarenko,R.D. |
| EPI_ISL_16287706, EPI_ISL_16287707, EPI_ISL_16287708, EPI_ISL_16287709, EPI_ISL_16287710, EPI_ISL_16287711, EPI_ISL_16287712, EPI_ISL_16287713, EPI_ISL_16287714 | All India Institute of Medical Sciences Rishikesh | All India Institute of Medical Sciences Rishikesh | Badoni,G., Gupta,P., Ratho,R.K., Kaistha,N. and Pai,M.O. |
| EPI_ISL_16287715 | All India Institute of Medical Sciences Rishikesh | All India Institute of Medical Sciences Rishikesh | Badoni,G. |
| EPI_ISL_16287716, EPI_ISL_16299723, EPI_ISL_16299844, EPI_ISL_16299903, EPI_ISL_16299904, EPI_ISL_16299905 | All India Institute of Medical Sciences Rishikesh | All India Institute of Medical Sciences Rishikesh | Badoni,G., Gupta,P., Ratho,R.K., Kaistha,N. and Pai,M.O. |
| EPI_ISL_16312445, EPI_ISL_16312645, EPI_ISL_16312646, EPI_ISL_16312647, EPI_ISL_16312648, EPI_ISL_16312649, EPI_ISL_16312718, EPI_ISL_16312739, EPI_ISL_16312740, EPI_ISL_16312741, EPI_ISL_16312742, EPI_ISL_16312743, EPI_ISL_16312744, EPI_ISL_16312745, EPI_ISL_16312746, EPI_ISL_16312747, EPI_ISL_16312748, EPI_ISL_16312749, EPI_ISL_16312750, EPI_ISL_16312751, EPI_ISL_16312752, EPI_ISL_16312753, EPI_ISL_16312754, EPI_ISL_16312755, EPI_ISL_16312756, EPI_ISL_16312757, EPI_ISL_16312758, EPI_ISL_16312759, EPI_ISL_16312760, EPI_ISL_16312761, EPI_ISL_16312762 | | | |
| see above | Centers for Disease Control and Prevention | Centers for Disease Control and Prevention | Santiago,G.A., Gonzalez-Morales,G., Flores,B., Charriez,K.N., Reller,M., Matute,A., Reyes,Y., Centeno-Cuadra,E., Bucardo,F. and Munoz-Jordan,J.L. |
| EPI_ISL_16350698, EPI_ISL_16350704, EPI_ISL_16350805, EPI_ISL_16352470 | All India Institute of Medical Sciences Rishikesh | All India Institute of Medical Sciences Rishikesh | Badoni,G., Gupta,P., Ratho,R.K., Kaistha,N. and Pai,M.O. |
| EPI_ISL_16480244 | Universidade Federal de Mato Grosso (UFMT) | Universidade Federal de Mato Grosso (UFMT) | Santos,M.A.M., Pavon,J.A.R., Viniski,A.E., Souza,C.L.C., Oliveira,E.C., Medeiros,D.B.A., Silva,S.P., Nunes,M.R.T. and Slhessarenko,R.D. |
| EPI_ISL_16482531 | Federal University of Mato Grosso | Federal University of Mato Grosso | Santos, M.A.M., Pavon,J.A.R., Viniski,A.E., Souza,C.L.C., Oliveira,E.C., Medeiros,D.B.A., Silva,S.P., Nunes,M.R.T. and Slhessarenko, R.D. |
| EPI_ISL_17410726 | Centers for Disease Control and Prevention, Arboviral Diseases Branch | Centers for Disease Control and Prevention, Arboviral Diseases Branch | Hughes,H.R. and Russell,B.J. |
| EPI_ISL_17448340, EPI_ISL_17448341, EPI_ISL_17448342, EPI_ISL_17448343, EPI_ISL_17448344, EPI_ISL_17448345, EPI_ISL_17448346, EPI_ISL_17448347 | Unit of Omics, Vector Control Research Centre | Unit of Omics, Vector Control Research Centre | Sankari,T., Jayasree,A., Rameela Sanya,K., Varghese,B., Kumar,M., Kumar,A. and Niranjan,R. |
| EPI_ISL_17448597 | Virginia-Maryland College of Veterinary Medicine, Biomedical Sciences and Pathobiology, | Virginia-Maryland College of Veterinary Medicine, Biomedical Sciences and Pathobiology, | Marano,J.M. and Weger-Lucarelli,J. |
| EPI_ISL_17450850 | Infection and Immunology, Translational Health Science and Technology Institute | Infection and Immunology, Translational Health Science and Technology Institute | Maurya,R., Agrawal,T., Saraswat,S., Ananthraj,A., Chandele,A., Kumar,S., Kanakan,A., Chattopadhyay,P., Swaminathan,A., Saifi,S., Shashi,P., Pandey,R. and Medigeshi,G.R. |
| EPI_ISL_17450851 | Infection and Immunology, Translational Health Science and Technology Institute | Infection and Immunology, Translational Health Science and Technology Institute | Medigeshi,G., Pandey,R., Agrawal,T., Saraswat,S., Ananthraj,A., Chandele,A., Maurya,R., Kanakan,A., Chattopadhyay,P., Swaminathan,A. and Saifi,S. |
| EPI_ISL_17450852 | Infection and Immunology, Translational Health Science and Technology Institute | Infection and Immunology, Translational Health Science and Technology Institute | Maurya,R., Agrawal,T., Saraswat,S., Ananthraj,A., Chandele,A., Kumar,S., Kanakan,A., Chattopadhyay,P., Swaminathan,A., Saifi,S., Shashi,P., Pandey,R. and Medigeshi,G.R. |
| EPI_ISL_17450853 | Infection and Immunology, Translational Health Science and Technology Institute | Infection and Immunology, Translational Health Science and Technology Institute | Agrawal,T., Saraswat,S., Shashi,P., Ananthraj,A., Kumar,S., Chandele,A., Lodha,R., Singh,B. and Medigeshi,G.R. |
| EPI_ISL_17450854 | Infection and Immunology, Translational Health Science and Technology Institute | Infection and Immunology, Translational Health Science and Technology Institute | Maurya,R., Agrawal,T., Saraswat,S., Ananthraj,A., Chandele,A., Kumar,S., Kanakan,A., Chattopadhyay,P., Swaminathan,A., Saifi,S., Shashi,P., Pandey,R. and Medigeshi,G.R. |
| EPI_ISL_17450855 | Infection and Immunology, Translational Health Science and Technology Institute | Infection and Immunology, Translational Health Science and Technology Institute | Agrawal,T., Saraswat,S., Shashi,P., Ananthraj,A., Kumar,S., Chandele,A., Lodha,R., Singh,B. and Medigeshi,G.R. |
| EPI_ISL_17450856 | Infection and Immunology, Translational Health Science and Technology Institute | Infection and Immunology, Translational Health Science and Technology Institute | Maurya,R., Agrawal,T., Saraswat,S., Ananthraj,A., Chandele,A., Kumar,S., Kanakan,A., Chattopadhyay,P., Swaminathan,A., Saifi,S., Shashi,P., Pandey,R. and Medigeshi,G.R. |
| EPI_ISL_17450857, EPI_ISL_17450858 | Infection and Immunology, Translational Health Science and Technology Institute | Infection and Immunology, Translational Health Science and Technology Institute | Medigeshi,G., Pandey,R., Agrawal,T., Saraswat,S., Ananthraj,A., Chandele,A., Maurya,R., Kanakan,A., Chattopadhyay,P., Swaminathan,A. and Saifi,S. |
| EPI_ISL_17450859 | Infection and Immunology, Translational Health Science and Technology Institute | Infection and Immunology, Translational Health Science and Technology Institute | Agrawal,T., Saraswat,S., Shashi,P., Ananthraj,A., Kumar,S., Chandele,A., Lodha,R., Singh,B. and Medigeshi,G.R. |
| EPI_ISL_17450860 | Infection and Immunology, Translational Health Science and Technology Institute | Infection and Immunology, Translational Health Science and Technology Institute | Maurya,R., Agrawal,T., Saraswat,S., Ananthraj,A., Chandele,A., Kumar,S., Kanakan,A., Chattopadhyay,P., Swaminathan,A., Saifi,S., Shashi,P., Pandey,R. and Medigeshi,G.R. |
| EPI_ISL_17450861 | Infection and Immunology, Translational Health Science and Technology Institute | Infection and Immunology, Translational Health Science and Technology Institute | Agrawal,T., Saraswat,S., Shashi,P., Ananthraj,A., Kumar,S., Chandele,A., Lodha,R., Singh,B. and Medigeshi,G.R. |
| EPI_ISL_17450862, EPI_ISL_17450863 | Infection and Immunology, Translational Health Science and Technology Institute | Infection and Immunology, Translational Health Science and Technology Institute | Maurya,R., Agrawal,T., Saraswat,S., Ananthraj,A., Chandele,A., Kumar,S., Kanakan,A., Chattopadhyay,P., Swaminathan,A., Saifi,S., Shashi,P., Pandey,R. and Medigeshi,G.R. |
| EPI_ISL_17450864 | Infection and Immunology, Translational Health Science and Technology Institute | Infection and Immunology, Translational Health Science and Technology Institute | Agrawal,T., Saraswat,S., Shashi,P., Ananthraj,A., Kumar,S., Chandele,A., Lodha,R., Singh,B. and Medigeshi,G.R. |
| EPI_ISL_17450865 | Infection and Immunology, Translational Health Science and Technology Institute | Infection and Immunology, Translational Health Science and Technology Institute | Kumar,P., Shashi,P., Singh,B., Verma,C.K., Tripathi,A., Chandele,A., Lodha,R. and Medigeshi,G.R. |
| EPI_ISL_17450866 | Infection and Immunology, Translational Health Science and Technology Institute | Infection and Immunology, Translational Health Science and Technology Institute | Maurya,R., Agrawal,T., Saraswat,S., Ananthraj,A., Chandele,A., Kumar,S., Kanakan,A., Chattopadhyay,P., Swaminathan,A., Saifi,S., Shashi,P., Pandey,R. and Medigeshi,G.R. |
| EPI_ISL_17450867 | Infection and Immunology, Translational Health Science and Technology Institute | Infection and Immunology, Translational Health Science and Technology Institute | Maurya,R., Agrawal,T., Saraswat,S., Ananthraj,A., Chandele,A., Kumar,S., Kanakan,A., Chattopadhyay,P., Swaminathan,A., Saifi,S., Shashi,P., Singh,B., Pandey,R. and Medigeshi,G.R. |
| EPI_ISL_17450868, EPI_ISL_17450869, EPI_ISL_17450870, EPI_ISL_17450871 | Infection and Immunology, Translational Health Science and Technology Institute | Infection and Immunology, Translational Health Science and Technology Institute | Maurya,R., Agrawal,T., Saraswat,S., Ananthraj,A., Chandele,A., Kumar,S., Kanakan,A., Chattopadhyay,P., Swaminathan,A., Saifi,S., Shashi,P., Pandey,R. and Medigeshi,G.R. |
| EPI_ISL_17450872 | Infection and Immunology, Translational Health Science and Technology Institute | Infection and Immunology, Translational Health Science and Technology Institute | Maurya,R., Agrawal,T., Saraswat,S., Ananthraj,A., Chandele,A., Kumar,S., Kanakan,A., Chattopadhyay,P., Swaminathan,A., Saifi,S., Shashi,P., Singh,B., Pandey,R. and Medigeshi,G.R. |
| EPI_ISL_17450873, EPI_ISL_17450874, EPI_ISL_17450875 | Infection and Immunology, Translational Health Science and Technology Institute | Infection and Immunology, Translational Health Science and Technology Institute | Maurya,R., Agrawal,T., Saraswat,S., Ananthraj,A., Chandele,A., Kumar,S., Kanakan,A., Chattopadhyay,P., Swaminathan,A., Saifi,S., Shashi,P., Pandey,R. and Medigeshi,G.R. |

| | | | |
|---|---|---|---|
| EPI_ISL_17450876 | Infection and Immunology, Translational Health Science and Technology Institute | Infection and Immunology, Translational Health Science and Technology Institute | Agrawal,T., Saraswat,S., Shashi,P., Ananthraj,A., Kumar,S., Chandele,A., Lodha,R., Singh,B. and Medigeshi,G.R. |
| EPI_ISL_17450877, EPI_ISL_17450878 | Infection and Immunology, Translational Health Science and Technology Institute | Infection and Immunology, Translational Health Science and Technology Institute | Maurya,R., Agrawal,T., Saraswat,S., Ananthraj,A., Chandele,A., Kumar,S., Kanakan,A., Chattopadhyay,P., Swaminathan,A., Saifi,S., Shashi,P., Pandey,R. and Medigeshi,G.R. |
| EPI_ISL_17450879, EPI_ISL_17450880 | Infection and Immunology, Translational Health Science and Technology Institute | Infection and Immunology, Translational Health Science and Technology Institute | Agrawal,T., Saraswat,S., Shashi,P., Ananthraj,A., Kumar,S., Chandele,A., Lodha,R., Singh,B. and Medigeshi,G.R. |
| EPI_ISL_17450881 | Infection and Immunology, Translational Health Science and Technology Institute | Infection and Immunology, Translational Health Science and Technology Institute | Maurya,R., Agrawal,T., Saraswat,S., Ananthraj,A., Chandele,A., Kumar,S., Kanakan,A., Chattopadhyay,P., Swaminathan,A., Saifi,S., Shashi,P., Pandey,R. and Medigeshi,G.R. |
| EPI_ISL_17450882, EPI_ISL_17450883, EPI_ISL_17450884, EPI_ISL_17450885, EPI_ISL_17450886 | Infection and Immunology, Translational Health Science and Technology Institute | Infection and Immunology, Translational Health Science and Technology Institute | Agrawal,T., Saraswat,S., Shashi,P., Ananthraj,A., Kumar,S., Chandele,A., Lodha,R., Singh,B. and Medigeshi,G.R. |
| EPI_ISL_17450887, EPI_ISL_17450888, EPI_ISL_17450889, EPI_ISL_17450890 | Infection and Immunology, Translational Health Science and Technology Institute | Infection and Immunology, Translational Health Science and Technology Institute | Maurya,R., Agrawal,T., Saraswat,S., Ananthraj,A., Chandele,A., Kumar,S., Kanakan,A., Chattopadhyay,P., Swaminathan,A., Saifi,S., Shashi,P., Pandey,R. and Medigeshi,G.R. |
| EPI_ISL_17450891 | Infection and Immunology, Translational Health Science and Technology Institute | Infection and Immunology, Translational Health Science and Technology Institute | Agrawal,T., Saraswat,S., Shashi,P., Ananthraj,A., Kumar,S., Chandele,A., Lodha,R., Singh,B. and Medigeshi,G.R. |
| EPI_ISL_17450892 | Infection and Immunology, Translational Health Science and Technology Institute | Infection and Immunology, Translational Health Science and Technology Institute | Maurya,R., Agrawal,T., Saraswat,S., Ananthraj,A., Chandele,A., Kumar,S., Kanakan,A., Chattopadhyay,P., Swaminathan,A., Saifi,S., Shashi,P., Singh,B., Pandey,R. and Medigeshi,G.R. |
| EPI_ISL_17450893 | Infection and Immunology, Translational Health Science and Technology Institute | Infection and Immunology, Translational Health Science and Technology Institute | Maurya,R., Agrawal,T., Saraswat,S., Ananthraj,A., Chandele,A., Kumar,S., Kanakan,A., Chattopadhyay,P., Swaminathan,A., Saifi,S., Shashi,P., Pandey,R. and Medigeshi,G.R. |
| EPI_ISL_17450894, EPI_ISL_17450895, EPI_ISL_17450896, EPI_ISL_17450897, EPI_ISL_17450898, EPI_ISL_17450899 | Infection and Immunology, Translational Health Science and Technology Institute | Infection and Immunology, Translational Health Science and Technology Institute | Agrawal,T., Saraswat,S., Shashi,P., Ananthraj,A., Kumar,S., Chandele,A., Lodha,R., Singh,B. and Medigeshi,G.R. |
| EPI_ISL_17450900 | Infection and Immunology, Translational Health Science and Technology Institute | Infection and Immunology, Translational Health Science and Technology Institute | Maurya,R., Agrawal,T., Saraswat,S., Ananthraj,A., Chandele,A., Kumar,S., Shashi,P., Singh,B., Lodha,R., Pandey,R. and Medigeshi,G.R. |
| EPI_ISL_17450901, EPI_ISL_17450902 | Infection and Immunology, Translational Health Science and Technology Institute | Infection and Immunology, Translational Health Science and Technology Institute | Maurya,R., Agrawal,T., Saraswat,S., Ananthraj,A., Chandele,A., Kumar,S., Kanakan,A., Chattopadhyay,P., Swaminathan,A., Saifi,S., Shashi,P., Pandey,R. and Medigeshi,G.R. |
| EPI_ISL_17450903, EPI_ISL_17450904 | Infection and Immunology, Translational Health Science and Technology Institute | Infection and Immunology, Translational Health Science and Technology Institute | Agrawal,T., Saraswat,S., Shashi,P., Ananthraj,A., Kumar,S., Chandele,A., Lodha,R., Singh,B. and Medigeshi,G.R. |
| EPI_ISL_17450905 | Infection and Immunology, Translational Health Science and Technology Institute | Infection and Immunology, Translational Health Science and Technology Institute | Maurya,R., Agrawal,T., Saraswat,S., Ananthraj,A., Chandele,A., Kumar,S., Kanakan,A., Chattopadhyay,P., Swaminathan,A., Saifi,S., Shashi,P., Pandey,R. and Medigeshi,G.R. |
| EPI_ISL_17450906 | Infection and Immunology, Translational Health Science and Technology Institute | Infection and Immunology, Translational Health Science and Technology Institute | Agrawal,T., Saraswat,S., Shashi,P., Ananthraj,A., Kumar,S., Chandele,A., Lodha,R., Singh,B. and Medigeshi,G.R. |
| EPI_ISL_17450907 | Infection and Immunology, Translational Health Science and Technology Institute | Infection and Immunology, Translational Health Science and Technology Institute | Kumar,P., Shashi,P., Singh,B., Verma,C.K., Tripathi,A., Chandele,A., Lodha,R. and Medigeshi,G.R. |
| EPI_ISL_17450908, EPI_ISL_17450909 | Infection and Immunology, Translational Health Science and Technology Institute | Infection and Immunology, Translational Health Science and Technology Institute | Verma,C.K., Shashi,P., Singh,B., Kumar,P., Tripathi,A., Chandele,A., Lodha,R. and Medigeshi,G.R. |
| EPI_ISL_17450910, EPI_ISL_17450911 | Infection and Immunology, Translational Health Science and Technology Institute | Infection and Immunology, Translational Health Science and Technology Institute | Maurya,R., Agrawal,T., Saraswat,S., Ananthraj,A., Chandele,A., Kumar,S., Kanakan,A., Chattopadhyay,P., Swaminathan,A., Saifi,S., Shashi,P., Pandey,R. and Medigeshi,G.R. |
| EPI_ISL_17450912 | Infection and Immunology, Translational Health Science and Technology Institute | Infection and Immunology, Translational Health Science and Technology Institute | Agrawal,T., Saraswat,S., Shashi,P., Ananthraj,A., Kumar,S., Chandele,A., Lodha,R., Singh,B. and Medigeshi,G.R. |
| EPI_ISL_17450913 | Infection and Immunology, Translational Health Science and Technology Institute | Infection and Immunology, Translational Health Science and Technology Institute | Kumar,P., Shashi,P., Singh,B., Verma,C.K., Tripathi,A., Chandele,A., Lodha,R. and Medigeshi,G.R. |
| EPI_ISL_17498037, EPI_ISL_17498038 | Center for devices and radiological health - US Food and Drug Administration | Center for devices and radiological health - US Food and Drug Administration | Rios,M., Fares-Gusmao,R., Volkova,E., Grinev,A., Assis,F., Chancey,C., Ok,S., Rocha,B., Sippert,E., Tallon,L., Sadzewicz,L., Vavikolanu,K., Mehta,A., Aluvathingal,J., Nadendla,S., Yan,Y. and Sichtig,H. |
| EPI_ISL_17498073, EPI_ISL_17498074, EPI_ISL_17498075, EPI_ISL_17498076, EPI_ISL_17498077, EPI_ISL_17498078, EPI_ISL_17498079, EPI_ISL_17498080, EPI_ISL_17498081, EPI_ISL_17498082, EPI_ISL_17498083, EPI_ISL_17498084, EPI_ISL_17498085, EPI_ISL_17498086, EPI_ISL_17498087, EPI_ISL_17498088, EPI_ISL_17498089, EPI_ISL_17498090, EPI_ISL_17498091, EPI_ISL_17498092, EPI_ISL_17498093, EPI_ISL_17498094, EPI_ISL_17498095, EPI_ISL_17498096, EPI_ISL_17498097, EPI_ISL_17498098, EPI_ISL_17498099, EPI_ISL_17498100, EPI_ISL_17498101, EPI_ISL_17498102, EPI_ISL_17498103, EPI_ISL_17498104, EPI_ISL_17498105, EPI_ISL_17498106, EPI_ISL_17498107, EPI_ISL_17498108, EPI_ISL_17498109, EPI_ISL_17498110, EPI_ISL_17498111, EPI_ISL_17498112, EPI_ISL_17498113, EPI_ISL_17498114, EPI_ISL_17498115, EPI_ISL_17498116, EPI_ISL_17498117, EPI_ISL_17498118, EPI_ISL_17498119, EPI_ISL_17498120, EPI_ISL_17498121, EPI_ISL_17498122, EPI_ISL_17498123, EPI_ISL_17498124, EPI_ISL_17498125, EPI_ISL_17498126, EPI_ISL_17498127, EPI_ISL_17498128, EPI_ISL_17498129, EPI_ISL_17498130, EPI_ISL_17498131, EPI_ISL_17498132, EPI_ISL_17498133, EPI_ISL_17498134, EPI_ISL_17498135, EPI_ISL_17498136, EPI_ISL_17498137, EPI_ISL_17498138, EPI_ISL_17498139, EPI_ISL_17498140, EPI_ISL_17498141, EPI_ISL_17498142, EPI_ISL_17498143, EPI_ISL_17498144, EPI_ISL_17498145, EPI_ISL_17498146, EPI_ISL_17498147, EPI_ISL_17498148, EPI_ISL_17498149, EPI_ISL_17498150, EPI_ISL_17498151, EPI_ISL_17498152, EPI_ISL_17498153, EPI_ISL_17498154, EPI_ISL_17498155, EPI_ISL_17498156, EPI_ISL_17498157, EPI_ISL_17498158, EPI_ISL_17498159, EPI_ISL_17498160 | | | |
| see above | Centers for Disease Control and Prevention, Dengue Branch | Centers for Disease Control and Prevention, Dengue Branch | Santiago,G.A., Gonzalez-Morales,G., Flores,B., Charriez,K., Munoz-Jordan,J.L., Bunch,S., Cano,N., Jaber,R., Morrison,A., Panzera,C., Vergara,J., Zimler,R., Heberlein,L. and Kopp,E. |
| EPI_ISL_17498885 | Infection and Immunology, Translational Health Science and Technology Institute | Infection and Immunology, Translational Health Science and Technology Institute | Verma,C.K., Shashi,P., Singh,B., Kumar,P., Tripathi,A., Chandele,A., Lodha,R. and Medigeshi,G.R. |
| EPI_ISL_17498976, EPI_ISL_17498977, EPI_ISL_17498978, EPI_ISL_17498979, EPI_ISL_17498980, EPI_ISL_17498981, EPI_ISL_17498982, EPI_ISL_17498983, EPI_ISL_17498984, EPI_ISL_17498985, EPI_ISL_17498986, EPI_ISL_17498987, EPI_ISL_17498988 | | | |
| see above | Jilin Medical College | Jilin Medical College | Liang,C. |
| EPI_ISL_17499049 | National Institutes of Allergy and Infectious Diseases, Laboratory of Malaria and Vector Research | National Institutes of Allergy and Infectious Diseases, Laboratory of Malaria and Vector Research | Oum,M., Chea,S., Lay,S., Bohl,J.A., Yek,C. and Manning,J.E. |
| EPI_ISL_17499127, EPI_ISL_17499128, EPI_ISL_17499129, EPI_ISL_17499130, EPI_ISL_17499131, EPI_ISL_17499132, EPI_ISL_17499133, EPI_ISL_17499134, EPI_ISL_17499135, EPI_ISL_17499136, EPI_ISL_17499137, EPI_ISL_17499138, EPI_ISL_17499139, EPI_ISL_17499140, EPI_ISL_17499141, EPI_ISL_17499142, EPI_ISL_17499143, EPI_ISL_17499144, EPI_ISL_17499145, EPI_ISL_17499146, EPI_ISL_17499147, EPI_ISL_17499148, EPI_ISL_17499149, EPI_ISL_17499150, EPI_ISL_17499151, EPI_ISL_17499152, EPI_ISL_17499153, EPI_ISL_17499154, EPI_ISL_17499155, EPI_ISL_17499156, EPI_ISL_17499157, EPI_ISL_17499158, EPI_ISL_17499159, EPI_ISL_17499160, EPI_ISL_17499161, EPI_ISL_17499162, EPI_ISL_17499163, EPI_ISL_17499164, EPI_ISL_17499165, EPI_ISL_17499166, EPI_ISL_17499167, EPI_ISL_17499168, EPI_ISL_17499169, EPI_ISL_17499170 | | | |
| see above | U. S. Army Medical Component, Armed Forces Research Institute of Medical Sciences | U. S. Army Medical Component, Armed Forces Research Institute of Medical Sciences | Binay Thapa,B., Gyeltshen,S., Klungthong,C., Zangmo,S., Wangchuk,S., Chinnawirotpisan,P., Manasatienkij,W., Fernandez,S. and Jones,A.R. |
| EPI_ISL_17568981, EPI_ISL_17568982 | General Sir John Kotelawala Defence University | General Sir John Kotelawala Defence University | Dassanayake,A.K.K., De Silva,A.D., Kariyawasam,J.C., De Silva,A., Fernando,A.N., Tippalagama,R. and Rajendran,A. |
| EPI_ISL_17572824, EPI_ISL_17572825, EPI_ISL_17572826, EPI_ISL_17572827, EPI_ISL_17572828, EPI_ISL_17572829 | Unité de Mixte de Recherche, Processus Infectieux en Milieu Insulaire Tropical, Dynamique des Systèmes Infectieux Insulaires | Unité de Mixte de Recherche, Processus Infectieux en Milieu Insulaire Tropical, Dynamique des Systèmes Infectieux Insulaires | Hafsia,S., Barbar,T., Wilkinson,D.A., Atyame,C., Biscornet,L., Bibi,J., Louange,M., Gedeon,J., De Santis,O., Flahault,A., Cabie,A., Bertolotti,A. and Mavingui,P. |
| EPI_ISL_17595688, EPI_ISL_17595690, EPI_ISL_17595691, EPI_ISL_17595692 | Takeda Vaccines, Inc. | Takeda Vaccines, Inc. | Wallace,D. |
| EPI_ISL_17599631, EPI_ISL_17599632, EPI_ISL_17599633, EPI_ISL_17599634, EPI_ISL_17599635, EPI_ISL_17599636, EPI_ISL_17599637, EPI_ISL_17599638 | ICMR- National Institute for Research in Environmental Health | ICMR- National Institute for Research in Environmental Health | Sarma,D.K., Rathod,L., Mishra,S., Das,D. and Prakash,A. |
| EPI_ISL_17599669, EPI_ISL_17599670, EPI_ISL_17599671, EPI_ISL_17599672, EPI_ISL_17599673, EPI_ISL_17599674, EPI_ISL_17599675, EPI_ISL_17599676, EPI_ISL_17599677, EPI_ISL_17599678, EPI_ISL_17599679, EPI_ISL_17599680, EPI_ISL_17599681, EPI_ISL_17599682, EPI_ISL_17599683, EPI_ISL_17599684, EPI_ISL_17599685, EPI_ISL_17599686, EPI_ISL_17599687, EPI_ISL_17599688, EPI_ISL_17599689, EPI_ISL_17599690, EPI_ISL_17599691, EPI_ISL_17599692, EPI_ISL_17599693, EPI_ISL_17599694, EPI_ISL_17599695, EPI_ISL_17599696, EPI_ISL_17599697, EPI_ISL_17599698, EPI_ISL_17599699, EPI_ISL_17599700, EPI_ISL_17599701, EPI_ISL_17599702, EPI_ISL_17599703, EPI_ISL_17599704, EPI_ISL_17599705, EPI_ISL_17599706, EPI_ISL_17599707, EPI_ISL_17599708, EPI_ISL_17599709, EPI_ISL_17599710, EPI_ISL_17599711, EPI_ISL_17599712, EPI_ISL_17599713, EPI_ISL_17599714, EPI_ISL_17599715, EPI_ISL_17599716, EPI_ISL_17599717, EPI_ISL_17599718, EPI_ISL_17599719, EPI_ISL_17599720, EPI_ISL_17599721, EPI_ISL_17599722, EPI_ISL_17599723, EPI_ISL_17599724, EPI_ISL_17599725, EPI_ISL_17599726, EPI_ISL_17599727, EPI_ISL_17599728, EPI_ISL_17599729, EPI_ISL_17599730, EPI_ISL_17599731, EPI_ISL_17599732, EPI_ISL_17599752, EPI_ISL_17599790, EPI_ISL_17599828, EPI_ISL_17599864, EPI_ISL_17599904, EPI_ISL_17599944, EPI_ISL_17599984, EPI_ISL_17600022, EPI_ISL_17600061, EPI_ISL_17600101, EPI_ISL_17600105, EPI_ISL_17600106, EPI_ISL_17600107, EPI_ISL_17600108, EPI_ISL_17600109, EPI_ISL_17600110, EPI_ISL_17600111, EPI_ISL_17600112, EPI_ISL_17600113, EPI_ISL_17600114, EPI_ISL_17600115, EPI_ISL_17600116, EPI_ISL_17600117, EPI_ISL_17600118, EPI_ISL_17600119, EPI_ISL_17600120, EPI_ISL_17600121, EPI_ISL_17600122, EPI_ISL_17600123, EPI_ISL_17600124, EPI_ISL_17600125, EPI_ISL_17600126, EPI_ISL_17600127, EPI_ISL_17600128, EPI_ISL_17600129, EPI_ISL_17600130, EPI_ISL_17600131, EPI_ISL_17600132, EPI_ISL_17600133, EPI_ISL_17600134, EPI_ISL_17600135, EPI_ISL_17600136, EPI_ISL_17600137, EPI_ISL_17600138, EPI_ISL_17600139, EPI_ISL_17600140, EPI_ISL_17600141, EPI_ISL_17600142, EPI_ISL_17600143, EPI_ISL_17600144, EPI_ISL_17600145, EPI_ISL_17600146, EPI_ISL_17600147, EPI_ISL_17600148, EPI_ISL_17600149, EPI_ISL_17600150, EPI_ISL_17600151, EPI_ISL_17600152, EPI_ISL_17600153, EPI_ISL_17600154, EPI_ISL_17600155, EPI_ISL_17600156, EPI_ISL_17600157, EPI_ISL_17600158, EPI_ISL_17600159, EPI_ISL_17600160, EPI_ISL_17600161, EPI_ISL_17600162, EPI_ISL_17600163, EPI_ISL_17600164, EPI_ISL_17600165, EPI_ISL_17600166, EPI_ISL_17600167, EPI_ISL_17600168, EPI_ISL_17600169, EPI_ISL_17600170, EPI_ISL_17600171, EPI_ISL_17600172, EPI_ISL_17600173, EPI_ISL_17600174, EPI_ISL_17600175, EPI_ISL_17600176, EPI_ISL_17600177, EPI_ISL_17600178, EPI_ISL_17600179, EPI_ISL_17600180, EPI_ISL_17600181, EPI_ISL_17600182, EPI_ISL_17600183, EPI_ISL_17600184, EPI_ISL_17600185, EPI_ISL_17600186, EPI_ISL_17600187, EPI_ISL_17600188, EPI_ISL_17600189, EPI_ISL_17600190, EPI_ISL_17600191, EPI_ISL_17600192, EPI_ISL_17600193, EPI_ISL_17600194, EPI_ISL_17600195, EPI_ISL_17600196, EPI_ISL_17600197, EPI_ISL_17600198, EPI_ISL_17600199, EPI_ISL_17600200, EPI_ISL_17600201, EPI_ISL_17600202, EPI_ISL_17600203, EPI_ISL_17600204, EPI_ISL_17600205, EPI_ISL_17600206, EPI_ISL_17600207, EPI_ISL_17600208, EPI_ISL_17600209, EPI_ISL_17600210, EPI_ISL_17600211, EPI_ISL_17600212, EPI_ISL_17600213, EPI_ISL_17600214, EPI_ISL_17600215, EPI_ISL_17600216, EPI_ISL_17600217, EPI_ISL_17600218, EPI_ISL_17600219, EPI_ISL_17600220, EPI_ISL_17600221, EPI_ISL_17600222, EPI_ISL_17600223, EPI_ISL_17600224, EPI_ISL_17600225, EPI_ISL_17600226, EPI_ISL_17600227, EPI_ISL_17600228, EPI_ISL_17600229, EPI_ISL_17600230, EPI_ISL_17600231, EPI_ISL_17600232, EPI_ISL_17600233, EPI_ISL_17600234, EPI_ISL_17600235, EPI_ISL_17600236, EPI_ISL_17600237, EPI_ISL_17600238, EPI_ISL_17600239, EPI_ISL_17600240, EPI_ISL_17600241, EPI_ISL_17600242, EPI_ISL_17600243, EPI_ISL_17600244, EPI_ISL_17600245, EPI_ISL_17600246, EPI_ISL_17600247, EPI_ISL_17600248, EPI_ISL_17600249, EPI_ISL_17600250, EPI_ISL_17600251, EPI_ISL_17600252, EPI_ISL_17600253, EPI_ISL_17600254, EPI_ISL_17600255, EPI_ISL_17600256, EPI_ISL_17600257, EPI_ISL_17600258, EPI_ISL_17600259, EPI_ISL_17600260, EPI_ISL_17600261, EPI_ISL_17600262, EPI_ISL_17600263, EPI_ISL_17600264, EPI_ISL_17600265, EPI_ISL_17600266, EPI_ISL_17600267, EPI_ISL_17600268, EPI_ISL_17600269, EPI_ISL_17600270, EPI_ISL_17600271, EPI_ISL_17600272, EPI_ISL_17600273, EPI_ISL_17600274, EPI_ISL_17600275, EPI_ISL_17600276, EPI_ISL_17600277, EPI_ISL_17600278, EPI_ISL_17600279, EPI_ISL_17600280, EPI_ISL_17600281, EPI_ISL_17600282, EPI_ISL_17600283, EPI_ISL_17600284, EPI_ISL_17600285, EPI_ISL_17600286, EPI_ISL_17600287, EPI_ISL_17600288, EPI_ISL_17600289, EPI_ISL_17600290, EPI_ISL_17600291, EPI_ISL_17600292, EPI_ISL_17600293, EPI_ISL_17600294, EPI_ISL_17600295, EPI_ISL_17600296, EPI_ISL_17600297, EPI_ISL_17600298, EPI_ISL_17600299, EPI_ISL_17600300, EPI_ISL_17600301, EPI_ISL_17600302, EPI_ISL_17600303, EPI_ISL_17600304, EPI_ISL_17600305, EPI_ISL_17600306, EPI_ISL_17600307, EPI_ISL_17600308, EPI_ISL_17600309, EPI_ISL_17600310, EPI_ISL_17600311, EPI_ISL_17600312, EPI_ISL_17600313, EPI_ISL_17600314, EPI_ISL_17600315, EPI_ISL_17600316, EPI_ISL_17600317, EPI_ISL_17600318, EPI_ISL_17600319, EPI_ISL_17600320, EPI_ISL_17600321, EPI_ISL_17600322, EPI_ISL_17600323, EPI_ISL_17600324, EPI_ISL_17600325, EPI_ISL_17600326, EPI_ISL_17600327, EPI_ISL_17600328, EPI_ISL_17600329, EPI_ISL_17600330, EPI_ISL_17600331, EPI_ISL_17600332, EPI_ISL_17600333, EPI_ISL_17600334, EPI_ISL_17600335, EPI_ISL_17600336, EPI_ISL_17600337, EPI_ISL_17600338, EPI_ISL_17600339, EPI_ISL_17600340, EPI_ISL_17600341, EPI_ISL_17600342, EPI_ISL_17600343, EPI_ISL_17600344, EPI_ISL_17600345, EPI_ISL_17600346, EPI_ISL_17600347, EPI_ISL_17600348, EPI_ISL_17600349, EPI_ISL_17600350, EPI_ISL_17600351, EPI_ISL_17600352, EPI_ISL_17600353, EPI_ISL_17600354, EPI_ISL_17600355, EPI_ISL_17600356, EPI_ISL_17600357, EPI_ISL_17600358, EPI_ISL_17600360, EPI_ISL_17600361, EPI_ISL_17600362, EPI_ISL_17600363, EPI_ISL_17600364, EPI_ISL_17600365, EPI_ISL_17600366, EPI_ISL_17600367, EPI_ISL_17600368, EPI_ISL_17600369, EPI_ISL_17600370, EPI_ISL_17600371, EPI_ISL_17600372, EPI_ISL_17600373, EPI_ISL_17600374, EPI_ISL_17600375, EPI_ISL_17600376, EPI_ISL_17600377, EPI_ISL_17600378, EPI_ISL_17600379, EPI_ISL_17600380, EPI_ISL_17600381, EPI_ISL_17600382, EPI_ISL_17600383, EPI_ISL_17600384, EPI_ISL_17600385, EPI_ISL_17600386, EPI_ISL_17600387, EPI_ISL_17600388, EPI_ISL_17600389, EPI_ISL_17600390, EPI_ISL_17600391, EPI_ISL_17600392, EPI_ISL_17600393, EPI_ISL_17600394, EPI_ISL_17600395, EPI_ISL_17600396, EPI_ISL_17600397, EPI_ISL_17600398, EPI_ISL_17600399, EPI_ISL_17600400, EPI_ISL_17600401, EPI_ISL_17600402 | | | |
| see above | National Health Laboratory | National Health Laboratory | Kelly,M.E., Msafiri,F., Affara,M., Gehre,F., Moremi,N., Mghamba,J., Misinzo,G., Gatei,W., Whistler,T., Joachim,A., Lema,N. and Santiago,G.A. |
| EPI_ISL_17600403 | Universidad de Cordoba | Universidad de Cordoba | Rivero,R., Paternina,D., Tique,V. and Mattar,S. |
| EPI_ISL_17600404 | Centers for Disease Control and Prevention, Dengue Branch | Centers for Disease Control and Prevention, Dengue Branch | Santiago,G.A., Gonzalez-Morales,G.L., Charriez,K.N., Flores,B., Chow,A., Newsum,M., Mendoza,S., Poujilvaia,K., Fitzpatrick,K. and Munoz-Jordan,J.L. |
| EPI_ISL_17600405 | Centers for Disease Control and Prevention, | Centers for Disease Control and Prevention, Dengue | Santiago,G.A., Gonzalez-Morales,G., Flores,B., Charriez,K.N., Falcon,J. and Munoz-Jordan,J.L. |

| | | | |
|---|---|---|---|
| | Dengue Branch | Branch | |
| EPI_ISL_17600406 | Centers for Disease Control and Prevention, Dengue Branch | Centers for Disease Control and Prevention, Dengue Branch | Santiago,G.A., Gonzalez-Morales,G.L., Flores,B., Charriez,K.N., Medina,J., Falcon,J., Brown,G. and Munoz-Jordan,J.L. |
| EPI_ISL_17600407, EPI_ISL_17600408 | Centers for Disease Control and Prevention, Dengue Branch | Centers for Disease Control and Prevention, Dengue Branch | Santiago,G.A., Gonzalez-Morales,G.L., Charriez,K.N., Flores,B., Chow,A., Newsum,M., Mendoza,S., Poujilvaia,K., Fitzpatrick,K. and Munoz-Jordan,J.L. |
| EPI_ISL_17600409, EPI_ISL_17600410, EPI_ISL_17600411, EPI_ISL_17600412, EPI_ISL_17600413, EPI_ISL_17600414 | Centers for Disease Control and Prevention, Dengue Branch | Centers for Disease Control and Prevention, Dengue Branch | Santiago,G.A., Gonzalez-Morales,G.L., Flores,B., Charriez,K.N., Medina,J., Falcon,J., Brown,G. and Munoz-Jordan,J.L. |
| EPI_ISL_17600462, EPI_ISL_17600463 | Department of Epidemiology of Microbial Diseases, Yale School of Public Health | Department of Epidemiology of Microbial Diseases, Yale School of Public Health | Vogels,C.B.F., Breban,M.I., Chaguza,C., Hill,V., Paul,L., Michael,S., Bunch,S., Cano,N., Jaber,R., Morrison,A., Panzera,C., Stryker,I., Vergara,J., Zimler,R., Kopp,E., Herberlein,L. and Grubaugh,N.D. |
| EPI_ISL_17600464, EPI_ISL_17600465 | Department of Epidemiology of Microbial Diseases, Yale School of Public Health | Department of Epidemiology of Microbial Diseases, Yale School of Public Health | Vogels,C.B.F., Breban,M.I., Chaguza,C., Hill,V., Ovalle Segura,J.A., Campechano Perez,C., Jabier,M., Mueses,S., Peguero,A., Cuevas,P., Sancehz Morfe,N., Ruiz,I., Paulino-Ramirez,R. and Grubaugh,N.D. |
| EPI_ISL_17600466 | Department of Epidemiology of Microbial Diseases, Yale School of Public Health | Department of Epidemiology of Microbial Diseases, Yale School of Public Health | Vogels,C.B.F., Breban,M.I., Chaguza,C., Hill,V., Paul,L., Michael,S., Bunch,S., Cano,N., Jaber,R., Morrison,A., Panzera,C., Stryker,I., Vergara,J., Zimler,R., Kopp,E., Herberlein,L. and Grubaugh,N.D. |
| EPI_ISL_17600467 | Department of Epidemiology of Microbial Diseases, Yale School of Public Health | Department of Epidemiology of Microbial Diseases, Yale School of Public Health | Vogels,C.B.F., Breban,M.I., Chaguza,C., Hill,V., Ovalle Segura,J.A., Campechano Perez,C., Jabier,M., Mueses,S., Peguero,A., Cuevas,P., Sancehz Morfe,N., Ruiz,I., Paulino-Ramirez,R. and Grubaugh,N.D. |
| EPI_ISL_17600468, EPI_ISL_17600469, EPI_ISL_17600470, EPI_ISL_17600471, EPI_ISL_17600472, EPI_ISL_17600473, EPI_ISL_17600474, EPI_ISL_17600475, EPI_ISL_17600476, EPI_ISL_17600477, EPI_ISL_17600478, EPI_ISL_17600479, EPI_ISL_17600480, EPI_ISL_17600481 | | | |
| see above | Department of Epidemiology of Microbial Diseases, Yale School of Public Health | Department of Epidemiology of Microbial Diseases, Yale School of Public Health | Vogels,C.B.F., Breban,M.I., Chaguza,C., Hill,V., Paul,L., Michael,S., Bunch,S., Cano,N., Jaber,R., Morrison,A., Panzera,C., Stryker,I., Vergara,J., Zimler,R., Kopp,E., Herberlein,L. and Grubaugh,N.D. |
| EPI_ISL_17600482, EPI_ISL_17600483, EPI_ISL_17600484, EPI_ISL_17600485, EPI_ISL_17600486 | Department of Epidemiology of Microbial Diseases, Yale School of Public Health | Department of Epidemiology of Microbial Diseases, Yale School of Public Health | Vogels,C.B.F., Breban,M.I., Chaguza,C., Hill,V., Ovalle Segura,J.A., Campechano Perez,C., Jabier,M., Mueses,S., Peguero,A., Cuevas,P., Sancehz Morfe,N., Ruiz,I., Paulino-Ramirez,R. and Grubaugh,N.D. |
| EPI_ISL_17600487, EPI_ISL_17600488, EPI_ISL_17600489, EPI_ISL_17600490, EPI_ISL_17600491, EPI_ISL_17600492, EPI_ISL_17600493, EPI_ISL_17600494, EPI_ISL_17600495, EPI_ISL_17600496, EPI_ISL_17600497, EPI_ISL_17600498, EPI_ISL_17600499, EPI_ISL_17600500, EPI_ISL_17600501, EPI_ISL_17600502, EPI_ISL_17600503, EPI_ISL_17600504, EPI_ISL_17600505, EPI_ISL_17600506, EPI_ISL_17600507, EPI_ISL_17600508, EPI_ISL_17600509, EPI_ISL_17600510, EPI_ISL_17600511, EPI_ISL_17600512, EPI_ISL_17600513, EPI_ISL_17600514, EPI_ISL_17600515, EPI_ISL_17600516, EPI_ISL_17600517, EPI_ISL_17600518, EPI_ISL_17600519, EPI_ISL_17600520, EPI_ISL_17600521, EPI_ISL_17600522, EPI_ISL_17600523, EPI_ISL_17600524, EPI_ISL_17600525, EPI_ISL_17600526, EPI_ISL_17600527 | | | |
| see above | Department of Epidemiology of Microbial Diseases, Yale School of Public Health | Department of Epidemiology of Microbial Diseases, Yale School of Public Health | Vogels,C.B.F., Breban,M.I., Chaguza,C., Hill,V., Paul,L., Michael,S., Bunch,S., Cano,N., Jaber,R., Morrison,A., Panzera,C., Stryker,I., Vergara,J., Zimler,R., Kopp,E., Herberlein,L. and Grubaugh,N.D. |
| EPI_ISL_17600528, EPI_ISL_17600529 | Department of Epidemiology of Microbial Diseases, Yale School of Public Health | Department of Epidemiology of Microbial Diseases, Yale School of Public Health | Vogels,C.B.F., Breban,M.I., Chaguza,C., Hill,V., Ovalle Segura,J.A., Campechano Perez,C., Jabier,M., Mueses,S., Peguero,A., Cuevas,P., Sancehz Morfe,N., Ruiz,I., Paulino-Ramirez,R. and Grubaugh,N.D. |
| EPI_ISL_17600530, EPI_ISL_17600531 | Department of Epidemiology of Microbial Diseases, Yale School of Public Health | Department of Epidemiology of Microbial Diseases, Yale School of Public Health | Vogels,C.B.F., Breban,M.I., Chaguza,C., Hill,V., Norman,F., Galan,J.-C. and Grubaugh,N.D. |
| EPI_ISL_17600532, EPI_ISL_17600533 | Department of Epidemiology of Microbial Diseases, Yale School of Public Health | Department of Epidemiology of Microbial Diseases, Yale School of Public Health | Vogels,C.B.F., Breban,M.I., Chaguza,C., Hill,V., Paul,L., Michael,S., Bunch,S., Cano,N., Jaber,R., Morrison,A., Panzera,C., Stryker,I., Vergara,J., Zimler,R., Kopp,E., Herberlein,L. and Grubaugh,N.D. |
| EPI_ISL_17600534 | Department of Epidemiology of Microbial Diseases, Yale School of Public Health | Department of Epidemiology of Microbial Diseases, Yale School of Public Health | Vogels,C.B.F., Breban,M.I., Chaguza,C., Hill,V., Ovalle Segura,J.A., Campechano Perez,C., Jabier,M., Mueses,S., Peguero,A., Cuevas,P., Sancehz Morfe,N., Ruiz,I., Paulino-Ramirez,R. and Grubaugh,N.D. |
| EPI_ISL_17600535, EPI_ISL_17600536, EPI_ISL_17600537, EPI_ISL_17600538, EPI_ISL_17600539, EPI_ISL_17600540, EPI_ISL_17600541, EPI_ISL_17600542, EPI_ISL_17600543, EPI_ISL_17600544, EPI_ISL_17600545, EPI_ISL_17600546, EPI_ISL_17600547, EPI_ISL_17600548, EPI_ISL_17600549, EPI_ISL_17600550, EPI_ISL_17600551, EPI_ISL_17600552, EPI_ISL_17600553, EPI_ISL_17600554, EPI_ISL_17600555, EPI_ISL_17600556, EPI_ISL_17600557, EPI_ISL_17600558, EPI_ISL_17600559, EPI_ISL_17600560, EPI_ISL_17600561, EPI_ISL_17600562, EPI_ISL_17600563, EPI_ISL_17600564, EPI_ISL_17600565, EPI_ISL_17600566, EPI_ISL_17600567, EPI_ISL_17600568, EPI_ISL_17600569, EPI_ISL_17600570, EPI_ISL_17600571, EPI_ISL_17600572, EPI_ISL_17600573, EPI_ISL_17600574, EPI_ISL_17600575, EPI_ISL_17600576, EPI_ISL_17600577, EPI_ISL_17600578, EPI_ISL_17600579, EPI_ISL_17600580, EPI_ISL_17600581, EPI_ISL_17600582, EPI_ISL_17600583, EPI_ISL_17600584, EPI_ISL_17600585, EPI_ISL_17600586, EPI_ISL_17600587, EPI_ISL_17600588, EPI_ISL_17600589, EPI_ISL_17600590, EPI_ISL_17600591, EPI_ISL_17600592, EPI_ISL_17600593, EPI_ISL_17600594, EPI_ISL_17600595, EPI_ISL_17600596, EPI_ISL_17600597, EPI_ISL_17600598, EPI_ISL_17600599 | | | |
| see above | Department of Epidemiology of Microbial Diseases, Yale School of Public Health | Department of Epidemiology of Microbial Diseases, Yale School of Public Health | Vogels,C.B.F., Breban,M.I., Chaguza,C., Hill,V., Paul,L., Michael,S., Bunch,S., Cano,N., Jaber,R., Morrison,A., Panzera,C., Stryker,I., Vergara,J., Zimler,R., Kopp,E., Herberlein,L. and Grubaugh,N.D. |
| EPI_ISL_17600600 | Department of Epidemiology of Microbial Diseases, Yale School of Public Health | Department of Epidemiology of Microbial Diseases, Yale School of Public Health | Vogels,C.B.F., Breban,M.I., Chaguza,C., Hill,V., Ovalle Segura,J.A., Campechano Perez,C., Jabier,M., Mueses,S., Peguero,A., Cuevas,P., Sancehz Morfe,N., Ruiz,I., Paulino-Ramirez,R. and Grubaugh,N.D. |
| EPI_ISL_17600601, EPI_ISL_17600602, EPI_ISL_17600603, EPI_ISL_17600604, EPI_ISL_17600605, EPI_ISL_17600606, EPI_ISL_17600607, EPI_ISL_17600608, EPI_ISL_17600609, EPI_ISL_17600610, EPI_ISL_17600611, EPI_ISL_17600612, EPI_ISL_17600613, EPI_ISL_17600614, EPI_ISL_17600615, EPI_ISL_17600616, EPI_ISL_17600617, EPI_ISL_17600618, EPI_ISL_17600619, EPI_ISL_17600620, EPI_ISL_17600621, EPI_ISL_17600622, EPI_ISL_17600623, EPI_ISL_17600624, EPI_ISL_17600625, EPI_ISL_17600626, EPI_ISL_17600627, EPI_ISL_17600628, EPI_ISL_17600629, EPI_ISL_17600630, EPI_ISL_17600631, EPI_ISL_17600632, EPI_ISL_17600633, EPI_ISL_17600634, EPI_ISL_17600635, EPI_ISL_17600636, EPI_ISL_17600637, EPI_ISL_17600638, EPI_ISL_17600639, EPI_ISL_17600640, EPI_ISL_17600641, EPI_ISL_17600642, EPI_ISL_17600643, EPI_ISL_17600644, EPI_ISL_17600645, EPI_ISL_17600646, EPI_ISL_17600647, EPI_ISL_17600648, EPI_ISL_17600649, EPI_ISL_17600650, EPI_ISL_17600651, EPI_ISL_17600652, EPI_ISL_17600653, EPI_ISL_17600654, EPI_ISL_17600655, EPI_ISL_17600656, EPI_ISL_17600657, EPI_ISL_17600658, EPI_ISL_17600659, EPI_ISL_17600660, EPI_ISL_17600661, EPI_ISL_17600662, EPI_ISL_17600663, EPI_ISL_17600664, EPI_ISL_17600665, EPI_ISL_17600666, EPI_ISL_17600667, EPI_ISL_17600668, EPI_ISL_17600669, EPI_ISL_17600670, EPI_ISL_17600671, EPI_ISL_17600672, EPI_ISL_17600673, EPI_ISL_17600674, EPI_ISL_17600675, EPI_ISL_17600676, EPI_ISL_17600677, EPI_ISL_17600678, EPI_ISL_17600679, EPI_ISL_17600680, EPI_ISL_17600681, EPI_ISL_17600682, EPI_ISL_17600683, EPI_ISL_17600684, EPI_ISL_17600685, EPI_ISL_17600686, EPI_ISL_17600687, EPI_ISL_17600688, EPI_ISL_17600689, EPI_ISL_17600690, EPI_ISL_17600691, EPI_ISL_17600692, EPI_ISL_17600693, EPI_ISL_17600694, EPI_ISL_17600695, EPI_ISL_17600696, EPI_ISL_17600697, EPI_ISL_17600698, EPI_ISL_17600699, EPI_ISL_17600700, EPI_ISL_17600701, EPI_ISL_17600702, EPI_ISL_17600703, EPI_ISL_17600704, EPI_ISL_17600705, EPI_ISL_17600706, EPI_ISL_17600707, EPI_ISL_17600708, EPI_ISL_17600709, EPI_ISL_17600710, EPI_ISL_17600711, EPI_ISL_17600712, EPI_ISL_17600713, EPI_ISL_17600714, EPI_ISL_17600715, EPI_ISL_17600716, EPI_ISL_17600717, EPI_ISL_17600718, EPI_ISL_17600719, EPI_ISL_17600720, EPI_ISL_17600721, EPI_ISL_17600722, EPI_ISL_17600723, EPI_ISL_17600724, EPI_ISL_17600725, EPI_ISL_17600726, EPI_ISL_17600727, EPI_ISL_17600728, EPI_ISL_17600729, EPI_ISL_17600730, EPI_ISL_17600731, EPI_ISL_17600732, EPI_ISL_17600733, EPI_ISL_17600734, EPI_ISL_17600735, EPI_ISL_17600736, EPI_ISL_17600737, EPI_ISL_17600738, EPI_ISL_17600739, EPI_ISL_17600740, EPI_ISL_17600741, EPI_ISL_17600742, EPI_ISL_17600743, EPI_ISL_17600744, EPI_ISL_17600745, EPI_ISL_17600746, EPI_ISL_17600747, EPI_ISL_17600748, EPI_ISL_17600749, EPI_ISL_17600750, EPI_ISL_17600751, EPI_ISL_17600752, EPI_ISL_17600753, EPI_ISL_17600754, EPI_ISL_17600755, EPI_ISL_17600756, EPI_ISL_17600757, EPI_ISL_17600758, EPI_ISL_17600759, EPI_ISL_17600760, EPI_ISL_17600761, EPI_ISL_17600762, EPI_ISL_17600763, EPI_ISL_17600764, EPI_ISL_17600765, EPI_ISL_17600766, EPI_ISL_17600767, EPI_ISL_17600768, EPI_ISL_17600769, EPI_ISL_17600770, EPI_ISL_17600771, EPI_ISL_17600772, EPI_ISL_17600773, EPI_ISL_17600774, EPI_ISL_17600775, EPI_ISL_17600776, EPI_ISL_17600777, EPI_ISL_17600778, EPI_ISL_17600779, EPI_ISL_17600780, EPI_ISL_17600781, EPI_ISL_17600782, EPI_ISL_17600783, EPI_ISL_17600784, EPI_ISL_17600785, EPI_ISL_17600786, EPI_ISL_17600787, EPI_ISL_17600788, EPI_ISL_17600789, EPI_ISL_17600790, EPI_ISL_17600791, EPI_ISL_17600792, EPI_ISL_17600793, EPI_ISL_17600794, EPI_ISL_17600795, EPI_ISL_17600796, EPI_ISL_17600797, EPI_ISL_17600798, EPI_ISL_17600799, EPI_ISL_17600800, EPI_ISL_17600801 | | | |
| see above | Department of Epidemiology of Microbial Diseases, Yale School of Public Health | Department of Epidemiology of Microbial Diseases, Yale School of Public Health | Vogels,C.B.F., Breban,M.I., Chaguza,C., Hill,V., Paul,L., Michael,S., Bunch,S., Cano,N., Jaber,R., Morrison,A., Panzera,C., Stryker,I., Vergara,J., Zimler,R., Kopp,E., Herberlein,L. and Grubaugh,N.D. |
| EPI_ISL_17600802, EPI_ISL_17600803, EPI_ISL_17600804 | Department of Microbiology, King George's Medical University | Department of Microbiology, King George's Medical University | Mallikarjun,K., Santhosh,K.S. and Puneeth,T.G. |
| EPI_ISL_17600805, EPI_ISL_17600806, EPI_ISL_17600807, EPI_ISL_17600808, EPI_ISL_17600809, EPI_ISL_17600810, EPI_ISL_17600811, EPI_ISL_17600812, EPI_ISL_17600813, EPI_ISL_17600814, EPI_ISL_17600815, EPI_ISL_17600816, EPI_ISL_17600817, EPI_ISL_17600818, EPI_ISL_17600819, EPI_ISL_17600820, EPI_ISL_17600821, EPI_ISL_17600822 | | | |
| see above | Eijkman Institute for Molecular Biology, Dengue Unit | Eijkman Institute for Molecular Biology, Dengue Unit | Sasmono,R.T., Nara,M.B., Santoso,M.S., Nugroho,D.K., Yohan,B., Purnama,A., Boro,A.M., Hayati,R.F., Gae,E.P., Denis,D. and Rana,B. |
| EPI_ISL_17600823 | Infection and Immunology, Translational Health Science and Technology Institute | Infection and Immunology, Translational Health Science and Technology Institute | Verma,C.K., Singh,B., Shashi,P., Chandele,A., Lodha,R. and Medigeshi,G.R. |
| EPI_ISL_17600824 | Infection and Immunology, Translational Health Science and Technology Institute | Infection and Immunology, Translational Health Science and Technology Institute | Singh,B., Shashi,P., Verma,C.K., Chandele,A., Lodha,R. and Medigeshi,G.R. |
| EPI_ISL_17600825, EPI_ISL_17600826 | Infection and Immunology, Translational Health Science and Technology Institute | Infection and Immunology, Translational Health Science and Technology Institute | Verma,C.K., Singh,B., Shashi,P., Chandele,A., Lodha,R. and Medigeshi,G.R. |
| EPI_ISL_17600827, EPI_ISL_17600828, EPI_ISL_17600829, EPI_ISL_17600830, EPI_ISL_17600831, EPI_ISL_17600832, EPI_ISL_17600833, EPI_ISL_17600834 | Infection and Immunology, Translational Health Science and Technology Institute | Infection and Immunology, Translational Health Science and Technology Institute | Singh,B., Shashi,P., Verma,C.K., Chandele,A., Lodha,R. and Medigeshi,G.R. |
| EPI_ISL_17600835, EPI_ISL_17600836, EPI_ISL_17600837, EPI_ISL_17600838, EPI_ISL_17600839, EPI_ISL_17600840, EPI_ISL_17600841 | Institute of Medical Biology, Chinese Academy of Medical Sciences, Peking Union Medical College, Tsinghua University | Institute of Medical Biology, Chinese Academy of Medical Sciences, Peking Union Medical College, Tsinghua University | Zhou,Y., Zhou,Y., Liyang,S., Wang,P., An,J. and Liu,H. |
| EPI_ISL_17600842, EPI_ISL_17600843, EPI_ISL_17600844, EPI_ISL_17600845, EPI_ISL_17600846, EPI_ISL_17600847, EPI_ISL_17600848, EPI_ISL_17600849, EPI_ISL_17600850, EPI_ISL_17600851, EPI_ISL_17600852, EPI_ISL_17600853, EPI_ISL_17600854 | | | |
| see above | Institute of Virology, Wenzhou University | Institute of Virology, Wenzhou University | Liu,Y. |
| EPI_ISL_17600855 | Instituto Leonidas e Maria Deane, Fiocruz | Instituto Leonidas e Maria Deane, Fiocruz | Regina Maria Pinto de Figueiredo, Victor Costa de Souza, Luciana Goncalves, Dejanane Silva, Fernanda Oliveira do Nascimento, Matilde Mejia, Valdinete Alves do Nascimento and Felipe Naveca |
| EPI_ISL_17600856, EPI_ISL_17600857, EPI_ISL_17600858 | Laboratório Central de Saúde Pública de Roraima | Instituto Leonidas e Maria Deane, Fiocruz | Victor Costa de Souza, Goncalves,L., Silva,D., Fernanda Oliveira do Nascimento, Mejia,M., Nascimento,V. and Naveca,F. |
| EPI_ISL_17600859, EPI_ISL_17600860, EPI_ISL_17600861, EPI_ISL_17600862 | Laboratory of Malaria and Vector Research, National Institute of Allergy and Infectious Diseases | Laboratory of Malaria and Vector Research, National Institute of Allergy and Infectious Diseases | Bohl,J.A., Lay,S., Chea,S., Ahyong,V., Parker,D.M., Gallagher,S., Fintzi,J., Man,S., Ponce,A., Sreng,S., Kong,D., Oliveira,F., Kalantar,K., Tan,M., Fahsbender,L., Sheu,J., Neff,N., Detweiler,A.M., Yek,C., Ly,S., Sath,R., Huch,C., Kry,H., Leang,R., Huy,R., Lon,C., Tato,C.M., DeRisi,J.L. and Manning,J.E. |
| EPI_ISL_17600863 | Laboratory of Malaria and Vector Research, National Institute of Allergy and Infectious Diseases | Laboratory of Malaria and Vector Research, National Institute of Allergy and Infectious Diseases | Oum,M., Lay,S., Chea,S., Bohl,J.A., Yek,C. and Manning,J.E. |
| EPI_ISL_17600864 | Laboratory of Malaria and Vector Research, National Institute of Allergy and Infectious Diseases | Laboratory of Malaria and Vector Research, National Institute of Allergy and Infectious Diseases | Bohl,J.A., Lay,S., Chea,S., Ahyong,V., Parker,D.M., Gallagher,S., Fintzi,J., Man,S., Ponce,A., Sreng,S., Kong,D., Oliveira,F., Kalantar,K., Tan,M., Fahsbender,L., Sheu,J., Neff,N., Detweiler,A.M., Yek,C., Ly,S., Sath,R., Huch,C., Kry,H., Leang,R., Huy,R., Lon,C., Tato,C.M., DeRisi,J.L. and Manning,J.E. |
| EPI_ISL_17600865 | Laboratory of Malaria and Vector Research, National Institute of Allergy and Infectious Diseases | Laboratory of Malaria and Vector Research, National Institute of Allergy and Infectious Diseases | Oum,M., Lay,S., Chea,S., Bohl,J.A., Yek,C. and Manning,J.E. |
| EPI_ISL_17600866, EPI_ISL_17600867, EPI_ISL_17600868, EPI_ISL_17600869, EPI_ISL_17600870 | Laboratory of Malaria and Vector Research, National Institute of Allergy and Infectious Diseases | Laboratory of Malaria and Vector Research, National Institute of Allergy and Infectious Diseases | Bohl,J.A., Lay,S., Chea,S., Ahyong,V., Parker,D.M., Gallagher,S., Fintzi,J., Man,S., Ponce,A., Sreng,S., Kong,D., Oliveira,F., Kalantar,K., Tan,M., Fahsbender,L., Sheu,J., Neff,N., Detweiler,A.M., Yek,C., Ly,S., Sath,R., Huch,C., Kry,H., Leang,R., Huy,R., Lon,C., Tato,C.M., DeRisi,J.L. and Manning,J.E. |
| EPI_ISL_17600871 | Laboratory of Malaria and Vector Research, National Institute of Allergy and Infectious | Laboratory of Malaria and Vector Research, National Institute of Allergy and Infectious Diseases | Oum,M., Lay,S., Chea,S., Bohl,J.A., Yek,C. and Manning,J.E. |

| | | | |
|---|---|---|---|
| | Diseases | | |
| EPI_ISL_17600872 | Laboratory of Malaria and Vector Research, National Institute of Allergy and Infectious Diseases | Laboratory of Malaria and Vector Research, National Institute of Allergy and Infectious Diseases | Bohl,J.A., Lay,S., Chea,S., Ahyong,V., Parker,D.M., Gallagher,S., Fintzi,J., Man,S., Ponce,A., Sreng,S., Kong,D., Oliveira,F., Kalantar,K., Tan,M., Fahsbender,L., Sheu,J., Neff,N., Detweiler,A.M., Yek,C., Ly,S., Sath,R., Huch,C., Kry,H., Leang,R., Huy,R., Lon,C., Tato,C.M., DeRisi,J.L. and Manning,J.E. |
| EPI_ISL_17600873 | Laboratory of Malaria and Vector Research, National Institute of Allergy and Infectious Diseases | Laboratory of Malaria and Vector Research, National Institute of Allergy and Infectious Diseases | Oum,M., Lay,S., Chea,S., Bohl,J.A., Yek,C. and Manning,J.E. |
| EPI_ISL_17600874 | Laboratory of Malaria and Vector Research, National Institute of Allergy and Infectious Diseases | Laboratory of Malaria and Vector Research, National Institute of Allergy and Infectious Diseases | Bohl,J.A., Lay,S., Chea,S., Ahyong,V., Parker,D.M., Gallagher,S., Fintzi,J., Man,S., Ponce,A., Sreng,S., Kong,D., Oliveira,F., Kalantar,K., Tan,M., Fahsbender,L., Sheu,J., Neff,N., Detweiler,A.M., Yek,C., Ly,S., Sath,R., Huch,C., Kry,H., Leang,R., Huy,R., Lon,C., Tato,C.M., DeRisi,J.L. and Manning,J.E. |
| EPI_ISL_17600888, EPI_ISL_17600889, EPI_ISL_17600890, EPI_ISL_17600891, EPI_ISL_17600892, EPI_ISL_17600893, EPI_ISL_17600894, EPI_ISL_17600895, EPI_ISL_17600896 | Virology, Oswaldo Cruz Foundation | Virology, Oswaldo Cruz Foundation | Mendonca,M.C.L., Cavalcanti,A.C., Camargo,A.C., Rodrigues,C.D., Fabri,A.A., Santos,C.C. and Filippis,A.M. |
| EPI_ISL_17600897, EPI_ISL_17600898, EPI_ISL_17600899, EPI_ISL_17600900, EPI_ISL_17600901, EPI_ISL_17600902, EPI_ISL_17600903, EPI_ISL_17600904, EPI_ISL_17600905 | Virus Laboratory, Indian Council of Medical Research, National Institute of Cholera and Enteric Diseases | Virus Laboratory, Indian Council of Medical Research, National Institute of Cholera and Enteric Diseases | Baskey,U., Verma,P., Ghosh,A. and Sadhukhan,P.C. |
| EPI_ISL_17609476 | Center for Disease Control and Prevention of Henan Province | Center for Disease Control and Prevention of Henan Province | Li,D.X. and Li,Y. |
| EPI_ISL_17609477 | Centers for Disease Control and Prevention, Arboviral Diseases Branch | Centers for Disease Control and Prevention, Arboviral Diseases Branch | Hughes,H.R. and Russell,B.J. |
| EPI_ISL_17609844 | Laboratório Central do Estado do Paraná | Instituto Leonidas e Maria Deane, Fiocruz | Presibella,M., Marques,N.F., Riediger,I.N., Victor Costa de Souza, Goncalves,L., Silva,D., Fernanda Oliveira do Nascimento, Mejia,M., Nascimento,V. and Naveca,F. |
| EPI_ISL_17609855, EPI_ISL_17609856 | King George's Medical University, Department of Microbiology | King George's Medical University, Department of Microbiology | Prakash,O. |
| EPI_ISL_17609857, EPI_ISL_17609858, EPI_ISL_17609859, EPI_ISL_17609860, EPI_ISL_17609861, EPI_ISL_17609862, EPI_ISL_17609863, EPI_ISL_17609864, EPI_ISL_17609865, EPI_ISL_17609866, EPI_ISL_17609867, EPI_ISL_17609868, EPI_ISL_17609869, EPI_ISL_17609870, EPI_ISL_17609871, EPI_ISL_17609872, EPI_ISL_17609873, EPI_ISL_17609874, EPI_ISL_17609875, EPI_ISL_17609876, EPI_ISL_17609877, EPI_ISL_17609878, EPI_ISL_17609879, EPI_ISL_17609880, EPI_ISL_17609881, EPI_ISL_17609882, EPI_ISL_17609883, EPI_ISL_17609884, EPI_ISL_17609885, EPI_ISL_17609886, EPI_ISL_17609887, EPI_ISL_17609888, EPI_ISL_17609889, EPI_ISL_17609890, EPI_ISL_17609891, EPI_ISL_17609892, EPI_ISL_17609893, EPI_ISL_17609894, EPI_ISL_17609895, EPI_ISL_17609896, EPI_ISL_17609897, EPI_ISL_17609898, EPI_ISL_17609899, EPI_ISL_17609900, EPI_ISL_17609901, EPI_ISL_17609902, EPI_ISL_17609903, EPI_ISL_17609904, EPI_ISL_17609905, EPI_ISL_17609906, EPI_ISL_17609907, EPI_ISL_17609908, EPI_ISL_17609909, EPI_ISL_17609910, EPI_ISL_17609911, EPI_ISL_17609912, EPI_ISL_17609913, EPI_ISL_17609914, EPI_ISL_17609915, EPI_ISL_17609916, EPI_ISL_17609917, EPI_ISL_17609918, EPI_ISL_17609919, EPI_ISL_17609920, EPI_ISL_17609921, EPI_ISL_17609922, EPI_ISL_17609923, EPI_ISL_17609924, EPI_ISL_17609925, EPI_ISL_17609926, EPI_ISL_17609927, EPI_ISL_17609928, EPI_ISL_17609929, EPI_ISL_17609930, EPI_ISL_17609931, EPI_ISL_17609932, EPI_ISL_17609933, EPI_ISL_17609934, EPI_ISL_17609935, EPI_ISL_17609936, EPI_ISL_17609937, EPI_ISL_17609938, EPI_ISL_17609939, EPI_ISL_17609940, EPI_ISL_17609941, EPI_ISL_17609942, EPI_ISL_17609943, EPI_ISL_17609944, EPI_ISL_17609945, EPI_ISL_17609946, EPI_ISL_17609947, EPI_ISL_17609948, EPI_ISL_17609949, EPI_ISL_17609950, EPI_ISL_17609951, EPI_ISL_17609952, EPI_ISL_17609953, EPI_ISL_17609954, EPI_ISL_17609955, EPI_ISL_17609956, EPI_ISL_17609957, EPI_ISL_17609958, EPI_ISL_17609959, EPI_ISL_17609960, EPI_ISL_17609961, EPI_ISL_17609962, EPI_ISL_17609963, EPI_ISL_17609964, EPI_ISL_17609965, EPI_ISL_17609966, EPI_ISL_17609967, EPI_ISL_17609968, EPI_ISL_17609969, EPI_ISL_17609970, EPI_ISL_17609971, EPI_ISL_17609972, EPI_ISL_17609973, EPI_ISL_17609974, EPI_ISL_17609975, EPI_ISL_17609976, EPI_ISL_17609977, EPI_ISL_17609978, EPI_ISL_17609979, EPI_ISL_17609980, EPI_ISL_17609981, EPI_ISL_17609982, EPI_ISL_17609983, EPI_ISL_17609984, EPI_ISL_17609985, EPI_ISL_17609986, EPI_ISL_17609987, EPI_ISL_17609988, EPI_ISL_17609989, EPI_ISL_17609990, EPI_ISL_17609991, EPI_ISL_17609992, EPI_ISL_17609993, EPI_ISL_17609994, EPI_ISL_17609995, EPI_ISL_17609996, EPI_ISL_17609997, EPI_ISL_17609998, EPI_ISL_17609999, EPI_ISL_17610000, EPI_ISL_17610013, EPI_ISL_17610071, EPI_ISL_17610089, EPI_ISL_17610090, EPI_ISL_17610091, EPI_ISL_17610092, EPI_ISL_17610093, EPI_ISL_17610094, EPI_ISL_17610095, EPI_ISL_17610096, EPI_ISL_17610097, EPI_ISL_17610098, EPI_ISL_17610099, EPI_ISL_17610100, EPI_ISL_17610101, EPI_ISL_17610102, EPI_ISL_17610103, EPI_ISL_17610104, EPI_ISL_17610105, EPI_ISL_17610106, EPI_ISL_17610107, EPI_ISL_17610108, EPI_ISL_17610109, EPI_ISL_17610110, EPI_ISL_17610111, EPI_ISL_17610112, EPI_ISL_17610113, EPI_ISL_17610114, EPI_ISL_17610115, EPI_ISL_17610116, EPI_ISL_17610117, EPI_ISL_17610118, EPI_ISL_17610119, EPI_ISL_17610120, EPI_ISL_17610121, EPI_ISL_17610122, EPI_ISL_17610123, EPI_ISL_17610124, EPI_ISL_17610125, EPI_ISL_17610126, EPI_ISL_17610127, EPI_ISL_17610128, EPI_ISL_17610129, EPI_ISL_17610130, EPI_ISL_17610131, EPI_ISL_17610132, EPI_ISL_17610133, EPI_ISL_17610134, EPI_ISL_17610135, EPI_ISL_17610136, EPI_ISL_17610137, EPI_ISL_17610138, EPI_ISL_17610139, EPI_ISL_17610140, EPI_ISL_17610141, EPI_ISL_17610142, EPI_ISL_17610143, EPI_ISL_17610144, EPI_ISL_17610145, EPI_ISL_17610146, EPI_ISL_17610147, EPI_ISL_17610148, EPI_ISL_17610149, EPI_ISL_17610150, EPI_ISL_17610151, EPI_ISL_17610152, EPI_ISL_17610153, EPI_ISL_17610154, EPI_ISL_17610155, EPI_ISL_17610156, EPI_ISL_17610157, EPI_ISL_17610158, EPI_ISL_17610159, EPI_ISL_17610160, EPI_ISL_17610161, EPI_ISL_17610162, EPI_ISL_17610163, EPI_ISL_17610164, EPI_ISL_17610165, EPI_ISL_17610166, EPI_ISL_17610167, EPI_ISL_17610168, EPI_ISL_17610169, EPI_ISL_17610170, EPI_ISL_17610171, EPI_ISL_17610172, EPI_ISL_17610173, EPI_ISL_17610174, EPI_ISL_17610175, EPI_ISL_17610176, EPI_ISL_17610177, EPI_ISL_17610178, EPI_ISL_17610179, EPI_ISL_17610180, EPI_ISL_17610181, EPI_ISL_17610182, EPI_ISL_17610183, EPI_ISL_17610184, EPI_ISL_17610185, EPI_ISL_17610186, EPI_ISL_17610187, EPI_ISL_17610188, EPI_ISL_17610189, EPI_ISL_17610190, EPI_ISL_17610191, EPI_ISL_17610192, EPI_ISL_17610193, EPI_ISL_17610194, EPI_ISL_17610195, EPI_ISL_17610196, EPI_ISL_17610197, EPI_ISL_17610198, EPI_ISL_17610199, EPI_ISL_17610200, EPI_ISL_17610201, EPI_ISL_17610202, EPI_ISL_17610203, EPI_ISL_17610204, EPI_ISL_17610205, EPI_ISL_17610206, EPI_ISL_17610207, EPI_ISL_17610208, EPI_ISL_17610209, EPI_ISL_17610210, EPI_ISL_17610211, EPI_ISL_17610212, EPI_ISL_17610213, EPI_ISL_17610214, EPI_ISL_17610215, EPI_ISL_17610216, EPI_ISL_17610217, EPI_ISL_17610218, EPI_ISL_17610219, EPI_ISL_17610220, EPI_ISL_17610221, EPI_ISL_17610222, EPI_ISL_17610223, EPI_ISL_17610224, EPI_ISL_17610225, EPI_ISL_17610226, EPI_ISL_17610227, EPI_ISL_17610228, EPI_ISL_17610229, EPI_ISL_17610230, EPI_ISL_17610231, EPI_ISL_17610232, EPI_ISL_17610233, EPI_ISL_17610234, EPI_ISL_17610235, EPI_ISL_17610236, EPI_ISL_17610237, EPI_ISL_17610238, EPI_ISL_17610239, EPI_ISL_17610240, EPI_ISL_17610241, EPI_ISL_17610242, EPI_ISL_17610243, EPI_ISL_17610244, EPI_ISL_17610245, EPI_ISL_17610246, EPI_ISL_17610247, EPI_ISL_17610248, EPI_ISL_17610249, EPI_ISL_17610250, EPI_ISL_17610251, EPI_ISL_17610252, EPI_ISL_17610253, EPI_ISL_17610254, EPI_ISL_17610255, EPI_ISL_17610256, EPI_ISL_17610257, EPI_ISL_17610258, EPI_ISL_17610259, EPI_ISL_17610260, EPI_ISL_17610261, EPI_ISL_17610262, EPI_ISL_17610263, EPI_ISL_17610264, EPI_ISL_17610265, EPI_ISL_17610266, EPI_ISL_17610267, EPI_ISL_17610268, EPI_ISL_17610269, EPI_ISL_17610270, EPI_ISL_17610271, EPI_ISL_17610272, EPI_ISL_17610273, EPI_ISL_17610274, EPI_ISL_17610275, EPI_ISL_17610276, EPI_ISL_17610277, EPI_ISL_17610278, EPI_ISL_17610279, EPI_ISL_17610280, EPI_ISL_17610281, EPI_ISL_17610282, EPI_ISL_17610283, EPI_ISL_17610284, EPI_ISL_17610285, EPI_ISL_17610286, EPI_ISL_17610287, EPI_ISL_17610288, EPI_ISL_17610289, EPI_ISL_17610290, EPI_ISL_17610291, EPI_ISL_17610292, EPI_ISL_17610293, EPI_ISL_17610294, EPI_ISL_17610295, EPI_ISL_17610296, EPI_ISL_17610297, EPI_ISL_17610298, EPI_ISL_17610299, EPI_ISL_17610300, EPI_ISL_17610301, EPI_ISL_17610302, EPI_ISL_17610303, EPI_ISL_17610304, EPI_ISL_17610305, EPI_ISL_17610306, EPI_ISL_17610307, EPI_ISL_17610308, EPI_ISL_17610309, EPI_ISL_17610310, EPI_ISL_17610311, EPI_ISL_17610312, EPI_ISL_17610313, EPI_ISL_17610314, EPI_ISL_17610315, EPI_ISL_17610316, EPI_ISL_17610317, EPI_ISL_17610318, EPI_ISL_17610319, EPI_ISL_17610320, EPI_ISL_17610321, EPI_ISL_17610322, EPI_ISL_17610323, EPI_ISL_17610324, EPI_ISL_17610325, EPI_ISL_17610326, EPI_ISL_17610327, EPI_ISL_17610328, EPI_ISL_17610329, EPI_ISL_17610330, EPI_ISL_17610331, EPI_ISL_17610332, EPI_ISL_17610333, EPI_ISL_17610334, EPI_ISL_17610335, EPI_ISL_17610336, EPI_ISL_17610337, EPI_ISL_17610338, EPI_ISL_17610339, EPI_ISL_17610340, EPI_ISL_17610341, EPI_ISL_17610342, EPI_ISL_17610343, EPI_ISL_17610344, EPI_ISL_17610345, EPI_ISL_17610346, EPI_ISL_17610347, EPI_ISL_17610348, EPI_ISL_17610349, EPI_ISL_17610350, EPI_ISL_17610351, EPI_ISL_17610352, EPI_ISL_17610353, EPI_ISL_17610354, EPI_ISL_17610355, EPI_ISL_17610356, EPI_ISL_17610357, EPI_ISL_17610358, EPI_ISL_17610359, EPI_ISL_17610360, EPI_ISL_17610361, EPI_ISL_17610362, EPI_ISL_17610363, EPI_ISL_17610364, EPI_ISL_17610365, EPI_ISL_17610366, EPI_ISL_17610367, EPI_ISL_17610368, EPI_ISL_17610369, EPI_ISL_17610370, EPI_ISL_17610371, EPI_ISL_17610372, EPI_ISL_17610373, EPI_ISL_17610374, EPI_ISL_17610375, EPI_ISL_17610376, EPI_ISL_17610377, EPI_ISL_17610378, EPI_ISL_17610379, EPI_ISL_17610380, EPI_ISL_17610381, EPI_ISL_17610382, EPI_ISL_17610383, EPI_ISL_17610384, EPI_ISL_17610385, EPI_ISL_17610386, EPI_ISL_17610411, EPI_ISL_17610412, EPI_ISL_17610413, EPI_ISL_17610414, EPI_ISL_17610415, EPI_ISL_17610416, EPI_ISL_17610417, EPI_ISL_17610418, EPI_ISL_17610419, EPI_ISL_17610420, EPI_ISL_17610421, EPI_ISL_17610422, EPI_ISL_17610423, EPI_ISL_17610424, EPI_ISL_17610425, EPI_ISL_17610426, EPI_ISL_17610427, EPI_ISL_17610428, EPI_ISL_17610429, EPI_ISL_17610430, EPI_ISL_17610431, EPI_ISL_17610432, EPI_ISL_17610433, EPI_ISL_17610434, EPI_ISL_17610435, EPI_ISL_17610436, EPI_ISL_17610437, EPI_ISL_17610438, EPI_ISL_17610439, EPI_ISL_17610440, EPI_ISL_17610441, EPI_ISL_17610442, EPI_ISL_17610443, EPI_ISL_17610444, EPI_ISL_17610445, EPI_ISL_17610446, EPI_ISL_17610447, EPI_ISL_17610448, EPI_ISL_17610449, EPI_ISL_17610450, EPI_ISL_17610451, EPI_ISL_17610452, EPI_ISL_17610453, EPI_ISL_17610454 | | | |
| see above | London School of Hygiene and Tropical Medicine | London School of Hygiene and Tropical Medicine | Ashall,J., Shah,S., Biggs,J.R., Chang,R., Jafari,Y., Brady,O.J., Mai,H.K., Lien,L.T., Thai,H.D., Nhuyen,H.A.T., Anh,D., Iwasaki,C., Kitamura,N., Van Loock,M., Herrera-Taracena,G., Rasschaert,F., Van Wesenbeeck,L., Yoshida,L.-M., Hafalla,J.C.R., Hue,S. and Hibberd,M.L. |
| EPI_ISL_17610808, EPI_ISL_17610809, EPI_ISL_17610810, EPI_ISL_17610811, EPI_ISL_17610812, EPI_ISL_17610813, EPI_ISL_17610814, EPI_ISL_17610815, EPI_ISL_17610816, EPI_ISL_17610817, EPI_ISL_17610818, EPI_ISL_17610819, EPI_ISL_17610820, EPI_ISL_17610821, EPI_ISL_17610822, EPI_ISL_17610823, EPI_ISL_17610824 | | | |
| see above | Sapporo Medical University, School of Medicine, Department of Hygiene | Sapporo Medical University, School of Medicine, Department of Hygiene | Jahan,A., Shyamal,K., Aung,M.S. and Kobayashi,N. |
| EPI_ISL_17611178, EPI_ISL_17611179, EPI_ISL_17611180, EPI_ISL_17611181, EPI_ISL_17611182, EPI_ISL_17611183 | University of Nebraska Medical Center, Department of Environmental, Agricultural and Occupational Health | University of Nebraska Medical Center, Department of Environmental, Agricultural and Occupational Health | Agbodzi,B., Yousseu,F.B.S., Simo,F.B.N., Kumordjie,S., Yeboah,C., Mosore,M.-T., Bentil,R.E., Attram,N., Nimo-Paintsil,S., Fox,A.T., Bonney,J.H.K., Ampofo,W., Sanders,T., Wiley,M.R., Demanou,M. and Letizia,A.G. |
| EPI_ISL_17611185 | Uppsala University, Zoonosis Science Center | Uppsala University, Zoonosis Science Center | Nguyen-Tien,T., Ling,J. and Lindalh,J. |
| EPI_ISL_17611186 | Virus Laboratory, Indian Council of Medical Research, National Institute of Cholera and Enteric Diseases | Virus Laboratory, Indian Council of Medical Research, National Institute of Cholera and Enteric Diseases | Baskey,U., Verma,P., Ghosh,A. and Sadhukhan,P.C. |
| EPI_ISL_17658722 | Yanbian University | Yanbian University | Li,C. |
| EPI_ISL_17658723, EPI_ISL_17658724, EPI_ISL_17658725, EPI_ISL_17658726, EPI_ISL_17658727, EPI_ISL_17658728 | Department of Epidemiology of Microbial Diseases, Yale School of Public Health | Department of Epidemiology of Microbial Diseases, Yale School of Public Health | Vogels,C.B.F., Breban,M.I., Chaguza,C., Hill,V., Paul,L., Michael,S., Bunch,S., Cano,N., Jaber,R., Morrison,A., Panzera,C., Stryker,I., Vergara,J., Zimler,R., Kopp,E., Herberlein,L. and Grubaugh,N.D. |
| EPI_ISL_17658729, EPI_ISL_17658730, EPI_ISL_17658731, EPI_ISL_17658732, EPI_ISL_17658733, EPI_ISL_17658734, EPI_ISL_17658735, EPI_ISL_17658736, EPI_ISL_17658737, EPI_ISL_17658738, EPI_ISL_17658739, EPI_ISL_17658740, EPI_ISL_17658741 | | | |
| see above | Institute of Medical Biology, Chinese Academy of Medical Sciences, Peking Union Medical College, Tsinghua University | Institute of Medical Biology, Chinese Academy of Medical Sciences, Peking Union Medical College, Tsinghua University | Zhou,Y., Zhou,Y., Liyang,S., Wang,P., An,J. and Liu,H. |
| EPI_ISL_17658742 | National Institutes of Health, National Center for Biotechnology Information | National Institutes of Health, National Center for Biotechnology Information | Osatomi,K. and Sumiyoshi,H. |
| EPI_ISL_17658743 | National Institutes of Health, National Center for Biotechnology Information | National Institutes of Health, National Center for Biotechnology Information | Durbin,A.P., Karron,R.A., Sun,W., Vaughn,D.W., Reynolds,M.J., Perreault,J.R., Thumar,B., Men,R., Lai,C.J., Elkins,W.R., Chanock,R.M., Murphy,B.R. and Whitehead,S.S. |
| EPI_ISL_17658986 | Virology Unit, Institut Pasteur de Dakar | Virology Unit, Institut Pasteur de Dakar | Lagare,A., Faye,M., Fintan,G., Fall,G., Ousmane,H., Ibrahim,E.T., Diagne,M.M., Amadou,S., Sankhe,S., Ibrahim,L., Seini,H., Faye,O. and Jambou,R. |
| EPI_ISL_17658987 | Department of Microbiology, Calcutta School of Tropical Medicine | Department of Microbiology, Calcutta School of Tropical Medicine | Tripathi,A. and Mukherjee,S. |
| EPI_ISL_17658988 | Virology Unit, Institut Pasteur de Dakar | Virology Unit, Institut Pasteur de Dakar | Dieng,I. |
| EPI_ISL_17658989 | Department of Microbiology, King George's Medical University | Department of Microbiology, King George's Medical University | Mallikarjun,K., Santhosh,K.S. and Puneeth,T.G. |
| EPI_ISL_17673145, EPI_ISL_17673146, EPI_ISL_17673147, EPI_ISL_17673148, EPI_ISL_17673149, EPI_ISL_17673150 | Instituto Adolfo Lutz - Strategic Laboratory | Instituto Adolfo Lutz - Strategic Laboratory | Claudio Tavares Sacchi, Karoline Rodrigues Campos, Marlon Benedito Nascimento Santos, Juliana Silva Nogueira |
| EPI_ISL_17673494 | Vector-borne Diseases Laboratory, Instituto Adolfo Lutz | Vector-borne Diseases Laboratory, Instituto Adolfo Lutz | Cunha,M.S., de Moura Coletti,T., Guerra,J.M., Ponce,C.C., Fernandes,N.C.C.A., Resio,R.A., Claro,I.M., Salles,F., Lima Neto,D.F. and Sabino,E. |
| EPI_ISL_17673495 | Instituto Adolfo Lutz - Strategic Laboratory | Instituto Adolfo Lutz - Strategic Laboratory | Sacchi,C.T., Campos,K.R., Santos,M.B., Reis,A.D., Nogueira,J.S. and Maeda,A.Y. |
| EPI_ISL_17673496, EPI_ISL_17673497, EPI_ISL_17673498, EPI_ISL_17673499, EPI_ISL_17673500, EPI_ISL_17673501, EPI_ISL_17673502, EPI_ISL_17673503, EPI_ISL_17673504, EPI_ISL_17673505, EPI_ISL_17673506, EPI_ISL_17673507, EPI_ISL_17673508, EPI_ISL_17673509, EPI_ISL_17673510, EPI_ISL_17673511, EPI_ISL_17673512, EPI_ISL_17673513, EPI_ISL_17673514, EPI_ISL_17673515, EPI_ISL_17673516, EPI_ISL_17673517, EPI_ISL_17673518, EPI_ISL_17673519, EPI_ISL_17673520, EPI_ISL_17673521, EPI_ISL_17673522, EPI_ISL_17673523, EPI_ISL_17673524, EPI_ISL_17673525, EPI_ISL_17673526, EPI_ISL_17673527, EPI_ISL_17673528, EPI_ISL_17673529, EPI_ISL_17673530, EPI_ISL_17673531, EPI_ISL_17673532, EPI_ISL_17673533, EPI_ISL_17673534, EPI_ISL_17673535, EPI_ISL_17673536 | | | |
| see above | Instituto Adolfo Lutz - Strategic Laboratory | Instituto Adolfo Lutz - Strategic Laboratory | Claudio Tavares Sacchi, Karoline Rodrigues Campos, Marlon Benedito Nascimento Santos, Juliana Silva Nogueira |
| EPI_ISL_17689847 | LESP Chiapas | Instituto de Diagnostico y Referencia Epidemiologicos (INDRE) | Claudia Wong-Arámbula; Abril Rodríguez-Maldonado; Silvia Rivero-Arredondo; Ruth Madera-Sandoval; Joaquín Quiroz-Mercado; Daniel Duran-Ayala; Luz Torres Rodríguez; Claudia Rosales-Jiménez; Carlos Fuentes-Cuevas; Ernesto Ramírez-González; Fernando González-Domínguez; Lucia Hernández-Rivas, Irma López-Martínez; Mauricio Vázquez-Pichardo. |
| EPI_ISL_17689848 | LESP Chihuahua | Instituto de Diagnostico y Referencia Epidemiologicos (INDRE) | Claudia Wong-Arámbula; Abril Rodríguez-Maldonado; Silvia Rivero-Arredondo; Ruth Madera-Sandoval; Joaquín Quiroz-Mercado; Daniel Duran-Ayala; Luz Torres Rodríguez; Claudia Rosales-Jiménez; Carlos Fuentes-Cuevas; Ernesto Ramírez-González; Fernando González-Domínguez; Lucia Hernández-Rivas, Irma López-Martínez; Mauricio Vázquez-Pichardo. |
| EPI_ISL_17689849 | LESP Sinaloa | Instituto de Diagnostico y Referencia Epidemiologicos (INDRE) | Claudia Wong-Arámbula; Abril Rodríguez-Maldonado; Silvia Rivero-Arredondo; Ruth Madera-Sandoval; Joaquín Quiroz-Mercado; Daniel Duran-Ayala; Luz Torres Rodríguez; Claudia Rosales-Jiménez; Carlos Fuentes-Cuevas; Ernesto Ramírez-González; Fernando González-Domínguez; Lucia Hernández-Rivas, Irma López-Martínez; Mauricio Vázquez-Pichardo. |
| EPI_ISL_17689850 | LESP Sonora | Instituto de Diagnostico y Referencia Epidemiologicos (INDRE) | Claudia Wong-Arámbula; Abril Rodríguez-Maldonado; Silvia Rivero-Arredondo; Ruth Madera-Sandoval; Joaquín Quiroz-Mercado; Daniel Duran-Ayala; Luz Torres Rodríguez; Claudia Rosales-Jiménez; Carlos Fuentes-Cuevas; Ernesto Ramírez-González; Fernando González-Domínguez; Lucia Hernández-Rivas, Irma López-Martínez; Mauricio Vázquez-Pichardo. |

| | | | |
|---|---|---|---|
| EPI_ISL_17689851 | LESP Puebla | Instituto de Diagnostico y Referencia Epidemiologicos (INDRE) | Claudia Wong-Arámbula; Abril Rodríguez-Maldonado; Silvia Rivero-Arredondo; Ruth Madera-Sandoval; Joaquín Quiroz-Mercado; Daniel Duran-Ayala; Luz Torres Rodríguez; Claudia Rosales-Jiménez; Carlos Fuentes-Cuevas; Ernesto Ramírez-González; Fernando González-Domínguez; Lucia Hernández-Rivas, Irma López-Martínez; Mauricio Vázquez-Pichardo. |
| EPI_ISL_17689852, EPI_ISL_17689853 | LESP State of Mexico | Instituto de Diagnostico y Referencia Epidemiologicos (INDRE) | Claudia Wong-Arámbula; Abril Rodríguez-Maldonado; Silvia Rivero-Arredondo; Ruth Madera-Sandoval; Joaquín Quiroz-Mercado; Daniel Duran-Ayala; Luz Torres Rodríguez; Claudia Rosales-Jiménez; Carlos Fuentes-Cuevas; Ernesto Ramírez-González; Fernando González-Domínguez; Lucia Hernández-Rivas, Irma López-Martínez; Mauricio Vázquez-Pichardo. |
| EPI_ISL_17689854, EPI_ISL_17689855, EPI_ISL_17689856 | LESP Baja California | Instituto de Diagnostico y Referencia Epidemiologicos (INDRE) | Claudia Wong-Arámbula; Abril Rodríguez-Maldonado; Silvia Rivero-Arredondo; Ruth Madera-Sandoval; Joaquín Quiroz-Mercado; Daniel Duran-Ayala; Luz Torres Rodríguez; Claudia Rosales-Jiménez; Carlos Fuentes-Cuevas; Ernesto Ramírez-González; Fernando González-Domínguez; Lucia Hernández-Rivas, Irma López-Martínez; Mauricio Vázquez-Pichardo. |
| EPI_ISL_17689857, EPI_ISL_17689858 | LESP Sinaloa | Instituto de Diagnostico y Referencia Epidemiologicos (INDRE) | Claudia Wong-Arámbula; Abril Rodríguez-Maldonado; Silvia Rivero-Arredondo; Ruth Madera-Sandoval; Joaquín Quiroz-Mercado; Daniel Duran-Ayala; Luz Torres Rodríguez; Claudia Rosales-Jiménez; Carlos Fuentes-Cuevas; Ernesto Ramírez-González; Fernando González-Domínguez; Lucia Hernández-Rivas, Irma López-Martínez; Mauricio Vázquez-Pichardo. |
| EPI_ISL_17689859 | LESP State of Mexico | Instituto de Diagnostico y Referencia Epidemiologicos (INDRE) | Claudia Wong-Arámbula; Abril Rodríguez-Maldonado; Silvia Rivero-Arredondo; Ruth Madera-Sandoval; Joaquín Quiroz-Mercado; Daniel Duran-Ayala; Luz Torres Rodríguez; Claudia Rosales-Jiménez; Carlos Fuentes-Cuevas; Ernesto Ramírez-González; Fernando González-Domínguez; Lucia Hernández-Rivas, Irma López-Martínez; Mauricio Vázquez-Pichardo. |
| EPI_ISL_17689860 | LESP Puebla | Instituto de Diagnostico y Referencia Epidemiologicos (INDRE) | Claudia Wong-Arámbula; Abril Rodríguez-Maldonado; Silvia Rivero-Arredondo; Ruth Madera-Sandoval; Joaquín Quiroz-Mercado; Daniel Duran-Ayala; Luz Torres Rodríguez; Claudia Rosales-Jiménez; Carlos Fuentes-Cuevas; Ernesto Ramírez-González; Fernando González-Domínguez; Lucia Hernández-Rivas, Irma López-Martínez; Mauricio Vázquez-Pichardo. |
| EPI_ISL_17689861, EPI_ISL_17689862 | LESP Veracruz | Instituto de Diagnostico y Referencia Epidemiologicos (INDRE) | Claudia Wong-Arámbula; Abril Rodríguez-Maldonado; Silvia Rivero-Arredondo; Ruth Madera-Sandoval; Joaquín Quiroz-Mercado; Daniel Duran-Ayala; Luz Torres Rodríguez; Claudia Rosales-Jiménez; Carlos Fuentes-Cuevas; Ernesto Ramírez-González; Fernando González-Domínguez; Lucia Hernández-Rivas, Irma López-Martínez; Mauricio Vázquez-Pichardo. |
| EPI_ISL_17689863 | LESP Sonora | Instituto de Diagnostico y Referencia Epidemiologicos (INDRE) | Claudia Wong-Arámbula; Abril Rodríguez-Maldonado; Silvia Rivero-Arredondo; Ruth Madera-Sandoval; Joaquín Quiroz-Mercado; Daniel Duran-Ayala; Luz Torres Rodríguez; Claudia Rosales-Jiménez; Carlos Fuentes-Cuevas; Ernesto Ramírez-González; Fernando González-Domínguez; Lucia Hernández-Rivas, Irma López-Martínez; Mauricio Vázquez-Pichardo. |
| EPI_ISL_17689864 | LESP Yucatan | Instituto de Diagnostico y Referencia Epidemiologicos (INDRE) | Claudia Wong-Arámbula; Abril Rodríguez-Maldonado; Silvia Rivero-Arredondo; Ruth Madera-Sandoval; Joaquín Quiroz-Mercado; Daniel Duran-Ayala; Luz Torres Rodríguez; Claudia Rosales-Jiménez; Carlos Fuentes-Cuevas; Ernesto Ramírez-González; Fernando González-Domínguez; Lucia Hernández-Rivas, Irma López-Martínez; Mauricio Vázquez-Pichardo. |
| EPI_ISL_17689865 | LESP Coahuila | Instituto de Diagnostico y Referencia Epidemiologicos (INDRE) | Claudia Wong-Arámbula; Abril Rodríguez-Maldonado; Silvia Rivero-Arredondo; Ruth Madera-Sandoval; Joaquín Quiroz-Mercado; Daniel Duran-Ayala; Luz Torres Rodríguez; Claudia Rosales-Jiménez; Carlos Fuentes-Cuevas; Ernesto Ramírez-González; Fernando González-Domínguez; Lucia Hernández-Rivas, Irma López-Martínez; Mauricio Vázquez-Pichardo. |
| EPI_ISL_17689866 | LESP Veracruz | Instituto de Diagnostico y Referencia Epidemiologicos (INDRE) | Claudia Wong-Arámbula; Abril Rodríguez-Maldonado; Silvia Rivero-Arredondo; Ruth Madera-Sandoval; Joaquín Quiroz-Mercado; Daniel Duran-Ayala; Luz Torres Rodríguez; Claudia Rosales-Jiménez; Carlos Fuentes-Cuevas; Ernesto Ramírez-González; Fernando González-Domínguez; Lucia Hernández-Rivas, Irma López-Martínez; Mauricio Vázquez-Pichardo. |
| EPI_ISL_17689867, EPI_ISL_17689868 | LESP Coahuila | Instituto de Diagnostico y Referencia Epidemiologicos (INDRE) | Claudia Wong-Arámbula; Abril Rodríguez-Maldonado; Silvia Rivero-Arredondo; Ruth Madera-Sandoval; Joaquín Quiroz-Mercado; Daniel Duran-Ayala; Luz Torres Rodríguez; Claudia Rosales-Jiménez; Carlos Fuentes-Cuevas; Ernesto Ramírez-González; Fernando González-Domínguez; Lucia Hernández-Rivas, Irma López-Martínez; Mauricio Vázquez-Pichardo. |
| EPI_ISL_17689869 | LESP Veracruz | Instituto de Diagnostico y Referencia Epidemiologicos (INDRE) | Claudia Wong-Arámbula; Abril Rodríguez-Maldonado; Silvia Rivero-Arredondo; Ruth Madera-Sandoval; Joaquín Quiroz-Mercado; Daniel Duran-Ayala; Luz Torres Rodríguez; Claudia Rosales-Jiménez; Carlos Fuentes-Cuevas; Ernesto Ramírez-González; Fernando González-Domínguez; Lucia Hernández-Rivas, Irma López-Martínez; Mauricio Vázquez-Pichardo. |
| EPI_ISL_17689870 | LESP Tabasco | Instituto de Diagnostico y Referencia Epidemiologicos (INDRE) | Claudia Wong-Arámbula; Abril Rodríguez-Maldonado; Silvia Rivero-Arredondo; Ruth Madera-Sandoval; Joaquín Quiroz-Mercado; Daniel Duran-Ayala; Luz Torres Rodríguez; Claudia Rosales-Jiménez; Carlos Fuentes-Cuevas; Ernesto Ramírez-González; Fernando González-Domínguez; Lucia Hernández-Rivas, Irma López-Martínez; Mauricio Vázquez-Pichardo. |
| EPI_ISL_17689871, EPI_ISL_17689872, EPI_ISL_17689873 | LESP Quintana Roo | Instituto de Diagnostico y Referencia Epidemiologicos (INDRE) | Claudia Wong-Arámbula; Abril Rodríguez-Maldonado; Silvia Rivero-Arredondo; Ruth Madera-Sandoval; Joaquín Quiroz-Mercado; Daniel Duran-Ayala; Luz Torres Rodríguez; Claudia Rosales-Jiménez; Carlos Fuentes-Cuevas; Ernesto Ramírez-González; Fernando González-Domínguez; Lucia Hernández-Rivas, Irma López-Martínez; Mauricio Vázquez-Pichardo. |
| EPI_ISL_17689874, EPI_ISL_17689875, EPI_ISL_17689876 | LESP Colima | Instituto de Diagnostico y Referencia Epidemiologicos (INDRE) | Claudia Wong-Arámbula; Abril Rodríguez-Maldonado; Silvia Rivero-Arredondo; Ruth Madera-Sandoval; Joaquín Quiroz-Mercado; Daniel Duran-Ayala; Luz Torres Rodríguez; Claudia Rosales-Jiménez; Carlos Fuentes-Cuevas; Ernesto Ramírez-González; Fernando González-Domínguez; Lucia Hernández-Rivas, Irma López-Martínez; Mauricio Vázquez-Pichardo. |
| EPI_ISL_17689877, EPI_ISL_17689878 | LESP Tabasco | Instituto de Diagnostico y Referencia Epidemiologicos (INDRE) | Claudia Wong-Arámbula; Abril Rodríguez-Maldonado; Silvia Rivero-Arredondo; Ruth Madera-Sandoval; Joaquín Quiroz-Mercado; Daniel Duran-Ayala; Luz Torres Rodríguez; Claudia Rosales-Jiménez; Carlos Fuentes-Cuevas; Ernesto Ramírez-González; Fernando González-Domínguez; Lucia Hernández-Rivas, Irma López-Martínez; Mauricio Vázquez-Pichardo. |
| EPI_ISL_17694637, EPI_ISL_17694638 | Department of Microbiology, Calcutta School of Tropical Medicine | Department of Microbiology, Calcutta School of Tropical Medicine | Tripathi,A. and Mukherjee,S. |
| EPI_ISL_17694639, EPI_ISL_17694640, EPI_ISL_17694641, EPI_ISL_17694642, EPI_ISL_17694643, EPI_ISL_17694644 | Institute of Medical Biology, Chinese Academy of Medical Sciences, Peking Union Medical College, Tsinghua University | Institute of Medical Biology, Chinese Academy of Medical Sciences, Peking Union Medical College, Tsinghua University | Zhou,Y., Zhou,Y., Liyang,S., Wang,P., An,J. and Liu,H. |
| EPI_ISL_17694645, EPI_ISL_17694646, EPI_ISL_17694647, EPI_ISL_17694648, EPI_ISL_17694649 | Department of Microbiology, Calcutta School of Tropical Medicine | Department of Microbiology, Calcutta School of Tropical Medicine | Tripathi,A. and Mukherjee,S. |
| EPI_ISL_17699727 | LACEN de Sergipe | Instituto Adolfo Lutz - Strategic Laboratory | Claudio Tavares Sacchi, Karoline Rodrigues Campos, Marlon Benedito Nascimento Santos, Gabriela Vasconcelos Brito Bezerra |
| EPI_ISL_17699728, EPI_ISL_17699729, EPI_ISL_17699730, EPI_ISL_17699731, EPI_ISL_17699732, EPI_ISL_17699733, EPI_ISL_17699734, EPI_ISL_17699735, EPI_ISL_17699736, EPI_ISL_17699737, EPI_ISL_17699738, EPI_ISL_17699739, EPI_ISL_17699740, EPI_ISL_17699741, EPI_ISL_17699742, EPI_ISL_17699743, EPI_ISL_17699744, EPI_ISL_17699745, EPI_ISL_17699746, EPI_ISL_17699747, EPI_ISL_17699748, EPI_ISL_17699749, EPI_ISL_17699750, EPI_ISL_17699751, EPI_ISL_17699752, EPI_ISL_17699753, EPI_ISL_17699754, EPI_ISL_17699755, EPI_ISL_17699756, EPI_ISL_17699757, EPI_ISL_17699758, EPI_ISL_17699759, EPI_ISL_17699760, EPI_ISL_17699761, EPI_ISL_17699762, EPI_ISL_17699763, EPI_ISL_17699764, EPI_ISL_17699766, EPI_ISL_17699767, EPI_ISL_17699768, EPI_ISL_17699769, EPI_ISL_17699770, EPI_ISL_17699771, EPI_ISL_17699772, EPI_ISL_17699773, EPI_ISL_17699774, EPI_ISL_17699775, EPI_ISL_17699776, EPI_ISL_17699777 | | | |
| see above | Instituto Adolfo Lutz - Strategic Laboratory | Instituto Adolfo Lutz - Strategic Laboratory | Claudio Tavares Sacchi, Karoline Rodrigues Campos, Marlon Benedito Nascimento Santos, Juliana Silva Nogueira |
| EPI_ISL_17699778, EPI_ISL_17699779, EPI_ISL_17699780, EPI_ISL_17699781, EPI_ISL_17699782, EPI_ISL_17699783, EPI_ISL_17699784, EPI_ISL_17699785 | LACEN de Santa Catarina | Instituto Adolfo Lutz - Strategic Laboratory | Claudio Tavares Sacchi, Karoline Rodrigues Campos, Marlon Benedito Nascimento Santos |
| EPI_ISL_17699786, EPI_ISL_17699787 | LACEN de Sergipe | Instituto Adolfo Lutz - Strategic Laboratory | Claudio Tavares Sacchi, Karoline Rodrigues Campos, Marlon Benedito Nascimento Santos, Gabriela Vasconcelos Brito Bezerra |
| EPI_ISL_17699788, EPI_ISL_17699789, EPI_ISL_17699796, EPI_ISL_17699797, EPI_ISL_17699798, EPI_ISL_17699799, EPI_ISL_17699800, EPI_ISL_17699801, EPI_ISL_17699802, EPI_ISL_17699803, EPI_ISL_17699804, EPI_ISL_17699805, EPI_ISL_17699806, EPI_ISL_17699807, EPI_ISL_17699808, EPI_ISL_17699809, EPI_ISL_17699810, EPI_ISL_17699811, EPI_ISL_17699812, EPI_ISL_17699813, EPI_ISL_17699814, EPI_ISL_17699815, EPI_ISL_17699816, EPI_ISL_17699817 | | | |
| see above | LACEN do Rio de Janeiro | Instituto Adolfo Lutz - Strategic Laboratory | Claudio Tavares Sacchi, Karoline Rodrigues Campos, Marlon Benedito Nascimento Santos, Leandro Magalhaes de Souza |
| EPI_ISL_17733558, EPI_ISL_17733559, EPI_ISL_17733560, EPI_ISL_17733561, EPI_ISL_17733562, EPI_ISL_17733563, EPI_ISL_17733565, EPI_ISL_17733566, EPI_ISL_17733567, EPI_ISL_17733568, EPI_ISL_17733569, EPI_ISL_17733570, EPI_ISL_17733571, EPI_ISL_17733572, EPI_ISL_17733573, EPI_ISL_17733574, EPI_ISL_17733575, EPI_ISL_17733576, EPI_ISL_17733577, EPI_ISL_17733578, EPI_ISL_17733580, EPI_ISL_17733581, EPI_ISL_17733582, EPI_ISL_17733583, EPI_ISL_17733584, EPI_ISL_17733585, EPI_ISL_17733586, EPI_ISL_17733587, EPI_ISL_17733588, EPI_ISL_17733589, EPI_ISL_17733590, EPI_ISL_17733591, EPI_ISL_17733592, EPI_ISL_17733594, EPI_ISL_17733595, EPI_ISL_17733596, EPI_ISL_17733597, EPI_ISL_17733598, EPI_ISL_17733599, EPI_ISL_17733600, EPI_ISL_17733601, EPI_ISL_17733602, EPI_ISL_17733603, EPI_ISL_17733604, EPI_ISL_17733605, EPI_ISL_17733606, EPI_ISL_17733608, EPI_ISL_17733609, EPI_ISL_17733610, EPI_ISL_17733611, EPI_ISL_17733612, EPI_ISL_17733613, EPI_ISL_17733614, EPI_ISL_17733615, EPI_ISL_17733616, EPI_ISL_17733617, EPI_ISL_17733618, EPI_ISL_17733619, EPI_ISL_17733620, EPI_ISL_17733621, EPI_ISL_17733623, EPI_ISL_17733624, EPI_ISL_17733625, EPI_ISL_17733626, EPI_ISL_17733627, EPI_ISL_17733628, EPI_ISL_17733629, EPI_ISL_17733630, EPI_ISL_17733631, EPI_ISL_17733632, EPI_ISL_17733633, EPI_ISL_17733634, EPI_ISL_17733635, EPI_ISL_17733637, EPI_ISL_17733638, EPI_ISL_17733639, EPI_ISL_17733640, EPI_ISL_17733641, EPI_ISL_17733642, EPI_ISL_17733643, EPI_ISL_17733644, EPI_ISL_17733645, EPI_ISL_17733646, EPI_ISL_17733647, EPI_ISL_17733648, EPI_ISL_17733649, EPI_ISL_17733650, EPI_ISL_17733651, EPI_ISL_17733652, EPI_ISL_17733653, EPI_ISL_17733654, EPI_ISL_17733655, EPI_ISL_17733656, EPI_ISL_17733657, EPI_ISL_17733658, EPI_ISL_17733659, EPI_ISL_17733660, EPI_ISL_17733661, EPI_ISL_17733662, EPI_ISL_17733663, EPI_ISL_17733664, EPI_ISL_17733665, EPI_ISL_17733666, EPI_ISL_17733667, EPI_ISL_17733668, EPI_ISL_17733669, EPI_ISL_17733670, EPI_ISL_17733671, EPI_ISL_17733672, EPI_ISL_17733673, EPI_ISL_17733674, EPI_ISL_17733675, EPI_ISL_17733676, EPI_ISL_17733677, EPI_ISL_17733678, EPI_ISL_17733679, EPI_ISL_17733680, EPI_ISL_17733681, EPI_ISL_17733682, EPI_ISL_17733683, EPI_ISL_17733684, EPI_ISL_17733685, EPI_ISL_17733686, EPI_ISL_17733687, EPI_ISL_17733688, EPI_ISL_17733689, EPI_ISL_17733690, EPI_ISL_17733691, EPI_ISL_17733692, EPI_ISL_17733693, EPI_ISL_17733694, EPI_ISL_17733695 | | | |
| see above | Central Public Health Laboratory, State Health Surveillance Center of the Rio Grande do Sul State Health Department (LACEN/CEVS/SES-RS) | Center for Scientific and Technological Development, State Center for Health Surveillance of the Secretary of Health of the State of Rio Grande do Sul (CDCT/CEVS/SES-RS) | Fernanda Godinho, Tatiana Schäffer Gregianini, Amanda Pellenz Ruivo, Júlio Augusto Schroder, Viviane Horn, Franciellen Machado dos Santos, Regina Bones Barcellos, Juliano de Oliveira Silveira, Fernanda Letícia Martiny, Érica Bortoli Möllmann, Gabriel da Luz Wallau, Richard Steiner Salvato |
| EPI_ISL_17733704, EPI_ISL_17733705, EPI_ISL_17733706, EPI_ISL_17733707, EPI_ISL_17733708, EPI_ISL_17733709, EPI_ISL_17733710, EPI_ISL_17733711, EPI_ISL_17733712, EPI_ISL_17733713, EPI_ISL_17733714, EPI_ISL_17733715, EPI_ISL_17733716, EPI_ISL_17733717, EPI_ISL_17733718, EPI_ISL_17733719, EPI_ISL_17733720, EPI_ISL_17733721, EPI_ISL_17733722, EPI_ISL_17733723, EPI_ISL_17733724, EPI_ISL_17733725, EPI_ISL_17733726, EPI_ISL_17733727, EPI_ISL_17733728, EPI_ISL_17733729, EPI_ISL_17733730, EPI_ISL_17733731, EPI_ISL_17733732, EPI_ISL_17733733, EPI_ISL_17733734, EPI_ISL_17733735, EPI_ISL_17733736, EPI_ISL_17733737, EPI_ISL_17733738, EPI_ISL_17733739, EPI_ISL_17733740, EPI_ISL_17733741, EPI_ISL_17733742, EPI_ISL_17733743, EPI_ISL_17733744, EPI_ISL_17733745, EPI_ISL_17733746, EPI_ISL_17733747, EPI_ISL_17733748, EPI_ISL_17733749, EPI_ISL_17733750, EPI_ISL_17733751, EPI_ISL_17733752, EPI_ISL_17733753, EPI_ISL_17733754, EPI_ISL_17733755, EPI_ISL_17733756, EPI_ISL_17733757, EPI_ISL_17733758, EPI_ISL_17733759, EPI_ISL_17733760, EPI_ISL_17733761, EPI_ISL_17733762, EPI_ISL_17733763, EPI_ISL_17733764, EPI_ISL_17733765, EPI_ISL_17733766, EPI_ISL_17733767, EPI_ISL_17733768, EPI_ISL_17733769, EPI_ISL_17733770, EPI_ISL_17733771, EPI_ISL_17733772, EPI_ISL_17733773, EPI_ISL_17733774, EPI_ISL_17733775, EPI_ISL_17733776, EPI_ISL_17733777, EPI_ISL_17733778, EPI_ISL_17733779, EPI_ISL_17733780, EPI_ISL_17733781, EPI_ISL_17733782, EPI_ISL_17733783, EPI_ISL_17733784, EPI_ISL_17733785, EPI_ISL_17733786, EPI_ISL_17733787, EPI_ISL_17733788, EPI_ISL_17733789, EPI_ISL_17733790, EPI_ISL_17733791, EPI_ISL_17733792, EPI_ISL_17733793, EPI_ISL_17733794, EPI_ISL_17733795, EPI_ISL_17733796, EPI_ISL_17733797, EPI_ISL_17733798, EPI_ISL_17733799, EPI_ISL_17733800, EPI_ISL_17733801, EPI_ISL_17733802, EPI_ISL_17733803, EPI_ISL_17733804, EPI_ISL_17733805, EPI_ISL_17733806, EPI_ISL_17733807, EPI_ISL_17733808, EPI_ISL_17733809, EPI_ISL_17733810, EPI_ISL_17733811, EPI_ISL_17733812, EPI_ISL_17733813, EPI_ISL_17733814, EPI_ISL_17733815, EPI_ISL_17733816, EPI_ISL_17733817, EPI_ISL_17733818, EPI_ISL_17733819, EPI_ISL_17733820, EPI_ISL_17733821, EPI_ISL_17733822, EPI_ISL_17733823, EPI_ISL_17733824, EPI_ISL_17733825, EPI_ISL_17733826, EPI_ISL_17733827, EPI_ISL_17733828, EPI_ISL_17733829, EPI_ISL_17733830, EPI_ISL_17733831, EPI_ISL_17733832, EPI_ISL_17733833, EPI_ISL_17733834, EPI_ISL_17733835, EPI_ISL_17733836, EPI_ISL_17733837, EPI_ISL_17733838, EPI_ISL_17733839, EPI_ISL_17733840, EPI_ISL_17733841, EPI_ISL_17733842, EPI_ISL_17733843, EPI_ISL_17733844, EPI_ISL_17733845, EPI_ISL_17733846, EPI_ISL_17733847, EPI_ISL_17733848, EPI_ISL_17733849, EPI_ISL_17733850, EPI_ISL_17733851, EPI_ISL_17733852, EPI_ISL_17733853, EPI_ISL_17733854, EPI_ISL_17733855, EPI_ISL_17733856, EPI_ISL_17733857, EPI_ISL_17733858, EPI_ISL_17733859, EPI_ISL_17733860, EPI_ISL_17733861, EPI_ISL_17733862, EPI_ISL_17733863, EPI_ISL_17733864, EPI_ISL_17733865, EPI_ISL_17733866, EPI_ISL_17733867, EPI_ISL_17733868, EPI_ISL_17733869, EPI_ISL_17733870, EPI_ISL_17733871, EPI_ISL_17733872, EPI_ISL_17733873, EPI_ISL_17733874, EPI_ISL_17733875, EPI_ISL_17733876, EPI_ISL_17733877, EPI_ISL_17733878, EPI_ISL_17733879, EPI_ISL_17733880, EPI_ISL_17733881, EPI_ISL_17733882, EPI_ISL_17733883, EPI_ISL_17733884, EPI_ISL_17733885, EPI_ISL_17733886, EPI_ISL_17733887, EPI_ISL_17733888, EPI_ISL_17733889, EPI_ISL_17733890, EPI_ISL_17733891, EPI_ISL_17733892, EPI_ISL_17733893, EPI_ISL_17733894, EPI_ISL_17733895, EPI_ISL_17733896, EPI_ISL_17733897, EPI_ISL_17733898, EPI_ISL_17733899, EPI_ISL_17733900, EPI_ISL_17733901, EPI_ISL_17733902, EPI_ISL_17733903, EPI_ISL_17733904, EPI_ISL_17733905, EPI_ISL_17733906, EPI_ISL_17733907, EPI_ISL_17733908, EPI_ISL_17733909, EPI_ISL_17733910, EPI_ISL_17733911, EPI_ISL_17733912, EPI_ISL_17733913, EPI_ISL_17733914, EPI_ISL_17733915, EPI_ISL_17733916, EPI_ISL_17733917, EPI_ISL_17733918, EPI_ISL_17733919, EPI_ISL_17733920, EPI_ISL_17733921, EPI_ISL_17733922, EPI_ISL_17733923, EPI_ISL_17733924, EPI_ISL_17733925, EPI_ISL_17733926, EPI_ISL_17733927, EPI_ISL_17733928, EPI_ISL_17733929, EPI_ISL_17733930, EPI_ISL_17733931, EPI_ISL_17733932, EPI_ISL_17733933, EPI_ISL_17733934, EPI_ISL_17733935, EPI_ISL_17733936, EPI_ISL_17733937, EPI_ISL_17733938, EPI_ISL_17733939, EPI_ISL_17733940, EPI_ISL_17733941, EPI_ISL_17733942, EPI_ISL_17733943, EPI_ISL_17733944, EPI_ISL_17733945, EPI_ISL_17733946, EPI_ISL_17733947, EPI_ISL_17733948, EPI_ISL_17733949, EPI_ISL_17733950, EPI_ISL_17733951, EPI_ISL_17733952, EPI_ISL_17733953, EPI_ISL_17733954, EPI_ISL_17733955 | | | |
| see above | Guangdong Provincial Center for Disease Control and Prevention | Guangdong Provincial Center for Disease Control and Prevention | Sun,J. |
| EPI_ISL_17734039, EPI_ISL_17734040, EPI_ISL_17734041, EPI_ISL_17734045, EPI_ISL_17734046, EPI_ISL_17734047, EPI_ISL_17734048, EPI_ISL_17734049, EPI_ISL_17734050, EPI_ISL_17734051, EPI_ISL_17734052, EPI_ISL_17734053, EPI_ISL_17734054, EPI_ISL_17734055, EPI_ISL_17734056, EPI_ISL_17734057, EPI_ISL_17734058, EPI_ISL_17734059, EPI_ISL_17734060, EPI_ISL_17734061, EPI_ISL_17734062, EPI_ISL_17734063, EPI_ISL_17734064 | | | |
| see above | Fundação Oswaldo Cruz, Instituto Carlos Chagas | Fundação Oswaldo Cruz, Instituto Carlos Chagas | Graf,T., Ferreira,C.N., Presibella,M., Fernandes,S.B. and Duarte dos Santos,C.N. |
| EPI_ISL_17744309 | Laboratorio Central de Saude Publica "Dr. Milton Bezerra Sobral" LACEN PE | Fiocruz Pernambuco - Instituto Aggeu Magalhaes | Gustavo Barbosa de Lima, Raul Emídio de Lima, Lais Ceschini Machado, Tulio de Lima Campos, Gabriel Luz Wallau |
| EPI_ISL_17772334, EPI_ISL_17772335 | Department of Microbiology, Calcutta School of Tropical Medicine | Department of Microbiology, Calcutta School of Tropical Medicine | Tripathi,A. and Mukherjee,S. |
| EPI_ISL_17783293, EPI_ISL_17783294, EPI_ISL_17783295, EPI_ISL_17783296, EPI_ISL_17783297, EPI_ISL_17783298, EPI_ISL_17783299, EPI_ISL_17783300, EPI_ISL_17783301, EPI_ISL_17783302, EPI_ISL_17783303, EPI_ISL_17783304, EPI_ISL_17783305, EPI_ISL_17783306, EPI_ISL_17783307, EPI_ISL_17783308, EPI_ISL_17783309, EPI_ISL_17783310, EPI_ISL_17783311, EPI_ISL_17783312, EPI_ISL_17783313, EPI_ISL_17783314, EPI_ISL_17783315, EPI_ISL_17783316, EPI_ISL_17783317, EPI_ISL_17783318, EPI_ISL_17783319, EPI_ISL_17783320, EPI_ISL_17783321, EPI_ISL_17783322, EPI_ISL_17783323, EPI_ISL_17783325, EPI_ISL_17783326, EPI_ISL_17783327, EPI_ISL_17783328, EPI_ISL_17783329, EPI_ISL_17783330, EPI_ISL_17783331, EPI_ISL_17783332, EPI_ISL_17783333, EPI_ISL_17783334, EPI_ISL_17783335, EPI_ISL_17783336 | | | |
| see above | Grupo de Morfologia Celular, Instituto Nacional de Salud | Grupo de Morfologia Celular, Instituto Nacional de Salud | Laiton-Donato,K.D., Alvarez-Diaz,D.A., Rosales-Munar,A., Pelaez-Carvajal,D., Ajami,N.J. and Usme-Ciro,J.A. |
| EPI_ISL_17783337, EPI_ISL_17783338 | Tokyo Metropolitan Institute of Public Health | Tokyo Metropolitan Institute of Public Health | Okada,W., Kasuya,F., Harada,S., Kumagai,R., Hasegawa,M., Miyake,H., Nagashima,M. and Sadamasu,K. |

| | | | |
|---|---|---|---|
| EPI_ISL_17783339, EPI_ISL_17783340, EPI_ISL_17783341, EPI_ISL_17783342, EPI_ISL_17783343, EPI_ISL_17783344, EPI_ISL_17783345, EPI_ISL_17783346, EPI_ISL_17783347, EPI_ISL_17783348, EPI_ISL_17783349, EPI_ISL_17783350, EPI_ISL_17783351, EPI_ISL_17783352, EPI_ISL_17783353 | | | |
| see above | Grupo de Morfologia Celular, Instituto Nacional de Salud | Grupo de Morfologia Celular, Instituto Nacional de Salud | Laiton-Donato,K.D., Alvarez-Diaz,D.A., Rosales-Munar,A., Pelaez-Carvajal,D., Ajami,N.J. and Usme-Ciro,J.A. |
| EPI_ISL_17798172, EPI_ISL_17798173, EPI_ISL_17798174, EPI_ISL_17798175, EPI_ISL_17798176, EPI_ISL_17798177, EPI_ISL_17798178, EPI_ISL_17798179, EPI_ISL_17798180, EPI_ISL_17798181, EPI_ISL_17798182, EPI_ISL_17798183, EPI_ISL_17798184, EPI_ISL_17798185, EPI_ISL_17798186, EPI_ISL_17798187, EPI_ISL_17798188, EPI_ISL_17798189, EPI_ISL_17798190, EPI_ISL_17798191, EPI_ISL_17798192, EPI_ISL_17798193, EPI_ISL_17798194, EPI_ISL_17798195, EPI_ISL_17798196, EPI_ISL_17798197, EPI_ISL_17798198, EPI_ISL_17798199, EPI_ISL_17798200, EPI_ISL_17798201, EPI_ISL_17798202, EPI_ISL_17798203, EPI_ISL_17798204, EPI_ISL_17798205, EPI_ISL_17798206, EPI_ISL_17798207, EPI_ISL_17798208, EPI_ISL_17798209, EPI_ISL_17798210, EPI_ISL_17798211, EPI_ISL_17798212, EPI_ISL_17798213, EPI_ISL_17798214, EPI_ISL_17798215, EPI_ISL_17798216, EPI_ISL_17798217, EPI_ISL_17798218, EPI_ISL_17798219, EPI_ISL_17798220, EPI_ISL_17798221, EPI_ISL_17798222, EPI_ISL_17798223, EPI_ISL_17798224, EPI_ISL_17798225, EPI_ISL_17798226, EPI_ISL_17798227, EPI_ISL_17798228, EPI_ISL_17798229, EPI_ISL_17798230, EPI_ISL_17798231, EPI_ISL_17798232, EPI_ISL_17798233, EPI_ISL_17798234, EPI_ISL_17798235, EPI_ISL_17798236, EPI_ISL_17798237, EPI_ISL_17798238, EPI_ISL_17798239, EPI_ISL_17798240, EPI_ISL_17798241, EPI_ISL_17798242, EPI_ISL_17798243, EPI_ISL_17798244, EPI_ISL_17798245, EPI_ISL_17798246, EPI_ISL_17798247, EPI_ISL_17798248, EPI_ISL_17798249, EPI_ISL_17798250, EPI_ISL_17798251, EPI_ISL_17798252, EPI_ISL_17798253, EPI_ISL_17798254, EPI_ISL_17798255, EPI_ISL_17798256, EPI_ISL_17798257, EPI_ISL_17798258, EPI_ISL_17798259, EPI_ISL_17798260, EPI_ISL_17798261, EPI_ISL_17798262, EPI_ISL_17798263, EPI_ISL_17798264, EPI_ISL_17798265, EPI_ISL_17798266, EPI_ISL_17798267, EPI_ISL_17798268, EPI_ISL_17798269, EPI_ISL_17798270, EPI_ISL_17798271, EPI_ISL_17798272, EPI_ISL_17798273, EPI_ISL_17798274, EPI_ISL_17798275, EPI_ISL_17798276, EPI_ISL_17798277, EPI_ISL_17798278, EPI_ISL_17798279, EPI_ISL_17798280, EPI_ISL_17798281, EPI_ISL_17798282, EPI_ISL_17798283, EPI_ISL_17798284, EPI_ISL_17798285, EPI_ISL_17798286, EPI_ISL_17798287, EPI_ISL_17798288, EPI_ISL_17798289, EPI_ISL_17798290, EPI_ISL_17798291, EPI_ISL_17798292, EPI_ISL_17798293, EPI_ISL_17798294, EPI_ISL_17798295, EPI_ISL_17798296, EPI_ISL_17798297, EPI_ISL_17798298, EPI_ISL_17798299, EPI_ISL_17798300, EPI_ISL_17798301, EPI_ISL_17798302, EPI_ISL_17798303, EPI_ISL_17798304, EPI_ISL_17798305, EPI_ISL_17798306, EPI_ISL_17798307, EPI_ISL_17798308, EPI_ISL_17798309, EPI_ISL_17798310, EPI_ISL_17798311, EPI_ISL_17798312, EPI_ISL_17798313, EPI_ISL_17798314, EPI_ISL_17798315, EPI_ISL_17798316, EPI_ISL_17798317, EPI_ISL_17798318, EPI_ISL_17798319, EPI_ISL_17798320, EPI_ISL_17798321, EPI_ISL_17798322, EPI_ISL_17798323, EPI_ISL_17798324, EPI_ISL_17798325, EPI_ISL_17798326, EPI_ISL_17798327, EPI_ISL_17798328, EPI_ISL_17798329, EPI_ISL_17798330, EPI_ISL_17798331, EPI_ISL_17798332, EPI_ISL_17798333, EPI_ISL_17798334, EPI_ISL_17798335, EPI_ISL_17798336, EPI_ISL_17798337, EPI_ISL_17798338, EPI_ISL_17798339, EPI_ISL_17798340, EPI_ISL_17798341, EPI_ISL_17798342, EPI_ISL_17798343, EPI_ISL_17798344, EPI_ISL_17798345, EPI_ISL_17798346, EPI_ISL_17798347, EPI_ISL_17798348, EPI_ISL_17798349, EPI_ISL_17798350, EPI_ISL_17798351, EPI_ISL_17798352, EPI_ISL_17798353, EPI_ISL_17798354, EPI_ISL_17798355, EPI_ISL_17798356, EPI_ISL_17798357, EPI_ISL_17798358, EPI_ISL_17798359, EPI_ISL_17798360, EPI_ISL_17798361, EPI_ISL_17798362, EPI_ISL_17798363, EPI_ISL_17798364, EPI_ISL_17798365, EPI_ISL_17798366, EPI_ISL_17798414, EPI_ISL_17798435, EPI_ISL_17798436, EPI_ISL_17798437, EPI_ISL_17798438, EPI_ISL_17798439, EPI_ISL_17798440, EPI_ISL_17798441, EPI_ISL_17798442, EPI_ISL_17798443, EPI_ISL_17798444, EPI_ISL_17798445, EPI_ISL_17798446, EPI_ISL_17798447, EPI_ISL_17798448, EPI_ISL_17798449, EPI_ISL_17798450, EPI_ISL_17798451, EPI_ISL_17798452, EPI_ISL_17798453, EPI_ISL_17798454, EPI_ISL_17798455, EPI_ISL_17798456, EPI_ISL_17798457, EPI_ISL_17798458, EPI_ISL_17798459, EPI_ISL_17798460, EPI_ISL_17798461, EPI_ISL_17798462, EPI_ISL_17798463, EPI_ISL_17798464, EPI_ISL_17798465, EPI_ISL_17798466, EPI_ISL_17798467, EPI_ISL_17798468, EPI_ISL_17798469, EPI_ISL_17798470, EPI_ISL_17798471, EPI_ISL_17798472, EPI_ISL_17798473, EPI_ISL_17798474, EPI_ISL_17798475, EPI_ISL_17798476, EPI_ISL_17798477, EPI_ISL_17798478, EPI_ISL_17798479, EPI_ISL_17798480, EPI_ISL_17798481, EPI_ISL_17798482, EPI_ISL_17798483, EPI_ISL_17798484, EPI_ISL_17798485, EPI_ISL_17798486, EPI_ISL_17798487, EPI_ISL_17798488, EPI_ISL_17798489, EPI_ISL_17798490, EPI_ISL_17798491, EPI_ISL_17798492, EPI_ISL_17798493, EPI_ISL_17798494, EPI_ISL_17798495, EPI_ISL_17798496, EPI_ISL_17798497, EPI_ISL_17798498, EPI_ISL_17798499, EPI_ISL_17798500, EPI_ISL_17798501, EPI_ISL_17798502, EPI_ISL_17798503, EPI_ISL_17798504, EPI_ISL_17798505, EPI_ISL_17798506, EPI_ISL_17798507, EPI_ISL_17798508, EPI_ISL_17798509, EPI_ISL_17798510, EPI_ISL_17798511, EPI_ISL_17798512, EPI_ISL_17798513, EPI_ISL_17798514, EPI_ISL_17798515, EPI_ISL_17798516, EPI_ISL_17798517, EPI_ISL_17798518, EPI_ISL_17798519, EPI_ISL_17798520, EPI_ISL_17798521, EPI_ISL_17798522, EPI_ISL_17798523, EPI_ISL_17798524, EPI_ISL_17798525, EPI_ISL_17798526, EPI_ISL_17798527, EPI_ISL_17798528, EPI_ISL_17798529, EPI_ISL_17798530, EPI_ISL_17798531, EPI_ISL_17798532, EPI_ISL_17798533, EPI_ISL_17798534, EPI_ISL_17798535, EPI_ISL_17798536, EPI_ISL_17798537, EPI_ISL_17798538, EPI_ISL_17798539, EPI_ISL_17798540, EPI_ISL_17798541, EPI_ISL_17798542, EPI_ISL_17798543, EPI_ISL_17798544, EPI_ISL_17798545, EPI_ISL_17798546, EPI_ISL_17798547, EPI_ISL_17798548, EPI_ISL_17798549, EPI_ISL_17798550, EPI_ISL_17798551, EPI_ISL_17798552, EPI_ISL_17798553, EPI_ISL_17798554, EPI_ISL_17798555, EPI_ISL_17798556, EPI_ISL_17798557, EPI_ISL_17798558, EPI_ISL_17798559, EPI_ISL_17798560, EPI_ISL_17798561, EPI_ISL_17798562, EPI_ISL_17798563, EPI_ISL_17798564, EPI_ISL_17798565, EPI_ISL_17798566, EPI_ISL_17798567, EPI_ISL_17798568, EPI_ISL_17798569, EPI_ISL_17798570, EPI_ISL_17798571, EPI_ISL_17798572, EPI_ISL_17798573, EPI_ISL_17798574, EPI_ISL_17798575, EPI_ISL_17798576, EPI_ISL_17798577, EPI_ISL_17798578, EPI_ISL_17798579, EPI_ISL_17798580, EPI_ISL_17798581, EPI_ISL_17798582, EPI_ISL_17798583, EPI_ISL_17798584, EPI_ISL_17798585, EPI_ISL_17798586, EPI_ISL_17798587, EPI_ISL_17798588, EPI_ISL_17798589, EPI_ISL_17798590, EPI_ISL_17798591 | | | |
| see above | Environmental Health Institute, National Environment Agency | Environmental Health Institute, National Environment Agency | Sy,A.K., Koo,C., Privaldos,K.J.R., Quinones,M.A.T., Igoy,M.A.U., Villanueva,S.Y.A.M., Hibberd,M.L., Ng,L.C. and Hapuarachchi,H.C. |
| EPI_ISL_17798592, EPI_ISL_17798593, EPI_ISL_17798594, EPI_ISL_17798595, EPI_ISL_17798596, EPI_ISL_17798597, EPI_ISL_17798598, EPI_ISL_17798599, EPI_ISL_17798600, EPI_ISL_17798601, EPI_ISL_17798602, EPI_ISL_17798603, EPI_ISL_17798604, EPI_ISL_17798605, EPI_ISL_17798606, EPI_ISL_17798607, EPI_ISL_17798608, EPI_ISL_17798609, EPI_ISL_17798610, EPI_ISL_17798611, EPI_ISL_17798612, EPI_ISL_17798613, EPI_ISL_17798614, EPI_ISL_17798615, EPI_ISL_17798616, EPI_ISL_17798617, EPI_ISL_17798618, EPI_ISL_17798619, EPI_ISL_17798620, EPI_ISL_17798621, EPI_ISL_17798622, EPI_ISL_17798623, EPI_ISL_17798624, EPI_ISL_17798625, EPI_ISL_17798626, EPI_ISL_17798627, EPI_ISL_17798628, EPI_ISL_17798629, EPI_ISL_17798630, EPI_ISL_17798631, EPI_ISL_17798632, EPI_ISL_17798633, EPI_ISL_17798634, EPI_ISL_17798635, EPI_ISL_17798636, EPI_ISL_17798637, EPI_ISL_17798638, EPI_ISL_17798639, EPI_ISL_17798640, EPI_ISL_17798641, EPI_ISL_17798642, EPI_ISL_17798643, EPI_ISL_17798644, EPI_ISL_17798645, EPI_ISL_17798646, EPI_ISL_17798647, EPI_ISL_17798648, EPI_ISL_17798649, EPI_ISL_17798650, EPI_ISL_17798651, EPI_ISL_17798652, EPI_ISL_17798653, EPI_ISL_17798654, EPI_ISL_17798655, EPI_ISL_17798656, EPI_ISL_17798657, EPI_ISL_17798658, EPI_ISL_17798659, EPI_ISL_17798660, EPI_ISL_17798661, EPI_ISL_17798662, EPI_ISL_17798663, EPI_ISL_17798664, EPI_ISL_17798665, EPI_ISL_17798666, EPI_ISL_17798667, EPI_ISL_17798668, EPI_ISL_17798669, EPI_ISL_17798670, EPI_ISL_17798671, EPI_ISL_17798672, EPI_ISL_17798673, EPI_ISL_17798674, EPI_ISL_17798675, EPI_ISL_17798676, EPI_ISL_17798677, EPI_ISL_17798678, EPI_ISL_17798679, EPI_ISL_17798680, EPI_ISL_17798681, EPI_ISL_17798682, EPI_ISL_17798683, EPI_ISL_17798684, EPI_ISL_17798685, EPI_ISL_17798686, EPI_ISL_17798687, EPI_ISL_17798688, EPI_ISL_17798689, EPI_ISL_17798690, EPI_ISL_17798691, EPI_ISL_17798692, EPI_ISL_17798693, EPI_ISL_17798694, EPI_ISL_17798695, EPI_ISL_17798696, EPI_ISL_17798697, EPI_ISL_17798698, EPI_ISL_17798699, EPI_ISL_17798700, EPI_ISL_17798701, EPI_ISL_17798702, EPI_ISL_17798703, EPI_ISL_17798704, EPI_ISL_17798705, EPI_ISL_17798706, EPI_ISL_17798707, EPI_ISL_17798708, EPI_ISL_17798709, EPI_ISL_17798710, EPI_ISL_17798711, EPI_ISL_17798712, EPI_ISL_17798713, EPI_ISL_17798714, EPI_ISL_17798715, EPI_ISL_17798716, EPI_ISL_17798717, EPI_ISL_17798718, EPI_ISL_17798719, EPI_ISL_17798720, EPI_ISL_17798721, EPI_ISL_17798722, EPI_ISL_17798723, EPI_ISL_17798724, EPI_ISL_17798725, EPI_ISL_17798726, EPI_ISL_17798727, EPI_ISL_17798728, EPI_ISL_17798729, EPI_ISL_17798730, EPI_ISL_17798731, EPI_ISL_17798732, EPI_ISL_17798733, EPI_ISL_17798734, EPI_ISL_17798735, EPI_ISL_17798736, EPI_ISL_17798737, EPI_ISL_17798738, EPI_ISL_17798739, EPI_ISL_17798740, EPI_ISL_17798741, EPI_ISL_17798742, EPI_ISL_17798743, EPI_ISL_17798744, EPI_ISL_17798745, EPI_ISL_17798746, EPI_ISL_17798747, EPI_ISL_17798748, EPI_ISL_17798749, EPI_ISL_17798750, EPI_ISL_17798751, EPI_ISL_17798752, EPI_ISL_17798753, EPI_ISL_17798754, EPI_ISL_17798755, EPI_ISL_17798756, EPI_ISL_17798757, EPI_ISL_17798758, EPI_ISL_17798759, EPI_ISL_17798760, EPI_ISL_17798761, EPI_ISL_17798762, EPI_ISL_17798763, EPI_ISL_17798764, EPI_ISL_17798765, EPI_ISL_17798766, EPI_ISL_17798767, EPI_ISL_17798768, EPI_ISL_17798769, EPI_ISL_17798770 | | | |
| see above | Evercare Hospital Dhaka (Ex Apollo Hospitals Dhaka) | Research Institute for Microbial Diseases, Osaka University, Department of Viral Infections | Rahim,R., Hasan,A., Phadungsombat,J., Hasan,N., Ara,N., Biswas,S.M., Nakayama,E.E., Rahman,M. and Shioda,T. |
| EPI_ISL_17798771, EPI_ISL_17798772, EPI_ISL_17798773, EPI_ISL_17798774, EPI_ISL_17798775, EPI_ISL_17798776, EPI_ISL_17798777, EPI_ISL_17798778, EPI_ISL_17798779, EPI_ISL_17798780, EPI_ISL_17798781, EPI_ISL_17798782, EPI_ISL_17798783, EPI_ISL_17798784, EPI_ISL_17798785, EPI_ISL_17798786, EPI_ISL_17798787, EPI_ISL_17798788, EPI_ISL_17798789, EPI_ISL_17798790, EPI_ISL_17798791, EPI_ISL_17798792, EPI_ISL_17798793, EPI_ISL_17798794, EPI_ISL_17798795 | | | |
| see above | Sun Yat-Sen University, Sun Yat-Sen Memorial Hospital, Guangdong Provincial Key Laboratory of Malignant Tumor Epigenetics and Gene Regulation | Sun Yat-Sen University, Sun Yat-Sen Memorial Hospital, Guangdong Provincial Key Laboratory of Malignant Tumor Epigenetics and Gene Regulation | Meng,J.X., Hu,Q.M., Zhang,L.M., Li,N., He,Y.W., Yang,Z.X., Sun,Y. and Wang,J.L. |
| EPI_ISL_17804503, EPI_ISL_17804504, EPI_ISL_17804505, EPI_ISL_17804506, EPI_ISL_17804507, EPI_ISL_17804508, EPI_ISL_17804509, EPI_ISL_17804510, EPI_ISL_17804511, EPI_ISL_17804512, EPI_ISL_17804513, EPI_ISL_17804514, EPI_ISL_17804515, EPI_ISL_17804516, EPI_ISL_17804517, EPI_ISL_17804518, EPI_ISL_17804519, EPI_ISL_17804520, EPI_ISL_17804521, EPI_ISL_17804522, EPI_ISL_17804523, EPI_ISL_17804524, EPI_ISL_17804525, EPI_ISL_17804526, EPI_ISL_17804527, EPI_ISL_17804528, EPI_ISL_17804529, EPI_ISL_17804530, EPI_ISL_17804531, EPI_ISL_17804532, EPI_ISL_17804533, EPI_ISL_17804534 | | | |
| see above | California Academy of Sciences | California Academy of Sciences | Thongrsipong,P., Edgerton,S.V., Bos,S., Saborio,S., Kuan,G., Balmaseda,A., Harris,E. and Bennett,S.N. |
| EPI_ISL_17831892 | State Center for Health Surveillance/Secretary of State for Health | State Center for Health Surveillance/Secretary of State for Health | Salvato,R.S., Wallau,G.L. and Graf,T. |
| EPI_ISL_17831894, EPI_ISL_17831895 | Research Medical Center of Lambarene | Research Medical Center of Lambarene | Bikangui,R. |
| EPI_ISL_17831896 | Blood Center of Ribeirao Preto, University of Sao Paulo | Blood Center of Ribeirao Preto, University of Sao Paulo | La-Roque,D.G.L., Santos,E.V., Oliveira,R.A.M., Slavov,S.N., Rodrigues,E.S., Fonseca,V., Martins,A.J., Giomo,D.B., Torres,P.M.A., Gentil,D.C.D., Catoia,E.A., Chiquito,N.C., Medeiros,A.P.S.S., Yamamoto,A.Y., Passos,L.M.R., Calado,R.T., Elias,M.C., Sampaio,S.C., Giovanetti,M., Alcantara,L.C., Covas,D.T. and Kashima,S. |
| EPI_ISL_17831897, EPI_ISL_17831898, EPI_ISL_17831899, EPI_ISL_17831900 | Department of Virology and Immunity, Guangzhou Center for Disease Control and Prevention | Department of Virology and Immunity, Guangzhou Center for Disease Control and Prevention | Su,W., Lu,W. and Cao,Y. |
| EPI_ISL_17831901, EPI_ISL_17831902, EPI_ISL_17831903, EPI_ISL_17831904, EPI_ISL_17831905, EPI_ISL_17831906, EPI_ISL_17831907, EPI_ISL_17831908, EPI_ISL_17831909, EPI_ISL_17831910, EPI_ISL_17831911, EPI_ISL_17831912, EPI_ISL_17831913, EPI_ISL_17831914, EPI_ISL_17831915, EPI_ISL_17831916, EPI_ISL_17831917, EPI_ISL_17831918, EPI_ISL_17831919, EPI_ISL_17831920, EPI_ISL_17831921, EPI_ISL_17831922, EPI_ISL_17831923, EPI_ISL_17831924, EPI_ISL_17831925, EPI_ISL_17831926, EPI_ISL_17831927, EPI_ISL_17831928, EPI_ISL_17831929, EPI_ISL_17831930, EPI_ISL_17831931, EPI_ISL_17831932, EPI_ISL_17831933, EPI_ISL_17831934, EPI_ISL_17831935, EPI_ISL_17831936 | | | |
| see above | National Hospital for Tropical Disease (NHTD) | Research Institute for Microbial Diseases, Osaka University, Department of Viral Infections | Phadungsombat,J., Vu,H.T.T., Nguyen,Q.T., Nguyen,H.T.V., Nguyen,H.T.N., Dang,B.T., Nakayama,E.E., Ishizaki,A., Ichimura,H., Shioda,T. and Pham,T.N. |
| EPI_ISL_17848715, EPI_ISL_17848716 | Department of Microbiology, Zhongshan School of Medicine, Sun Yat-Sen University, Key Laboratory of Tropical Disease Control, Ministry of Education | Department of Microbiology, Zhongshan School of Medicine, Sun Yat-Sen University, Key Laboratory of Tropical Disease Control, Ministry of Education | Li,Y. |
| EPI_ISL_17848717 | Department of Microbiology, Zhongshan School of Medicine, Sun Yat-Sen University, Key Laboratory of Tropical Disease Control, Ministry of Education | Department of Microbiology, Zhongshan School of Medicine, Sun Yat-Sen University, Key Laboratory of Tropical Disease Control, Ministry of Education | Li,Y.-P., Hu,M., Wu,D. and Wu,T. |
| EPI_ISL_17859404, EPI_ISL_17859405, EPI_ISL_17859406, EPI_ISL_17859407, EPI_ISL_17859408, EPI_ISL_17859409, EPI_ISL_17859410, EPI_ISL_17859411, EPI_ISL_17859412, EPI_ISL_17859413, EPI_ISL_17859414, EPI_ISL_17859415, EPI_ISL_17859416, EPI_ISL_17859417, EPI_ISL_17859418, EPI_ISL_17859419, EPI_ISL_17859420, EPI_ISL_17859421, EPI_ISL_17859422, EPI_ISL_17859423, EPI_ISL_17859424, EPI_ISL_17859425, EPI_ISL_17859426, EPI_ISL_17859427, EPI_ISL_17859428, EPI_ISL_17859429, EPI_ISL_17859430, EPI_ISL_17859431, EPI_ISL_17859432, EPI_ISL_17859433, EPI_ISL_17859434, EPI_ISL_17859435, EPI_ISL_17859436, EPI_ISL_17859437, EPI_ISL_17859438, EPI_ISL_17859439, EPI_ISL_17859440, EPI_ISL_17859441, EPI_ISL_17859442 | | | |
| see above | Coordenacao Geral da Laboratorio em Saude Publica | Coordenacao Geral da Laboratorio em Saude Publica | Ribeiro,G., Parise,D., Paiva,A., Franz,H., Junior,W. and Arcanjo,A.R. |
| EPI_ISL_17859443 | Bureau of Public Health Laboratories, Florida Department of Health | Bureau of Public Health Laboratories, Florida Department of Health | Schmedes,S., Tekin,O., Tarigopula,N. and Dong,Y. |
| EPI_ISL_17859444, EPI_ISL_17859445, EPI_ISL_17859446, EPI_ISL_17859447, EPI_ISL_17859448 | Bureau of Public Health Laboratories, Florida Department of Health | Bureau of Public Health Laboratories, Florida Department of Health | Schmedes,S., Tarigopula,N., Tekin,O. and Dong,Y. |
| EPI_ISL_17859449 | Bureau of Public Health Laboratories, Florida Department of Health | Bureau of Public Health Laboratories, Florida Department of Health | Schmedes,S., Tekin,O., Tarigopula,N. and Dong,Y. |
| EPI_ISL_17859450, EPI_ISL_17859451 | Bureau of Public Health Laboratories, Florida Department of Health | Bureau of Public Health Laboratories, Florida Department of Health | Schmedes,S., Tarigopula,N., Tekin,O. and Dong,Y. |
| EPI_ISL_17859452 | Bureau of Public Health Laboratories, Florida Department of Health | Bureau of Public Health Laboratories, Florida Department of Health | Schmedes,S., Tekin,O., Tarigopula,N. and Dong,Y. |
| EPI_ISL_17859453, EPI_ISL_17859454 | Bureau of Public Health Laboratories, Florida Department of Health | Bureau of Public Health Laboratories, Florida Department of Health | Schmedes,S., Tarigopula,N., Tekin,O. and Dong,Y. |
| EPI_ISL_17859455, EPI_ISL_17859456 | Bureau of Public Health Laboratories, Florida Department of Health | Bureau of Public Health Laboratories, Florida Department of Health | Schmedes,S., Tekin,O., Tarigopula,N. and Dong,Y. |
| EPI_ISL_17859457 | Bureau of Public Health Laboratories, Florida Department of Health | Bureau of Public Health Laboratories, Florida Department of Health | Schmedes,S., Tarigopula,N., Tekin,O. and Dong,Y. |
| EPI_ISL_17859458, EPI_ISL_17859459, EPI_ISL_17859460 | Bureau of Public Health Laboratories, Florida Department of Health | Bureau of Public Health Laboratories, Florida Department of Health | Schmedes,S., Tekin,O., Tarigopula,N. and Dong,Y. |
| EPI_ISL_17859461, EPI_ISL_17859462 | Bureau of Public Health Laboratories, Florida Department of Health | Bureau of Public Health Laboratories, Florida Department of Health | Schmedes,S., Tarigopula,N., Tekin,O. and Dong,Y. |
| EPI_ISL_17859463 | Bureau of Public Health Laboratories, Florida Department of Health | Bureau of Public Health Laboratories, Florida Department of Health | Schmedes,S., Tekin,O., Tarigopula,N. and Dong,Y. |
| EPI_ISL_17884732, EPI_ISL_17884733, EPI_ISL_17884734, EPI_ISL_17884735, EPI_ISL_17884736, EPI_ISL_17884737, EPI_ISL_17884738, EPI_ISL_17884739, EPI_ISL_17884740, EPI_ISL_17884741, EPI_ISL_17884742, EPI_ISL_17884743, EPI_ISL_17884744, EPI_ISL_17884745, EPI_ISL_17884746, EPI_ISL_17884747, EPI_ISL_17884748, EPI_ISL_17884749, EPI_ISL_17884750, EPI_ISL_17884751, EPI_ISL_17884752, EPI_ISL_17884753, EPI_ISL_17884754, EPI_ISL_17884755, EPI_ISL_17884756, EPI_ISL_17884757, EPI_ISL_17884758, EPI_ISL_17884759, EPI_ISL_17884760, EPI_ISL_17884761, EPI_ISL_17884762, EPI_ISL_17884763, EPI_ISL_17884764, EPI_ISL_17884765, EPI_ISL_17884766, EPI_ISL_17884767, EPI_ISL_17884768, EPI_ISL_17884769, EPI_ISL_17884770, EPI_ISL_17884771, EPI_ISL_17884772, EPI_ISL_17884773, EPI_ISL_17884774, EPI_ISL_17884775, EPI_ISL_17884776, EPI_ISL_17884777, EPI_ISL_17884778, EPI_ISL_17884779, EPI_ISL_17884780, EPI_ISL_17884781, EPI_ISL_17884782, EPI_ISL_17884783, EPI_ISL_17884784, EPI_ISL_17884785, EPI_ISL_17884786, EPI_ISL_17884787, EPI_ISL_17884788, EPI_ISL_17884789, EPI_ISL_17884790, EPI_ISL_17884791, EPI_ISL_17884792, EPI_ISL_17884793, EPI_ISL_17884794, EPI_ISL_17884795, EPI_ISL_17884796, EPI_ISL_17884797, EPI_ISL_17884798, EPI_ISL_17884799, EPI_ISL_17884800, EPI_ISL_17884801, EPI_ISL_17884802, EPI_ISL_17884803, EPI_ISL_17884804, EPI_ISL_17884805, EPI_ISL_17884806, EPI_ISL_17884807, EPI_ISL_17884808, EPI_ISL_17884809, EPI_ISL_17884810, EPI_ISL_17884811, EPI_ISL_17884812, EPI_ISL_17884813, EPI_ISL_17884814, EPI_ISL_17884815, EPI_ISL_17884816, EPI_ISL_17884817, EPI_ISL_17884818, EPI_ISL_17884819, EPI_ISL_17884820, EPI_ISL_17884821, EPI_ISL_17884822, EPI_ISL_17884823, EPI_ISL_17884824, EPI_ISL_17884825, EPI_ISL_17884826, EPI_ISL_17884827, EPI_ISL_17884828, EPI_ISL_17884829, EPI_ISL_17884830, EPI_ISL_17884831, EPI_ISL_17884832, EPI_ISL_17884833, EPI_ISL_17884834, EPI_ISL_17884835, EPI_ISL_17884836, EPI_ISL_17884837, EPI_ISL_17884838, EPI_ISL_17884839, EPI_ISL_17884840, EPI_ISL_17884841, EPI_ISL_17884842, EPI_ISL_17884843, EPI_ISL_17884844, EPI_ISL_17884845, EPI_ISL_17884846, EPI_ISL_17884847, EPI_ISL_17884848, EPI_ISL_17884849 | | | |
| see above | Central Public Health Laboratory Dr. Giovanni | Central Public Health Laboratory Dr. Giovanni | Mendonca,A.F., Andrade,C.F., Sobreiro,M.B., Almeida,R.M., Silva,V.L. and Pereira,L.A. |

| | | | |
|---|---|---|---|
| | Cysneiros (LACEN-GO) | Cysneiros (LACEN-GO) | |
| EPI_ISL_17884850, EPI_ISL_17884851, EPI_ISL_17884852, EPI_ISL_17884853, EPI_ISL_17884854, EPI_ISL_17884855, EPI_ISL_17884856, EPI_ISL_17884857, EPI_ISL_17884858, EPI_ISL_17884859, EPI_ISL_17884860, EPI_ISL_17884861, EPI_ISL_17884862, EPI_ISL_17884863, EPI_ISL_17884864, EPI_ISL_17884865, EPI_ISL_17884866, EPI_ISL_17884867, EPI_ISL_17884868, EPI_ISL_17884869, EPI_ISL_17884870, EPI_ISL_17884871 | | | |
| see above | Coordenacao Geral da Laboratorio em Saude Publica | Coordenacao Geral da Laboratorio em Saude Publica | Ribeiro,G., Parise,D., Paiva,A., Franz,H., Junior,W. and Arcanjo,A.R. |
| EPI_ISL_17884872 | Virus Laboratory, Indian Council of Medical Research, National Institute of Cholera and Enteric Diseases | Virus Laboratory, Indian Council of Medical Research, National Institute of Cholera and Enteric Diseases | Baskey,U., Verma,P., Ghosh,A. and Sadhukhan,P.C. |
| EPI_ISL_17959701, EPI_ISL_17959702, EPI_ISL_17959703, EPI_ISL_17959704, EPI_ISL_17959705, EPI_ISL_17959706, EPI_ISL_17959707, EPI_ISL_17959708, EPI_ISL_17959709, EPI_ISL_17959710, EPI_ISL_17959711, EPI_ISL_17959712, EPI_ISL_17959713, EPI_ISL_17959714, EPI_ISL_17959715, EPI_ISL_17959716, EPI_ISL_17959718, EPI_ISL_17959719, EPI_ISL_17959720, EPI_ISL_17959721, EPI_ISL_17959722, EPI_ISL_17959723, EPI_ISL_17959724, EPI_ISL_17959725, EPI_ISL_17959726 | | | |
| see above | Institute of Microbiology, Universidad San Francisco de Quito | Institute of Microbiology, Universidad San Francisco de Quito | Sully Márquez, Gwenyth Lee, Bernardo Gutiérrez, Joseph Eisenberg, Gabriel Trueba, Josefina Coloma |
| EPI_ISL_17975657, EPI_ISL_17975658, EPI_ISL_17975659, EPI_ISL_17975660, EPI_ISL_17975661, EPI_ISL_17975662, EPI_ISL_17975663, EPI_ISL_17975664, EPI_ISL_17975665, EPI_ISL_17975666, EPI_ISL_17975667, EPI_ISL_17975668, EPI_ISL_17975669, EPI_ISL_17975670, EPI_ISL_17975671, EPI_ISL_17975672, EPI_ISL_17975673, EPI_ISL_17975674, EPI_ISL_17975675, EPI_ISL_17975676, EPI_ISL_17975677, EPI_ISL_17975678, EPI_ISL_17975679, EPI_ISL_17975680, EPI_ISL_17975681 | | | |
| see above | Gandhi Medical College | Gandhi Medical College | Agarwal,A., Ganvir,R., Kale,D., Chaurasia,D. and Kapoor,G. |
| EPI_ISL_17975682, EPI_ISL_17975683, EPI_ISL_17975684, EPI_ISL_17975685, EPI_ISL_17975686, EPI_ISL_17975687, EPI_ISL_17975688, EPI_ISL_17975689, EPI_ISL_17975690, EPI_ISL_17975691, EPI_ISL_17975692, EPI_ISL_17975693, EPI_ISL_17975694, EPI_ISL_17975695, EPI_ISL_17975696, EPI_ISL_17975697, EPI_ISL_17975698, EPI_ISL_17975699 | | | |
| see above | Armauer Hansen Research Institute | Armauer Hansen Research Institute | Mengesha,M., Tayachew,A., Tsega,D., Hailu,D., Abeje,D., Seyoum,T., Adnew,B., Konde,A., Biru,M., Agunie,A., Asrat,Y., Wossen,M., Gelanew,T., Piantadosi,A., Mihret,A. and Mulu,A. |
| EPI_ISL_17976304, EPI_ISL_17977444 | Instituto de Investigaciones Biologicas del Tropico, Universidad de Córdoba | Instituto de Investigaciones Biologicas del Tropico, Universidad de Córdoba | Ricardo Rivero, Daniela Paternina, Vaneza Tique, Salim Mattar |
| EPI_ISL_17980694, EPI_ISL_17980695, EPI_ISL_17980696, EPI_ISL_17980697, EPI_ISL_17980698, EPI_ISL_17980699, EPI_ISL_17980700, EPI_ISL_17980701, EPI_ISL_17980702, EPI_ISL_17980703, EPI_ISL_17980704, EPI_ISL_17980705 | | | |
| see above | Department of Microbiology | Department of Microbiology | Okada,W., Kasuya,F., Harada,S., Kumagai,R., Amano,A., Hasegawa,M., Miyake,H., Nagashima,M. and Sadamasu,K. |
| EPI_ISL_17980809, EPI_ISL_17980810 | Hospital Maternidade Sao Jose (Guaraciaba Do Norte) | Laboratório Central de Saúde Pública do Ceará | Tayna da Silva Fiuza, Vânia Angélica Feitosa Viana, Shirlene Telmos Silva de Lima |
| EPI_ISL_17980811 | Laboratorio De Patologia Clinica De Morada Nova | Laboratório Central de Saúde Pública do Ceará | Tayna da Silva Fiuza, Vânia Angélica Feitosa Viana, Shirlene Telmos Silva de Lima |
| EPI_ISL_17980812 | Hospital Municipal Jaime Osterno (Marco) | Laboratório Central de Saúde Pública do Ceará | Tayna da Silva Fiuza, Vânia Angélica Feitosa Viana, Shirlene Telmos Silva de Lima |
| EPI_ISL_17980813 | Centro De Saude De Bela Cruzfns(Bela Cruz) | Laboratório Central de Saúde Pública do Ceará | Tayna da Silva Fiuza, Vânia Angélica Feitosa Viana, Shirlene Telmos Silva de Lima |
| EPI_ISL_17980814 | Hospital Municipal De Jaguaribe | Laboratório Central de Saúde Pública do Ceará | Tayna da Silva Fiuza, Vânia Angélica Feitosa Viana, Shirlene Telmos Silva de Lima |
| EPI_ISL_17980815, EPI_ISL_17980816, EPI_ISL_17980817, EPI_ISL_17980818, EPI_ISL_17980819, EPI_ISL_17980820, EPI_ISL_17980821, EPI_ISL_17980822, EPI_ISL_17980823, EPI_ISL_17980824, EPI_ISL_17980825, EPI_ISL_17980826, EPI_ISL_17980827 | | | |
| see above | Centro De Saude De Bela Cruzfns(Bela Cruz) | Laboratório Central de Saúde Pública do Ceará | Tayna da Silva Fiuza, Vânia Angélica Feitosa Viana, Shirlene Telmos Silva de Lima |
| EPI_ISL_17980828 | Lab De Analises Clinicas De Cruz | Laboratório Central de Saúde Pública do Ceará | Tayna da Silva Fiuza, Vânia Angélica Feitosa Viana, Shirlene Telmos Silva de Lima |
| EPI_ISL_17980829 | Hosp Munic Dr Deoclecio Lima Verde (Limoeiro) | Laboratório Central de Saúde Pública do Ceará | Tayna da Silva Fiuza, Vânia Angélica Feitosa Viana, Shirlene Telmos Silva de Lima |
| EPI_ISL_17980830 | Centro De Saude De Bela Cruzfns(Bela Cruz) | Laboratório Central de Saúde Pública do Ceará | Tayna da Silva Fiuza, Vânia Angélica Feitosa Viana, Shirlene Telmos Silva de Lima |
| EPI_ISL_17980831, EPI_ISL_17980832 | Hospital Municipal De Jaguaribe | Laboratório Central de Saúde Pública do Ceará | Tayna da Silva Fiuza, Vânia Angélica Feitosa Viana, Shirlene Telmos Silva de Lima |
| EPI_ISL_17980833 | Centro De Saude De Bela Cruzfns(Bela Cruz) | Laboratório Central de Saúde Pública do Ceará | Tayna da Silva Fiuza, Vânia Angélica Feitosa Viana, Shirlene Telmos Silva de Lima |
| EPI_ISL_17980834, EPI_ISL_17980835, EPI_ISL_17980836, EPI_ISL_17980837 | Hospital Municipal De Jaguaribe | Laboratório Central de Saúde Pública do Ceará | Tayna da Silva Fiuza, Vânia Angélica Feitosa Viana, Shirlene Telmos Silva de Lima |
| EPI_ISL_17980838, EPI_ISL_17980839, EPI_ISL_17980840, EPI_ISL_17980841, EPI_ISL_17980842, EPI_ISL_17980843, EPI_ISL_17980844, EPI_ISL_17980845, EPI_ISL_17980846, EPI_ISL_17980847, EPI_ISL_17980848 | | | |
| see above | Centro De Saude De Bela Cruzfns(Bela Cruz) | Laboratório Central de Saúde Pública do Ceará | Tayna da Silva Fiuza, Vânia Angélica Feitosa Viana, Shirlene Telmos Silva de Lima |
| EPI_ISL_17980849 | Hospital Infantil Albert Sabin (Fortaleza) | Laboratório Central de Saúde Pública do Ceará | Tayna da Silva Fiuza, Vânia Angélica Feitosa Viana, Shirlene Telmos Silva de Lima |
| EPI_ISL_17983067 | LACEN de Tocantins | Instituto Adolfo Lutz - Strategic Laboratory | Claudio Tavares Sacchi, Karoline Rodrigues Campos, Marlon Benedito Nascimento Santos, Franciano Dias Pereira Cardoso |
| EPI_ISL_17983068, EPI_ISL_17983069, EPI_ISL_17983070, EPI_ISL_17983071 | LACEN de Sergipe | Instituto Adolfo Lutz - Strategic Laboratory | Claudio Tavares Sacchi, Karoline Rodrigues Campos, Marlon Benedito Nascimento Santos, Gabriela Vasconcelos Brito Bezerra |
| EPI_ISL_17983072, EPI_ISL_17983073, EPI_ISL_17983074, EPI_ISL_17983075, EPI_ISL_17983076, EPI_ISL_17983077, EPI_ISL_17983078, EPI_ISL_17983079, EPI_ISL_17983080, EPI_ISL_17983081, EPI_ISL_17983082, EPI_ISL_17983083, EPI_ISL_17983084, EPI_ISL_17983085, EPI_ISL_17983086, EPI_ISL_17983087 | | | |
| see above | Instituto Adolfo Lutz | Instituto Adolfo Lutz - Strategic Laboratory | Claudio Tavares Sacchi, Karoline Rodrigues Campos, Marlon Benedito Nascimento Santos, Juliana Silva Nogueira |
| EPI_ISL_17983088, EPI_ISL_17983089, EPI_ISL_17983090, EPI_ISL_17983091 | LACEN de Sergipe | Instituto Adolfo Lutz - Strategic Laboratory | Claudio Tavares Sacchi, Karoline Rodrigues Campos, Marlon Benedito Nascimento Santos, Gabriela Vasconcelos Brito Bezerra |
| EPI_ISL_17983092, EPI_ISL_17983093, EPI_ISL_17983094 | LACEN de Tocantins | Instituto Adolfo Lutz - Strategic Laboratory | Claudio Tavares Sacchi, Karoline Rodrigues Campos, Marlon Benedito Nascimento Santos, Franciano Dias Pereira Cardoso |
| EPI_ISL_17983095 | LACEN de Tocantins | Instituto Adolfo Lutz - Strategic Laboratory | Claudio Tavares Sacchi, Karoline Rodrigues Campos, Marlon Benedito Nascimento Santos, Gabriela Vasconcelos Brito Bezerra |
| EPI_ISL_18001716 | Armauer Hansen Research Institute | Armauer Hansen Research Institute | Mengesha,M., Tayachew,A., Tsega,D., Hailu,D., Abeje,D., Seyoum,T., Adnew,B., Konde,A., Biru,M., Agunie,A., Asrat,Y., Wossen,M., Gelanew,T., Piantadosi,A., Mihret,A. and Mulu,A. |
| EPI_ISL_18001717, EPI_ISL_18001718, EPI_ISL_18001719, EPI_ISL_18001720, EPI_ISL_18001721, EPI_ISL_18001722, EPI_ISL_18001723, EPI_ISL_18001724, EPI_ISL_18001725, EPI_ISL_18001726, EPI_ISL_18001727, EPI_ISL_18001728 | | | |
| see above | Department of Microbiology | Department of Microbiology | Okada,W., Kasuya,F., Harada,S., Kumagai,R., Amano,A., Hasegawa,M., Miyake,H., Nagashima,M. and Sadamasu,K. |
| EPI_ISL_18001729, EPI_ISL_18001730, EPI_ISL_18001731, EPI_ISL_18001732, EPI_ISL_18001733, EPI_ISL_18001734, EPI_ISL_18001735, EPI_ISL_18001736, EPI_ISL_18001737, EPI_ISL_18001738, EPI_ISL_18001739, EPI_ISL_18001740, EPI_ISL_18001741, EPI_ISL_18001742, EPI_ISL_18001743, EPI_ISL_18001744, EPI_ISL_18001745 | | | |
| see above | Gandhi Medical College | Gandhi Medical College | Agarwal,A., Ganvir,R., Kale,D., Chaurasia,D. and Kapoor,G. |
| EPI_ISL_18007204, EPI_ISL_18007205, EPI_ISL_18007206, EPI_ISL_18007207, EPI_ISL_18007208, EPI_ISL_18007209, EPI_ISL_18007210, EPI_ISL_18007211, EPI_ISL_18007212 | Centro Nacional de Enfermedades Tropicales | Centro Nacional de Enfermedades Tropicales | Roca,Y., Baronti,C., Revollo,R.J., Cook,S., Loayza,R., Ninove,L., Fernandez,R.T., Flores,J.V., Herve,J.P. and de Lamballerie,X. |
| EPI_ISL_18007213 | Department of Virology and Immunity, Guangzhou Center for Disease Control and Prevention | Department of Virology and Immunity, Guangzhou Center for Disease Control and Prevention | Su,W., Lu,W. and Cao,Y. |
| EPI_ISL_18009962, EPI_ISL_18009963 | LACEN-AL | Laboratório Central de Saúde Pública do Ceará | Tayna da Silva Fiuza, Vânia Angélica Feitosa Viana, Shirlene Telmos Silva de Lima |
| EPI_ISL_18009964 | LACEN-MA | Laboratório Central de Saúde Pública do Ceará | Tayna da Silva Fiuza, Vânia Angélica Feitosa Viana, Shirlene Telmos Silva de Lima on behalf of LACEN-MA |
| EPI_ISL_18009965 | Unidade Mista Dr Edmar Da Costa Barroso (Pacatuba) | Laboratório Central de Saúde Pública do Ceará | Tayna da Silva Fiuza, Vânia Angélica Feitosa Viana, Shirlene Telmos Silva de Lima |
| EPI_ISL_18009966, EPI_ISL_18009967, EPI_ISL_18009968, EPI_ISL_18009969, EPI_ISL_18009970, EPI_ISL_18009971, EPI_ISL_18009972 | Hospital Municipal De Jaguaribe | Laboratório Central de Saúde Pública do Ceará | Tayna da Silva Fiuza, Vânia Angélica Feitosa Viana, Shirlene Telmos Silva de Lima |
| EPI_ISL_18009973 | Lab De Analises Clinicas De Cruz | Laboratório Central de Saúde Pública do Ceará | Tayna da Silva Fiuza, Vânia Angélica Feitosa Viana, Shirlene Telmos Silva de Lima |
| EPI_ISL_18009974 | Hospital Municipal De Jaguaribe | Laboratório Central de Saúde Pública do Ceará | Tayna da Silva Fiuza, Vânia Angélica Feitosa Viana, Shirlene Telmos Silva de Lima |
| EPI_ISL_18009975 | Hospital Municipal Jaime Osterno (Marco) | Laboratório Central de Saúde Pública do Ceará | Tayna da Silva Fiuza, Vânia Angélica Feitosa Viana, Shirlene Telmos Silva de Lima |
| EPI_ISL_18009976 | Lab De Analises Clinicas De Cruz | Laboratório Central de Saúde Pública do Ceará | Tayna da Silva Fiuza, Vânia Angélica Feitosa Viana, Shirlene Telmos Silva de Lima |
| EPI_ISL_18009977, EPI_ISL_18009978 | Hospital Municipal De Jaguaribe | Laboratório Central de Saúde Pública do Ceará | Tayna da Silva Fiuza, Vânia Angélica Feitosa Viana, Shirlene Telmos Silva de Lima |
| EPI_ISL_18009979 | Hospital Da Mulher De Fortaleza | Laboratório Central de Saúde Pública do Ceará | Tayna da Silva Fiuza, Vânia Angélica Feitosa Viana, Shirlene Telmos Silva de Lima |
| EPI_ISL_18009980 | Hospital Municipal Jaime Osterno (Marco) | Laboratório Central de Saúde Pública do Ceará | Tayna da Silva Fiuza, Vânia Angélica Feitosa Viana, Shirlene Telmos Silva de Lima |
| EPI_ISL_18009981, EPI_ISL_18009982, EPI_ISL_18009983 | Hospital Municipal De Jaguaribe | Laboratório Central de Saúde Pública do Ceará | Tayna da Silva Fiuza, Vânia Angélica Feitosa Viana, Shirlene Telmos Silva de Lima |
| EPI_ISL_18009984 | Lab De Analises Clinicas De Cruz | Laboratório Central de Saúde Pública do Ceará | Tayna da Silva Fiuza, Vânia Angélica Feitosa Viana, Shirlene Telmos Silva de Lima |
| EPI_ISL_18009985, EPI_ISL_18009986 | Hospital Municipal De Jaguaribe | Laboratório Central de Saúde Pública do Ceará | Tayna da Silva Fiuza, Vânia Angélica Feitosa Viana, Shirlene Telmos Silva de Lima |
| EPI_ISL_18009987 | Hospital Sao Jose (Fortaleza) | Laboratório Central de Saúde Pública do Ceará | Tayna da Silva Fiuza, Vânia Angélica Feitosa Viana, Shirlene Telmos Silva de Lima |
| EPI_ISL_18009988, EPI_ISL_18009989, EPI_ISL_18009990, EPI_ISL_18009991 | Lab De Analises Clinicas De Cruz | Laboratório Central de Saúde Pública do Ceará | Tayna da Silva Fiuza, Vânia Angélica Feitosa Viana, Shirlene Telmos Silva de Lima |
| EPI_ISL_18009992, EPI_ISL_18009993, EPI_ISL_18009994, EPI_ISL_18009995 | Hospital Municipal De Jaguaribe | Laboratório Central de Saúde Pública do Ceará | Tayna da Silva Fiuza, Vânia Angélica Feitosa Viana, Shirlene Telmos Silva de Lima |
| EPI_ISL_18009996 | Casa De Saude Adilia Maria (Boa Viagem) | Laboratório Central de Saúde Pública do Ceará | Tayna da Silva Fiuza, Vânia Angélica Feitosa Viana, Shirlene Telmos Silva de Lima |

| | | | |
|---|---|---|---|
| EPI_ISL_18009997 | Lab De Analises Clinicas De Cruz | Laboratório Central de Saúde Pública do Ceará | Tayna da Silva Fiuza, Vânia Angélica Feitosa Viana, Shirlene Telmos Silva de Lima |
| EPI_ISL_18009998 | Hospital Municipal De Jaguaribe | Laboratório Central de Saúde Pública do Ceará | Tayna da Silva Fiuza, Vânia Angélica Feitosa Viana, Shirlene Telmos Silva de Lima |
| EPI_ISL_18009999 | Centro De Saude Dr Nestor De Paula Pessoa (Acarau) | Laboratório Central de Saúde Pública do Ceará | Tayna da Silva Fiuza, Vânia Angélica Feitosa Viana, Shirlene Telmos Silva de Lima |
| EPI_ISL_18010000, EPI_ISL_18010001, EPI_ISL_18010002 | Lab De Analises Clinicas De Cruz | Laboratório Central de Saúde Pública do Ceará | Tayna da Silva Fiuza, Vânia Angélica Feitosa Viana, Shirlene Telmos Silva de Lima |
| EPI_ISL_18010003, EPI_ISL_18010004, EPI_ISL_18010005, EPI_ISL_18010006 | Hospital Municipal De Jaguaribe | Laboratório Central de Saúde Pública do Ceará | Tayna da Silva Fiuza, Vânia Angélica Feitosa Viana, Shirlene Telmos Silva de Lima |
| EPI_ISL_18010007, EPI_ISL_18010008, EPI_ISL_18010009 | Lab De Analises Clinicas De Cruz | Laboratório Central de Saúde Pública do Ceará | Tayna da Silva Fiuza, Vânia Angélica Feitosa Viana, Shirlene Telmos Silva de Lima |
| EPI_ISL_18010010, EPI_ISL_18010011 | Hospital Municipal De Jaguaribe | Laboratório Central de Saúde Pública do Ceará | Tayna da Silva Fiuza, Vânia Angélica Feitosa Viana, Shirlene Telmos Silva de Lima |
| EPI_ISL_18010012 | Lamac Laboratorio Municipal De Analises Clinicas (Camocim) | Laboratório Central de Saúde Pública do Ceará | Tayna da Silva Fiuza, Vânia Angélica Feitosa Viana, Shirlene Telmos Silva de Lima |
| EPI_ISL_18010013 | Hospital Municipal De Jaguaribe | Laboratório Central de Saúde Pública do Ceará | Tayna da Silva Fiuza, Vânia Angélica Feitosa Viana, Shirlene Telmos Silva de Lima |
| EPI_ISL_18010014 | Hospital Geral Dr Waldemar Alcantara | Laboratório Central de Saúde Pública do Ceará | Tayna da Silva Fiuza, Vânia Angélica Feitosa Viana, Shirlene Telmos Silva de Lima |
| EPI_ISL_18010015, EPI_ISL_18010016 | Hospital Municipal De Jaguaribe | Laboratório Central de Saúde Pública do Ceará | Tayna da Silva Fiuza, Vânia Angélica Feitosa Viana, Shirlene Telmos Silva de Lima |
| EPI_ISL_18010017, EPI_ISL_18010018 | Centro De Saude De Groairas | Laboratório Central de Saúde Pública do Ceará | Tayna da Silva Fiuza, Vânia Angélica Feitosa Viana, Shirlene Telmos Silva de Lima |
| EPI_ISL_18010019, EPI_ISL_18010020, EPI_ISL_18010021, EPI_ISL_18010022, EPI_ISL_18010023, EPI_ISL_18010024 | Centro De Saude De Bela Cruzfns(Bela Cruz) | Laboratório Central de Saúde Pública do Ceará | Tayna da Silva Fiuza, Vânia Angélica Feitosa Viana, Shirlene Telmos Silva de Lima |
| EPI_ISL_18010025 | Hospital Municipal De Jaguaribe | Laboratório Central de Saúde Pública do Ceará | Tayna da Silva Fiuza, Vânia Angélica Feitosa Viana, Shirlene Telmos Silva de Lima |
| EPI_ISL_18010026 | Centro De Saude De Bela Cruzfns(Bela Cruz) | Laboratório Central de Saúde Pública do Ceará | Tayna da Silva Fiuza, Vânia Angélica Feitosa Viana, Shirlene Telmos Silva de Lima |
| EPI_ISL_18010027, EPI_ISL_18010028 | Hospital Municipal De Jaguaribe | Laboratório Central de Saúde Pública do Ceará | Tayna da Silva Fiuza, Vânia Angélica Feitosa Viana, Shirlene Telmos Silva de Lima |
| EPI_ISL_18010029, EPI_ISL_18010030 | Centro De Saude De Bela Cruzfns(Bela Cruz) | Laboratório Central de Saúde Pública do Ceará | Tayna da Silva Fiuza, Vânia Angélica Feitosa Viana, Shirlene Telmos Silva de Lima |
| EPI_ISL_18029956, EPI_ISL_18029957, EPI_ISL_18029958, EPI_ISL_18029959, EPI_ISL_18029960, EPI_ISL_18029961, EPI_ISL_18029962, EPI_ISL_18029963, EPI_ISL_18029964, EPI_ISL_18029965, EPI_ISL_18029966, EPI_ISL_18029967, EPI_ISL_18029968, EPI_ISL_18029969, EPI_ISL_18029970, EPI_ISL_18029971, EPI_ISL_18029972, EPI_ISL_18029973 | | | |
| see above | Department of Microbiology | Department of Microbiology | Okada,W., Kasuya,F., Harada,S., Kumagai,R., Amano,A., Hasegawa,M., Miyake,H., Nagashima,M. and Sadamasu,K. |
| EPI_ISL_18059883, EPI_ISL_18059884, EPI_ISL_18059885 | Ministry of Health, Central Virology Laboratory | Ministry of Health, Central Virology Laboratory | Zuckerman,N.S., Schwartz,E., Pandey,P., Halpern,O., Bucris,E., Morad,H., Geva,M., Wax,M. and Lustig,Y. |
| EPI_ISL_18103416, EPI_ISL_18103417, EPI_ISL_18103418 | Undisclosed Laboratory in Brazil for the Health Emergency Information and Risk Assessment (HIM) Unit of PAHO | Undisclosed Laboratory in Brazil for the Health Emergency Information and Risk Assessment (HIM) Unit of PAHO | Vazquez,C., Luiz Carlos Junior Alcantara, Vagner Fonseca, Mauricio Lima, Joilson Xavier, Adelino,T., Hegger Fritsch, Castro,E., de Oliveira,C., Schuab,G., Lima,A.R.J., Villalba,S., Gomez de la Fuente,A., Rojas,A., Cantero,C., Fleitas,F., Aquino,C., Ojeda,A., Sequera,G., Torales,J., Barrios,J., Elias,M.C., Iani,F.C.M., Ortega,M.J., Gamarra,M.L., Montoya,R., Rodrigues,E.S., Simone Kashima, Sampaio,S.C., Coluchi,N., Leite,J., Gresh,L., Franco,L., Lourenco,J., Rico,J.M., Bispo de Filippis,A.M. and Marta Giovanetti |
| EPI_ISL_18103419 | Undisclosed Laboratory in Brazil for the Health Emergency Information and Risk Assessment (HIM) Unit of PAHO | Undisclosed Laboratory in Brazil for the Health Emergency Information and Risk Assessment (HIM) Unit of PAHO | Hegger Fritsch, Moreno,K., Lima,I.A.B., Santos,C.S., Costa,B.G.G., de Almeida,B.L., Dos Santos,R.A., Francisco,M.V.L.O., Sampaio,M.P.S., de Mauricio LimaM., Pereira,F.M., Vagner Fonseca, Tosta,S., Joilson Xavier, de Oliveira,C., Adelino,T., de Mello,A.L.E.S., Graf,T., Luiz Carlos Junior Alcantara, Marta Giovanetti and de Siqueira,I.C. |
| EPI_ISL_18103420, EPI_ISL_18103421, EPI_ISL_18103422, EPI_ISL_18103423, EPI_ISL_18103424, EPI_ISL_18103425, EPI_ISL_18103426, EPI_ISL_18103427, EPI_ISL_18103428, EPI_ISL_18103429, EPI_ISL_18103430, EPI_ISL_18103431, EPI_ISL_18103432 | | | |
| see above | Undisclosed Laboratory in Brazil for the Health Emergency Information and Risk Assessment (HIM) Unit of PAHO | Undisclosed Laboratory in Brazil for the Health Emergency Information and Risk Assessment (HIM) Unit of PAHO | Vazquez,C., Luiz Carlos Junior Alcantara, Vagner Fonseca, Mauricio Lima, Joilson Xavier, Adelino,T., Hegger Fritsch, Castro,E., de Oliveira,C., Schuab,G., Lima,A.R.J., Villalba,S., Gomez de la Fuente,A., Rojas,A., Cantero,C., Fleitas,F., Aquino,C., Ojeda,A., Sequera,G., Torales,J., Barrios,J., Elias,M.C., Iani,F.C.M., Ortega,M.J., Gamarra,M.L., Montoya,R., Rodrigues,E.S., Simone Kashima, Sampaio,S.C., Coluchi,N., Leite,J., Gresh,L., Franco,L., Lourenco,J., Rico,J.M., Bispo de Filippis,A.M. and Marta Giovanetti |
| EPI_ISL_18103433 | Undisclosed Laboratory in Brazil for the Health Emergency Information and Risk Assessment (HIM) Unit of PAHO | Undisclosed Laboratory in Brazil for the Health Emergency Information and Risk Assessment (HIM) Unit of PAHO | Hegger Fritsch, Moreno,K., Lima,I.A.B., Santos,C.S., Costa,B.G.G., de Almeida,B.L., Dos Santos,R.A., Francisco,M.V.L.O., Sampaio,M.P.S., de Mauricio LimaM., Pereira,F.M., Vagner Fonseca, Tosta,S., Joilson Xavier, de Oliveira,C., Adelino,T., de Mello,A.L.E.S., Graf,T., Luiz Carlos Junior Alcantara, Marta Giovanetti and de Siqueira,I.C. |
| EPI_ISL_18103434, EPI_ISL_18103435, EPI_ISL_18103436, EPI_ISL_18103437, EPI_ISL_18103438, EPI_ISL_18103439, EPI_ISL_18103440 | Undisclosed Laboratory in Brazil for the Health Emergency Information and Risk Assessment (HIM) Unit of PAHO | Undisclosed Laboratory in Brazil for the Health Emergency Information and Risk Assessment (HIM) Unit of PAHO | Vazquez,C., Luiz Carlos Junior Alcantara, Vagner Fonseca, Mauricio Lima, Joilson Xavier, Adelino,T., Hegger Fritsch, Castro,E., de Oliveira,C., Schuab,G., Lima,A.R.J., Villalba,S., Gomez de la Fuente,A., Rojas,A., Cantero,C., Fleitas,F., Aquino,C., Ojeda,A., Sequera,G., Torales,J., Barrios,J., Elias,M.C., Iani,F.C.M., Ortega,M.J., Gamarra,M.L., Montoya,R., Rodrigues,E.S., Simone Kashima, Sampaio,S.C., Coluchi,N., Leite,J., Gresh,L., Franco,L., Lourenco,J., Rico,J.M., Bispo de Filippis,A.M. and Marta Giovanetti |
| EPI_ISL_18103441 | Undisclosed Laboratory in Brazil for the Health Emergency Information and Risk Assessment (HIM) Unit of PAHO | Undisclosed Laboratory in Brazil for the Health Emergency Information and Risk Assessment (HIM) Unit of PAHO | Hegger Fritsch, Moreno,K., Lima,I.A.B., Santos,C.S., Costa,B.G.G., de Almeida,B.L., Dos Santos,R.A., Francisco,M.V.L.O., Sampaio,M.P.S., de Mauricio LimaM., Pereira,F.M., Vagner Fonseca, Tosta,S., Joilson Xavier, de Oliveira,C., Adelino,T., de Mello,A.L.E.S., Graf,T., Luiz Carlos Junior Alcantara, Marta Giovanetti and de Siqueira,I.C. |
| EPI_ISL_18103442, EPI_ISL_18103443, EPI_ISL_18103444 | Undisclosed Laboratory in Brazil for the Health Emergency Information and Risk Assessment (HIM) Unit of PAHO | Undisclosed Laboratory in Brazil for the Health Emergency Information and Risk Assessment (HIM) Unit of PAHO | Vazquez,C., Luiz Carlos Junior Alcantara, Vagner Fonseca, Mauricio Lima, Joilson Xavier, Adelino,T., Hegger Fritsch, Castro,E., de Oliveira,C., Schuab,G., Lima,A.R.J., Villalba,S., Gomez de la Fuente,A., Rojas,A., Cantero,C., Fleitas,F., Aquino,C., Ojeda,A., Sequera,G., Torales,J., Barrios,J., Elias,M.C., Iani,F.C.M., Ortega,M.J., Gamarra,M.L., Montoya,R., Rodrigues,E.S., Simone Kashima, Sampaio,S.C., Coluchi,N., Leite,J., Gresh,L., Franco,L., Lourenco,J., Rico,J.M., Bispo de Filippis,A.M. and Marta Giovanetti |
| EPI_ISL_18103445 | Undisclosed Laboratory in Brazil for the Health Emergency Information and Risk Assessment (HIM) Unit of PAHO | Undisclosed Laboratory in Brazil for the Health Emergency Information and Risk Assessment (HIM) Unit of PAHO | Hegger Fritsch, Moreno,K., Lima,I.A.B., Santos,C.S., Costa,B.G.G., de Almeida,B.L., Dos Santos,R.A., Francisco,M.V.L.O., Sampaio,M.P.S., de Mauricio LimaM., Pereira,F.M., Vagner Fonseca, Tosta,S., Joilson Xavier, de Oliveira,C., Adelino,T., de Mello,A.L.E.S., Graf,T., Luiz Carlos Junior Alcantara, Marta Giovanetti and de Siqueira,I.C. |
| EPI_ISL_18103446, EPI_ISL_18103447, EPI_ISL_18103448 | Undisclosed Laboratory in Brazil for the Health Emergency Information and Risk Assessment (HIM) Unit of PAHO | Undisclosed Laboratory in Brazil for the Health Emergency Information and Risk Assessment (HIM) Unit of PAHO | Vazquez,C., Luiz Carlos Junior Alcantara, Vagner Fonseca, Mauricio Lima, Joilson Xavier, Adelino,T., Hegger Fritsch, Castro,E., de Oliveira,C., Schuab,G., Lima,A.R.J., Villalba,S., Gomez de la Fuente,A., Rojas,A., Cantero,C., Fleitas,F., Aquino,C., Ojeda,A., Sequera,G., Torales,J., Barrios,J., Elias,M.C., Iani,F.C.M., Ortega,M.J., Gamarra,M.L., Montoya,R., Rodrigues,E.S., Simone Kashima, Sampaio,S.C., Coluchi,N., Leite,J., Gresh,L., Franco,L., Lourenco,J., Rico,J.M., Bispo de Filippis,A.M. and Marta Giovanetti |
| EPI_ISL_18103449, EPI_ISL_18103450, EPI_ISL_18103451 | Undisclosed Laboratory in Brazil for the Health Emergency Information and Risk Assessment (HIM) Unit of PAHO | Undisclosed Laboratory in Brazil for the Health Emergency Information and Risk Assessment (HIM) Unit of PAHO | Hegger Fritsch, Moreno,K., Lima,I.A.B., Santos,C.S., Costa,B.G.G., de Almeida,B.L., Dos Santos,R.A., Francisco,M.V.L.O., Sampaio,M.P.S., de Mauricio LimaM., Pereira,F.M., Vagner Fonseca, Tosta,S., Joilson Xavier, de Oliveira,C., Adelino,T., de Mello,A.L.E.S., Graf,T., Luiz Carlos Junior Alcantara, Marta Giovanetti and de Siqueira,I.C. |
| EPI_ISL_18103452, EPI_ISL_18103453, EPI_ISL_18103454, EPI_ISL_18103455, EPI_ISL_18103456, EPI_ISL_18103457, EPI_ISL_18103458, EPI_ISL_18103459 | Undisclosed Laboratory in Brazil for the Health Emergency Information and Risk Assessment (HIM) Unit of PAHO | Undisclosed Laboratory in Brazil for the Health Emergency Information and Risk Assessment (HIM) Unit of PAHO | Vazquez,C., Luiz Carlos Junior Alcantara, Vagner Fonseca, Mauricio Lima, Joilson Xavier, Adelino,T., Hegger Fritsch, Castro,E., de Oliveira,C., Schuab,G., Lima,A.R.J., Villalba,S., Gomez de la Fuente,A., Rojas,A., Cantero,C., Fleitas,F., Aquino,C., Ojeda,A., Sequera,G., Torales,J., Barrios,J., Elias,M.C., Iani,F.C.M., Ortega,M.J., Gamarra,M.L., Montoya,R., Rodrigues,E.S., Simone Kashima, Sampaio,S.C., Coluchi,N., Leite,J., Gresh,L., Franco,L., Lourenco,J., Rico,J.M., Bispo de Filippis,A.M. and Marta Giovanetti |
| EPI_ISL_18103460 | Undisclosed Laboratory in Brazil for the Health Emergency Information and Risk Assessment (HIM) Unit of PAHO | Undisclosed Laboratory in Brazil for the Health Emergency Information and Risk Assessment (HIM) Unit of PAHO | Hegger Fritsch, Moreno,K., Lima,I.A.B., Santos,C.S., Costa,B.G.G., de Almeida,B.L., Dos Santos,R.A., Francisco,M.V.L.O., Sampaio,M.P.S., de Mauricio LimaM., Pereira,F.M., Vagner Fonseca, Tosta,S., Joilson Xavier, de Oliveira,C., Adelino,T., de Mello,A.L.E.S., Graf,T., Luiz Carlos Junior Alcantara, Marta Giovanetti and de Siqueira,I.C. |
| EPI_ISL_18103461, EPI_ISL_18103462, EPI_ISL_18103463 | Undisclosed Laboratory in Brazil for the Health Emergency Information and Risk Assessment (HIM) Unit of PAHO | Undisclosed Laboratory in Brazil for the Health Emergency Information and Risk Assessment (HIM) Unit of PAHO | Vazquez,C., Luiz Carlos Junior Alcantara, Vagner Fonseca, Mauricio Lima, Joilson Xavier, Adelino,T., Hegger Fritsch, Castro,E., de Oliveira,C., Schuab,G., Lima,A.R.J., Villalba,S., Gomez de la Fuente,A., Rojas,A., Cantero,C., Fleitas,F., Aquino,C., Ojeda,A., Sequera,G., Torales,J., Barrios,J., Elias,M.C., Iani,F.C.M., Ortega,M.J., Gamarra,M.L., Montoya,R., Rodrigues,E.S., Simone Kashima, Sampaio,S.C., Coluchi,N., Leite,J., Gresh,L., Franco,L., Lourenco,J., Rico,J.M., Bispo de Filippis,A.M. and Marta Giovanetti |
| EPI_ISL_18103464 | Undisclosed Laboratory in Brazil for the Health Emergency Information and Risk Assessment (HIM) Unit of PAHO | Undisclosed Laboratory in Brazil for the Health Emergency Information and Risk Assessment (HIM) Unit of PAHO | Hegger Fritsch, Moreno,K., Lima,I.A.B., Santos,C.S., Costa,B.G.G., de Almeida,B.L., Dos Santos,R.A., Francisco,M.V.L.O., Sampaio,M.P.S., de Mauricio LimaM., Pereira,F.M., Vagner Fonseca, Tosta,S., Joilson Xavier, de Oliveira,C., Adelino,T., de Mello,A.L.E.S., Graf,T., Luiz Carlos Junior Alcantara, Marta Giovanetti and de Siqueira,I.C. |
| EPI_ISL_18103465 | Undisclosed Laboratory in Brazil for the Health Emergency Information and Risk Assessment (HIM) Unit of PAHO | Undisclosed Laboratory in Brazil for the Health Emergency Information and Risk Assessment (HIM) Unit of PAHO | Vazquez,C., Luiz Carlos Junior Alcantara, Vagner Fonseca, Mauricio Lima, Joilson Xavier, Adelino,T., Hegger Fritsch, Castro,E., de Oliveira,C., Schuab,G., Lima,A.R.J., Villalba,S., Gomez de la Fuente,A., Rojas,A., Cantero,C., Fleitas,F., Aquino,C., Ojeda,A., Sequera,G., Torales,J., Barrios,J., Elias,M.C., Iani,F.C.M., Ortega,M.J., Gamarra,M.L., Montoya,R., Rodrigues,E.S., Simone Kashima, Sampaio,S.C., Coluchi,N., Leite,J., Gresh,L., Franco,L., Lourenco,J., Rico,J.M., Bispo de Filippis,A.M. and Marta Giovanetti |
| EPI_ISL_18103466, EPI_ISL_18103467 | Undisclosed Laboratory in Brazil for the Health Emergency Information and Risk Assessment (HIM) Unit of PAHO | Undisclosed Laboratory in Brazil for the Health Emergency Information and Risk Assessment (HIM) Unit of PAHO | Hegger Fritsch, Moreno,K., Lima,I.A.B., Santos,C.S., Costa,B.G.G., de Almeida,B.L., Dos Santos,R.A., Francisco,M.V.L.O., Sampaio,M.P.S., de Mauricio LimaM., Pereira,F.M., Vagner Fonseca, Tosta,S., Joilson Xavier, de Oliveira,C., Adelino,T., de Mello,A.L.E.S., Graf,T., Luiz Carlos Junior Alcantara, Marta Giovanetti and de Siqueira,I.C. |
| EPI_ISL_18103468 | Undisclosed Laboratory in Brazil for the Health Emergency Information and Risk Assessment (HIM) Unit of PAHO | Undisclosed Laboratory in Brazil for the Health Emergency Information and Risk Assessment (HIM) Unit of PAHO | Vazquez,C., Luiz Carlos Junior Alcantara, Vagner Fonseca, Mauricio Lima, Joilson Xavier, Adelino,T., Hegger Fritsch, Castro,E., de Oliveira,C., Schuab,G., Lima,A.R.J., Villalba,S., Gomez de la Fuente,A., Rojas,A., Cantero,C., Fleitas,F., Aquino,C., Ojeda,A., Sequera,G., Torales,J., Barrios,J., Elias,M.C., Iani,F.C.M., Ortega,M.J., Gamarra,M.L., Montoya,R., Rodrigues,E.S., Simone Kashima, Sampaio,S.C., Coluchi,N., Leite,J., Gresh,L., Franco,L., Lourenco,J., Rico,J.M., Bispo de Filippis,A.M. and Marta Giovanetti |
| EPI_ISL_18103469 | Undisclosed Laboratory in Brazil for the Health Emergency Information and Risk Assessment (HIM) Unit of PAHO | Undisclosed Laboratory in Brazil for the Health Emergency Information and Risk Assessment (HIM) Unit of PAHO | Hegger Fritsch, Moreno,K., Lima,I.A.B., Santos,C.S., Costa,B.G.G., de Almeida,B.L., Dos Santos,R.A., Francisco,M.V.L.O., Sampaio,M.P.S., de Mauricio LimaM., Pereira,F.M., Vagner Fonseca, Tosta,S., Joilson Xavier, de Oliveira,C., Adelino,T., de Mello,A.L.E.S., Graf,T., Luiz Carlos Junior Alcantara, Marta Giovanetti and de Siqueira,I.C. |
| EPI_ISL_18103470, EPI_ISL_18103471, EPI_ISL_18103472 | Undisclosed Laboratory in Brazil for the Health Emergency Information and Risk Assessment (HIM) Unit of PAHO | Undisclosed Laboratory in Brazil for the Health Emergency Information and Risk Assessment (HIM) Unit of PAHO | Vazquez,C., Luiz Carlos Junior Alcantara, Vagner Fonseca, Mauricio Lima, Joilson Xavier, Adelino,T., Hegger Fritsch, Castro,E., de Oliveira,C., Schuab,G., Lima,A.R.J., Villalba,S., Gomez de la Fuente,A., Rojas,A., Cantero,C., Fleitas,F., Aquino,C., Ojeda,A., Sequera,G., Torales,J., Barrios,J., Elias,M.C., Iani,F.C.M., Ortega,M.J., Gamarra,M.L., Montoya,R., Rodrigues,E.S., Simone Kashima, Sampaio,S.C., Coluchi,N., Leite,J., Gresh,L., Franco,L., Lourenco,J., Rico,J.M., Bispo de Filippis,A.M. and Marta Giovanetti |
| EPI_ISL_18103473 | Undisclosed Laboratory in Brazil for the Health Emergency Information and Risk Assessment (HIM) Unit of PAHO | Undisclosed Laboratory in Brazil for the Health Emergency Information and Risk Assessment (HIM) Unit of PAHO | Hegger Fritsch, Moreno,K., Lima,I.A.B., Santos,C.S., Costa,B.G.G., de Almeida,B.L., Dos Santos,R.A., Francisco,M.V.L.O., Sampaio,M.P.S., de Mauricio LimaM., Pereira,F.M., Vagner Fonseca, Tosta,S., Joilson Xavier, de Oliveira,C., Adelino,T., de Mello,A.L.E.S., Graf,T., Luiz Carlos Junior Alcantara, Marta Giovanetti and de Siqueira,I.C. |
| EPI_ISL_18103474, EPI_ISL_18103475 | Undisclosed Laboratory in Brazil for the Health Emergency Information and Risk Assessment (HIM) Unit of PAHO | Undisclosed Laboratory in Brazil for the Health Emergency Information and Risk Assessment (HIM) Unit of PAHO | Vazquez,C., Luiz Carlos Junior Alcantara, Vagner Fonseca, Mauricio Lima, Joilson Xavier, Adelino,T., Hegger Fritsch, Castro,E., de Oliveira,C., Schuab,G., Lima,A.R.J., Villalba,S., Gomez de la Fuente,A., Rojas,A., Cantero,C., Fleitas,F., Aquino,C., Ojeda,A., Sequera,G., Torales,J., Barrios,J., Elias,M.C., Iani,F.C.M., Ortega,M.J., Gamarra,M.L., Montoya,R., Rodrigues,E.S., Simone Kashima, Sampaio,S.C., Coluchi,N., Leite,J., Gresh,L., Franco,L., Lourenco,J., Rico,J.M., Bispo de Filippis,A.M. and Marta Giovanetti |
| EPI_ISL_18103476 | Undisclosed Laboratory in Brazil for the Health Emergency Information and Risk Assessment (HIM) Unit of PAHO | Undisclosed Laboratory in Brazil for the Health Emergency Information and Risk Assessment (HIM) Unit of PAHO | Hegger Fritsch, Moreno,K., Lima,I.A.B., Santos,C.S., Costa,B.G.G., de Almeida,B.L., Dos Santos,R.A., Francisco,M.V.L.O., Sampaio,M.P.S., de Mauricio LimaM., Pereira,F.M., Vagner Fonseca, Tosta,S., Joilson Xavier, de Oliveira,C., Adelino,T., de Mello,A.L.E.S., Graf,T., Luiz Carlos Junior Alcantara, Marta Giovanetti and de Siqueira,I.C. |

| | | | |
|---|---|---|---|
| EPI_ISL_18103477, EPI_ISL_18103478, EPI_ISL_18103479, EPI_ISL_18103480, EPI_ISL_18103481, EPI_ISL_18103482, EPI_ISL_18103483, EPI_ISL_18103484, EPI_ISL_18103485, EPI_ISL_18103486, EPI_ISL_18103487, EPI_ISL_18103488, EPI_ISL_18103489 | | | |
| see above | Undisclosed Laboratory in Brazil for the Health Emergency Information and Risk Assessment (HIM) Unit of PAHO | Undisclosed Laboratory in Brazil for the Health Emergency Information and Risk Assessment (HIM) Unit of PAHO | Vazquez,C., Luiz Carlos Junior Alcantara, Vagner Fonseca, Mauricio Lima, Joilson Xavier, Adelino,T., Hegger Fritsch, Castro,E., de Oliveira,C., Schuab,G., Lima,A.R.J., Villalba,S., Gomez de la Fuente,A., Rojas,A., Cantero,C., Fleitas,F., Aquino,C., Ojeda,A., Sequera,G., Torales,J., Barrios,J., Elias,M.C., Iani,F.C.M., Ortega,M.J., Gamarra,M.L., Montoya,R., Rodrigues,E.S., Simone Kashima, Sampaio,S.C., Coluchi,N., Leite,J., Gresh,L., Franco,L., Lourenco,J., Rico,J.M., Bispo de Filippis,A.M. and Marta Giovanetti |
| EPI_ISL_18103490 | Undisclosed Laboratory in Brazil for the Health Emergency Information and Risk Assessment (HIM) Unit of PAHO | Undisclosed Laboratory in Brazil for the Health Emergency Information and Risk Assessment (HIM) Unit of PAHO | Hegger Fritsch, Moreno,K., Lima,I.A.B., Santos,C.S., Costa,B.G.G., de Almeida,B.L., Dos Santos,R.A., Francisco,M.V.L.O., Sampaio,M.P.S., de Mauricio LimaM., Pereira,F.M., Vagner Fonseca, Tosta,S., Joilson Xavier, de Oliveira,C., Adelino,T., de Mello,A.L.E.S., Graf,T., Luiz Carlos Junior Alcantara, Marta Giovanetti and de Siqueira,I.C. |
| EPI_ISL_18103491, EPI_ISL_18103492, EPI_ISL_18103493, EPI_ISL_18103494, EPI_ISL_18103495, EPI_ISL_18103496, EPI_ISL_18103497, EPI_ISL_18103498, EPI_ISL_18103499, EPI_ISL_18103500, EPI_ISL_18103501, EPI_ISL_18103502, EPI_ISL_18103503, EPI_ISL_18103504 | | | |
| see above | Undisclosed Laboratory in Brazil for the Health Emergency Information and Risk Assessment (HIM) Unit of PAHO | Undisclosed Laboratory in Brazil for the Health Emergency Information and Risk Assessment (HIM) Unit of PAHO | Vazquez,C., Luiz Carlos Junior Alcantara, Vagner Fonseca, Mauricio Lima, Joilson Xavier, Adelino,T., Hegger Fritsch, Castro,E., de Oliveira,C., Schuab,G., Lima,A.R.J., Villalba,S., Gomez de la Fuente,A., Rojas,A., Cantero,C., Fleitas,F., Aquino,C., Ojeda,A., Sequera,G., Torales,J., Barrios,J., Elias,M.C., Iani,F.C.M., Ortega,M.J., Gamarra,M.L., Montoya,R., Rodrigues,E.S., Simone Kashima, Sampaio,S.C., Coluchi,N., Leite,J., Gresh,L., Franco,L., Lourenco,J., Rico,J.M., Bispo de Filippis,A.M. and Marta Giovanetti |
| EPI_ISL_18103505 | Undisclosed Laboratory in Brazil for the Health Emergency Information and Risk Assessment (HIM) Unit of PAHO | Undisclosed Laboratory in Brazil for the Health Emergency Information and Risk Assessment (HIM) Unit of PAHO | Hegger Fritsch, Moreno,K., Lima,I.A.B., Santos,C.S., Costa,B.G.G., de Almeida,B.L., Dos Santos,R.A., Francisco,M.V.L.O., Sampaio,M.P.S., de Mauricio LimaM., Pereira,F.M., Vagner Fonseca, Tosta,S., Joilson Xavier, de Oliveira,C., Adelino,T., de Mello,A.L.E.S., Graf,T., Luiz Carlos Junior Alcantara, Marta Giovanetti and de Siqueira,I.C. |
| EPI_ISL_18103506 | Undisclosed Laboratory in Brazil for the Health Emergency Information and Risk Assessment (HIM) Unit of PAHO | Undisclosed Laboratory in Brazil for the Health Emergency Information and Risk Assessment (HIM) Unit of PAHO | Vazquez,C., Luiz Carlos Junior Alcantara, Vagner Fonseca, Mauricio Lima, Joilson Xavier, Adelino,T., Hegger Fritsch, Castro,E., de Oliveira,C., Schuab,G., Lima,A.R.J., Villalba,S., Gomez de la Fuente,A., Rojas,A., Cantero,C., Fleitas,F., Aquino,C., Ojeda,A., Sequera,G., Torales,J., Barrios,J., Elias,M.C., Iani,F.C.M., Ortega,M.J., Gamarra,M.L., Montoya,R., Rodrigues,E.S., Simone Kashima, Sampaio,S.C., Coluchi,N., Leite,J., Gresh,L., Franco,L., Lourenco,J., Rico,J.M., Bispo de Filippis,A.M. and Marta Giovanetti |
| EPI_ISL_18103507 | Undisclosed Laboratory in Brazil for the Health Emergency Information and Risk Assessment (HIM) Unit of PAHO | Undisclosed Laboratory in Brazil for the Health Emergency Information and Risk Assessment (HIM) Unit of PAHO | Hegger Fritsch, Moreno,K., Lima,I.A.B., Santos,C.S., Costa,B.G.G., de Almeida,B.L., Dos Santos,R.A., Francisco,M.V.L.O., Sampaio,M.P.S., de Mauricio LimaM., Pereira,F.M., Vagner Fonseca, Tosta,S., Joilson Xavier, de Oliveira,C., Adelino,T., de Mello,A.L.E.S., Graf,T., Luiz Carlos Junior Alcantara, Marta Giovanetti and de Siqueira,I.C. |
| EPI_ISL_18103508, EPI_ISL_18103509 | Undisclosed Laboratory in Brazil for the Health Emergency Information and Risk Assessment (HIM) Unit of PAHO | Undisclosed Laboratory in Brazil for the Health Emergency Information and Risk Assessment (HIM) Unit of PAHO | Vazquez,C., Luiz Carlos Junior Alcantara, Vagner Fonseca, Mauricio Lima, Joilson Xavier, Adelino,T., Hegger Fritsch, Castro,E., de Oliveira,C., Schuab,G., Lima,A.R.J., Villalba,S., Gomez de la Fuente,A., Rojas,A., Cantero,C., Fleitas,F., Aquino,C., Ojeda,A., Sequera,G., Torales,J., Barrios,J., Elias,M.C., Iani,F.C.M., Ortega,M.J., Gamarra,M.L., Montoya,R., Rodrigues,E.S., Simone Kashima, Sampaio,S.C., Coluchi,N., Leite,J., Gresh,L., Franco,L., Lourenco,J., Rico,J.M., Bispo de Filippis,A.M. and Marta Giovanetti |
| EPI_ISL_18103510 | Undisclosed Laboratory in Brazil for the Health Emergency Information and Risk Assessment (HIM) Unit of PAHO | Undisclosed Laboratory in Brazil for the Health Emergency Information and Risk Assessment (HIM) Unit of PAHO | Hegger Fritsch, Moreno,K., Lima,I.A.B., Santos,C.S., Costa,B.G.G., de Almeida,B.L., Dos Santos,R.A., Francisco,M.V.L.O., Sampaio,M.P.S., de Mauricio LimaM., Pereira,F.M., Vagner Fonseca, Tosta,S., Joilson Xavier, de Oliveira,C., Adelino,T., de Mello,A.L.E.S., Graf,T., Luiz Carlos Junior Alcantara, Marta Giovanetti and de Siqueira,I.C. |
| EPI_ISL_18103511, EPI_ISL_18103512, EPI_ISL_18103513, EPI_ISL_18103514, EPI_ISL_18103515, EPI_ISL_18103516, EPI_ISL_18103517, EPI_ISL_18103518, EPI_ISL_18103519, EPI_ISL_18103520, EPI_ISL_18103521, EPI_ISL_18103522 | | | |
| see above | Undisclosed Laboratory in Brazil for the Health Emergency Information and Risk Assessment (HIM) Unit of PAHO | Undisclosed Laboratory in Brazil for the Health Emergency Information and Risk Assessment (HIM) Unit of PAHO | Vazquez,C., Luiz Carlos Junior Alcantara, Vagner Fonseca, Mauricio Lima, Joilson Xavier, Adelino,T., Hegger Fritsch, Castro,E., de Oliveira,C., Schuab,G., Lima,A.R.J., Villalba,S., Gomez de la Fuente,A., Rojas,A., Cantero,C., Fleitas,F., Aquino,C., Ojeda,A., Sequera,G., Torales,J., Barrios,J., Elias,M.C., Iani,F.C.M., Ortega,M.J., Gamarra,M.L., Montoya,R., Rodrigues,E.S., Simone Kashima, Sampaio,S.C., Coluchi,N., Leite,J., Gresh,L., Franco,L., Lourenco,J., Rico,J.M., Bispo de Filippis,A.M. and Marta Giovanetti |
| EPI_ISL_18103523 | Undisclosed Laboratory in Brazil for the Health Emergency Information and Risk Assessment (HIM) Unit of PAHO | Undisclosed Laboratory in Brazil for the Health Emergency Information and Risk Assessment (HIM) Unit of PAHO | Hegger Fritsch, Moreno,K., Lima,I.A.B., Santos,C.S., Costa,B.G.G., de Almeida,B.L., Dos Santos,R.A., Francisco,M.V.L.O., Sampaio,M.P.S., de Mauricio LimaM., Pereira,F.M., Vagner Fonseca, Tosta,S., Joilson Xavier, de Oliveira,C., Adelino,T., de Mello,A.L.E.S., Graf,T., Luiz Carlos Junior Alcantara, Marta Giovanetti and de Siqueira,I.C. |
| EPI_ISL_18103524, EPI_ISL_18103525, EPI_ISL_18103526, EPI_ISL_18103527, EPI_ISL_18103528, EPI_ISL_18103529, EPI_ISL_18103530, EPI_ISL_18103531, EPI_ISL_18103532, EPI_ISL_18103533 | Undisclosed Laboratory in Brazil for the Health Emergency Information and Risk Assessment (HIM) Unit of PAHO | Undisclosed Laboratory in Brazil for the Health Emergency Information and Risk Assessment (HIM) Unit of PAHO | Vazquez,C., Luiz Carlos Junior Alcantara, Vagner Fonseca, Mauricio Lima, Joilson Xavier, Adelino,T., Hegger Fritsch, Castro,E., de Oliveira,C., Schuab,G., Lima,A.R.J., Villalba,S., Gomez de la Fuente,A., Rojas,A., Cantero,C., Fleitas,F., Aquino,C., Ojeda,A., Sequera,G., Torales,J., Barrios,J., Elias,M.C., Iani,F.C.M., Ortega,M.J., Gamarra,M.L., Montoya,R., Rodrigues,E.S., Simone Kashima, Sampaio,S.C., Coluchi,N., Leite,J., Gresh,L., Franco,L., Lourenco,J., Rico,J.M., Bispo de Filippis,A.M. and Marta Giovanetti |
| EPI_ISL_18358948, EPI_ISL_18358949, EPI_ISL_18358950, EPI_ISL_18358951, EPI_ISL_18358952, EPI_ISL_18358953, EPI_ISL_18358954, EPI_ISL_18358955, EPI_ISL_18358956, EPI_ISL_18358957, EPI_ISL_18358958, EPI_ISL_18358959, EPI_ISL_18358960, EPI_ISL_18358961, EPI_ISL_18358962, EPI_ISL_18358963, EPI_ISL_18358964, EPI_ISL_18358965, EPI_ISL_18358966, EPI_ISL_18358967, EPI_ISL_18358968, EPI_ISL_18358969, EPI_ISL_18358970, EPI_ISL_18358971, EPI_ISL_18358972, EPI_ISL_18358973, EPI_ISL_18358974, EPI_ISL_18358975, EPI_ISL_18358976, EPI_ISL_18358977, EPI_ISL_18358978, EPI_ISL_18358979 | | | |
| see above | Institut Pasteur de la Guadeloupe | Institut Pasteur de la Guadeloupe | Garcia-Van Smevoorde,M., Piorkowski,G., Emboule,L., Dos Santos,G., Loraux,C., Guyomard-Rabenirina,S., Joannes,M.O., Fagour,L., Najioullah,F., Cabie,A., de Lamballerie,X., Vega-Rua,A., Cesaire,R. and Calvez,E. |
| EPI_ISL_18360767, EPI_ISL_18360768, EPI_ISL_18360769, EPI_ISL_18360770, EPI_ISL_18360771, EPI_ISL_18360772, EPI_ISL_18360773, EPI_ISL_18360774, EPI_ISL_18360775, EPI_ISL_18360776, EPI_ISL_18360777, EPI_ISL_18360778, EPI_ISL_18360779, EPI_ISL_18360780, EPI_ISL_18360781, EPI_ISL_18360782, EPI_ISL_18360783, EPI_ISL_18360784, EPI_ISL_18360785, EPI_ISL_18360786, EPI_ISL_18360787, EPI_ISL_18360788, EPI_ISL_18360789, EPI_ISL_18360790, EPI_ISL_18360791, EPI_ISL_18360792, EPI_ISL_18360793, EPI_ISL_18360794, EPI_ISL_18360795, EPI_ISL_18360796, EPI_ISL_18360797, EPI_ISL_18360798, EPI_ISL_18360799, EPI_ISL_18360800, EPI_ISL_18360801, EPI_ISL_18360802, EPI_ISL_18360803, EPI_ISL_18360804, EPI_ISL_18360805, EPI_ISL_18360806, EPI_ISL_18360807, EPI_ISL_18360808, EPI_ISL_18360809, EPI_ISL_18360810, EPI_ISL_18360811, EPI_ISL_18360812, EPI_ISL_18360813, EPI_ISL_18360814, EPI_ISL_18360815, EPI_ISL_18360816, EPI_ISL_18360817, EPI_ISL_18360818, EPI_ISL_18360819, EPI_ISL_18360820, EPI_ISL_18360821, EPI_ISL_18360822, EPI_ISL_18360823, EPI_ISL_18360824, EPI_ISL_18360825, EPI_ISL_18360826, EPI_ISL_18360827, EPI_ISL_18360828, EPI_ISL_18360829, EPI_ISL_18360830, EPI_ISL_18360831, EPI_ISL_18360832, EPI_ISL_18360833, EPI_ISL_18360834, EPI_ISL_18360835, EPI_ISL_18360836, EPI_ISL_18360837, EPI_ISL_18360838, EPI_ISL_18360839, EPI_ISL_18360840, EPI_ISL_18360841, EPI_ISL_18360842, EPI_ISL_18360843, EPI_ISL_18360844, EPI_ISL_18360845, EPI_ISL_18360846, EPI_ISL_18360847, EPI_ISL_18360848, EPI_ISL_18360849, EPI_ISL_18360850, EPI_ISL_18360851, EPI_ISL_18360852, EPI_ISL_18360853, EPI_ISL_18360854, EPI_ISL_18360855, EPI_ISL_18360856, EPI_ISL_18360857, EPI_ISL_18360858, EPI_ISL_18360859, EPI_ISL_18360860, EPI_ISL_18360861, EPI_ISL_18360862, EPI_ISL_18360863, EPI_ISL_18360864, EPI_ISL_18360865, EPI_ISL_18360866, EPI_ISL_18360867, EPI_ISL_18360868, EPI_ISL_18360869, EPI_ISL_18360870, EPI_ISL_18360871, EPI_ISL_18360872, EPI_ISL_18360873, EPI_ISL_18360874, EPI_ISL_18360875, EPI_ISL_18360876, EPI_ISL_18360877, EPI_ISL_18360878, EPI_ISL_18360879, EPI_ISL_18360880, EPI_ISL_18360881, EPI_ISL_18360882, EPI_ISL_18360883, EPI_ISL_18360884, EPI_ISL_18360885, EPI_ISL_18360886, EPI_ISL_18360887, EPI_ISL_18360888, EPI_ISL_18360889, EPI_ISL_18360890, EPI_ISL_18360891, EPI_ISL_18360892, EPI_ISL_18360893, EPI_ISL_18360894, EPI_ISL_18360895, EPI_ISL_18360896, EPI_ISL_18360897, EPI_ISL_18360898, EPI_ISL_18360899, EPI_ISL_18360900, EPI_ISL_18360901, EPI_ISL_18360902, EPI_ISL_18360903, EPI_ISL_18360904, EPI_ISL_18360905, EPI_ISL_18360906, EPI_ISL_18360907, EPI_ISL_18360908, EPI_ISL_18360909, EPI_ISL_18360910, EPI_ISL_18360911, EPI_ISL_18360912, EPI_ISL_18360913, EPI_ISL_18360914, EPI_ISL_18360915, EPI_ISL_18360916, EPI_ISL_18360917, EPI_ISL_18360918, EPI_ISL_18360919, EPI_ISL_18360920, EPI_ISL_18360921, EPI_ISL_18360922, EPI_ISL_18360923, EPI_ISL_18360924, EPI_ISL_18360925, EPI_ISL_18360926, EPI_ISL_18360927, EPI_ISL_18360928, EPI_ISL_18360929, EPI_ISL_18360930, EPI_ISL_18360931, EPI_ISL_18360932, EPI_ISL_18360933, EPI_ISL_18360934, EPI_ISL_18360935, EPI_ISL_18360936, EPI_ISL_18360937, EPI_ISL_18360938, EPI_ISL_18360939, EPI_ISL_18360940, EPI_ISL_18360941, EPI_ISL_18360942, EPI_ISL_18360943 | | | |
| see above | Laboratório Central de Saúde Pública do Estado de Mato Grosso do Sul | Laboratório Central de Saúde Pública do Estado de Mato Grosso do Sul | Larissa Domingues Castilho de Arruda, Marta Giovanetti, Vagner Fonseca, Marina Castilhos Souza Umaki Zardin, Gislene Garcia de Castro Lichs, Silvia Asato, Ana Olivia Pascoto Esposito, Miriam Tokeshi Müller, Joilson Xavier, Hegger Fritsch, Mauricio Lima, Carla de Oliveira, Elaine Vieira Santos, Livia de Mello Almeida Maziero, Danila Fernanda Rodrigues Frias, Danielle Ahad das Neves, Liliane Ferreira da Silva, Ellen Caroline Rodrigues Barretos, Paulo Eduardo Tsuha Oshiro, Bianca Modafari Goday, Jessica Klener Lemos dos Santos, Simone Kashima, Carlos F. C. de Albuquerque, Rodrigo Fabiano do Carmo Said, Alexander Rosewell, Luiz Henrique Ferraz Demarchi, Julio Croda, Luiz Carlos Junior Alcantara and Crhistinne Cavalheiro Maymone Gonçalves |
| EPI_ISL_18414933 | National University of Singapore | National University of Singapore | Chin,W.X., Teo,Z.Y. and Chu,J.J.H. |
| EPI_ISL_18474688, EPI_ISL_18474689 | Universidad de Antioquia | Universidad de Antioquia | Ciuoderis,K.A., Usuga,J., Moreno-Lopez,I., Perez-Restrepo,L.S., Florez,D.Y., Hernandez-Ortiz,J.P., Cloherty,G.A. and Osorio,J.E. |
| EPI_ISL_18474690 | Universidad de Antioquia | Universidad de Antioquia | Ciuoderis,K.A., Moreno-Lopez,I., Usuga-Restrepo,J.A., Perez-Restrepo,L.S., Florez,D.Y., Hernandez-Ortiz,J.P., Cloherty,G.A. and Osorio,J.E. |
| EPI_ISL_18474691 | Universidad de Antioquia | Universidad de Antioquia | Ciuoderis,K.A., Usuga,J., Moreno-Lopez,I., Perez-Restrepo,L.S., Florez,D.Y., Hernandez-Ortiz,J.P., Cloherty,G.A. and Osorio,J.E. |
| EPI_ISL_18474692 | Universidad de Antioquia | Universidad de Antioquia | Ciuoderis,K., Usuga,J., Moreno-Lopez,I., Perez-Restrepo,L., Florez,D., Hernandez-Ortiz,J., Cloherty,G. and Osorio,J. |
| EPI_ISL_18474693, EPI_ISL_18474694 | Universidad de Antioquia | Universidad de Antioquia | Ciuoderis,K.A., Moreno-Lopez,I., Usuga-Restrepo,J.A., Perez-Restrepo,L.S., Florez,D.Y., Hernandez-Ortiz,J.P., Cloherty,G.A. and Osorio,J.E. |
| EPI_ISL_18474695, EPI_ISL_18474696, EPI_ISL_18474697, EPI_ISL_18474698, EPI_ISL_18474699, EPI_ISL_18474700, EPI_ISL_18474701 | Universidad de Antioquia | Universidad de Antioquia | Ciuoderis,K.A., Usuga,J., Moreno-Lopez,I., Perez-Restrepo,L.S., Florez,D.Y., Hernandez-Ortiz,J.P., Cloherty,G.A. and Osorio,J.E. |
| EPI_ISL_18474702 | Universidad de Antioquia | Universidad de Antioquia | Ciuoderis,K.A., Moreno-Lopez,I., Usuga-Restrepo,J.A., Perez-Restrepo,L.S., Florez,D.Y., Hernandez-Ortiz,J.P., Cloherty,G.A. and Osorio,J.E. |
| EPI_ISL_18474703, EPI_ISL_18474704, EPI_ISL_18474705, EPI_ISL_18474706, EPI_ISL_18474707, EPI_ISL_18474708 | Universidad de Antioquia | Universidad de Antioquia | Ciuoderis,K.A., Usuga,J., Moreno-Lopez,I., Perez-Restrepo,L.S., Florez,D.Y., Hernandez-Ortiz,J.P., Cloherty,G.A. and Osorio,J.E. |
| EPI_ISL_18474709 | Universidad de Antioquia | Universidad de Antioquia | Ciuoderis,K.A., Moreno-Lopez,I., Usuga-Restrepo,J.A., Perez-Restrepo,L.S., Florez,D.Y., Hernandez-Ortiz,J.P., Cloherty,G.A. and Osorio,J.E. |
| EPI_ISL_18474710 | Universidad de Antioquia | Universidad de Antioquia | Ciuoderis,K.A., Usuga,J., Moreno-Lopez,I., Perez-Restrepo,L.S., Florez,D.Y., Hernandez-Ortiz,J.P., Cloherty,G.A. and Osorio,J.E. |
| EPI_ISL_18474711, EPI_ISL_18474712, EPI_ISL_18474713, EPI_ISL_18474714, EPI_ISL_18474715 | Universidad de Antioquia | Universidad de Antioquia | Ciuoderis,K.A., Moreno-Lopez,I., Usuga-Restrepo,J.A., Perez-Restrepo,L.S., Florez,D.Y., Hernandez-Ortiz,J.P., Cloherty,G.A. and Osorio,J.E. |
| EPI_ISL_18474716, EPI_ISL_18474717, EPI_ISL_18474718 | Universidad de Antioquia | Universidad de Antioquia | Ciuoderis,K.A., Usuga,J., Moreno-Lopez,I., Perez-Restrepo,L.S., Florez,D.Y., Hernandez-Ortiz,J.P., Cloherty,G.A. and Osorio,J.E. |
| EPI_ISL_18474719 | Universidad de Antioquia | Universidad de Antioquia | Ciuoderis,K.A., Moreno-Lopez,I., Usuga-Restrepo,J.A., Perez-Restrepo,L.S., Florez,D.Y., Hernandez-Ortiz,J.P., Cloherty,G.A. and Osorio,J.E. |
| EPI_ISL_18474720, EPI_ISL_18474721, EPI_ISL_18474722, EPI_ISL_18474723, EPI_ISL_18474724 | Universidad de Antioquia | Universidad de Antioquia | Ciuoderis,K.A., Usuga,J., Moreno-Lopez,I., Perez-Restrepo,L.S., Florez,D.Y., Hernandez-Ortiz,J.P., Cloherty,G.A. and Osorio,J.E. |
| EPI_ISL_18474725 | Universidad de Antioquia | Universidad de Antioquia | Ciuoderis,K.A., Moreno-Lopez,I., Usuga-Restrepo,J.A., Perez-Restrepo,L.S., Florez,D.Y., Hernandez-Ortiz,J.P., Cloherty,G.A. and Osorio,J.E. |
| EPI_ISL_18474726, EPI_ISL_18474727, EPI_ISL_18474728, EPI_ISL_18474729 | Universidad de Antioquia | Universidad de Antioquia | Ciuoderis,K.A., Usuga,J., Moreno-Lopez,I., Perez-Restrepo,L.S., Florez,D.Y., Hernandez-Ortiz,J.P., Cloherty,G.A. and Osorio,J.E. |
| EPI_ISL_18474730, EPI_ISL_18474736, EPI_ISL_18474752, EPI_ISL_18474770, EPI_ISL_18474787, EPI_ISL_18474814, EPI_ISL_18474819, EPI_ISL_18474834 | unknown | unknown | Fonseca,V., Giovanetti,M., Xavier,J., Oliveira,C., Lima,M., Barbosa,E., Guimaraes,N., Adelino,T., Fritsch,H., Mendonca,M., Bispo Filippis,A. and Alcantara,L. |
| EPI_ISL_18560746, EPI_ISL_18560747, EPI_ISL_18560748 | NC - Biological Sciences,NC - Redeemer's University | NC - Biological Sciences,NC - Redeemer's University | Onoja,B.A., Oguzie,J.U., George,U.E., Asoh,K.E., Ajayi,P., Omofaye,T.F., Igeleke,I.O., Eromon,P., Harouna,S., Parker,E., Adeniji,A.J. and Happi,C.T. |
| EPI_ISL_18579214, EPI_ISL_18579215, EPI_ISL_18579216, EPI_ISL_18579217, EPI_ISL_18579218, EPI_ISL_18579219, EPI_ISL_18579220, EPI_ISL_18579221, EPI_ISL_18579222, EPI_ISL_18579223, EPI_ISL_18579224, EPI_ISL_18579225, EPI_ISL_18579226, EPI_ISL_18579227, EPI_ISL_18579228, EPI_ISL_18579229, EPI_ISL_18579230, EPI_ISL_18579231, EPI_ISL_18579232, EPI_ISL_18579233, EPI_ISL_18579234, EPI_ISL_18579235, EPI_ISL_18579236, EPI_ISL_18579237, EPI_ISL_18579238, EPI_ISL_18579239, EPI_ISL_18579240, EPI_ISL_18579241, EPI_ISL_18579242, EPI_ISL_18579243, EPI_ISL_18579244, EPI_ISL_18579245, EPI_ISL_18579246, EPI_ISL_18579247, EPI_ISL_18579248, EPI_ISL_18579249, EPI_ISL_18579250, EPI_ISL_18579251, EPI_ISL_18579252, EPI_ISL_18579253, EPI_ISL_18579254, EPI_ISL_18579255, EPI_ISL_18579256, EPI_ISL_18579257, EPI_ISL_18579258, EPI_ISL_18579259, EPI_ISL_18579260, EPI_ISL_18579261, EPI_ISL_18579262, EPI_ISL_18579263, EPI_ISL_18579264, EPI_ISL_18579265, EPI_ISL_18579266, EPI_ISL_18579267, EPI_ISL_18579268, EPI_ISL_18579269, EPI_ISL_18579270, EPI_ISL_18579271, EPI_ISL_18579272, EPI_ISL_18579273, EPI_ISL_18579274, EPI_ISL_18579275, EPI_ISL_18579276, EPI_ISL_18579277, EPI_ISL_18579278, EPI_ISL_18579279, EPI_ISL_18579280 | | | |
| see above | University of Western Australia | University of Western Australia | Harapan,H., Ernst,T., Panta,K., McCarthy,S., Michie,A., Sasmono,R.T., Smith,D.W., Effler,P.V. and Imrie,A. |
| EPI_ISL_18607159, EPI_ISL_18607160, EPI_ISL_18607161, EPI_ISL_18607162, EPI_ISL_18607163, EPI_ISL_18607164, EPI_ISL_18607165, EPI_ISL_18607166, EPI_ISL_18607167 | Oswaldo Cruz Foundation, Instituto Carlos Chagas | Oswaldo Cruz Foundation, Instituto Carlos Chagas | de Arruda,T.B., Bavia,L., Mosimann,A.L.P., Aoki,M.N., Sarzi,M.L., Conchon-Costa,I., Wowk,P.F., Duarte Dos Santos,C.N., Pavanelli,W.R., Silveira,G.F. and Bordignon,J. |
| EPI_ISL_18689287, EPI_ISL_18689288, EPI_ISL_18689289, EPI_ISL_18689290, EPI_ISL_18689291, EPI_ISL_18689292, EPI_ISL_18689293, EPI_ISL_18689294, EPI_ISL_18689295, EPI_ISL_18689296, EPI_ISL_18689297, EPI_ISL_18689298, EPI_ISL_18689299, EPI_ISL_18689300, EPI_ISL_18689301, EPI_ISL_18689302, EPI_ISL_18689303, EPI_ISL_18689304, EPI_ISL_18689305, EPI_ISL_18689306, EPI_ISL_18689307, EPI_ISL_18689308, EPI_ISL_18689309, EPI_ISL_18689310, EPI_ISL_18689311, EPI_ISL_18689312, EPI_ISL_18689313, EPI_ISL_18689314, EPI_ISL_18689315, EPI_ISL_18689316, EPI_ISL_18689317, EPI_ISL_18689318, EPI_ISL_18689319, EPI_ISL_18689320, EPI_ISL_18689321, EPI_ISL_18689322, EPI_ISL_18689323, EPI_ISL_18689324, EPI_ISL_18689325, EPI_ISL_18689326, EPI_ISL_18689327, EPI_ISL_18689328, EPI_ISL_18689329, EPI_ISL_18689330, EPI_ISL_18689331, EPI_ISL_18689332, EPI_ISL_18689333, EPI_ISL_18689334, EPI_ISL_18689335, EPI_ISL_18689336, EPI_ISL_18689337, EPI_ISL_18689338, EPI_ISL_18689339, EPI_ISL_18689340, EPI_ISL_18689341, EPI_ISL_18689342, EPI_ISL_18689343, EPI_ISL_18689344, EPI_ISL_18689345, EPI_ISL_18689346, EPI_ISL_18689347, EPI_ISL_18689348 | | | |

| | | | |
|---|---|---|---|
| see above | Instituto de Investigaciones en Ciencias de la Salud, Universidad Nacional de Asuncion, Production Department | Instituto de Investigaciones en Ciencias de la Salud, Universidad Nacional de Asuncion, Production Department | Rojas,A., Shen,J., Cardozo,F., Bernal,C., Caballero,O., Ping,S., Key,A., Haider,A., Stittleburg,V., de Guillen,Y., Langjahr,P., Acosta,M.E., Aria,L., Mendoza,L., Paez,M., Von-Horoch,M., Luraschi,P., Cabral,S., Sanchez,M.C., Torres,A., Pinsky,B.A., Piantadosi,A. and Waggoner,J. |
| EPI_ISL_18689389, EPI_ISL_18689392, EPI_ISL_18689398, EPI_ISL_18689404, EPI_ISL_18689409, EPI_ISL_18689417, EPI_ISL_18689422, EPI_ISL_18689424 | University of Sao Paulo | University of Sao Paulo | Souza,C. and Romano,C. |
| EPI_ISL_18708170, EPI_ISL_18708171, EPI_ISL_18708172, EPI_ISL_18708173, EPI_ISL_18708174, EPI_ISL_18708175, EPI_ISL_18708176, EPI_ISL_18708177, EPI_ISL_18708178, EPI_ISL_18708179, EPI_ISL_18708180, EPI_ISL_18708181, EPI_ISL_18708182 | | | |
| see above | University of Bengkulu, Microbiology | University of Bengkulu, Microbiology | Nugraheni,E., Syahrurachman,A., Nainggolan,L., Sipriyadi,S., Sariyanti,M., Nugroho,M.A., Fithriyah,F. and Dewi,B.E. |
| EPI_ISL_18708233, EPI_ISL_18708234, EPI_ISL_18708235, EPI_ISL_18708236, EPI_ISL_18708237 | Viral Immunology Laboratory, Oswaldo Cruz Institute | Viral Immunology Laboratory, Oswaldo Cruz Institute | de Mendonca,M.C.L., Oliveira,C., Rodrigues,C.D.S., Santos,C.S., Sampaio,S.A., Fabri,A., de Filippis,A.M.B. and de Bruycker-Nogueira,F. |
| EPI_ISL_18713032, EPI_ISL_18713033, EPI_ISL_18713034, EPI_ISL_18713035 | Evandro Chagas Institute, Department of Arbovirology and Hemorrhagic Fevers | Evandro Chagas Institute, Department of Arbovirology and Hemorrhagic Fevers | Amorim,M.T., Hernandez,L.H.A., Naveca,F.G., Essashika Prazeres,I.T., Wanzeller,A.L.M., Silva,E.V.P.D., Casseb,L.M.N., Silva,F.S.D., da Silva,S.P., Nunes,B.T.D. and Cruz,A.C.R. |
| EPI_ISL_18714550, EPI_ISL_18714551, EPI_ISL_18714552, EPI_ISL_18714553, EPI_ISL_18714554, EPI_ISL_18714555, EPI_ISL_18714556, EPI_ISL_18714557, EPI_ISL_18714558, EPI_ISL_18714559, EPI_ISL_18714560, EPI_ISL_18714561, EPI_ISL_18714562, EPI_ISL_18714563, EPI_ISL_18714564, EPI_ISL_18714565, EPI_ISL_18714566, EPI_ISL_18714567, EPI_ISL_18714568, EPI_ISL_18714569, EPI_ISL_18714570, EPI_ISL_18714571, EPI_ISL_18714572, EPI_ISL_18714573, EPI_ISL_18714574, EPI_ISL_18714575, EPI_ISL_18714576, EPI_ISL_18714577, EPI_ISL_18714578, EPI_ISL_18714579, EPI_ISL_18714580, EPI_ISL_18714581, EPI_ISL_18714582, EPI_ISL_18714583, EPI_ISL_18714584, EPI_ISL_18714585, EPI_ISL_18714586, EPI_ISL_18714587, EPI_ISL_18714588, EPI_ISL_18714589, EPI_ISL_18714590, EPI_ISL_18714591, EPI_ISL_18714592, EPI_ISL_18714593, EPI_ISL_18714594, EPI_ISL_18714595, EPI_ISL_18714596, EPI_ISL_18714597, EPI_ISL_18714598, EPI_ISL_18714599, EPI_ISL_18714600, EPI_ISL_18714601, EPI_ISL_18714602, EPI_ISL_18714603, EPI_ISL_18714604, EPI_ISL_18714605, EPI_ISL_18714606, EPI_ISL_18714607, EPI_ISL_18714608, EPI_ISL_18714609, EPI_ISL_18714610, EPI_ISL_18714611, EPI_ISL_18714612, EPI_ISL_18714613, EPI_ISL_18714614, EPI_ISL_18714615, EPI_ISL_18714616, EPI_ISL_18714617, EPI_ISL_18714618, EPI_ISL_18714619, EPI_ISL_18714620, EPI_ISL_18714621, EPI_ISL_18714622, EPI_ISL_18714623, EPI_ISL_18714624, EPI_ISL_18714625, EPI_ISL_18714626, EPI_ISL_18714627, EPI_ISL_18714628, EPI_ISL_18714629, EPI_ISL_18714630, EPI_ISL_18714631, EPI_ISL_18714632, EPI_ISL_18714633, EPI_ISL_18714634, EPI_ISL_18714635, EPI_ISL_18714636, EPI_ISL_18714637, EPI_ISL_18714638, EPI_ISL_18714639, EPI_ISL_18714640, EPI_ISL_18714641, EPI_ISL_18714642, EPI_ISL_18714643, EPI_ISL_18714644, EPI_ISL_18714645, EPI_ISL_18714646, EPI_ISL_18714647, EPI_ISL_18714648, EPI_ISL_18714649, EPI_ISL_18714650, EPI_ISL_18714651, EPI_ISL_18714652 | | | |
| see above | Undisclosed Laboratory in Brazil for the Health Emergency Information and Risk Assessment (HIM) Unit of PAHO | Undisclosed Laboratory in Brazil for the Health Emergency Information and Risk Assessment (HIM) Unit of PAHO | Fonseca,V., Giovanetti,M., Xavier,J., Oliveira,C., Lima,M., Barbosa,E., Guimaraes,N., Adelino,T., Fritsch,H., Mendonca,M., Bispo Filippis,A. and Alcantara,L. |
| EPI_ISL_18737456, EPI_ISL_18737457, EPI_ISL_18737458, EPI_ISL_18737459, EPI_ISL_18737460, EPI_ISL_18737461, EPI_ISL_18737462, EPI_ISL_18737464, EPI_ISL_18737465, EPI_ISL_18737466, EPI_ISL_18737467, EPI_ISL_18737468, EPI_ISL_18737469, EPI_ISL_18737470, EPI_ISL_18737471, EPI_ISL_18737473, EPI_ISL_18737474, EPI_ISL_18737475, EPI_ISL_18737476, EPI_ISL_18737477, EPI_ISL_18737478, EPI_ISL_18737479, EPI_ISL_18737480, EPI_ISL_18737482, EPI_ISL_18737483, EPI_ISL_18737484, EPI_ISL_18737485, EPI_ISL_18737486, EPI_ISL_18737487, EPI_ISL_18737488, EPI_ISL_18737489, EPI_ISL_18737491, EPI_ISL_18737492, EPI_ISL_18737493, EPI_ISL_18737494, EPI_ISL_18737495, EPI_ISL_18737496, EPI_ISL_18737497, EPI_ISL_18737499, EPI_ISL_18737500, EPI_ISL_18737501, EPI_ISL_18737502, EPI_ISL_18737503, EPI_ISL_18737504, EPI_ISL_18737505, EPI_ISL_18737506, EPI_ISL_18737508, EPI_ISL_18737509, EPI_ISL_18737510, EPI_ISL_18737511, EPI_ISL_18737512, EPI_ISL_18737513, EPI_ISL_18737514, EPI_ISL_18737515, EPI_ISL_18737517, EPI_ISL_18737518 | | | |
| see above | Center for Vectors and Infectious Diseases Research (CEVDI), National Health Institute Doutor Ricardo Jorge, IP (INSA) | Center for Vectors and Infectious Diseases Research (CEVDI), National Health Institute Doutor Ricardo Jorge, IP (INSA) | Ze-Ze,L. and Alves,M.J. |
| EPI_ISL_18742163, EPI_ISL_18742164, EPI_ISL_18742165 | Ministry of Health, Central Virology Laboratory | Ministry of Health, Central Virology Laboratory | Zuckerman,N.S., Schwartz,E., Pandey,P., Erster,O., Halpern,O., Bucris,E., Morad-Eliyahu,H., Wax,M. and Lustig,Y. |
| EPI_ISL_18951576, EPI_ISL_18951577, EPI_ISL_18951578, EPI_ISL_18951579, EPI_ISL_18951580, EPI_ISL_18951581, EPI_ISL_18951582, EPI_ISL_18951583, EPI_ISL_18951584, EPI_ISL_18951585, EPI_ISL_18951586, EPI_ISL_18951587, EPI_ISL_18951588, EPI_ISL_18951589, EPI_ISL_18951590, EPI_ISL_18951591, EPI_ISL_18951592, EPI_ISL_18951593, EPI_ISL_18951594, EPI_ISL_18951595, EPI_ISL_18951596, EPI_ISL_18951597, EPI_ISL_18951598, EPI_ISL_18951599, EPI_ISL_18951600, EPI_ISL_18951601, EPI_ISL_18951602, EPI_ISL_18951603 | | | |
| see above | Institute of Tropical Medicine, Nagasaki University | Institute of Tropical Medicine, Nagasaki University | Nabeshima,T., Thi Thanh,N.N., Matsuzaki,A., Xayavong,D., Fraenkel,S., Qiang,X., Nwe,K.M., Palma,J.C.B., Tun,M.M.N., Thuy Duong,N.T., Mai,L.T.Q., Morita,K., Hasebe,F. and Takamatsu,Y. |
| EPI_ISL_18951604 | Institute of Tropical Medicine, Nagasaki University | | Nabeshima,T., Thi Thanh,N.N., Matsuzaki,A., Xayavong,D., Fraenkel,S., Qiang,X., Nwe,K.M., Palma,J.C.B., Tun,M.M.N., Thuy Duong,N.T., Mai,L.T.Q., Morita,K., Hasebe,F. and Takamatsu,Y. |
| EPI_ISL_18951605, EPI_ISL_18951606, EPI_ISL_18951607, EPI_ISL_18951608, EPI_ISL_18951609, EPI_ISL_18951610, EPI_ISL_18951611, EPI_ISL_18951612, EPI_ISL_18951613, EPI_ISL_18951614, EPI_ISL_18951615, EPI_ISL_18951616, EPI_ISL_18951617, EPI_ISL_18951618, EPI_ISL_18951619, EPI_ISL_18951620, EPI_ISL_18951621, EPI_ISL_18951622, EPI_ISL_18951623, EPI_ISL_18951624, EPI_ISL_18951625, EPI_ISL_18951626, EPI_ISL_18951627, EPI_ISL_18951628, EPI_ISL_18951629, EPI_ISL_18951630, EPI_ISL_18951631, EPI_ISL_18951632, EPI_ISL_18951633, EPI_ISL_18951634, EPI_ISL_18951635, EPI_ISL_18951636, EPI_ISL_18951637, EPI_ISL_18951638, EPI_ISL_18951639, EPI_ISL_18951640, EPI_ISL_18951641, EPI_ISL_18951642, EPI_ISL_18951643, EPI_ISL_18951644, EPI_ISL_18951645, EPI_ISL_18951646, EPI_ISL_18951647, EPI_ISL_18951648, EPI_ISL_18951649, EPI_ISL_18951650, EPI_ISL_18951651, EPI_ISL_18951652, EPI_ISL_18951653 | | | |
| see above | Institute of Tropical Medicine, Nagasaki University | Institute of Tropical Medicine, Nagasaki University | Nabeshima,T., Thi Thanh,N.N., Matsuzaki,A., Xayavong,D., Fraenkel,S., Qiang,X., Nwe,K.M., Palma,J.C.B., Tun,M.M.N., Thuy Duong,N.T., Mai,L.T.Q., Morita,K., Hasebe,F. and Takamatsu,Y. |
| EPI_ISL_18956187 | Evandro Chagas Institute, Department of Arbovirology and Hemorrhagic Fevers | Evandro Chagas Institute, Department of Arbovirology and Hemorrhagic Fevers | Amorim,M.T., Silva,S.P., Hernandez,L.H. and Cruz,A.C.R. |
| EPI_ISL_19007009, EPI_ISL_19007010, EPI_ISL_19007011, EPI_ISL_19007012, EPI_ISL_19007013, EPI_ISL_19007014, EPI_ISL_19007015, EPI_ISL_19007016, EPI_ISL_19007017, EPI_ISL_19007018, EPI_ISL_19007019, EPI_ISL_19007020, EPI_ISL_19007021, EPI_ISL_19007022, EPI_ISL_19007023, EPI_ISL_19007024, EPI_ISL_19007025, EPI_ISL_19007026, EPI_ISL_19007027, EPI_ISL_19007028, EPI_ISL_19007029, EPI_ISL_19007030, EPI_ISL_19007031, EPI_ISL_19007032, EPI_ISL_19007033, EPI_ISL_19007034, EPI_ISL_19007035, EPI_ISL_19007036, EPI_ISL_19007037, EPI_ISL_19007038, EPI_ISL_19007039, EPI_ISL_19007040, EPI_ISL_19007041, EPI_ISL_19007042, EPI_ISL_19007043, EPI_ISL_19007044, EPI_ISL_19007045, EPI_ISL_19007046, EPI_ISL_19007047, EPI_ISL_19007048, EPI_ISL_19007049, EPI_ISL_19007050, EPI_ISL_19007051, EPI_ISL_19007052, EPI_ISL_19007053, EPI_ISL_19007054, EPI_ISL_19007055, EPI_ISL_19007056, EPI_ISL_19007057, EPI_ISL_19007058, EPI_ISL_19007059, EPI_ISL_19007060, EPI_ISL_19007061, EPI_ISL_19007062, EPI_ISL_19007063, EPI_ISL_19007064, EPI_ISL_19007065, EPI_ISL_19007066, EPI_ISL_19007067, EPI_ISL_19007068, EPI_ISL_19007069, EPI_ISL_19007070, EPI_ISL_19007071, EPI_ISL_19007072, EPI_ISL_19007073, EPI_ISL_19007074, EPI_ISL_19007075, EPI_ISL_19007076, EPI_ISL_19007077, EPI_ISL_19007078, EPI_ISL_19007079, EPI_ISL_19007080, EPI_ISL_19007081, EPI_ISL_19007082, EPI_ISL_19007083, EPI_ISL_19007084, EPI_ISL_19007085, EPI_ISL_19007086, EPI_ISL_19007087, EPI_ISL_19007088, EPI_ISL_19007089, EPI_ISL_19007090, EPI_ISL_19007091, EPI_ISL_19007092, EPI_ISL_19007093, EPI_ISL_19007094, EPI_ISL_19007095, EPI_ISL_19007096, EPI_ISL_19007097, EPI_ISL_19007098, EPI_ISL_19007099, EPI_ISL_19007100, EPI_ISL_19007101, EPI_ISL_19007102, EPI_ISL_19007103, EPI_ISL_19007104, EPI_ISL_19007105, EPI_ISL_19007106, EPI_ISL_19007107, EPI_ISL_19007108, EPI_ISL_19007109, EPI_ISL_19007110, EPI_ISL_19007111, EPI_ISL_19007112, EPI_ISL_19007113, EPI_ISL_19007114, EPI_ISL_19007115, EPI_ISL_19007116, EPI_ISL_19007117, EPI_ISL_19007118, EPI_ISL_19007119, EPI_ISL_19007120, EPI_ISL_19007121, EPI_ISL_19007122, EPI_ISL_19007123, EPI_ISL_19007124, EPI_ISL_19007125, EPI_ISL_19007126, EPI_ISL_19007127, EPI_ISL_19007128, EPI_ISL_19007129, EPI_ISL_19007130, EPI_ISL_19007131, EPI_ISL_19007132, EPI_ISL_19007133, EPI_ISL_19007134, EPI_ISL_19007135, EPI_ISL_19007136, EPI_ISL_19007137, EPI_ISL_19007138, EPI_ISL_19007139, EPI_ISL_19007140, EPI_ISL_19007141, EPI_ISL_19007142, EPI_ISL_19007143, EPI_ISL_19007144, EPI_ISL_19007145, EPI_ISL_19007146, EPI_ISL_19007147, EPI_ISL_19007148, EPI_ISL_19007149, EPI_ISL_19007150, EPI_ISL_19007151, EPI_ISL_19007152, EPI_ISL_19007153, EPI_ISL_19007154, EPI_ISL_19007155, EPI_ISL_19007156, EPI_ISL_19007157, EPI_ISL_19007158, EPI_ISL_19007159, EPI_ISL_19007160, EPI_ISL_19007161, EPI_ISL_19007162, EPI_ISL_19007163, EPI_ISL_19007164, EPI_ISL_19007165, EPI_ISL_19007166, EPI_ISL_19007167, EPI_ISL_19007168, EPI_ISL_19007169, EPI_ISL_19007170, EPI_ISL_19007171, EPI_ISL_19007172, EPI_ISL_19007173, EPI_ISL_19007174, EPI_ISL_19007175, EPI_ISL_19007176, EPI_ISL_19007177, EPI_ISL_19007178, EPI_ISL_19007179, EPI_ISL_19007180, EPI_ISL_19007181, EPI_ISL_19007182, EPI_ISL_19007183, EPI_ISL_19007184, EPI_ISL_19007185, EPI_ISL_19007186, EPI_ISL_19007187, EPI_ISL_19007188, EPI_ISL_19007189, EPI_ISL_19007190, EPI_ISL_19007191, EPI_ISL_19007192, EPI_ISL_19007193, EPI_ISL_19007194, EPI_ISL_19007195, EPI_ISL_19007196, EPI_ISL_19007197, EPI_ISL_19007198, EPI_ISL_19007199, EPI_ISL_19007200, EPI_ISL_19007201, EPI_ISL_19007202, EPI_ISL_19007203, EPI_ISL_19007204, EPI_ISL_19007205, EPI_ISL_19007206, EPI_ISL_19007207, EPI_ISL_19007208, EPI_ISL_19007209, EPI_ISL_19007210, EPI_ISL_19007211, EPI_ISL_19007212, EPI_ISL_19007213, EPI_ISL_19007214, EPI_ISL_19007215, EPI_ISL_19007216, EPI_ISL_19007217, EPI_ISL_19007218, EPI_ISL_19007219, EPI_ISL_19007220, EPI_ISL_19007221, EPI_ISL_19007222, EPI_ISL_19007223, EPI_ISL_19007224, EPI_ISL_19007225, EPI_ISL_19007226, EPI_ISL_19007227, EPI_ISL_19007228, EPI_ISL_19007229, EPI_ISL_19007230, EPI_ISL_19007231, EPI_ISL_19007232, EPI_ISL_19007233, EPI_ISL_19007234, EPI_ISL_19007235, EPI_ISL_19007236, EPI_ISL_19007237, EPI_ISL_19007238, EPI_ISL_19007239, EPI_ISL_19007240, EPI_ISL_19007241, EPI_ISL_19007242, EPI_ISL_19007243, EPI_ISL_19007244, EPI_ISL_19007245, EPI_ISL_19007246, EPI_ISL_19007247, EPI_ISL_19007248, EPI_ISL_19007249, EPI_ISL_19007250, EPI_ISL_19007251, EPI_ISL_19007252, EPI_ISL_19007253, EPI_ISL_19007254, EPI_ISL_19007255, EPI_ISL_19007256, EPI_ISL_19007257, EPI_ISL_19007258, EPI_ISL_19007259, EPI_ISL_19007260, EPI_ISL_19007261, EPI_ISL_19007262, EPI_ISL_19007263, EPI_ISL_19007264, EPI_ISL_19007265, EPI_ISL_19007266, EPI_ISL_19007267, EPI_ISL_19007268, EPI_ISL_19007269, EPI_ISL_19007270, EPI_ISL_19007271, EPI_ISL_19007272, EPI_ISL_19007273, EPI_ISL_19007274, EPI_ISL_19007275, EPI_ISL_19007276, EPI_ISL_19007277, EPI_ISL_19007278, EPI_ISL_19007279, EPI_ISL_19007280, EPI_ISL_19007281, EPI_ISL_19007282, EPI_ISL_19007283, EPI_ISL_19007284, EPI_ISL_19007285, EPI_ISL_19007286, EPI_ISL_19007287, EPI_ISL_19007288, EPI_ISL_19007289, EPI_ISL_19007290, EPI_ISL_19007291, EPI_ISL_19007292, EPI_ISL_19007293, EPI_ISL_19007294, EPI_ISL_19007295, EPI_ISL_19007296, EPI_ISL_19007297, EPI_ISL_19007298, EPI_ISL_19007299, EPI_ISL_19007300, EPI_ISL_19007301, EPI_ISL_19007302, EPI_ISL_19007303, EPI_ISL_19007304, EPI_ISL_19007305, EPI_ISL_19007306, EPI_ISL_19007307, EPI_ISL_19007308, EPI_ISL_19007309, EPI_ISL_19007310, EPI_ISL_19007311, EPI_ISL_19007312, EPI_ISL_19007313, EPI_ISL_19007314, EPI_ISL_19007315, EPI_ISL_19007316, EPI_ISL_19007317, EPI_ISL_19007318, EPI_ISL_19007319, EPI_ISL_19007320, EPI_ISL_19007321, EPI_ISL_19007322, EPI_ISL_19007323, EPI_ISL_19007324, EPI_ISL_19007325, EPI_ISL_19007326, EPI_ISL_19007327, EPI_ISL_19007328, EPI_ISL_19007329, EPI_ISL_19007330, EPI_ISL_19007331, EPI_ISL_19007332, EPI_ISL_19007333, EPI_ISL_19007334, EPI_ISL_19007335, EPI_ISL_19007336, EPI_ISL_19007337, EPI_ISL_19007338, EPI_ISL_19007339, EPI_ISL_19007340, EPI_ISL_19007341, EPI_ISL_19007342, EPI_ISL_19007343, EPI_ISL_19007344, EPI_ISL_19007345, EPI_ISL_19007346, EPI_ISL_19007347, EPI_ISL_19007348, EPI_ISL_19007349, EPI_ISL_19007350, EPI_ISL_19007351, EPI_ISL_19007352, EPI_ISL_19007353, EPI_ISL_19007354, EPI_ISL_19007355, EPI_ISL_19007356, EPI_ISL_19007357, EPI_ISL_19007358, EPI_ISL_19007359, EPI_ISL_19007360, EPI_ISL_19007361, EPI_ISL_19007362, EPI_ISL_19007363, EPI_ISL_19007364, EPI_ISL_19007365, EPI_ISL_19007366, EPI_ISL_19007367, EPI_ISL_19007368, EPI_ISL_19007369, EPI_ISL_19007370, EPI_ISL_19007371, EPI_ISL_19007372, EPI_ISL_19007373, EPI_ISL_19007374, EPI_ISL_19007375, EPI_ISL_19007376, EPI_ISL_19007377, EPI_ISL_19007378, EPI_ISL_19007379, EPI_ISL_19007380, EPI_ISL_19007381, EPI_ISL_19007382, EPI_ISL_19007383, EPI_ISL_19007384, EPI_ISL_19007385, EPI_ISL_19007386, EPI_ISL_19007387, EPI_ISL_19007388, EPI_ISL_19007389, EPI_ISL_19007390, EPI_ISL_19007391, EPI_ISL_19007392, EPI_ISL_19007393, EPI_ISL_19007394, EPI_ISL_19007395, EPI_ISL_19007396, EPI_ISL_19007397, EPI_ISL_19007398, EPI_ISL_19007399, EPI_ISL_19007400, EPI_ISL_19007401, EPI_ISL_19007402, EPI_ISL_19007403, EPI_ISL_19007404, EPI_ISL_19007405, EPI_ISL_19007406, EPI_ISL_19007407, EPI_ISL_19007408, EPI_ISL_19007409, EPI_ISL_19007410, EPI_ISL_19007411, EPI_ISL_19007412, EPI_ISL_19007413, EPI_ISL_19007414, EPI_ISL_19007415, EPI_ISL_19007416, EPI_ISL_19007417, EPI_ISL_19007418, EPI_ISL_19007419, EPI_ISL_19007420, EPI_ISL_19007421, EPI_ISL_19007422, EPI_ISL_19007423, EPI_ISL_19007424, EPI_ISL_19007425, EPI_ISL_19007426, EPI_ISL_19007427, EPI_ISL_19007428, EPI_ISL_19007429, EPI_ISL_19007430, EPI_ISL_19007431, EPI_ISL_19007432, EPI_ISL_19007433, EPI_ISL_19007434, EPI_ISL_19007435, EPI_ISL_19007436, EPI_ISL_19007437, EPI_ISL_19007438, EPI_ISL_19007439, EPI_ISL_19007440, EPI_ISL_19007441, EPI_ISL_19007442, EPI_ISL_19007443, EPI_ISL_19007444, EPI_ISL_19007445, EPI_ISL_19007446, EPI_ISL_19007447, EPI_ISL_19007448, EPI_ISL_19007449, EPI_ISL_19007450, EPI_ISL_19007451, EPI_ISL_19007452, EPI_ISL_19007453, EPI_ISL_19007454, EPI_ISL_19007455, EPI_ISL_19007456, EPI_ISL_19007457, EPI_ISL_19007458, EPI_ISL_19007459, EPI_ISL_19007460, EPI_ISL_19007461, EPI_ISL_19007462, EPI_ISL_19007463, EPI_ISL_19007464, EPI_ISL_19007465, EPI_ISL_19007466, EPI_ISL_19007467, EPI_ISL_19007468, EPI_ISL_19007469, EPI_ISL_19007470, EPI_ISL_19007471, EPI_ISL_19007472, EPI_ISL_19007473, EPI_ISL_19007474, EPI_ISL_19007475, EPI_ISL_19007476, EPI_ISL_19007477, EPI_ISL_19007478, EPI_ISL_19007479, EPI_ISL_19007480, EPI_ISL_19007481, EPI_ISL_19007482, EPI_ISL_19007483, EPI_ISL_19007484, EPI_ISL_19007485, EPI_ISL_19007486, EPI_ISL_19007487, EPI_ISL_19007488, EPI_ISL_19007489, EPI_ISL_19007490, EPI_ISL_19007491, EPI_ISL_19007492, EPI_ISL_19007493, EPI_ISL_19007494, EPI_ISL_19007495, EPI_ISL_19007496, EPI_ISL_19007497, EPI_ISL_19007498, EPI_ISL_19007499, EPI_ISL_19007500, EPI_ISL_19007501, EPI_ISL_19007502, EPI_ISL_19007503, EPI_ISL_19007504, EPI_ISL_19007505, EPI_ISL_19007506, EPI_ISL_19007507, EPI_ISL_19007508, EPI_ISL_19007509, EPI_ISL_19007510, EPI_ISL_19007511, EPI_ISL_19007512, EPI_ISL_19007513, EPI_ISL_19007514, EPI_ISL_19007515, EPI_ISL_19007516, EPI_ISL_19007517, EPI_ISL_19007518, EPI_ISL_19007519, EPI_ISL_19007520, EPI_ISL_19007521, EPI_ISL_19007522, EPI_ISL_19007523, EPI_ISL_19007524, EPI_ISL_19007525, EPI_ISL_19007526, EPI_ISL_19007527, EPI_ISL_19007528, EPI_ISL_19007529, EPI_ISL_19007530, EPI_ISL_19007531, EPI_ISL_19007532, EPI_ISL_19007533, EPI_ISL_19007534, EPI_ISL_19007535, EPI_ISL_19007536, EPI_ISL_19007537, EPI_ISL_19007538, EPI_ISL_19007539, EPI_ISL_19007540, EPI_ISL_19007541, EPI_ISL_19007542, EPI_ISL_19007543, EPI_ISL_19007544, EPI_ISL_19007545, EPI_ISL_19007546, EPI_ISL_19007547, EPI_ISL_19007548, EPI_ISL_19007549, EPI_ISL_19007550, EPI_ISL_19007551, EPI_ISL_19007552, EPI_ISL_19007553, EPI_ISL_19007554, EPI_ISL_19007555, EPI_ISL_19007556, EPI_ISL_19007557, EPI_ISL_19007558, EPI_ISL_19007559, EPI_ISL_19007560, EPI_ISL_19007561, EPI_ISL_19007562, EPI_ISL_19007563, EPI_ISL_19007564, EPI_ISL_19007565, EPI_ISL_19007566, EPI_ISL_19007567, EPI_ISL_19007568, EPI_ISL_19007569, EPI_ISL_19007570, EPI_ISL_19007571, EPI_ISL_19007572, EPI_ISL_19007573, EPI_ISL_19007574, EPI_ISL_19007575, EPI_ISL_19007576, EPI_ISL_19007577, EPI_ISL_19007578, EPI_ISL_19007579, EPI_ISL_19007580, EPI_ISL_19007581, EPI_ISL_19007582, EPI_ISL_19007583, EPI_ISL_19007584, EPI_ISL_19007585, EPI_ISL_19007586, EPI_ISL_19007587, EPI_ISL_19007588, EPI_ISL_19007589, EPI_ISL_19007590, EPI_ISL_19007591, EPI_ISL_19007592, EPI_ISL_19007593, EPI_ISL_19007594, EPI_ISL_19007595, EPI_ISL_19007596, EPI_ISL_19007597, EPI_ISL_19007598, EPI_ISL_19007599, EPI_ISL_19007600, EPI_ISL_19007601, EPI_ISL_19007602, EPI_ISL_19007603, EPI_ISL_19007604, EPI_ISL_19007605, EPI_ISL_19007606, EPI_ISL_19007607, EPI_ISL_19007608, EPI_ISL_19007609, EPI_ISL_19007610, EPI_ISL_19007611, EPI_ISL_19007612, EPI_ISL_19007613, EPI_ISL_19007614, EPI_ISL_19007615, EPI_ISL_19007616, EPI_ISL_19007617, EPI_ISL_19007618, EPI_ISL_19007619, EPI_ISL_19007620, EPI_ISL_19007621, EPI_ISL_19007622, EPI_ISL_19007623, EPI_ISL_19007624, EPI_ISL_19007625, EPI_ISL_19007626, EPI_ISL_19007627, EPI_ISL_19007628, EPI_ISL_19007629, EPI_ISL_19007630, EPI_ISL_19007631, EPI_ISL_19007632, EPI_ISL_19007633, EPI_ISL_19007634, EPI_ISL_19007635, EPI_ISL_19007636, EPI_ISL_19007637, EPI_ISL_19007638, EPI_ISL_19007639, EPI_ISL_19007640, EPI_ISL_19007641, EPI_ISL_19007642, EPI_ISL_19007643, EPI_ISL_19007644, EPI_ISL_19007645, EPI_ISL_19007646, EPI_ISL_19007647, EPI_ISL_19007648, EPI_ISL_19007649, EPI_ISL_19007650, EPI_ISL_19007651, EPI_ISL_19007652, EPI_ISL_19007653, EPI_ISL_19007654, EPI_ISL_19007655, EPI_ISL_19007656, EPI_ISL_19007657, EPI_ISL_19007658, EPI_ISL_19007659, EPI_ISL_19007660, EPI_ISL_19007661, EPI_ISL_19007662, EPI_ISL_19007663, EPI_ISL_19007664, EPI_ISL_19007665, EPI_ISL_19007666, EPI_ISL_19007667, EPI_ISL_19007668, EPI_ISL_19007669, EPI_ISL_19007670, EPI_ISL_19007671, EPI_ISL_19007672, EPI_ISL_19007673, EPI_ISL_19007674, EPI_ISL_19007675, EPI_ISL_19007676, EPI_ISL_19007677, EPI_ISL_19007678, EPI_ISL_19007679, EPI_ISL_19007680, EPI_ISL_19007681, EPI_ISL_19007682, EPI_ISL_19007683, EPI_ISL_19007684, EPI_ISL_19007685, EPI_ISL_19007686, EPI_ISL_19007687, EPI_ISL_19007688, EPI_ISL_19007689, EPI_ISL_19007690, EPI_ISL_19007691, EPI_ISL_19007692, EPI_ISL_19007693, EPI_ISL_19007694, EPI_ISL_19007695, EPI_ISL_19007696, EPI_ISL_19007697, EPI_ISL_19007698, EPI_ISL_19007699, EPI_ISL_19007700, EPI_ISL_19007701, EPI_ISL_19007702, EPI_ISL_19007703, EPI_ISL_19007704, EPI_ISL_19007705, EPI_ISL_19007706, EPI_ISL_19007707, EPI_ISL_19007708, EPI_ISL_19007709, EPI_ISL_19007710, EPI_ISL_19007711, EPI_ISL_19007712, EPI_ISL_19007713, EPI_ISL_19007714, EPI_ISL_19007715, EPI_ISL_19007716, EPI_ISL_19007717, EPI_ISL_19007718, EPI_ISL_19007719, EPI_ISL_19007720, EPI_ISL_19007721, EPI_ISL_19007722, EPI_ISL_19007723, EPI_ISL_19007724, EPI_ISL_19007725, EPI_ISL_19007726, EPI_ISL_19007727, EPI_ISL_19007728, EPI_ISL_19007729, EPI_ISL_19007730, EPI_ISL_19007731, EPI_ISL_19007732, EPI_ISL_19007733, EPI_ISL_19007734, EPI_ISL_19007735, EPI_ISL_19007736, EPI_ISL_19007737, EPI_ISL_19007738, EPI_ISL_19007739, EPI_ISL_19007740, EPI_ISL_19007741, EPI_ISL_19007742, EPI_ISL_19007743, EPI_ISL_19007744, EPI_ISL_19007745, EPI_ISL_19007746, EPI_ISL_19007747, EPI_ISL_19007748, EPI_ISL_19007749, EPI_ISL_19007750, EPI_ISL_19007751, EPI_ISL_19007752, EPI_ISL_19007753, EPI_ISL_19007754, EPI_ISL_19007755, EPI_ISL_19007756, EPI_ISL_19007757, EPI_ISL_19007758, EPI_ISL_19007759, EPI_ISL_19007760, EPI_ISL_19007761, EPI_ISL_19007762, EPI_ISL_19007763, EPI_ISL_19007764, EPI_ISL_19007765, EPI_ISL_19007766, EPI_ISL_19007767, EPI_ISL_19007768, EPI_ISL_19007769, EPI_ISL_19007770, EPI_ISL_19007771, EPI_ISL_19007772, EPI_ISL_19007773, EPI_ISL_19007774, EPI_ISL_19007775, EPI_ISL_19007776, EPI_ISL_19007777, EPI_ISL_19007778, EPI_ISL_19007779, EPI_ISL_19007780, EPI_ISL_19007781, EPI_ISL_19007782, EPI_ISL_19007783, EPI_ISL_19007784, EPI_ISL_19007785, EPI_ISL_19007786, EPI_ISL_19007787, EPI_ISL_19007788, EPI_ISL_19007789, EPI_ISL_19007790, EPI_ISL_19007791, EPI_ISL_19007792, EPI_ISL_19007793, EPI_ISL_19007794, EPI_ISL_19007795, EPI_ISL_19007796, EPI_ISL_19007797, EPI_ISL_19007798, EPI_ISL_19007799, EPI_ISL_19007800, EPI_ISL_19007801, EPI_ISL_19007802, EPI_ISL_19007803, EPI_ISL_19007804, EPI_ISL_19007805, EPI_ISL_19007806, EPI_ISL_19007807, EPI_ISL_19007808, EPI_ISL_19007809, EPI_ISL_19007810, EPI_ISL_19007811, EPI_ISL_19007812, EPI_ISL_19007813, EPI_ISL_19007814, EPI_ISL_19007815, EPI_ISL_19007816, EPI_ISL_19007817, EPI_ISL_19007818, EPI_ISL_19007819, EPI_ISL_19007820, EPI_ISL_19007821, EPI_ISL_19007822, EPI_ISL_19007823, EPI_ISL_19007824, EPI_ISL_19007825, EPI_ISL_19007826, EPI_ISL_19007827, EPI_ISL_19007828, EPI_ISL_19007829, EPI_ISL_19007830, EPI_ISL_19007831, EPI_ISL_19007832, EPI_ISL_19007833, EPI_ISL_19007834, EPI_ISL_19007835, EPI_ISL_19007836, EPI_ISL_19007837, EPI_ISL_19007838, EPI_ISL_19007839, EPI_ISL_19007840, EPI_ISL_19007841, EPI_ISL_19007842, EPI_ISL_19007843, EPI_ISL_19007844, EPI_ISL_19007845, EPI_ISL_19007846, EPI_ISL_19007847, EPI_ISL_19007848, EPI_ISL_19007849, EPI_ISL_19007850, EPI_ISL_19007851, EPI_ISL_19007852, EPI_ISL_19007853, EPI_ISL_19007854, EPI_ISL_19007855, EPI_ISL_19007856, EPI_ISL_19007857, EPI_ISL_19007858, EPI_ISL_19007859, EPI_ISL_19007860, EPI_ISL_19007861, EPI_ISL_19007862, EPI_ISL_19007863, EPI_ISL_19007864, EPI_ISL_19007865, EPI_ISL_19007866, EPI_ISL_19007867, EPI_ISL_19007868, EPI_ISL_19007869, EPI_ISL_19007870, EPI_ISL_19007871, EPI_ISL_19007872, EPI_ISL_19007873, EPI_ISL_19007874, EPI_ISL_19007875, EPI_ISL_19007876, EPI_ISL_19007877, EPI_ISL_19007878, EPI_ISL_19007879, EPI_ISL_19007880, EPI_ISL_19007881, EPI_ISL_19007882, EPI_ISL_19007883, EPI_ISL_19007884, EPI_ISL_19007885, EPI_ISL_19007886, EPI_ISL_19007887, EPI_ISL_19007888, EPI_ISL_19007889, EPI_ISL_19007890, EPI_ISL_19007891, EPI_ISL_19007892, EPI_ISL_19007893, EPI_ISL_19007894, EPI_ISL_19007895, EPI_ISL_19007896, EPI_ISL_19007897, EPI_ISL_19007898, EPI_ISL_19007899, EPI_ISL_19007900, EPI_ISL_19007901, EPI_ISL_19007902, EPI_ISL_19007903, EPI_ISL_19007904, EPI_ISL_19007905, EPI_ISL_19007906, EPI_ISL_19007907, EPI_ISL_19007908, EPI_ISL_19007909, EPI_ISL_19007910, EPI_ISL_19007911, EPI_ISL_19007912, EPI_ISL_19007913, EPI_ISL_19007914, EPI_ISL_19007915, EPI_ISL_19007916 | | | |
| see above | Laboratoire de virologie, Centre National de Référence arbovirus Associe (CNR), Centre Hospitalier Universitaire de la Reunion (CHU) | Laboratoire de virologie, Centre National de Référence arbovirus Associe (CNR), Centre Hospitalier Universitaire de la Reunion (CHU) | Frumence,E., Traversier,N. and Jaffar Bandjee,M.-C. |
| EPI_ISL_19026416, EPI_ISL_19026417, EPI_ISL_19026418, EPI_ISL_19026419, EPI_ISL_19026420, EPI_ISL_19026421, EPI_ISL_19026422, EPI_ISL_19026423, EPI_ISL_19026424, EPI_ISL_19026425, EPI_ISL_19026426, EPI_ISL_19026427, EPI_ISL_19026428, EPI_ISL_19026429 | | | |
| see above | Nagasaki University, Department of Virology | Nagasaki University, Department of Virology | Ngwe Tun,M.M., Kyaw,A.K., Nabeshima,T., Dumre,S.P., Soe,A.M., Nwe,K.M., Myaing,S.S., Lwin,E.P., Win,Y.T., Inoue,S., Takamatsu,Y., Urano,T., Thu,H.M., Thant,K.Z., Htun,Z.T. and Morita,K. |
| EPI_ISL_19091309, EPI_ISL_19091310, EPI_ISL_19091311, EPI_ISL_19091312, EPI_ISL_19091313, EPI_ISL_19091314, EPI_ISL_19091315, EPI_ISL_19091316, EPI_ISL_19091317, EPI_ISL_19091318, EPI_ISL_19091319, EPI_ISL_19091320, EPI_ISL_19091321, EPI_ISL_19091322 | | | |
| see above | Virology, US Army Medical Directorate of the Armed Forces Research Institute of Medical Sciences (USAMD-AFRIMS) | Virology, US Army Medical Directorate of the Armed Forces Research Institute of Medical Sciences (USAMD-AFRIMS) | Gyeltshen,S., Dorji,K., Zangmo,S., Adhikari,L.M., Dorji,T., Wangchuk,S., Rodpradit,P., Phonpakobsin,T., Poolpanichupatam,Y., Chinnawirotpisan,P., Manasatienkij,W., Cotrone,T.S., Fernandez,S. and Klungthong,C. |
| EPI_ISL_8636670, EPI_ISL_8636671, EPI_ISL_8636672, EPI_ISL_8636673, EPI_ISL_8636674, EPI_ISL_8636675, EPI_ISL_8636676 | Guangxi Zhuang Autonomous Region Center for Disease Prevention and Control | Guangxi Zhuang Autonomous Region Center for Disease Prevention and Control | Wang,J. |

| | | | |
|---|---|---|---|
| EPI_ISL_8766816, EPI_ISL_8766817, EPI_ISL_8766818, EPI_ISL_8766819, EPI_ISL_8766820, EPI_ISL_8766821, EPI_ISL_8766822, EPI_ISL_8766823, EPI_ISL_8766824, EPI_ISL_8766825, EPI_ISL_8766826, EPI_ISL_8766827, EPI_ISL_8766828, EPI_ISL_8766876, EPI_ISL_8767061, EPI_ISL_8767062, EPI_ISL_8767063, EPI_ISL_8767064, EPI_ISL_8767626, EPI_ISL_8768614, EPI_ISL_8768615, EPI_ISL_8768616, EPI_ISL_8768617, EPI_ISL_8768889, EPI_ISL_8768890, EPI_ISL_8768891, EPI_ISL_8768892, EPI_ISL_8768893, EPI_ISL_8768894, EPI_ISL_8768895, EPI_ISL_8768896 | | | |
| see above | Shanghai Municipal Center for Disease Control and Prevention | Shanghai Municipal Center for Disease Control and Prevention | Wang,W. |
| EPI_ISL_8824344, EPI_ISL_8824350 | Emory University School of Medicine | Emory University School of Medicine | Key,A., Waggoner,J., Ali,I.M., Piantadosi,A.L. and Collins,M. |
| EPI_ISL_9004460, EPI_ISL_9004461, EPI_ISL_9004462, EPI_ISL_9004463, EPI_ISL_9004464 | UNIVERSITY OF HYDERABAD | UNIVERSITY OF HYDERABAD | Maisnam,D. and Musturi,V. |
| EPI_ISL_9004465, EPI_ISL_9004466, EPI_ISL_9004467, EPI_ISL_9004468, EPI_ISL_9004469 | Guangzhou Center for Disease Control and Prevention | Guangzhou Center for Disease Control and Prevention | Su,W., Cao,Y. and Lu,W. |
| EPI_ISL_9004470 | Guangzhou Center for Disease Control and Prevention | Guangzhou Center for Disease Control and Prevention | Su,W., Jiang,L. and Lu,W. |
| EPI_ISL_9103208, EPI_ISL_9103209, EPI_ISL_9103210, EPI_ISL_9103211 | Chinese Academy of Sciences | Chinese Academy of Sciences | lihong,H. |
| EPI_ISL_9152287, EPI_ISL_9152288, EPI_ISL_9152289, EPI_ISL_9152290, EPI_ISL_9152291, EPI_ISL_9152292, EPI_ISL_9437081, EPI_ISL_9437082 | Secretaria de Salud | Secretaria de Salud | Wong-Arambula,C.E., Rodriguez-Maldonado,A.P., Fuentes-Cuevas,C.H., Duran-Ayala,D., Torres-Rodriguez,Mdl.L., Rivero-Arredondo,V., Serrano-Gutierrez,S., Aparicio-Antonio,M., Rosales-Jimenez,C., del Mazo-Lopez,J.C., Hernandez-Rivas,L., Lopez-Martinez,I., Ramirez-Gonzalez,J.E. and Vazquez-Pichardo,M. |
| EPI_ISL_9636328, EPI_ISL_9636329, EPI_ISL_9636330, EPI_ISL_9636331, EPI_ISL_9636332 | University Of Hyderabad | UNIVERSITY OF HYDERABAD | Maisnam,D. and Musturi,V. |
| EPI_ISL_9711778, EPI_ISL_9711914, EPI_ISL_9711959, EPI_ISL_9712018, EPI_ISL_9712019, EPI_ISL_9712020, EPI_ISL_9712021, EPI_ISL_9712022, EPI_ISL_9712023, EPI_ISL_9712024, EPI_ISL_9713936, EPI_ISL_9713937, EPI_ISL_9713980, EPI_ISL_9713981, EPI_ISL_9713982, EPI_ISL_9713983, EPI_ISL_9713984, EPI_ISL_9713985, EPI_ISL_9713986, EPI_ISL_9713987, EPI_ISL_9713988, EPI_ISL_9713989, EPI_ISL_9713990, EPI_ISL_9713991 | | | |
| see above | National Institute of Virology | National Institute of Virology | Yadav,P.D. |
| EPI_ISL_9713992 | Adolfo Lutiz Institute | Adolfo Lutiz Institute | Cunha,M.S., de Moura Coletti,T., Guerra,J.M., Ponce,C.C., Fernandes,N.C.C.A., Resio,R.A., Claro,I.M., Salles,F., Lima Neto,D.F. and Sabino,E. |
| EPI_ISL_9768996, EPI_ISL_9769066 | Fundacion Universitaria Autonoma de las Americas | Fundacion Universitaria Autonoma de las Americas | Rojas-Gallardo,D.M., Restrepo-Chica,J., Key,A., Acosta,A., Castrillon-Spitia,J.D., Tabares-Villa,F.A., Trujillo,A.M., JJimenez-Posada,E.V., Martinez-Gutierrez,M., Ruiz-Saenz,J., Waggoner,J., Collins,M.H., Piantadosi,A. and Cardona-Ospina,J.A. |
| EPI_ISL_9845835, EPI_ISL_9845836, EPI_ISL_9882074, EPI_ISL_9882077, EPI_ISL_9935327, EPI_ISL_9935714, EPI_ISL_9936221, EPI_ISL_9988597, EPI_ISL_9988598 | National Institute of Virology | National Institute of Virology | Yadav,P.D. |
